# A Prospective Real-time Early Warning System to Anticipate Onsets and Peaks of Respiratory Diseases Outbreaks at the State Level in the U.S. A Transfer Learning Approach Leveraging Digital Traces

**DOI:** 10.1101/2025.10.10.25337739

**Authors:** Raul Garrido Garcia, Leonardo Clemente, Austin Meyer, George Dewey, Shihao Yang, Mauricio Santillana

## Abstract

Respiratory disease outbreaks burden U.S. healthcare systems with over one million hospitalizations annually, yet current surveillance systems lag 1-2 weeks behind real-time conditions, preventing timely intervention. We developed a machine learning early warning system that combines Google search trends with traditional epidemiological data using ensemble voting algorithms to predict the timing of outbreak onsets and peaks across multiple respiratory pathogens. The system applies anomaly detection and transfer learning to monitor syndromic Influenza-like illnesses (ILI), and hospitalizations caused by respiratory syncytial virus (RSV) or Influenza, simultaneously, across all 50 US states. During operational real-time deployment from August 2024 through the 2024-2025 season, the system detected 98.0% of outbreak onsets with 5-week average lead time and 97.0% of peaks with 2-week average lead time, achieving positive predictive values that exceed 82%. This framework transforms reactive public health responses into proactive epidemic preparedness by reducing historical timing uncertainty from 10-20 weeks to consistent 2-6 week prediction windows, providing a scalable approach for monitoring both seasonal outbreaks and emerging respiratory threats.

## 1 Introduction

Syndromic surveillance of influenza-like illnesses (ILI) in the United States has been conducted for over three decades and serves as a cornerstone of public health preparedness against respiratory disease outbreaks. The World Health Organization defines ILI as an acute respiratory infection characterized by fever ≥38°C and a cough (or sore throat) with onset within the past 10 days [1]. This broad definition enables surveillance systems to capture infections caused by influenza, respiratory syncytial virus (RSV), and SARS-CoV-2, among other respiratory pathogens. The public health burden due to ILI is substantial: during the 2024–2025 season alone, the U.S. experienced an estimated 47 million influenza cases with 610,000 hospitalizations, while RSV contributed 190,000 hospitalizations and COVID-19 added 250,000 more [2–4]. Accurate early detection of these outbreaks is essential for timely interventions, including vaccination campaigns, allocation of healthcare resources, and public health messaging before healthcare systems become overwhelmed.

Current surveillance systems face critical timing limitations that constrain proactive public health responses. For example, the United States Centers for Disease Control and Prevention (CDC)’s influenza surveillance network provides comprehensive data but often reflects events from 1 to 2 weeks earlier in the season, resulting in reports retrospective rather than predictive [5]. This delay hampers public health authorities’ efforts to implement preventive measures before outbreaks of respiratory diseases reach their peaks. State-of-the-art efforts to forecast the timing of the onset and peak of respiratory diseases outbreaks in the US, led by the FluSight CDC initiative, have remained experimental in nature [6–10]. The CDC FluSight models are evaluated for their ability to identify the timing of onsets and peaks only after the events in question have occurred. Even in this retrospective mode, the forecast scores (ranging 0.08-0.49 for the peak week and 0.01-0.41 for onset week) indicate that these models, on average, assign a probability between 1% and 49% that these events occurred in the correct week [7]. This broad range suggests that these forecasts are widely uncertain about the timing and occurrence of the onsets and peaks they aim to identify. Further complicating surveillance efforts are the multiple circulating types of respiratory infections which contribute to annual ILI incidence. Recent work has shown that influenza and RSV outbreaks have distinct timing relationships which vary across states; these differences are especially important in the context of healthcare infrastructure and the deployment of vaccinations and other therapeutics [11].

To enhance and refine surveillance efforts, researchers and public health authorities have embraced data from digital traces –information that is left behind by internet users navigating the web– which have emerged as promising indicators of infectious disease activity [12–18]. Indicators such as Google search trends for respiratory symptoms and treatments have been shown to capture population-level changes in health-seeking behavior that often precede clinical case reporting[13–17, 19–23]. Previous studies have demonstrated the utility of these digital traces for COVID-19 surveillance and their limitations[24–26], with Kogan et al. [27] developing Bayesian indicators that forecast outbreaks weeks in advance, and Stolerman et al. [28] creating county-level early warning systems using digital signals to estimate the reproductive number of county-level COVID-19 outbreaks. However, these approaches have primarily focused on single diseases and have not addressed the challenge of monitoring multiple respiratory diseases simultaneously across diverse geographic regions with varying outbreak dynamics.

Building on these advances, we developed a comprehensive machine learning framework that addresses key limitations in current respiratory disease surveillance. Unlike previous single-disease approaches, our early warning system integrates multiple data streams simultaneously to monitor ILI, influenza hospitalizations, and RSV activity across all 50 U.S. states. Rather than relying on complex reproductive number estimations which are prone to reporting delays and underascertainment [29–31], our approach leverages ensemble voting mechanisms with digital behavioral signals (primarily influenza-related Google search activity) to directly predict outbreak onsets and peaks. We additionally use transfer learning to extend our framework to different respiratory diseases despite the limited availability of historical data. We validated our methodology retrospectively using historical ILI data (2010–2020), tested retrospectively during the 2022–2024 ILI seasons, and deployed prospectively and in real-time for the 2024–2025 ILI season in operational collaboration with the CDC’s Center for Forecasting and Outbreak Analytics, with predictions shared continuously with public health decision-makers beginning in August 2024.

## 2 Results

Respiratory disease outbreak timing varies dramatically across U.S. states: our analysis of historical ILI data (2010–2020) shows onset timing varies by an average of 10.7 weeks and peak timing by up to 20 weeks (mean range 14.4 weeks) across states (Supplementary Fig. S1, Supplementary Fig. S2). This unpredictability—with some states beginning ILI season within a one-month window while others exhibit 2–4 months of temporal spread—makes historical patterns unreliable for prospective prediction and limits their utility for public health preparedness.

To address this critical gap, we developed and validated a machine learning early warning system that integrates digital behavioral signals (Google search trends) with traditional epidemiological data through ensemble voting algorithms to anticipate outbreak onsets and peaks 2–6 weeks in advance. We validated our approach through three phases: retrospective validation (2010–2020), out-of-sample testing (2022–2024), and real-time operational deployment (2024–2025) in collaboration with CDC’s Center for Forecasting and Outbreak Analytics, with prospective predictions shared continuously beginning in August 2024.

We assessed performance using three operationally relevant metrics: (i) detection sensitivity (proportion of onsets and peaks successfully anticipated), (ii) lead time (average weeks in advance), and (iii) positive predictive value (PPV; alarm reliability). These metrics, evaluated across all validation phases, quantified how well the EWS narrowed wide historical timing variability into consistent, actionable prediction windows. The main contributions are (1) development and real-time validation of the ILI early warning system across all 50 U.S. states, and (2) successful transfer learning extension to influenza hospitalizations and RSV activity with minimal training data.

### Early Warning System for ILI

We designed our early warning system by inspecting ILI temporal trends and the American public’s influenza-related Google searches at the state and national levels from 2010 to 2020. Specifically, we used time series anomaly detection approaches to characterize the timing of the onsets and peaks in the weekly temporal trends of ILI,. To label the onsets of outbreaks of ILI, we identified the beginning of time periods with consistent exponential growth in the number of reported ILI cases [27, 28] and implemented standard peak detection algorithms [32]. It is important to note that both of these strategies, when implemented in real-time and for prospective purposes, require future-looking data to detect the timing of the event. For example, generally speaking using standard peak detection packages, such as scipy in python [32] or scorepeak in R [33], a peak can only be identified when one observes that the value of the time series, at a given week, is higher than both the previous value of the time series and the next value of the time series. As as a result, both of our strategies label an event (either an onset or peak) about 3 weeks after it has occurred.

#### System Development and Retrospective Validation (2010–2020)

Our early warning system borrows elements from methodologies implemented for real-time onset identification in the context of COVID-19 [27, 28], and consists of identifying historical temporal anomalies in both the patterns of the general public’s disease-related Google searches and in events that may occur in neighboring states that anticipate the emergence of an event in a given state. Intuitively speaking, we hypothesize that a spike in cases of a disease may be detected in a population when sharp increases (exponential growth) in multiple disease-related search queries occur. We identify these queries and the number of queries needed to label an event using an ensemble voting methodology, which combines the results of multiple machine learning models to generate a final prediction. More details are presented in the subsequent sections.

To establish confidence in our methodology, we conducted extensive retrospective evaluations and validation using historical ILI data spanning 10 seasons, starting in the fall of 2010. These analyses were performed for all 50 U.S. states and the national level, with the exception of Florida, where ILINet data were not available. These evaluations allowed us to identify that a training set that consists of a rolling window of the most recent three years to predict subsequent outbreak patterns was optimal. We maintained strict temporal ordering in our experiments to simulate real-world deployment conditions.

With such a choice of training set length, we evaluated the performance of our approach retrospectively during the time period from the fall of 2014 to winter of 2020. In this time period, our EWS detected 99.3% of outbreak onsets with an average lead time of 3.4 weeks and achieved a positive predictive value (PPV) of 93.0% (meaning that 93.0% of alarms that were produced anticipated the occurrence of an onset and only 7.0% of alarms were false or too early). Peak detection during this retrospective validation period reached 84.2% sensitivity (the percent of true events that were correctly anticipated) with 3.9-week lead time and 69.3% PPV(Supplementary Fig. S3 and Fig. S4). This means that among 433 alarms 293 actually anticipated a peak. The fact that in 50 locations during the 6 years of validation we observed 316 peaks, suggests that during a given season there may be locations that may experience more than one peak, making this specific task more challenging than onset detection. These results established the foundational performance characteristics of our ensemble voting approach and validated the correlation-based feature selection methodology. Having established the system’s performance characteristics on historical data, we next evaluated whether these results would hold on previously unseen seasonal patterns.

#### Out-of-Sample Validation (2022–2024)

Out-of-sample testing on the 2022–2023 and 2023– 2024 seasons was used to further validate the system’s predictive capabilities. All hyperparameters established during development—including the onset and peak definitions, the three-season training window, and the correlation threshold for Google search term selection—were fixed during this evaluation. The only parameter adjusted was the voting system threshold (see Methods for details). The system successfully detected 99.0% of outbreak onsets with 3.8-week lead time and 96.0% PPV, while peak detection achieved 83.5% sensitivity with 2.6-week lead time and 82.7% PPV. These results demonstrated consistent performance across different seasonal patterns and confirmed the system’s readiness for real-time operational deployment. This high performance suggests that our EWS demonstrates robust generalizability of the digital behavioral signals as predictive variables and ensemble voting mechanisms across evolving epidemiological contexts. The consistent performance across the 2022-2024 seasons confirmed the system was ready for operational real-time deployment with prospective predictions shared with public health authorities before events occurred.

#### Prospective and Real-time Operational Deployment Evaluation (2024–2025)

We next deployed our EWS in real-time during the 2024-2025 ILI season through operational collaboration with the CDC’s Center for Forecasting and Outbreak Analytics. Prospective predictions were shared continuously with CDC researchers and decision-makers beginning in August 2024, providing independent validation of the system’s operational performance. During this time period, our early warning system demonstrated robust performance in detecting respiratory disease outbreaks across all 50 US states. The system successfully anticipated the timing of 98.0% of outbreak onsets with an average lead time of 5 weeks and achieved a PPV of 83.0% (Fig. 1). Only 14.0% of alarms represented false positives not followed by confirmed outbreaks in the subsequent 6 weeks. The system narrowed the historically wide onset detection window to a consistent 4 to 6 week range across nearly all states, representing a 2 to 4 fold reduction in timing uncertainty for states with the highest historical variability.

**Fig. 1:**
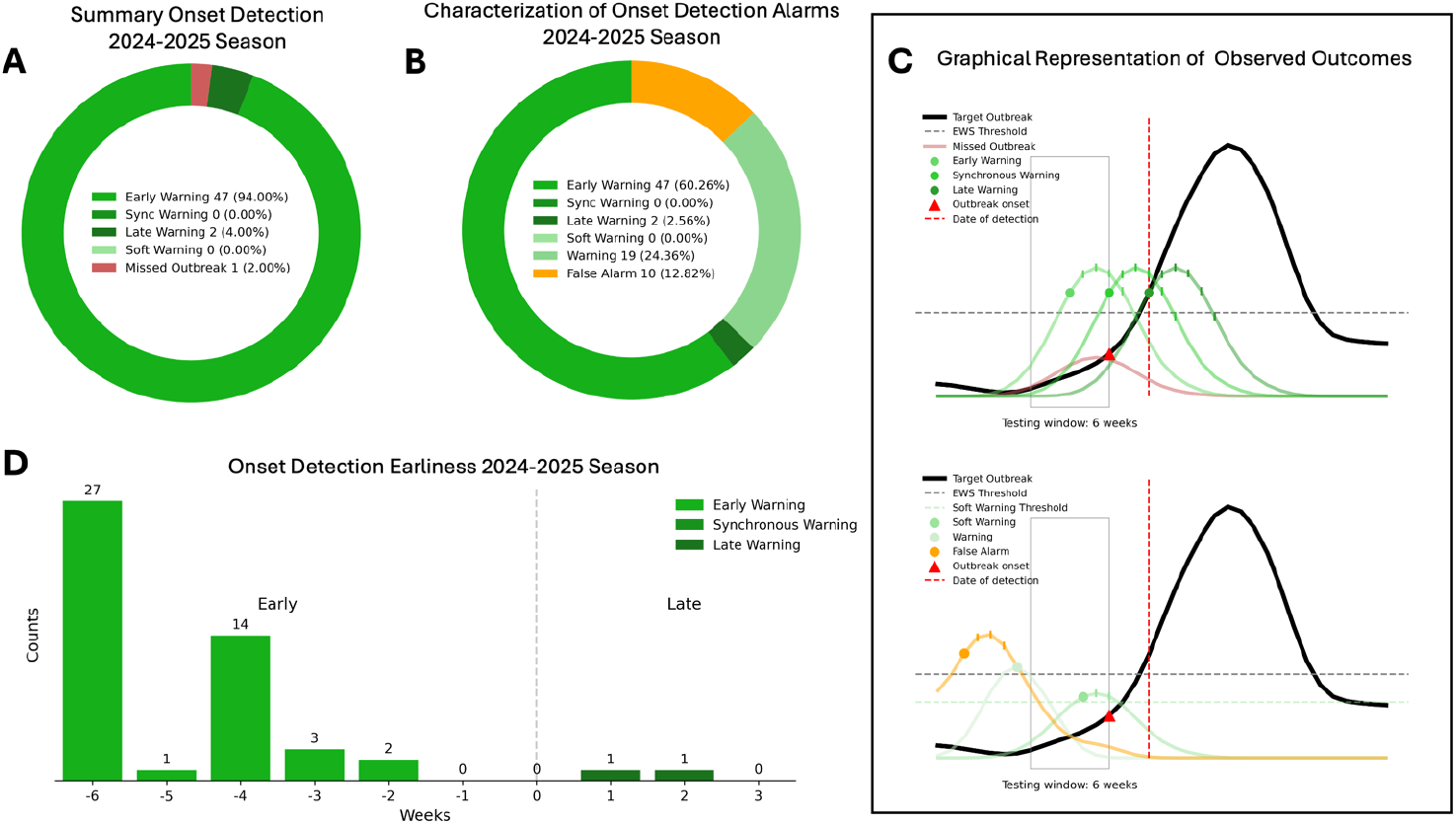
Real-time early warning system (EWS) onset detection performance for the 2024–2025 season, during operational deployment with CDC’s Center for Forecasting and Outbreak Analytics, achieving 98% detection with an average 5-week lead time and a positive predictive value of 83%. **A)** Overall classification of onset detections across U.S. states, categorized by warning type—Early, Synchronous, Late, Soft, and Missed. The donut chart shows that the majority of onsets were detected early, with a smaller portion detected synchronously or late, and a minimal proportion missed entirely. **B)**Distribution of all detection alarms, including false alarms. This breakdown illustrates the relative frequency of true positives, false positives, and the proportion of alarms that did not correspond to an actual onset. **C)** Schematic diagrams illustrating the temporal relationship between predicted and actual onsets across different detection types. Multiple scenarios are visualized, including early, synchronous, and late detections, as well as false alarms. These curves represent hypothetical trajectories of onset activity, with corresponding system predictions overlaid for clarity. **D)** Histogram of detection earliness for onsets. The x-axis shows the number of weeks between the detection and the actual onset, where negative values represent early warnings, zero represents detection during the onset week, and positive values indicate late warnings. The majority of detections occurred 3 to 5 weeks before the onset, indicating good anticipatory performance.

Peak detection performance was similarly strong, and the system accurately identified the timing of epidemic peaks in 97.0% states with an average lead time of 2 weeks, with PPV of 82.7%. 17.3% of peak alarms were classified as false positives (Fig. 2). This performance significantly outpaced the substantial historical uncertainty in peak timing, where natural variability spans 13–20 weeks in the most unpredictable states. The slight reduction in PPV compared to retrospective validation (83.1% vs. 92.9% for onsets) reflects the inherent challenges of real-time prediction in operational settings, including evolving disease dynamics and potential changes in digital behavior patterns. However, the maintained high sensitivity (98.0%) demonstrates the system’s reliability for public health decision-making.

**Fig. 2:**
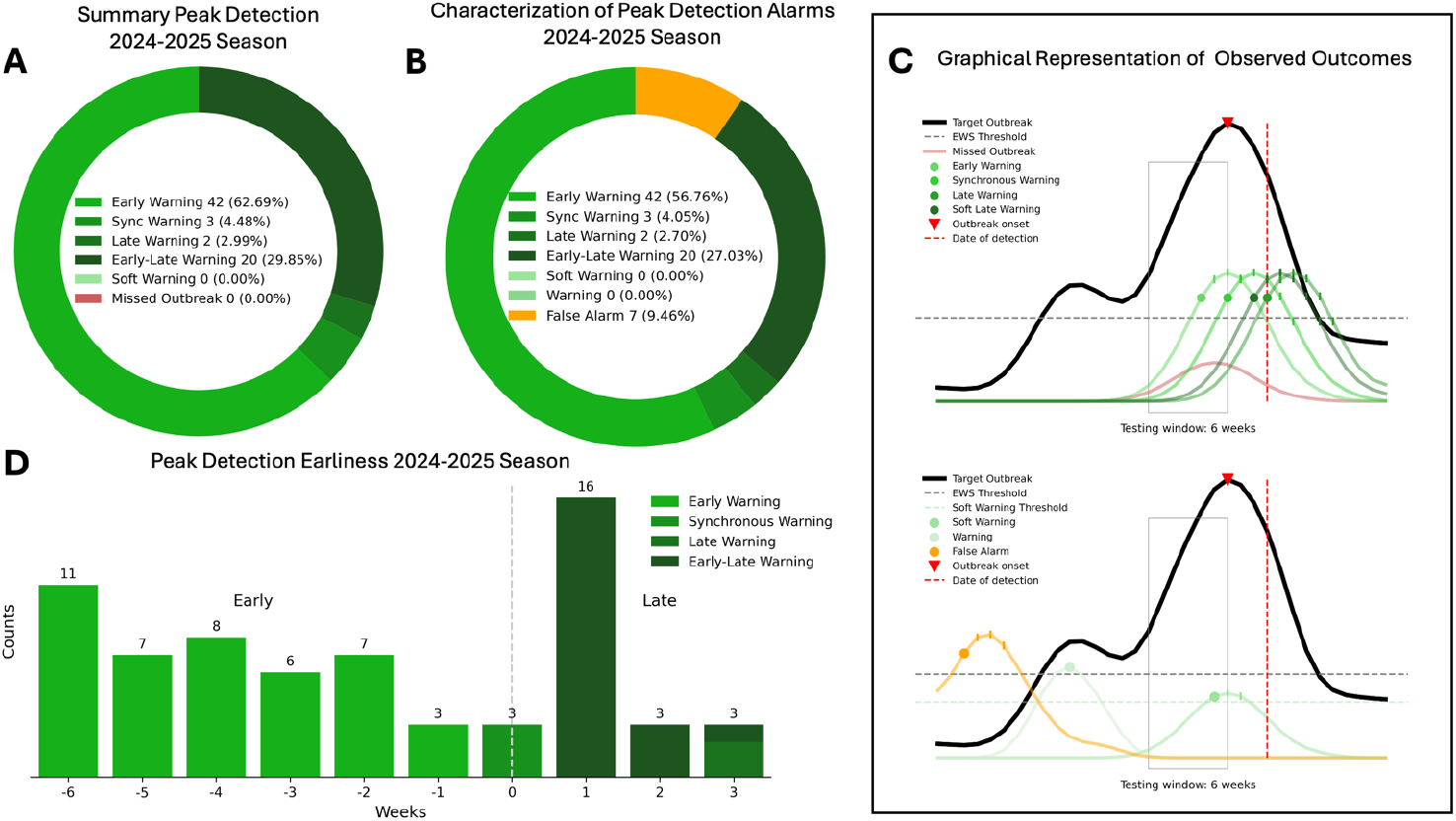
Real-time early warning system (EWS) peak detection performance for the 2024–2025 season, during operational deployment with CDC’s Center for Forecasting and Outbreak Analytics, achieving 97.0% detection with an average 2-week lead time and a positive predictive value of 82.7%. **A)**Overall classification of peak detections across U.S. states, categorized by warning type—Early, Early-Late, Synchronous, Late, Soft, and Missed. The donut chart shows that the majority of peaks were detected early, followed by a smaller proportion of synchronous, late, and early-late warnings. **B)**Distribution of all detection alarms, including false alarms. This breakdown illustrates the relative frequency of true positives, false positives, and the proportion of alarms that did not correspond to an actual peak. **C)** Schematic diagrams illustrating the temporal relationship between predicted and actual peaks across different detection types. Multiple scenarios are visualized, including early, early-late, synchronous, and late detections, as well as false alarms. These curves represent hypothetical trajectories of peak activity, with corresponding system predictions overlaid for clarity. **D)** Histogram of detection earliness for peaks. The x-axis shows the number of weeks between the detection and the actual peak, where negative values represent early warnings, zero represents detection during the peak week, and positive values indicate late warnings. The majority of detections occurred 3 to 6 weeks before the peak or the week after the peak preceding the algorithmic detection, indicating good anticipatory performance.

### Transfer Learning Extension to RSV and Influenza Hospitalizations

Based on the strong performance of our EWS using both retrospective and prospective data, we next used domain transfer learning to extend our framework to additional targets for respiratory disease. The feasibility of this approach is based on the hypothesis that the search activity related to respiratory disease will anticipate events for other indicators of respiratory diseases, such as hospitalizations for RSV and influenza. In practice, transfer learning was implemented by reusing the feature selection, model structure, and alarm-generation rules established for ILI, while recalibrating the alarm threshold with the limited available data for each new target disease. This strategy allows models trained on long ILI histories to be rapidly adapted to diseases with only one or two seasons of observations. This transfer learning approach has been shown to be viable despite the limited availability of historical data [34]. If such a hypothesis is correct, we could use the training approaches and feature selection techniques developed for ILI and rapidly design and train a new surveillance system for a related respiratory disease indicator with minimal training data.

#### Influenza Hospitalizations

For influenza hospitalizations, where only two seasons of training data were available (2022–2024), the system achieved 96.0% onset detection during the 2024–2025 season with 4-week average lead time and 82.8% PPV. Peak detection for influenza hospitalizations reached 60.8% sensitivity with a 2.6 week lead time and 94.4% PPV, with only 5.6% false alarms (Supplementary Fig. S5). The reduced peak detection sensitivity compared to ILI likely reflects differences in hospitalization reporting patterns and the more severe disease threshold, which may exhibit different relationships with early digital behavioral signals.

#### RSV Activity

RSV surveillance presented the greatest transfer learning challenge, with training data limited to just one historical season (2023–2024). Despite this severe constraint, the transfer learning approach successfully detected 87.2% of RSV activity onsets during 2024–2025 with 1.2-week lead time and achieved 100% PPV with zero false alarms. RSV peak detection reached 68.0% sensitivity with 2.0 weeks of lead time and 84.6% PPV (Supplementary Fig. S6).

The perfect PPV for RSV onset detection observed during the 2024–2025 season, despite limited training data, suggests that the digital behavioral signals for RSV may be particularly relevant to RSV activity and that the transfer learning approach offers a promising way to capture the essential predictive signals that arise during the RSV season. The shorter lead times (1.2 weeks vs. 5 weeks for ILI) may reflect more rapid progression of RSV outbreaks or differences in population search behavior patterns.

#### Comparative Performance Across Respiratory Pathogens

Our early warning system demonstrated consistent early detection capabilities across multiple respiratory disease targets during 2024–2025, achieving onset detection rates of 98.0% for ILI, 96.0% for influenza hospitalizations, and 87.2% for RSV activity, with lead times ranging from 1.2 to 5 weeks. Peak detection performance varied by disease target, with ILI showing the strongest performance (97.0% detection) followed by RSV (68.0%) and influenza hospitalizations (60.8%). Positive predictive values remained consistently high across all targets, ranging from 82.7% to 100%, indicating low false alarm rates suitable for operational public health decision-making. The performance gradation from the ILI EWS to those for influenza hospitalizations and RSV likely reflects both the reduced amount of available training data (10+ years vs. 2 seasons vs. 1 season) and inherent differences in disease progression patterns and their relationships to digital behavioral signals. The system’s ability to maintain operational performance across diseases with vastly different training data availability highlights the effectiveness of our transfer learning approach and suggests broad applicability to emerging respiratory threats. As additional training data becomes available, we expect continued performance improvements, particularly for peak detection in newer surveillance targets.

## 3 Discussion

Our early warning system achieved 98% onset detection and 97.0% peak detection for ILI during real-time deployment with average lead times of 5 weeks and 2 weeks, respectively. These results directly address the core problem that has limited implementation of early warning systems for public health preparedness: historical ILI onset timing varies by 10 to 20 weeks across states, while our system provides consistent 4 to 6 week prediction windows for onsets and 2 to 4 week windows for peaks. For states with the highest historical variability, this represents a 2–4 fold reduction in timing uncertainty, enabling health departments to plan vaccination campaigns, adjust hospital staffing, and coordinate resource allocation with specific timeframes rather than broad seasonal estimates. Furthermore, we demonstrate that the methods used in our ILI EWS can be applied to data from individual respiratory infections such as time series of influenza hospitalizations or RSV activity.

Importantly, these results were independently validated through our operational collaboration with CDC’s Center for Forecasting and Outbreak Analytics. By sharing prospective predictions continuously with CDC experts beginning in August 2024—before outbreaks occurred—we established transparent, real-time verification of system performance that distinguishes this work from retrospectively evaluated forecasting efforts.

### Overcoming Fundamental Limitations in Traditional Disease Surveillance

The results of this study suggest that our EWS methodology is a distinct improvement over traditional surveillance and forecasting methods [27, 28]. Rather than estimating effective reproductive numbers prone to reporting delays and under-ascertainment [29, 30], we directly leverage early behavioral signals through an ensemble voting strategy that aggregates Google search trends and neighboring state disease dynamics. By predicting outbreaks using only preceding signals, we avoid the pitfall of extrapolating estimates from delayed surveillance data that often becomes available only after events have occurred, resulting in models experiencing look-ahead bias.

As a result of these improvements, our models performed well compared to existing CDC FLUsight challenge results. While current forecasting models achieve onset detection scores of 0.01–0.41 (assigning 1–41% probability to the correct timing week) and peak detection scores of 0.08–0.49 [7], our system achieved 98% onset detection and 97.0% peak detection with 5-week and 2-week lead times, respectively.. This improvement reflects the advantage of utilizing behavioral signals that anticipate rather than follow epidemiological patterns. When people search for respiratory disease symptoms or treatments, they often do so before seeking clinical care, creating an early warning signal that our ensemble voting approach effectively captures [13, 14, 16, 19–21].Our Granger causality analysis quantifies this temporal relationship, with 92.7% of selected search terms demonstrating statistical precedence over disease activity, providing empirical support for the intuitive notion that health-seeking behavior manifests first in digital spaces before appearing in clinical settings.

### Transfer Learning Demonstrates Scalability Despite Data Constraints

The transfer learning framework we implemented successfully extended surveillance capabilities of our EWS to outbreaks caused by other respiratory diseases with varying data availability [34]. The progression from ILI (10+ years training, 98% onset detection) to influenza hospitalizations (2 seasons training, 96% onset detection) to RSV (1 season training, 87.2% onset detection) demonstrates both the framework’s adaptability and the expected relationship between historical data availability and detection performance.While performance declined as a result of the transfer learning process, the impacted performance was expected as a result of the decrease in the quantity of available training data. Despite minimal training data, the RSV implementation achieved 100% positive predictive value for onset detection, indicating that when the system triggered alarms, the alarms consistently preceded actual outbreaks. However, this finding should be interpreted cautiously given the single-season training constraint and relatively small sample size. The lower overall detection rate (87.2%) suggests that while the system produces few false alarms for RSV, predictions for RSV outbreaks may not conform to the limited historical patterns available for training.

The varying performance characteristics across disease targets provide insights into the relationship between clinical presentation and digital behavioral signals, though these interpretations require further validation. RSV’s shorter lead times (1.2 weeks vs. 5 weeks for ILI) may reflect the more rapid progression of RSV outbreaks or differences in population search behaviors, particularly given RSV’s disproportionate impact on vulnerable populations (such as infants and the elderly) who may have caregivers searching on their behalf rather than self-searching. Similarly, the reduced peak detection sensitivity for influenza hospitalizations (60.8%) compared to ILI (97.0%) likely reflects the higher severity threshold for hospitalization, which may have weaker relationships with early behavioral signals that precede milder symptomatic illness.

Taken together, these results suggest potential for rapid deployment of early warning systems to outbreaks of emerging respiratory threats, though with important caveats. The consistent detection of respiratory disease signals across different pathogens indicates that behavioral patterns may share common elements, but the limited training data for newer targets means that initial deployment performance may be constrained. The framework’s value for pandemic preparedness lies primarily in its ability to provide early warning capability quickly, even if initial performance is modest, with the expectation that accuracy would improve as outbreak data accumulates. This capability is precisely what’s needed for pandemic preparedness: the ability to deploy surveillance systems to novel pathogens within weeks rather than waiting years to accumulate sufficient training data, as demonstrated by our RSV implementation using only a single season of historical observations.

Implementation of such early warning systems will require buy-in from both scientists and public health authorities, in addition to careful coordination with administrators of healthcare facilities and leaders in local government.

### Immediate Operational Applications May Transform Public Health Response

The predicitive abilities of these early warning systems enable immediate practical applications that demonstrate the framework’s operational value for evidence-based public health decision-making. For example, current ACIP guidelines recommend uniform October–March administration of the RSV prophylactic nirsevimab that cannot account for substantial state-level variability in RSV timing. Our system’s ability to predict RSV onsets with 1 to 2 weeks of lead time with high positive predictive value may enable health departments and hospitals to optimize distribution of prophylactics or vaccines by adjusting administration start dates for early seasons, prioritizing shipments based on outbreak predictions, and minimizing both under-protection and wasted coverage from premature dosing [11, 35–37].

Similar applications extend to influenza vaccination campaigns, where 5-week lead time of ILI onsets enables targeting of high-risk populations before community transmission accelerates. Hospital surge planning benefits from 2-week peak detection lead time, allowing for staffing adjustments, supply chain preparation, and patient flow optimization. These applications demonstrate how the framework transforms reactive public health responses into proactive epidemic preparedness, delivering on the scalable monitoring capability promised in our approach.

### Current Limitations and Implications for Generalizability

This study has several limitations. First, constrained training data for newer surveillance targets and inherent challenges in peak detection methodology results in performance differences across pathogens, with peak detection ranging from 60.8% for influenza hospitalizations to 97.0% for ILI. These differences reflect both data availability constraints and fundamental differences in how severe disease manifestations relate to early behavioral signals. Short training histories limit generalizability, though performance trends suggest substantial improvement as data accumulate. Second, we selected a 6-week window as it represented a reasonable balance for identifying outbreak onsets for long outbreaks. Importantly, alternative choices of this parameter would have yielded qualitatively similar results across respiratory diseases, as the qualitative patterns remained consistent, see Supplementary Materials. This consistency arises because the same methodological framework (Supplementary Algorithm 1) was applied uniformly across all time series—ILI, RSV, influenza, and Google searches—so any modification of the parameter would have produced analogous effects across diseases and predictors without altering the overall conclusions. Third, our deliberate choice of binary outputs over probabilistic forecasts prioritizes interpretability for decision-makers while maintaining high positive predictive values (82–100%) across disease targets. We opted against using Bayesian probabilistic approaches given limited training data availability, which would yield poorly calibrated probabilities overly sensitive to modeling assumptions. This design choice reflects the operational reality that public

health officials need actionable yes or no decisions rather than complex probabilistic distributions. Lastly, the inherent challenge of peak detection — requiring post-peak data — results in some late warnings, though many soft warnings that approach alarm thresholds still provide valuable confirmatory situational awareness. Additionally, our reliance on digital behavioral signals creates potential vulnerabilities to changes in search behavior patterns or platform algorithms [24–26], though the consistency of performance across multiple seasons suggests reasonable robustness.

### Broader Implications for Pandemic Preparedness and Future Directions

In conclusion, our framework’s significance extends beyond seasonal respiratory disease surveillance to pandemic preparedness and emerging threat detection [18, 31]. Collaborative forecasting efforts during COVID-19, while valuable for longer-term projections[38], highlighted the continued need for early warning systems that can detect outbreak onsets and peaks prospectively with actionable lead times. The disease-agnostic approach we employed enables rapid deployment to novel pathogens by leveraging behavioral signals that precede clinical reporting, potentially serving as a generalized monitoring system for future respiratory threats. The successful real-time deployment during 2024–2025, achieving detection rates exceeding 95% for onsets across multiple pathogens with 2–5 week lead times, demonstrates that integrating digital behavioral signals with traditional epidemiological data can transform reactive public health responses into proactive epidemic preparedness. Future research directions should focus on expanding the behavioral signal repertoire beyond Google search trends to include internet searches from clinicians and other providers [39], social media patterns, mobility data, and healthcare utilization signals. Integration with genomic surveillance data could enhance specificity for emerging variants or novel pathogens. Development of adaptive weighting approaches that dynamically update ensemble parameters within seasons could improve performance for rapidly evolving outbreaks.

The international transferability of this approach presents both opportunities and challenges. While the underlying premise—that health-seeking behaviors precede clinical case reporting—likely holds across healthcare systems, differences in digital infrastructure, search behavior patterns, and healthcare access may require region-specific calibration. Collaborative international implementation could provide valuable comparative insights into digital behavioral signal universality. As digital data streams expand and training datasets accumulate, this approach offers a sustainable pathway for enhancing real-time disease surveillance capabilities at the scale and resolution needed for effective public health action. The demonstrated ability to maintain operational performance across diseases with vastly different training data availability suggests broad applicability to emerging respiratory threats, fulfilling the scalable monitoring promise essential for future pandemic preparedness.

## 4 Data and Methods

Our approach combines traditional epidemiological surveillance with digital behavioral signals through an ensemble voting mechanism that predicts outbreak onsets and peaks 2–6 weeks in advance. The system operates by identifying early signals from proxy data sources—Google search trends and neighboring state disease activity—that historically precede confirmed outbreaks in target locations.

### Data Sources

We integrated multiple data streams to develop and validate our early warning system across different respiratory disease targets. The following datasets provided the foundation for model training, validation, and real-time deployment.

### Official Influenza-Like Illness Reports

The dataset comprised a weekly time series of the percentage of outpatient visits due to Influenzalike illness (ILI), spanning from October 9, 2010, to April 12, 2025. This data covered each U.S. state and the national level, as provided by the Centers for Disease Control and Prevention (CDC) [40]. The extensive temporal coverage of ILI data enabled comprehensive model training and served as the primary target for initial system development and validation.

### Official Respiratory Syncytial Virus Emergency Department Visits Reports

The dataset comprised a daily time series of Respiratory Syncytial Virus (RSV) emergency department visits, spanning from October 8, 2022, to March 9, 2025. Data were available for all U.S. states except Missouri and were obtained from the National Syndromic Surveillance Program (NSSP) [41]. The limited temporal coverage of RSV data necessitated the use of transfer learning approaches to extend model capabilities to this respiratory disease target.

### Official Influenza Hospitalizations Reports

The dataset comprised a weekly time series of influenza hospitalizations, spanning from July 3, 2021, to May 24, 2025. This data covered each U.S. state and the national level, as provided by the Centers for Disease Control and Prevention (CDC) for the FLUsight challenge [42]. The intermediate temporal coverage of influenza hospitalization data provided an additional target for validating transfer learning capabilities.

### Google Trends Digital Behavioral Signals

We utilized weekly datasets from the Google Trends API, incorporating search terms related to influenza and respiratory diseases to capture population-level behavioral changes that precede clinical case reporting. Key search terms included specific disease identifiers such as ‘influenza b’, ‘rsv’, and ‘flu virus’. Additionally, terms representing common symptoms of respiratory illnesses, such as ‘flu headache’, ‘rsv symptoms’, and ‘human temperature’, were incorporated to capture symptom-related search behavior. Furthermore, terms related to treatment and prevention, including ‘over the counter flu’, ‘tamiflu wiki’, and ‘flu shot’, were used to enhance the dataset’s comprehensiveness in capturing health-seeking behavior patterns. All search terms used can be found in the Supplementary Materials Box 1.

### Data Preprocessing and Temporal Alignment

Our data streams experience specific availability delays that required temporal adjustments for proper alignment. Google Trends data are available up to 6 days before the current date, necessitating a temporal shift where data reported at time *t* were shifted to time *t* + 6 to address the 6-day reporting delay. This adjustment ensures that digital behavioral signals maintain their predictive value without introducing forward-looking bias in real-time applications.

### Early Warning System Methodology

The early warning system integrates epidemiological surveillance data with digital behavioral signals to identify outbreak patterns 2–6 weeks before peak healthcare demand. The methodology consists of three key components: outbreak event detection, predictive signal selection, and ensemble voting for alarm generation.

### 4.1 Outbreak Event Detection

We developed distinct algorithms to identify outbreak onsets and peaks in both target surveillance data (for model training) and proxy signals (for prediction). For onset detection, we adapted a methodology previously developed by Kogan et al. [27] and Stolerman et al. [28]. Specifically, for each week *t* of our time series, we assessed exponential growth patterns over the preceding 6 weeks by computing a retrospective linear regression model to determine the multiplicative constant *λ*_*t*_ that maps case numbers from one week to the next (i.e., the coefficient of a lag-1 autoregressive model without intercept). This parameter *λ*_*t*_ serves as a proxy for the effective reproductive number *R*_*t*_, a standard epidemiological measure indicating whether an epidemic is growing (*λ*_*t*_ *>* 1) or declining (*λ*_*t*_ *<* 1).

The methodologies then diverged based on data type to address temporal bias constraints. For target surveillance data used in model training, we could apply forward-looking validation and confirmed an onset when periods of *λ*_*t*_ *>* 1 were followed by six consecutive weeks of exponential growth where *λ*_*t*_ *>* 1. For proxy signals like Google Trends used in prediction, we avoided forward-looking bias by identifying onsets as the sixth week of exponential growth windows where *λ*_*t*_ *>* 1, ensuring that predictive signals remained strictly historical.

Peak identification employs the scipy library’s peak detection algorithm [32] with three filtering criteria: minimum duration of 2.5 weeks, case counts exceeding the 75th percentile of previous seasons, and temporal separation of at least 20 weeks between peaks (unless a subsequent peak is higher).

This approach inherently involves 1–3 week delays since peak confirmation requires observing both the rise and subsequent decline in disease activity.

### 4.2 Predictive Signal Selection and Ensemble Voting

For each target location, we perform correlation-based feature selection to identify the most predictive proxy signals from Google search trends and neighboring state surveillance data. Only proxies showing strong historical association (Pearson correlation *>* 0.7) with target outbreak patterns during training periods are retained for the ensemble. This threshold was determined during the System Development and Retrospective Validation phase (2010–2020), where we systematically evaluated correlation cutoffs ranging from 0.5 to 0.9 and found 0.7 to yield the best trade-off between sensitivity and specificity. To provide a supporting validation of this feature selection, we applied Granger causality tests to the retained proxies. Across rolling three-year windows, 92.7% of the selected terms were Granger-significant at least once (Supplementary Fig. S108), and 56.0% remained significant after adjustment for multiple comparisons using the false discovery rate (FDR; [43]) (Supplementary Fig. S107). This provides a heuristic but meaningful check that the selected features align with temporal predictive structure in the data.

The ensemble voting mechanism aggregates selected proxy signals based on their historical performance in predicting outbreak events. Each proxy is evaluated using standard classification metrics: true positives (proxy events preceding target outbreaks by ≤ 6 weeks), false positives (proxy events not followed by target outbreaks within 6 weeks), and false negatives (missed target outbreaks). The decision threshold *τ* for triggering early warning alarms is calibrated as the lowest number of correct predictions achieved across the three most recent training outbreaks:

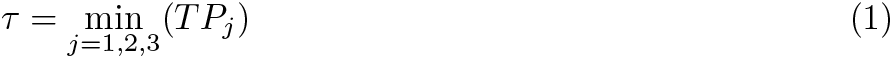

where *TP*_*j*_ represents the number of true positive votes from the three most recent training outbreaks. This conservative approach ensures high confidence in alarm generation while minimizing false alerts. The choice of a three-season training window was motivated by its use in similar early warning and machine learning frameworks [19, 27, 28] and was further validated during the System Development and Retrospective Validation phase (2010–2020), where we empirically tested training horizons ranging from one to five seasons and found three seasons provided the most stable balance between sensitivity and specificity.

At each time point, the early warning system calculates the number of proxy signals that have activated within the previous three weeks. When this count exceeds the threshold *τ*, the system issues a warning predicting an outbreak event within the following six weeks. Figures 3 and 4 illustrate this process for ILI onset prediction in Illinois and peak prediction in Massachusetts, respectively, showing how individual proxy signals aggregate to trigger early warning alarms.

**Fig. 3:**
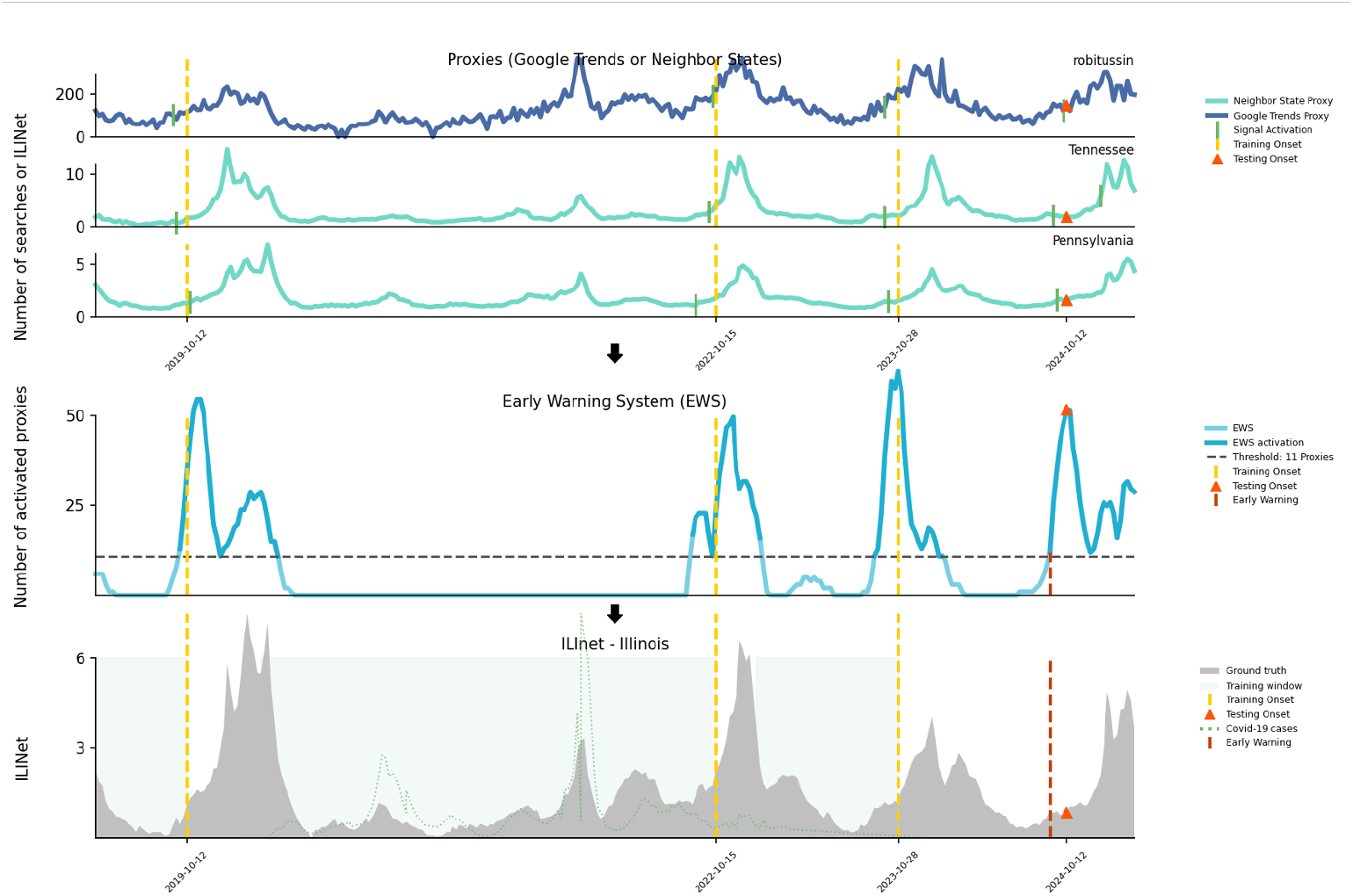
Early warning system for predicting ILI onset in Illinois. The top three panels show proxy signals from Google Trends (e.g., “robittussin”) and neighboring states (Tennessee and Pennsylvania). Green markers indicate signal activation events. The fourth panel displays the early warning system (EWS) output, with the number of activated proxies (light blue), and a dashed threshold line (11 proxies) above which an early warning is triggered (dark red dashed vertical lines). The bottom panel shows the official ILINet surveillance data for Illinois (gray shaded area) along with training and testing onsets (yellow and orange vertical lines, respectively), and overlaid COVID-19 cases (green dotted line). The EWS successfully triggers an early warning before the ILI onset for the 2024-2025 season.

**Fig. 4:**
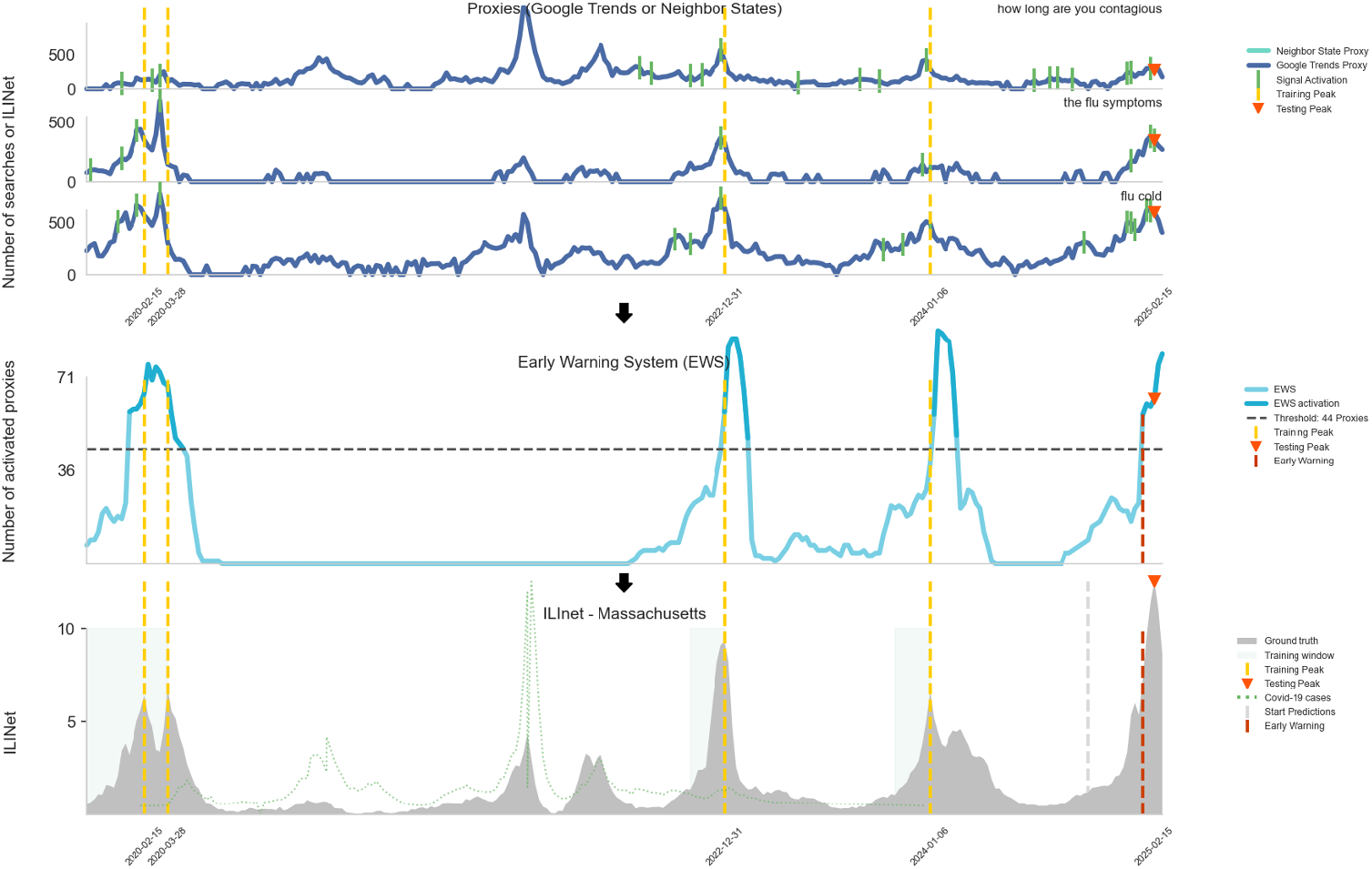
Early warning system for predicting ILI peak in Massachusetts. The top three panels present proxy signals including Google search queries (e.g., “how long are you contagious”, “flu symptoms”, “flu cold”) and data from neighboring states. Green ticks indicate proxy signal activation. The fourth panel shows the number of activated proxies used by the early warning system (EWS), with early warnings triggered once the activation threshold (44 proxies, dashed line) is exceeded. The bottom panel illustrates ILINet data for Massachusetts (gray shaded area) and the timing of training, testing, and early warnings. The system accurately anticipates peak ILI activity during the 2024–2025 season.

### 4.3 Transfer Learning for Influenza Hospitalizations and RSV Activity

To extend the framework to influenza hospitalizations and RSV activity despite limited historical data, we employed transfer learning by directly applying all model parameters, feature selection criteria, and methodological components from the trained ILI system (2010–2020). This approach leverages the assumption that digital behavioral signals exhibit similar patterns across respiratory diseases, allowing rapid deployment to new surveillance targets with minimal training data. The only component requiring adjustment for each new disease target was the alarm threshold (*τ*), which was fine-tuned using the limited available training data to optimize detection performance while maintaining appropriate false alarm rates.

#### Validation Strategy

A critical feature of our validation approach is strict temporal integrity: at no point did future data influence past predictions. This mirrors real-world deployment conditions where only historical data informs forecasts. We employed three validation phases for ILI: retrospective validation (2014–2020) for hyperparameter tuning and initial performance assessment, prospective testing (2022–2024) for out-of-sample validation, and real-time deployment (2024–2025) for operational performance evaluation. For influenza hospitalizations and RSV activity we employed a prospective testing (2024–2025) for out-of-sample validation.

The training strategy uses rolling three-season windows to capture evolving disease dynamics while maintaining temporal relevance. For each prediction target, the system trains on the three most recent historical seasons and predicts the subsequent season’s outbreak patterns. The 2024–2025 season data was completely withheld during development to provide an unbiased assessment of real-world performance.

#### Performance Metrics

We categorized system performance using temporal alignment between predicted and observed outbreak events:

– **Early Warnings:** alarms triggered 1–6 weeks before confirmed outbreak events
– **Synchronous Warnings:** alarms triggered during the same week as confirmed outbreak events
– **Late Warnings:** alarms triggered up to 3 weeks after confirmed outbreak events
– **Early-Late Warnings:** late warnings that still detected peaks before the algorithmic peak detection would have confirmed them
– **Soft Warnings:** cases where 75% of required proxy signals activated within 6 weeks of an outbreak, but the alarm threshold was not exceeded
– **Missed Outbreaks:** confirmed outbreak events with no corresponding alarm within the evaluation window

We distinguished between two types of false alarms to better understand system behavior:

– **True False Alarms:** alarms not followed by any detectable increase in disease activity within 6 weeks
– **Warning Signals:** alarms not followed by confirmed outbreaks but associated with measurable increases in disease activity, indicating elevated transmission risk

Performance evaluation emphasized positive predictive value (PPV) to quantify alarm reliability and detection sensitivity to assess outbreak capture rates. This dual focus ensured the system balanced early warning capabilities with practical utility for public health decision-making. Rather than traditional ROC curves, our evaluation focuses on aggregated detection rates and positive predictive values. This choice reflects our system’s architecture: dynamically retrained, location-specific classifiers that update as new data becomes available, rather than a single static global model. ROC curves for each classifier (with multiple models per location and time period) would not provide an interpretable summary of overall system performance. Our reported metrics—detection sensitivity, PPV, and lead times—directly quantify the system’s operational utility for public health decision-making.

## Acknowledgments

We thank Mr. Xi Chen for his valuable contributions to the implementation of the Granger causality analysis and the development of corresponding visualizations.

## Contributions

R.G.G., L.C., and M.S. designed research; R.G.G. and L.C. performed research; R.G.G. developed and implemented analytic tools; R.G.G., L.C. and M.S. analyzed data; R.G.G. and M.S. wrote the first draft of the paper; all authors reviewed the research, contributed to the manuscript, and approved the final version of the manuscript; and M.S. supervised the research.

## Data Availability

A sample of data and code used in this article can be found at *github*.*com/MIGHTE* − *lab/Respiratory EWS* − *Sample* and will be released to the public upon article publication. The original data is provided by the United States Centers for Disease Control and Prevention through data.cdc.gov and the google searches are provided by Google through trends.google.com.

## Funding

This manuscript was made possible in part by cooperative agreement CDC-RFA-FT-23-0069 from the CDC’s Center for Forecasting and Outbreak Analytics. Its contents are solely the responsibility of the authors and do not necessarily represent the official views of the Centers for Disease Control and Prevention.

## Competing interests

The authors have declared no competing interest.

## Notes

### Funding Statement

This manuscript was made possible in part by cooperative agreement CDC-RFA-FT-23-0069 from the CDCs Center for Forecasting and Outbreak Analytics. Its contents are solely the responsibility of the authors and do not necessarily represent the official views of the Centers for Disease Control and Prevention.

