## Supplementary Materials for "A Prospective Real-time Early Warning System to Anticipate Onsets and Peaks of Respiratory Diseases Outbreaks at the State Level in the U.S. A Transfer Learning Approach Leveraging Digital Traces"

#### Variability of Onsets and Peaks

Using historical state-level ILI onset weeks, we clustered states by timing variability (Supplementary Fig. 5). Low variability states (13 states) show onset ranges of 3–7 weeks with a median of 5 weeks, moderate variability states (22 states) exhibit ranges of 8–13 weeks with a median of 10 weeks, and high variability states (14 states) demonstrate ranges of 14–20 weeks with a median of 16.5 weeks. This means that while some states begin flu season within approximately one month most years, the majority experience 2–4 months of temporal spread in onset timing.

Peak timing uncertainty presents an even greater challenge (Supplementary Fig. 6). Low variability states (12 states) show peak ranges of 5–6 weeks, moderate variability states (22 states) exhibit ranges of 7–12 weeks, and high variability states (15 states) demonstrate ranges of 13–20 weeks. The dispersion in peak timing significantly exceeds that of onsets (mean range 14.39 vs. 10.67 weeks across states), underscoring that identifying peaks in real-time presents a greater challenge than onset detection. This substantial historical uncertainty makes traditional forecasting approaches unreliable and highlights the critical need for data-driven early warning systems that can operate independently of historical timing patterns.

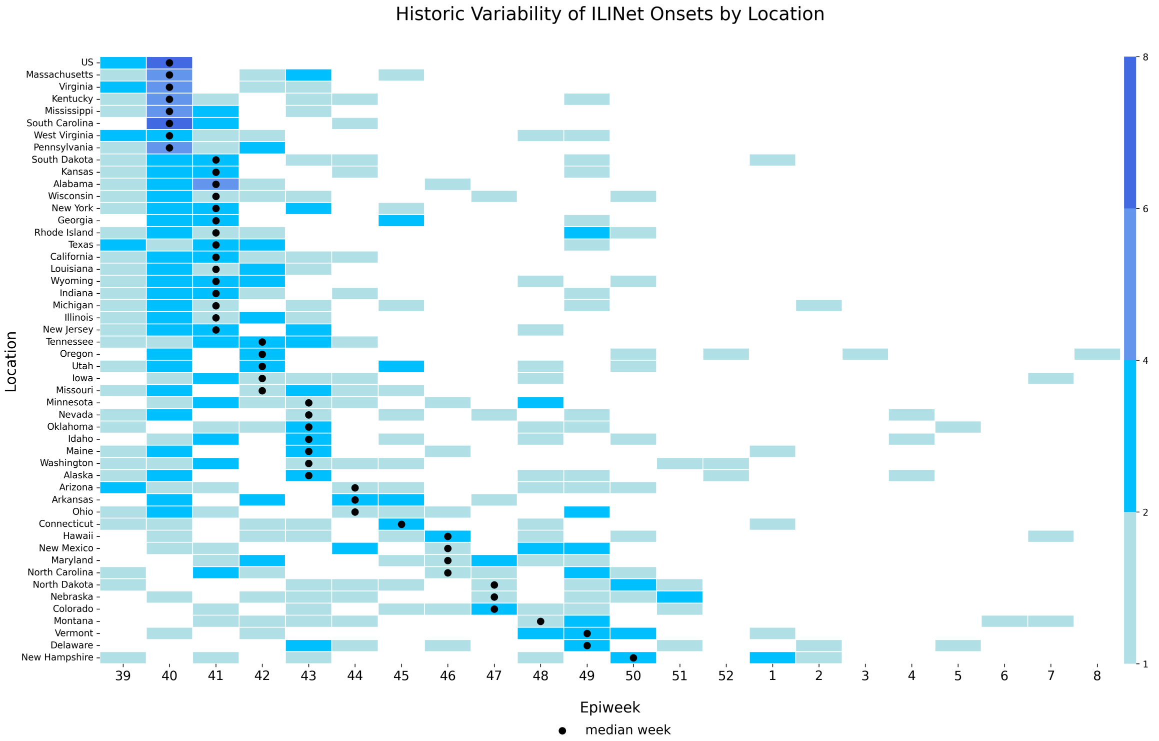


**Fig. S1**: **Variability of ILI Onsets at state and national levels in the U.S. from 2010-2020.** Each cell indicates the number of seasons in which a given location experienced onset during a particular epidemiological week. Darker shades represent higher frequencies, while black dots mark the median onset week for each location. This figure highlights substantial spatiotemporal heterogeneity in onset timing, illustrating both early and late season dynamics across states.

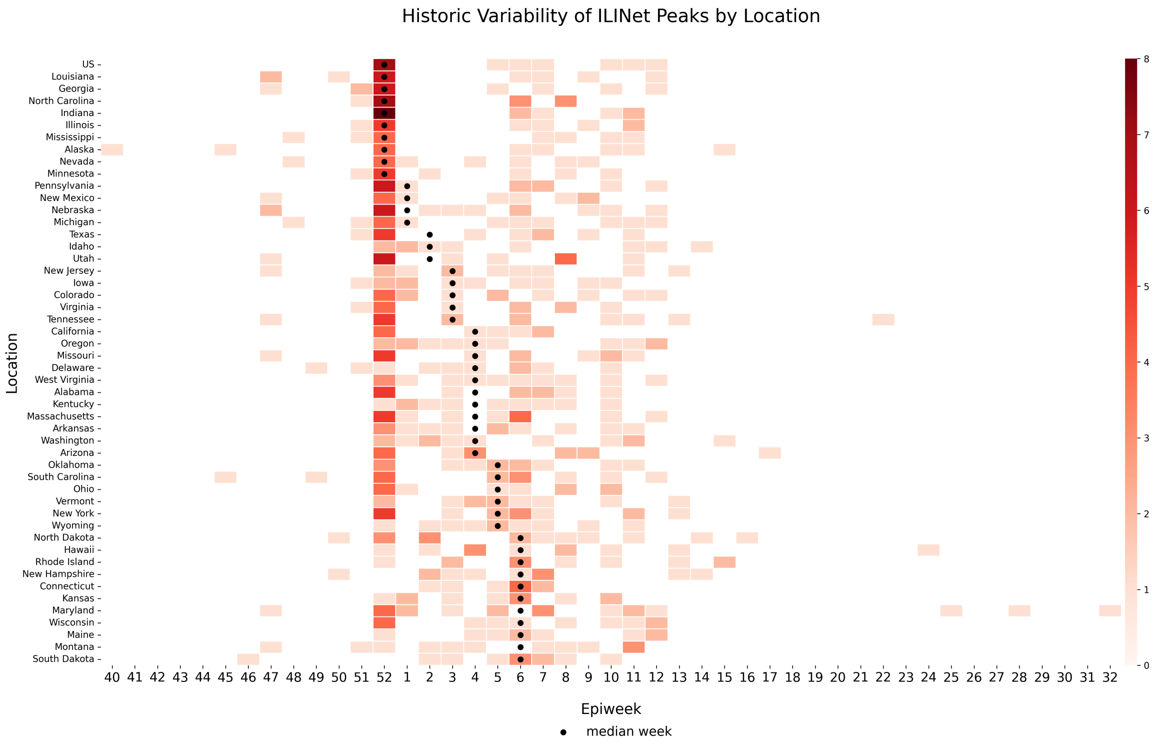


**Fig. S2**: **Variability of ILI Peaks at state and national levels in the U.S. from 2010-2020.** Each cell indicates the number of seasons in which a given location experienced peaks during a particular epidemiological week. Darker shades represent higher frequencies, while black dots mark the median peak week for each location. This figure highlights substantial spatiotemporal heterogeneity in peak timing, illustrating both early and late season dynamics across states.

#### Google Trends Used

##### Box 1: Google Search Terms

anosmia, chest pain, chest tightness, cold, cold symptoms, cold with fever, contagious flu, cough, cough and fever, cough fever, covid, covid nhs, covid symptoms, covid-19, covid-19 who, dry cough, feeling exhausted, feeling tired, fever, fever cough, flu and bronchitis, flu complications, how long are you contagious, how long does covid last, how to get over the flu, how to get rid of flu, how to get rid of the flu, how to reduce fever, influenza, influenza b symptoms, isolation, joints aching, loss of smell, loss smell, loss taste, nose bleed, oseltamivir, painful cough, pneumonia, pneumonia and have the flu, quarantine, remedies for the flu, respiratory flu, robitussin, robitussin cf., robitussin cough, rsv, runny nose, sars-cov 2, sars-cov-2, sore throat, stay home, strep, strep throat, symptoms of bronchitis, symptoms of flu, symptoms of influenza, symptoms of influenza b, symptoms of pneumonia, symptoms of rsv, tamiflu dosage, tamiflu dose, tamiflu drug, tamiflu generic, tamiflu side effects, tamiflu suspension, tamiflu while pregnant, tamiflu wiki, tessalon

#### Retrospective EWS Results

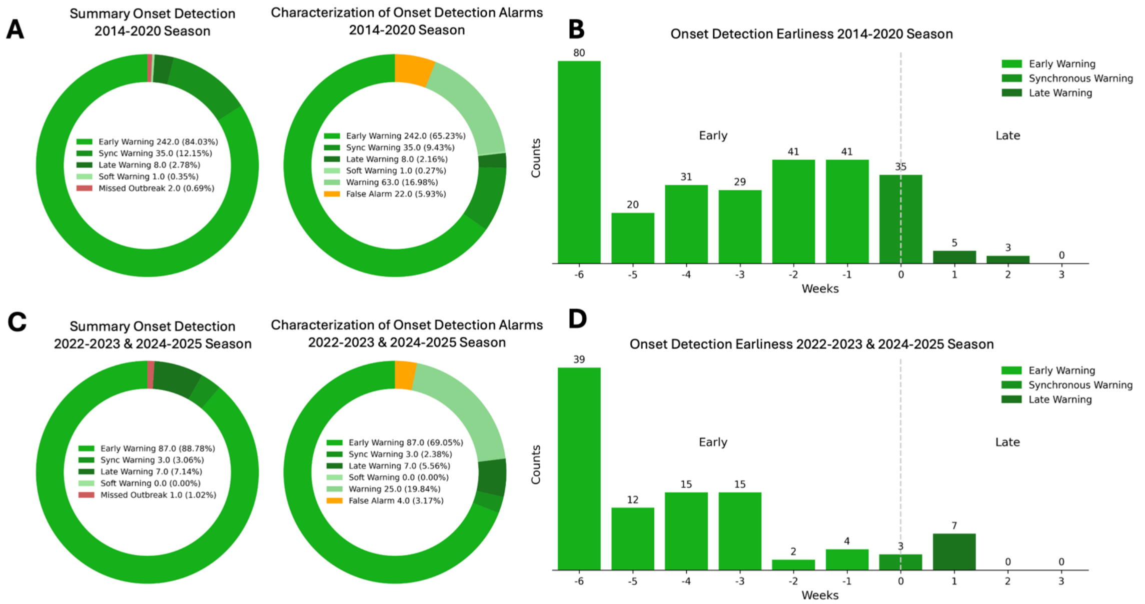


**Fig. S3**: **Onset Detection Performance During Retrospective Validation and Retrospective Experiment. Top Row: Validation Period (2014-2020 Seasons). A)** Left: Summary of onset detection outcomes across U.S. states using the Early Warning System (EWS). Most onsets were detected early (94.9%), with minimal synchronous (1.2%) and late warnings (1.6%). Only 0.3% of outbreaks were missed. Right: State-level quality of detection alarms, showing the proportion of true detections and false alarms. While early detections dominate, a small number of false alarms (5.9%) are also observed.**B)** Distribution of onset detection earliness during the validation period. The majority of detections occurred 3 to 6 weeks prior to the onset, peaking at 6 weeks in advance, indicating strong predictive lead time. **Bottom Row: Retrospective Experiment (2022–2023 and 2023–2024 Seasons). C)** Left: Summary of onset detection performance during the two most recent seasons. Early detections remain high (86.8%), with small increases in synchronous (6.1%) and late warnings (3.7%). Only one outbreak was missed. Right: Alarm characterization highlights continued accuracy, with early warnings leading (89.5%) and a false alarm rate of 3.7%. **D)** Distribution of detection earliness in the retrospective test phase. Most detections occurred 3 to 6 weeks before onset, with a strong peak at 6 weeks. While performance is slightly more variable than the validation period, early warning capacity remains robust.

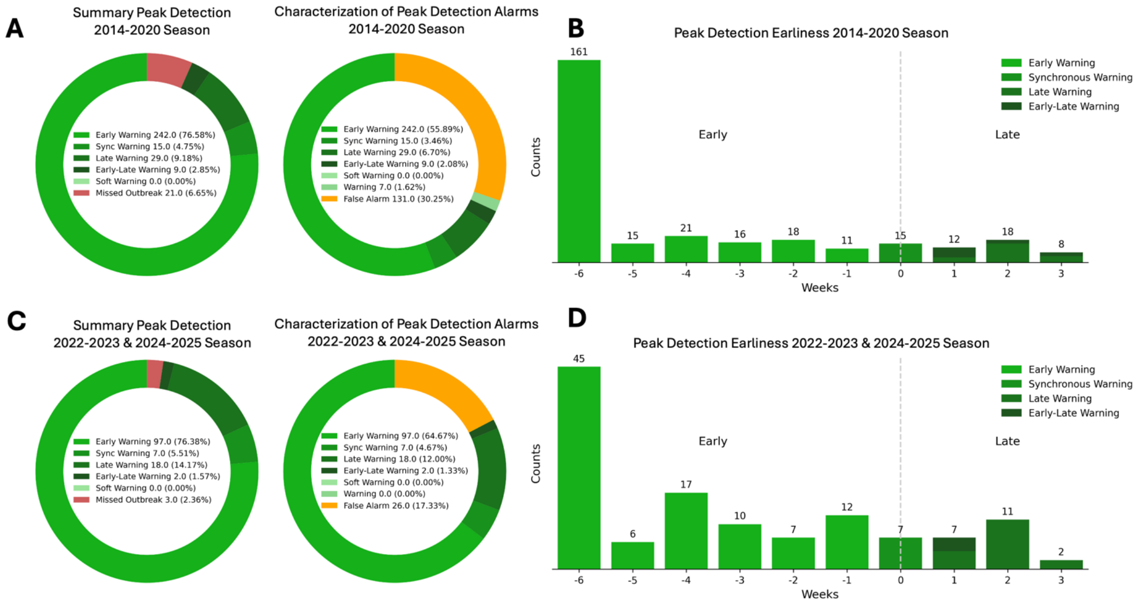


**Fig. S4**: **Onset Detection Performance During Retrospective Validation and Retrospective Experiment. Top Row: Validation Period (2014–2020 Seasons). A)** Left: Summary of peak detection outcomes across U.S. states using the Early Warning System (EWS). The majority of peaks were detected early (76.6%), with smaller proportions of synchronous (4.7%), late (4.3%), and early-late warnings (12.4%). A small number of outbreaks were missed (0.7%). Right: State-level quality of detection alarms, showing a high rate of early warnings (85.4%) and a false alarm rate of 6.3%. This indicates a strong balance between sensitivity and precision during the validation period. **B)** Distribution of peak detection earliness. Most detections occurred 3 to 6 weeks prior to the peak, with a pronounced concentration at 6 weeks. The early warning system consistently offered substantial lead time. **Bottom Row: Retrospective Experiment (2022-2023 and 2023-2024 Seasons). C)** Left: Summary of peak detection performance during the two most recent seasons. Early warnings remained high (76.4%), while synchronous, late, and early-late warnings comprised 5.4%, 11.4%, and 4.5% respectively. Three outbreaks were missed (3.6%). Right: Alarm classification continues to show high early-warning effectiveness (82.6%) with a slight reduction in precision, as indicated by a false alarm rate of 4.3%. **D)** Distribution of detection earliness in the retrospective test phase. While most detections still occurred early, the distribution was more spread, with notable counts even in the late warning range (1–3 weeks post-peak). Despite this variability, the early detection lead time was largely preserved.

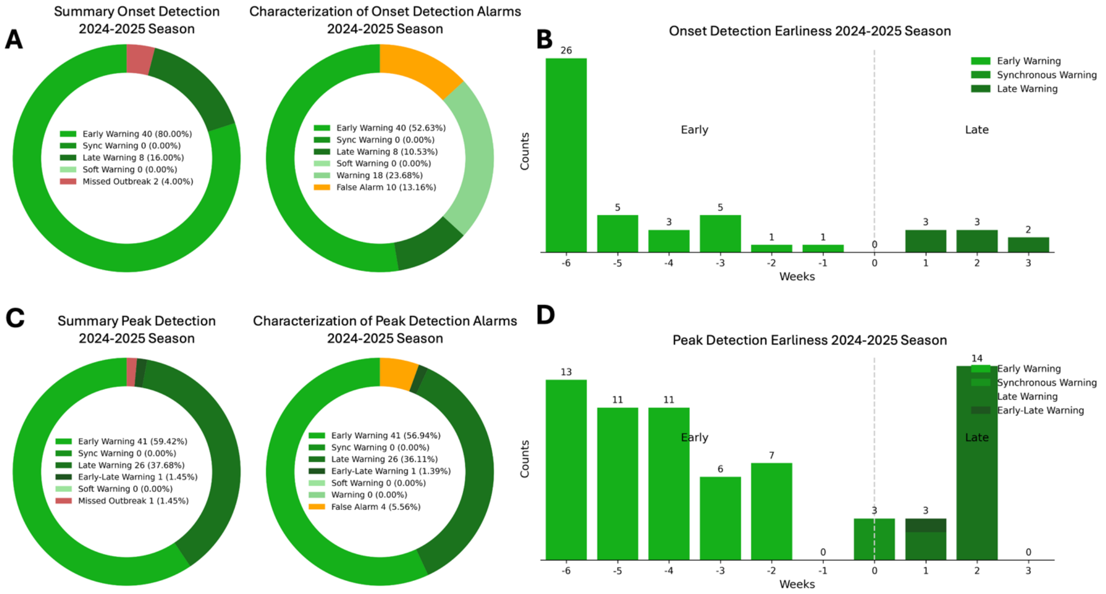


**Fig. S5**: **Performance of Influenza Early Warning System for the 2024–2025 Season. Top Row: Onset Detection. A)** Left: Summary of onset detection outcomes across U.S. states using the Early Warning System (EWS). Most onsets were detected early (80%), followed by late (16%) and missed outbreaks (4%). Only 2 onsets were missed. Right: Alarm classification based on onset detection, showing that early warnings comprised the majority (52.63%), followed by late warnings (10.53%), and false alarms (13.16%). **B)** Distribution of onset detection earliness. The majority of early detections occurred 3 to 6 weeks before onset, with a peak at 6 weeks prior. There were not many late warnings supporting the system's ability to provide timely alerts despite variability in signal dynamics. **Bottom Row: Peak Detection. C)** Left: Summary of peak detection outcomes, showing that over half of peaks were detected early (59.42%), followed by late warnings (37.68%), and early-late warnings (1.45%). Only one peak was missed. **D)** Right: Distribution of detection earliness for peak events. Most early warnings occurred 3 to 6 weeks in advance of the peak, with late warnings concentrated around 2 weeks post-peak.

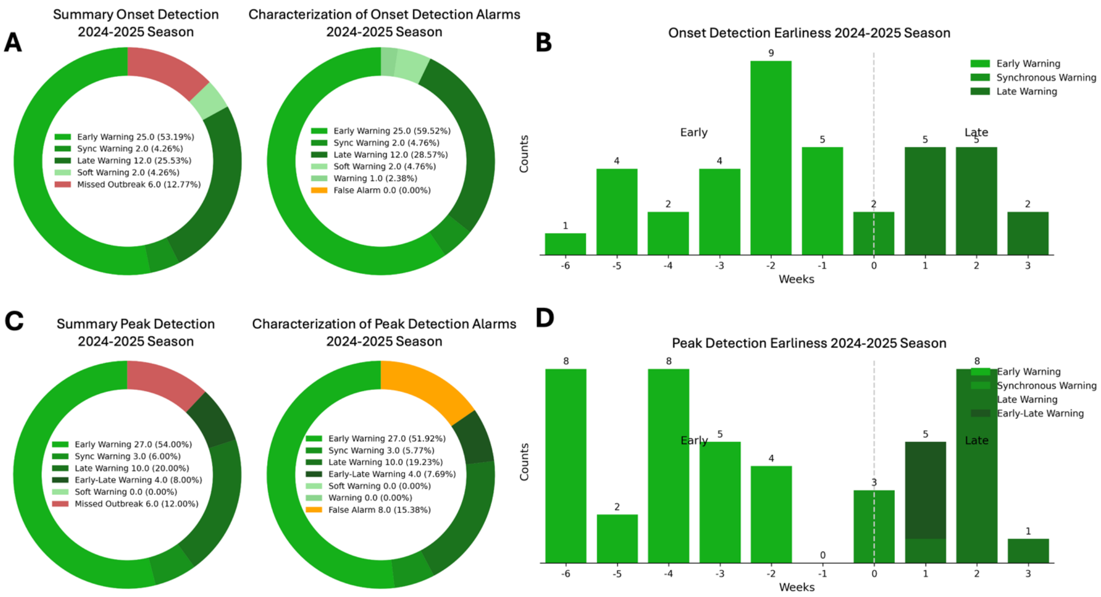


**Fig. S6**: **Performance of RSV Early Warning System for the 2024–2025 Season. Top Row: Onset Detection. A)** Left: Summary of onset detection outcomes across U.S. states using the Early Warning System (EWS). Most onsets were detected early (53.2%), followed by late (25.5%) and missed outbreaks (12.8%). Synchronous and soft warnings accounted for a smaller proportion (4.3% each). Right: Alarm classification based on onset detection, showing that early warnings comprised the majority (59.5%). No false alarms were recorded during this period. **B)** Distribution of onset detection earliness. The majority of early detections occurred 1 to 3 weeks before onset, with a peak at 2 weeks prior. Late warnings were most frequent 1–2 weeks after onset, supporting the system's ability to provide timely alerts despite variability in signal dynamics. **Bottom Row: Peak Detection. C)** Left: Summary of peak detection outcomes, showing that over half of peaks were detected early (54%), with smaller shares of synchronous (6%), late (20%), and early-late warnings (8%). A small number of peaks were missed (12%), with no soft warnings recorded. **D)** Right: Distribution of detection earliness for peak events. Most early warnings occurred 3–6 weeks in advance of the peak, with late warnings concentrated around 1–2 weeks post-peak.

#### Onset Examples 2024-2025 Real-Time Predictions

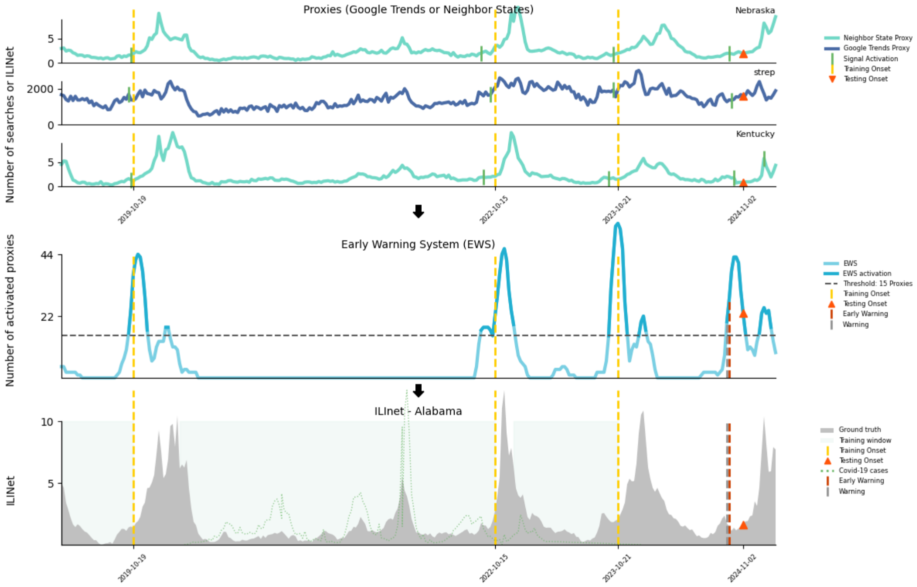


**Fig. S7**: Onset detection for Alabama during the 2024-2025 season

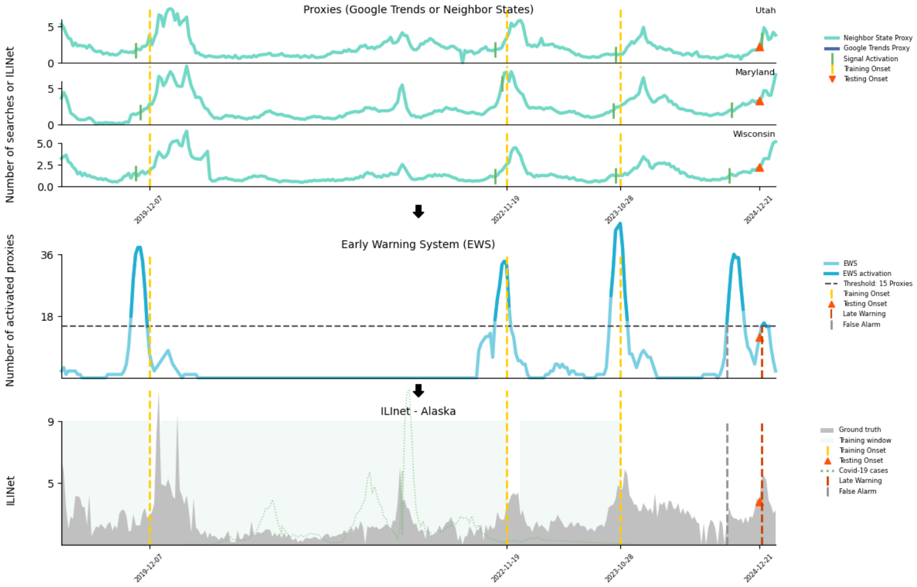


**Fig. S8**: Onset detection for Alaska during the 2024-2025 season

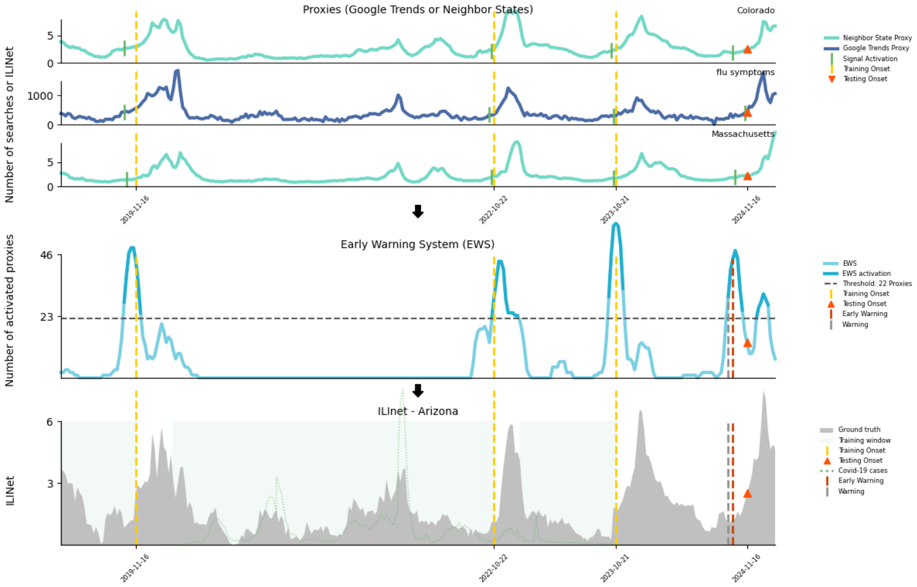

**Fig. S9**: Onset detection for Arizona during the 2024-2025 season

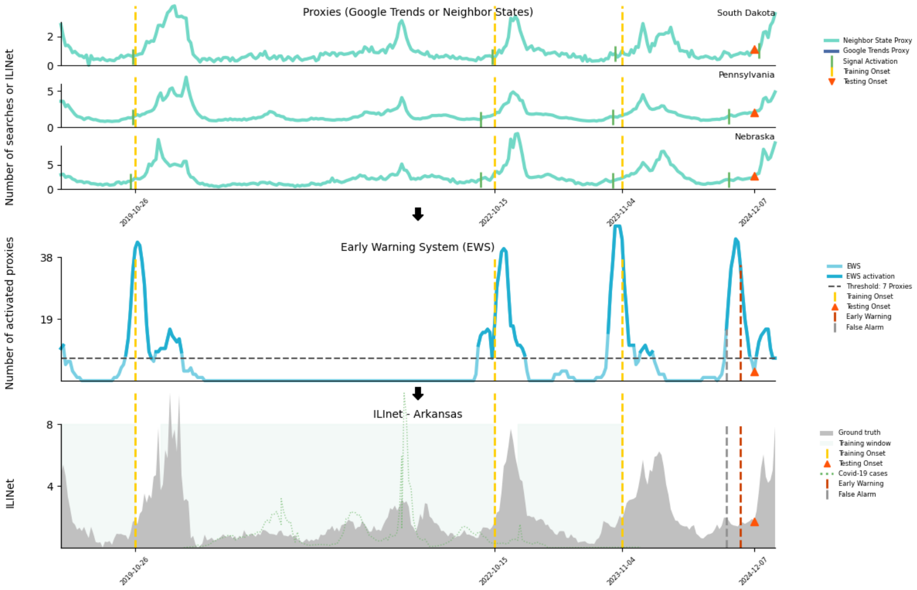

**Fig. S10**: Onset detection for Arkansas during the 2024-2025 season

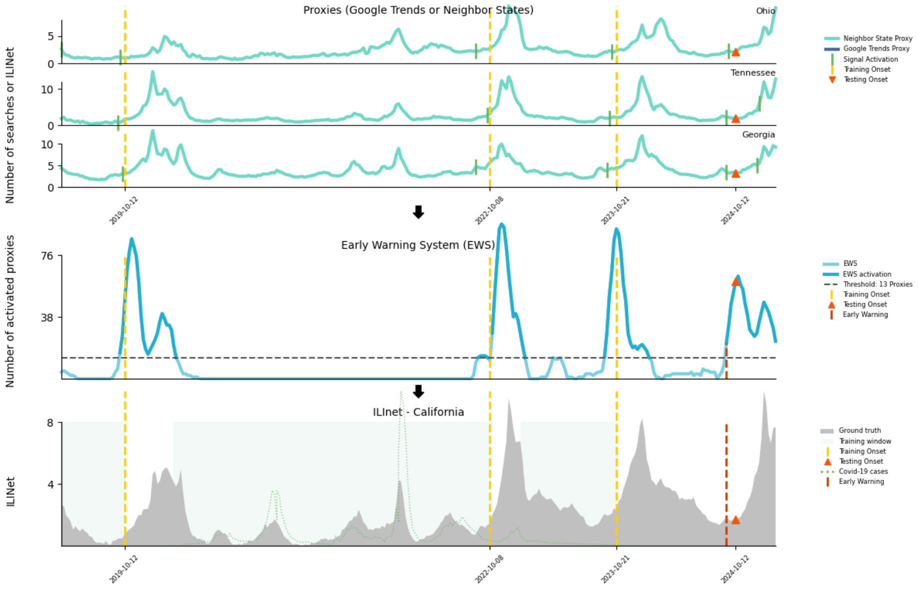


**Fig. S11**: Onset detection for California during the 2024-2025 season

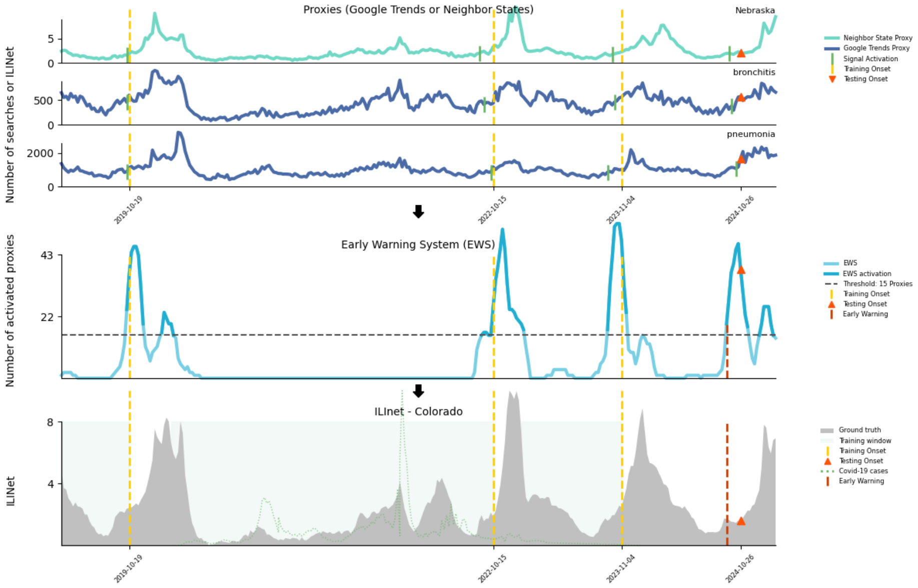


**Fig. S12**: Onset detection for Colorado during the 2024-2025 season

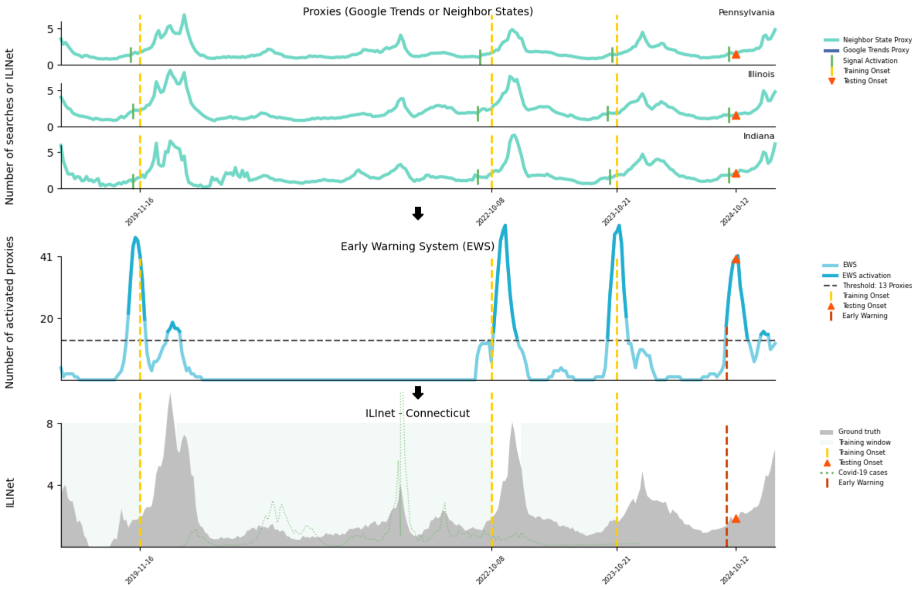


**Fig. S13**: Onset detection for Connecticut during the 2024-2025 season

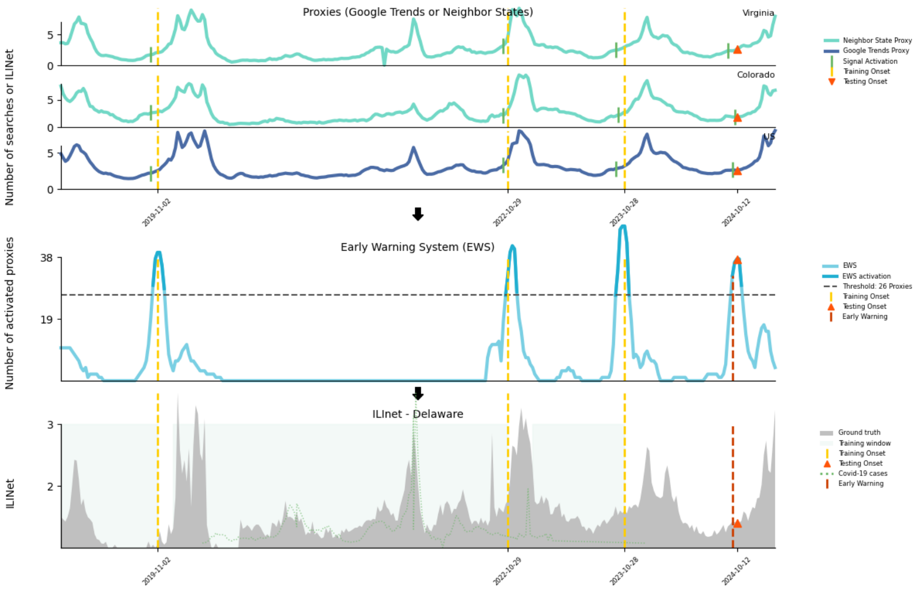


**Fig. S14**: Onset detection for Delaware during the 2024-2025 season

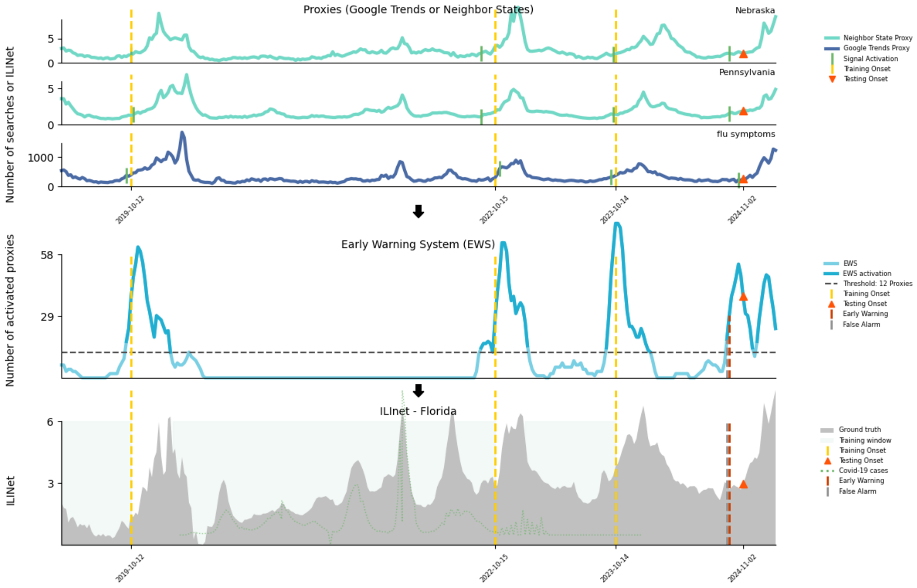


**Fig. S15**: Onset detection for Florida during the 2024-2025 season

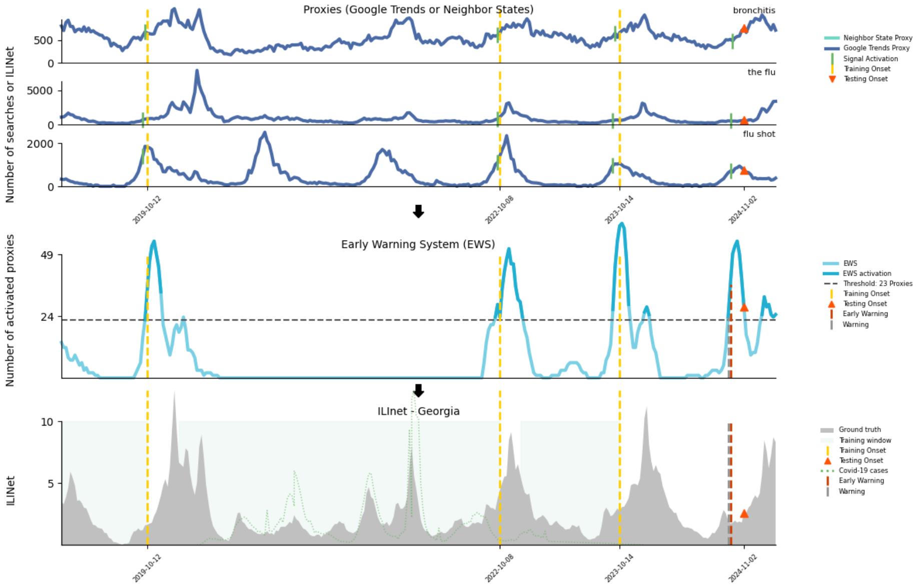


**Fig. S16**: Onset detection for Georgia during the 2024-2025 season

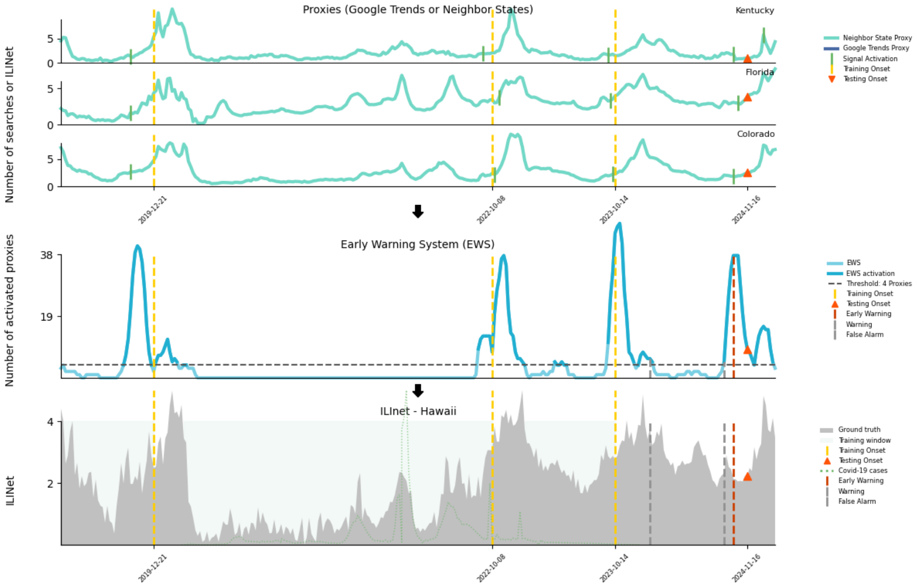


**Fig. S17**: Onset detection for Hawaii during the 2024-2025 season

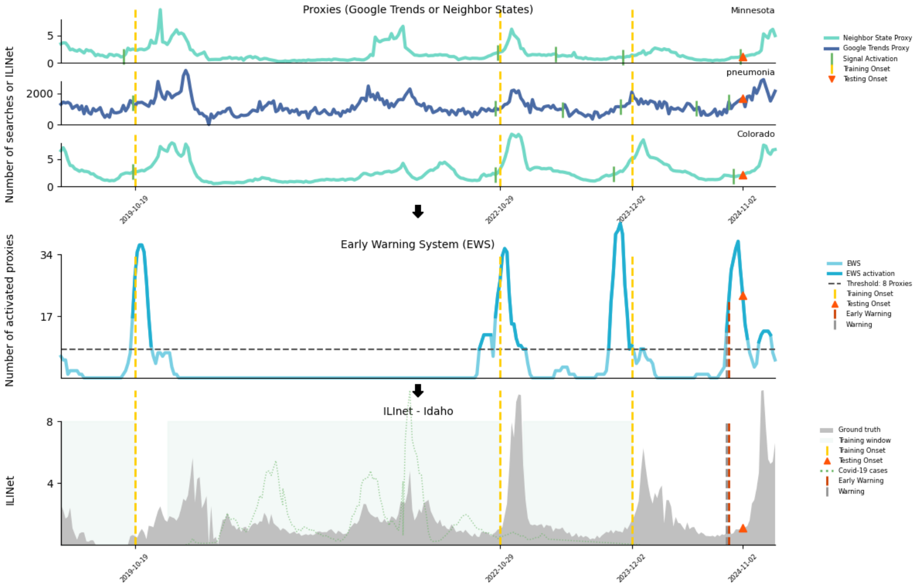


**Fig. S18**: Onset detection for Idaho during the 2024-2025 season

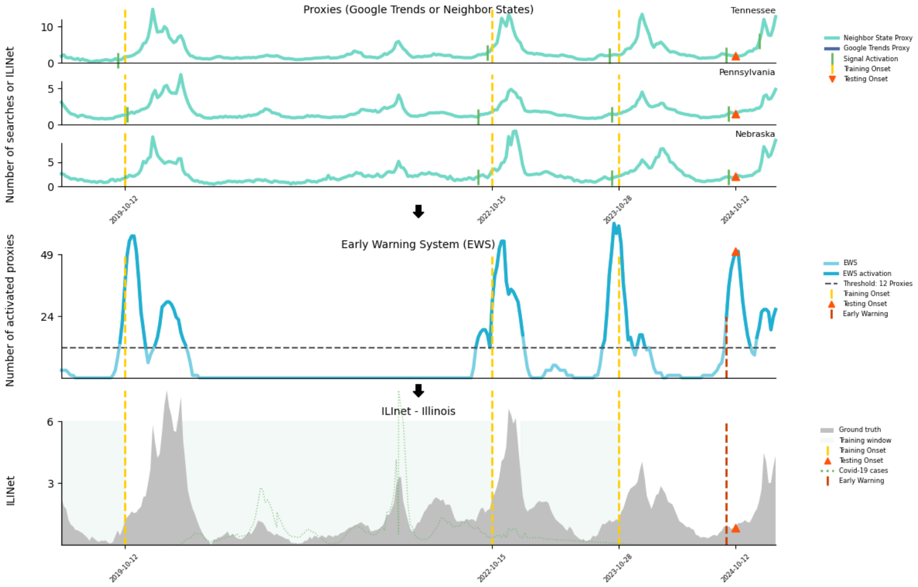


**Fig. S19**: Onset detection for Illinois during the 2024-2025 season

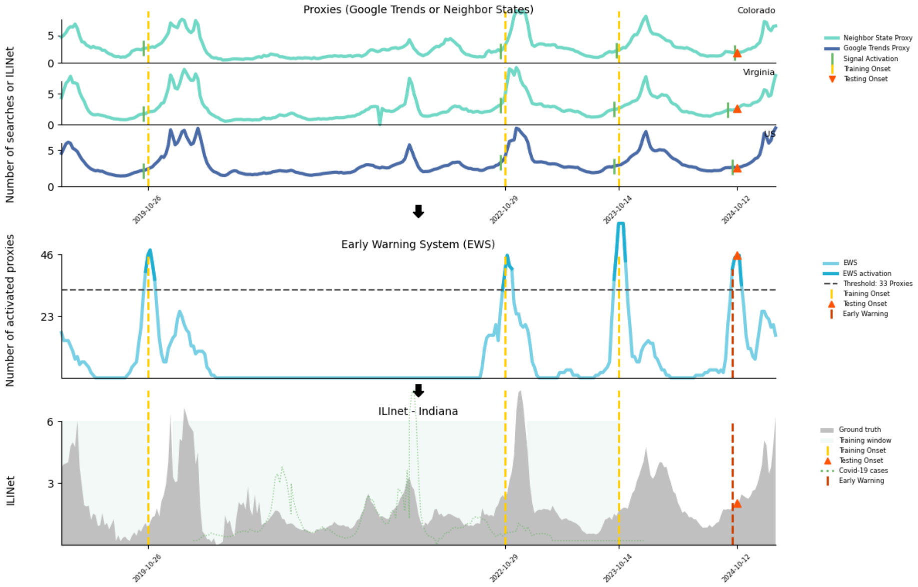


**Fig. S20**: Onset detection for Indiana during the 2024-2025 season

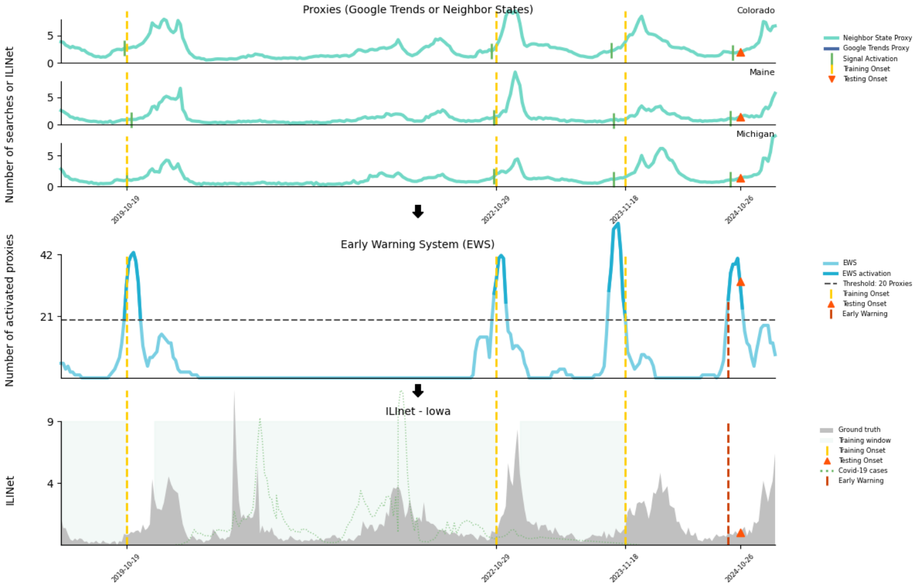


**Fig. S21**: Onset detection for Iowa during the 2024-2025 season

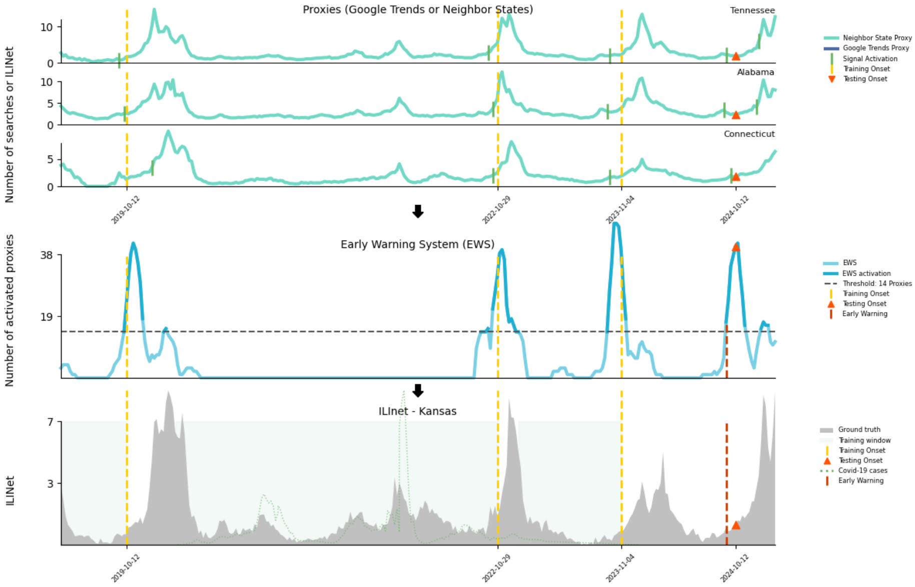


**Fig. S22**: Onset detection for Kansas during the 2024-2025 season

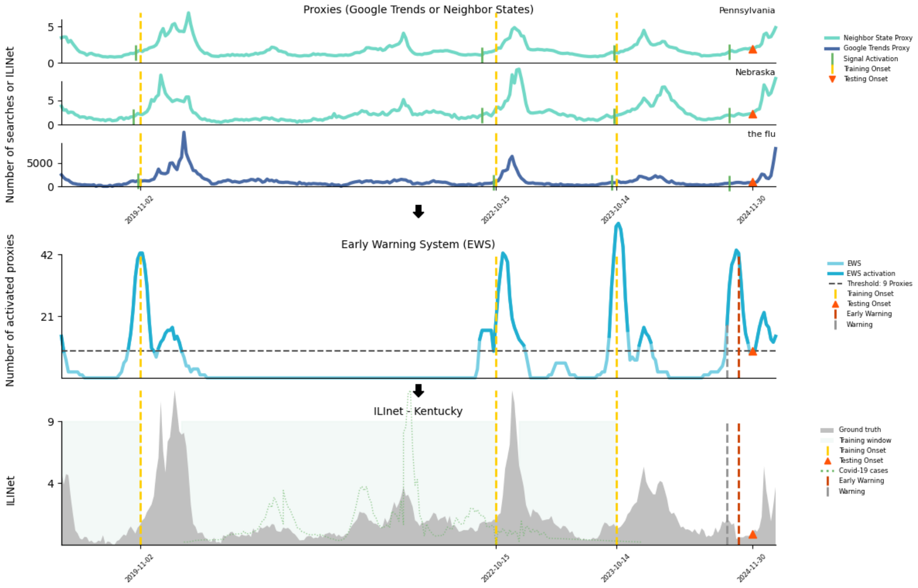


**Fig. S23**: Onset detection for Kentucky during the 2024-2025 season

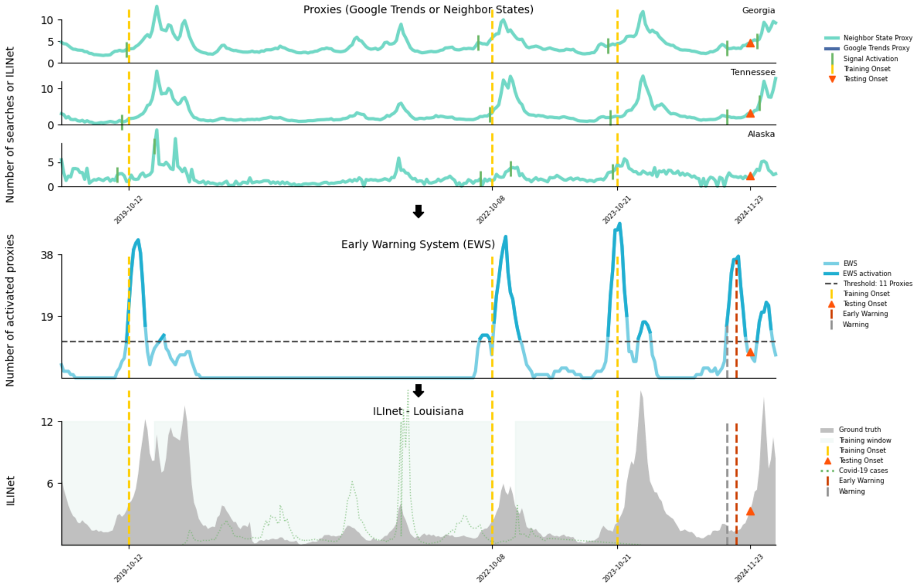


**Fig. S24**: Onset detection for Louisiana during the 2024-2025 season

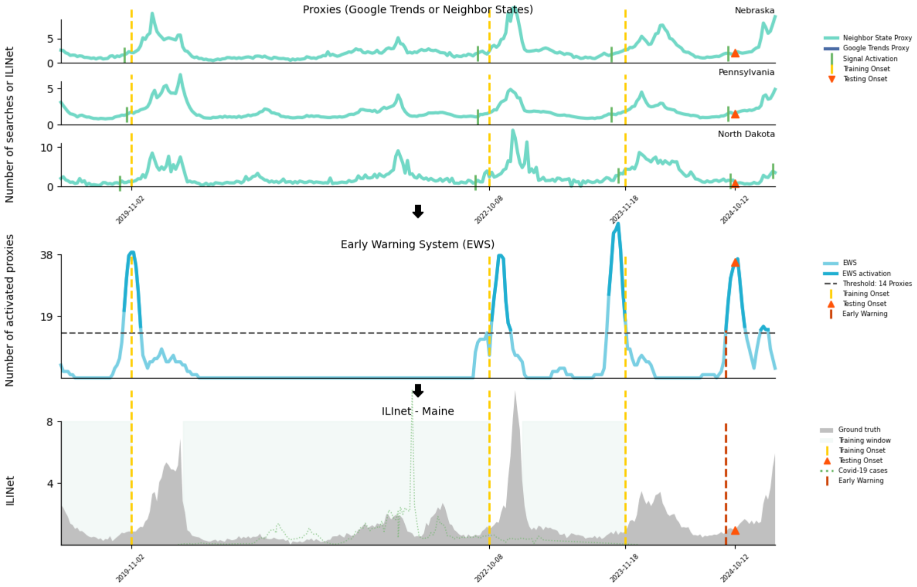


**Fig. S25**: Onset detection for Maine during the 2024-2025 season

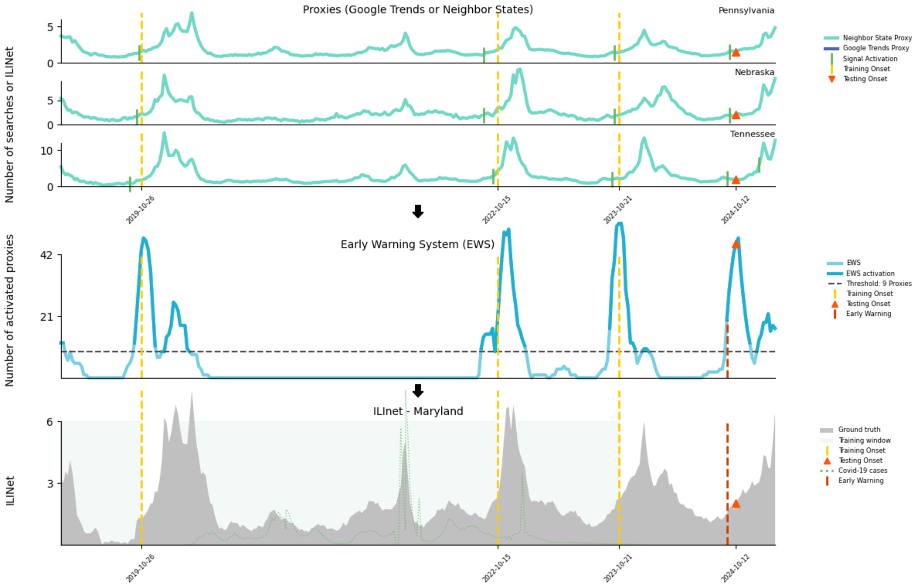


**Fig. S26**: Onset detection for Maryland during the 2024-2025 season

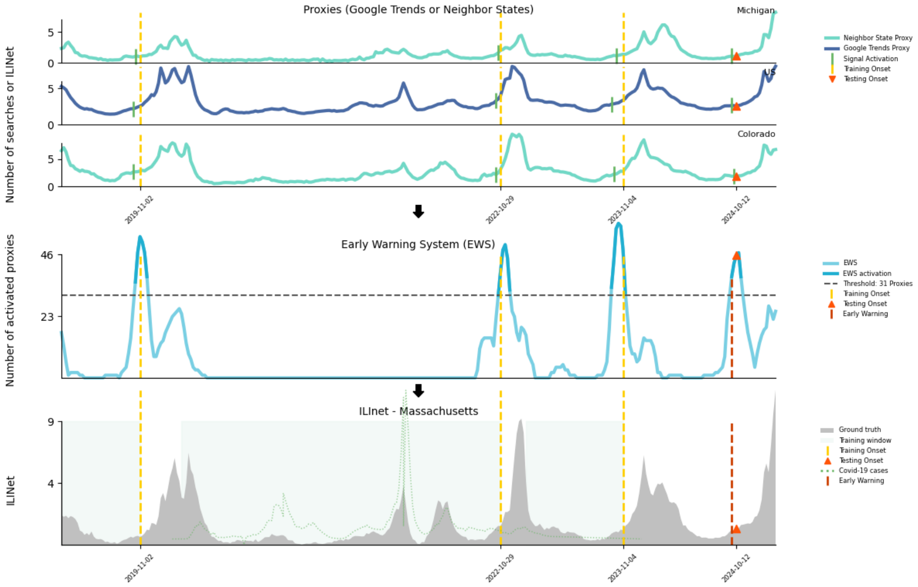


**Fig. S27**: Onset detection for Massachusetts during the 2024-2025 season

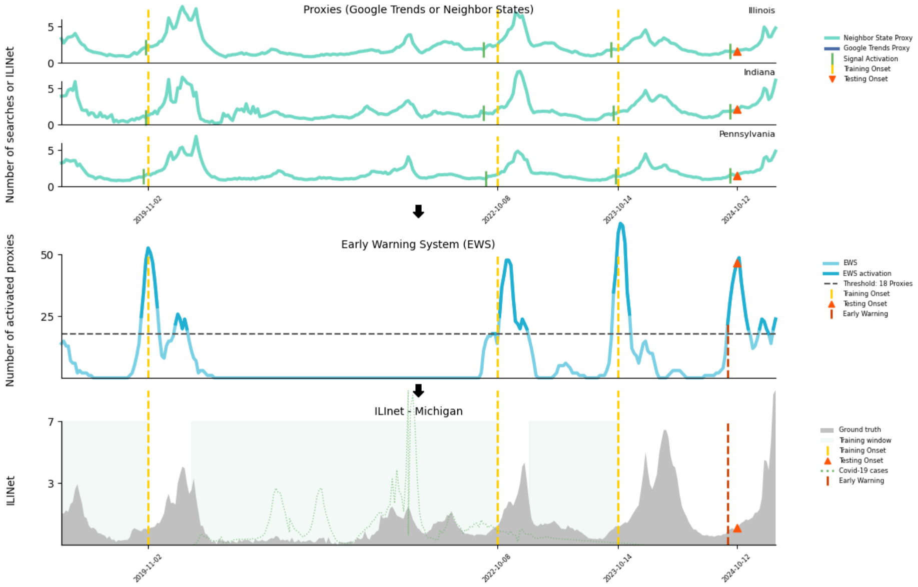


**Fig. S28**: Onset detection for Michigan during the 2024-2025 season

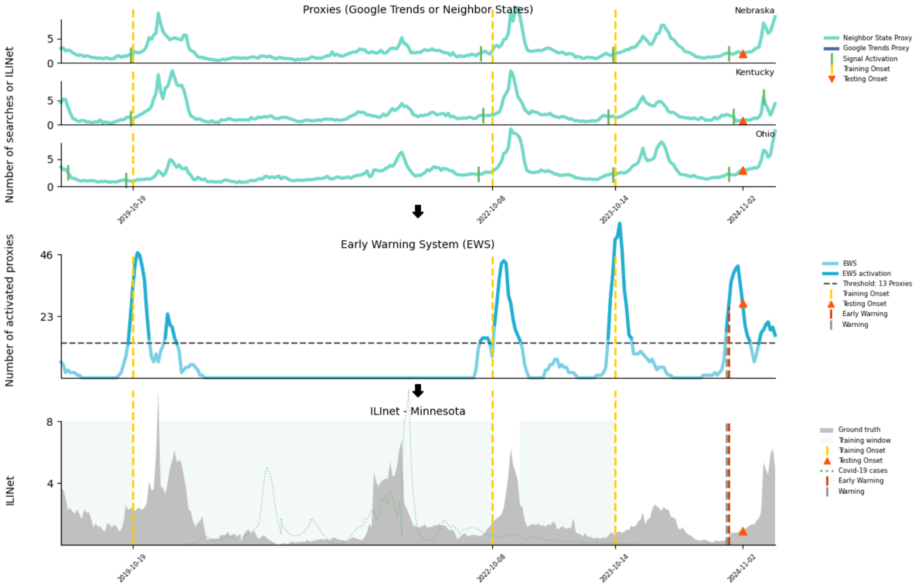


**Fig. S29**: Onset detection for Minnesota during the 2024-2025 season

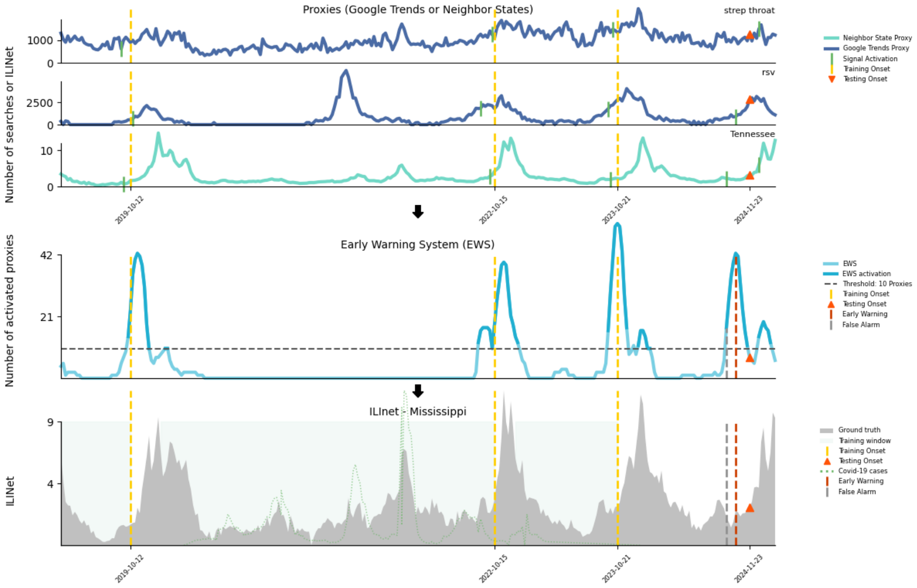


**Fig. S30**: Onset detection for Mississippi during the 2024-2025 season

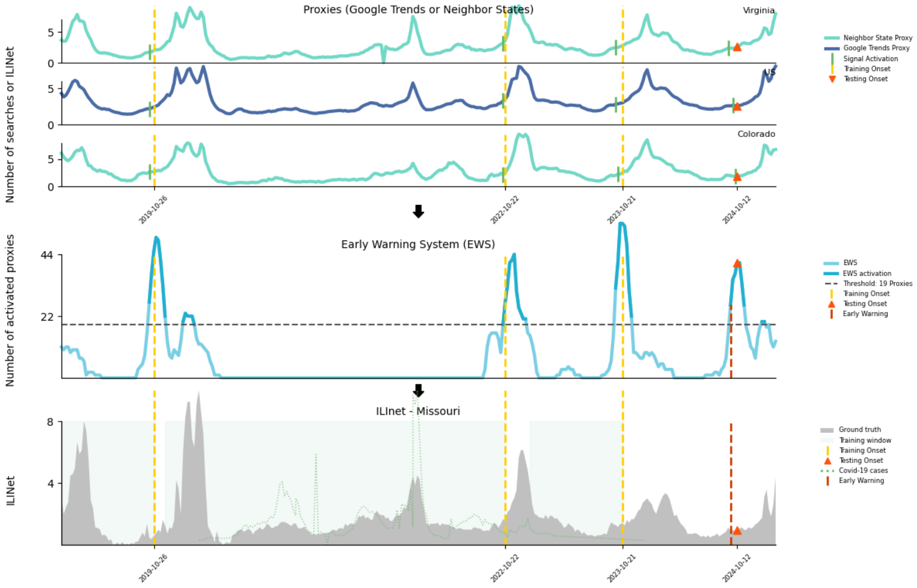


**Fig. S31**: Onset detection for Missouri during the 2024-2025 season

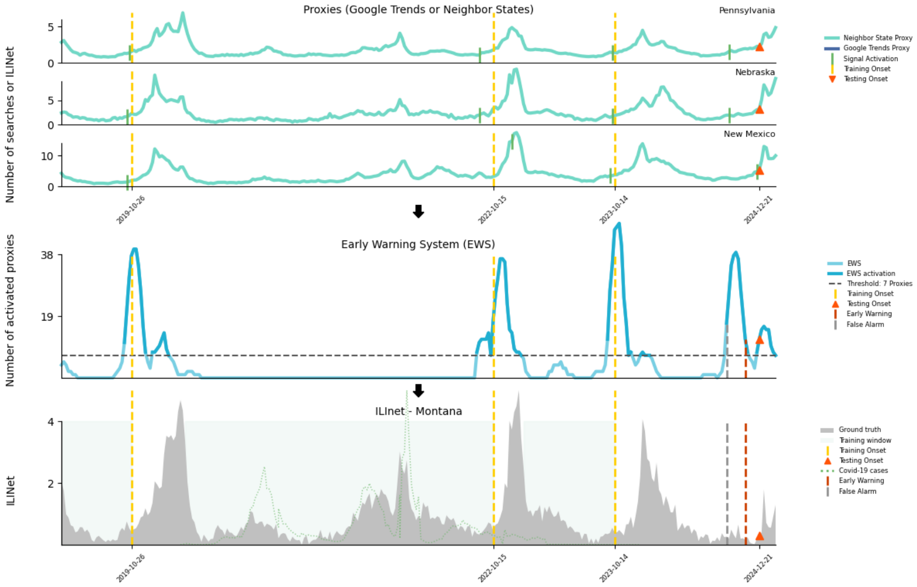


**Fig. S32**: Onset detection for Montana during the 2024-2025 season

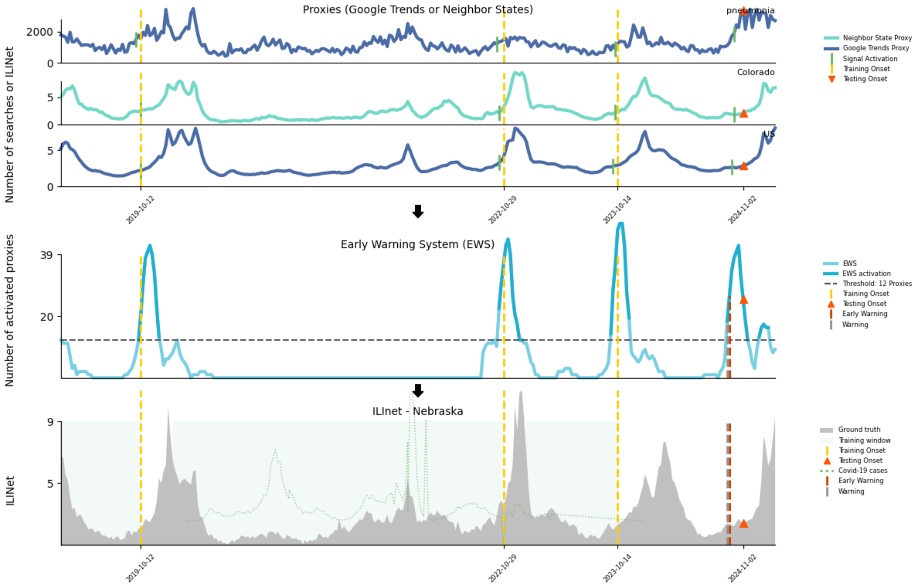


**Fig. S33**: Onset detection for Nebraska during the 2024-2025 season

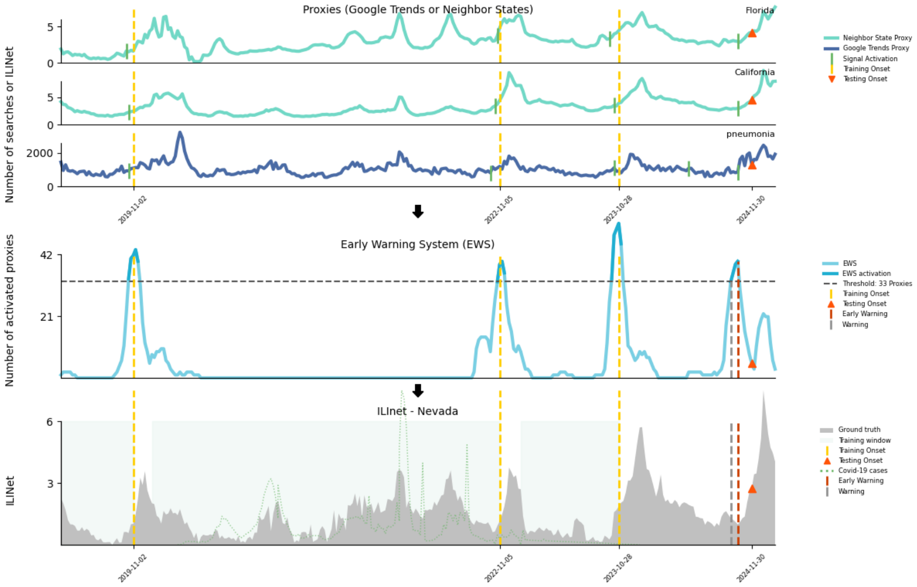


**Fig. S34**: Onset detection for Nevada during the 2024-2025 season

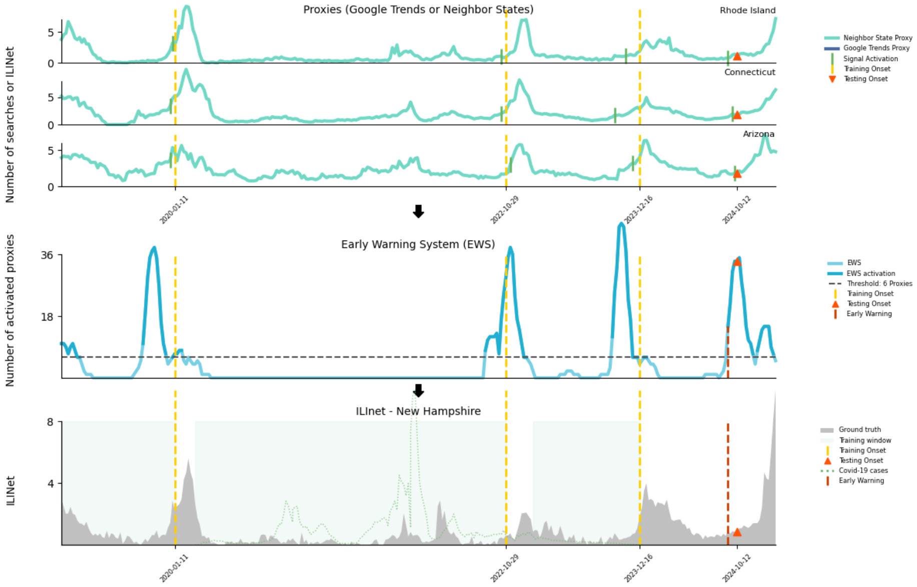


**Fig. S35**: Onset detection for New Hampshire during the 2024-2025 season

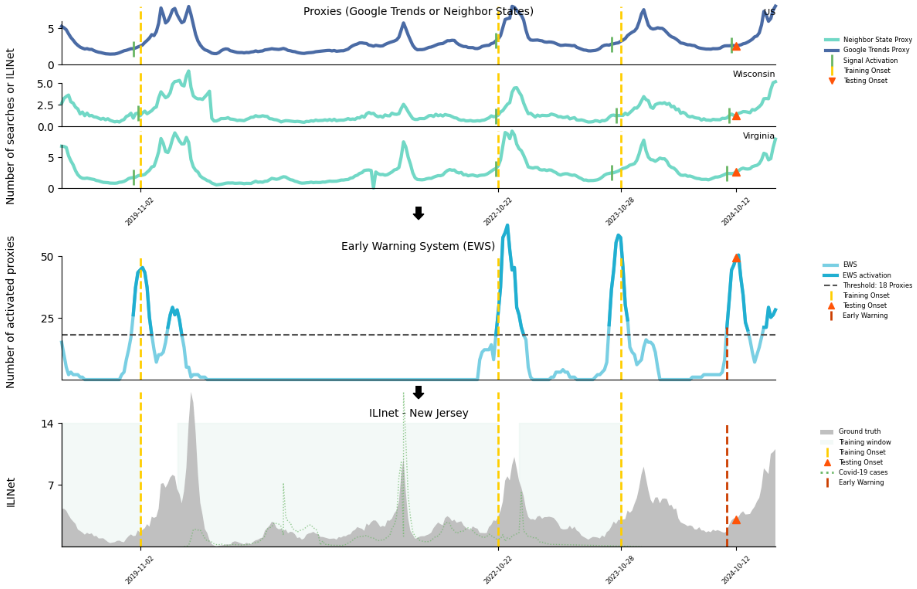


**Fig. S36**: Onset detection for New Jersey during the 2024-2025 season

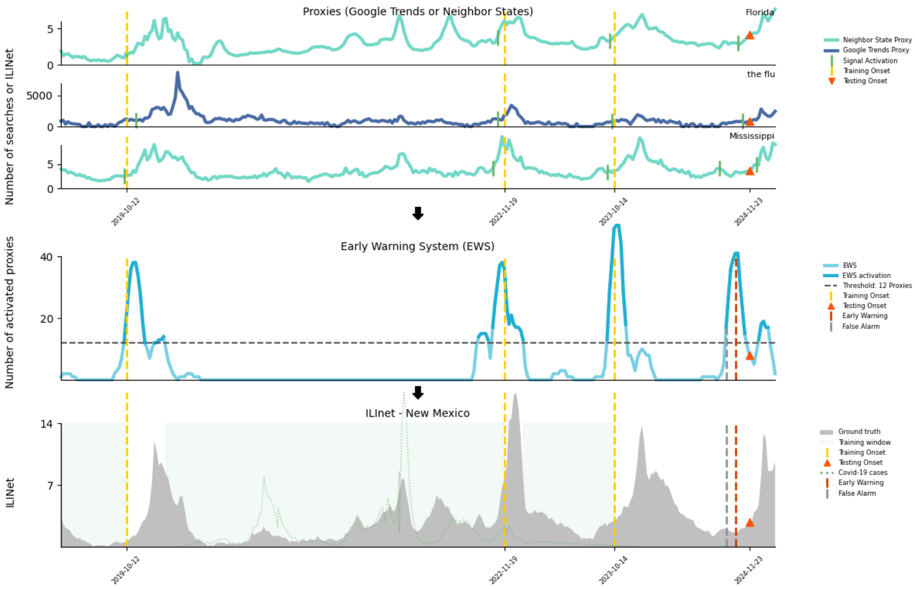


**Fig. S37**: Onset detection for New Mexico during the 2024-2025 season

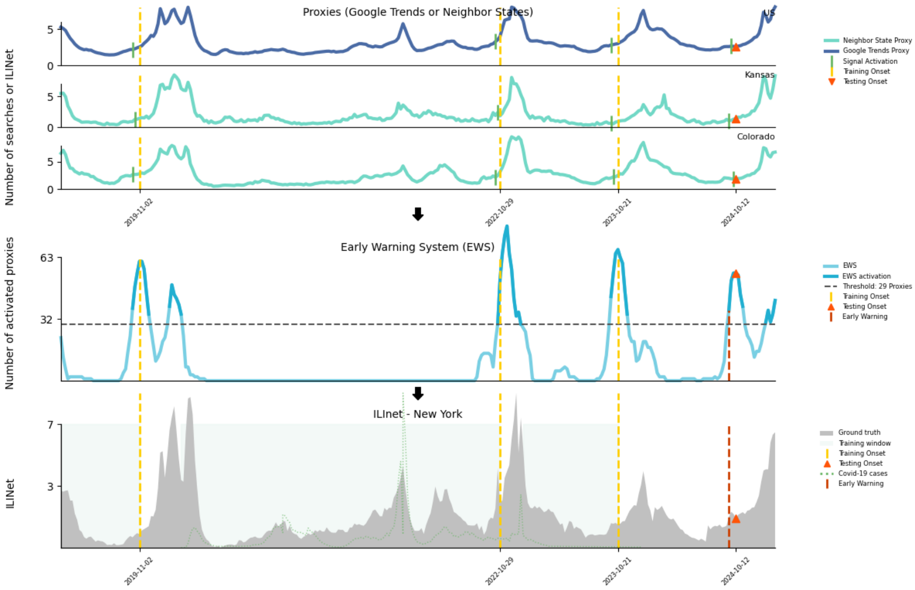


**Fig. S38**: Onset detection for New York during the 2024-2025 season

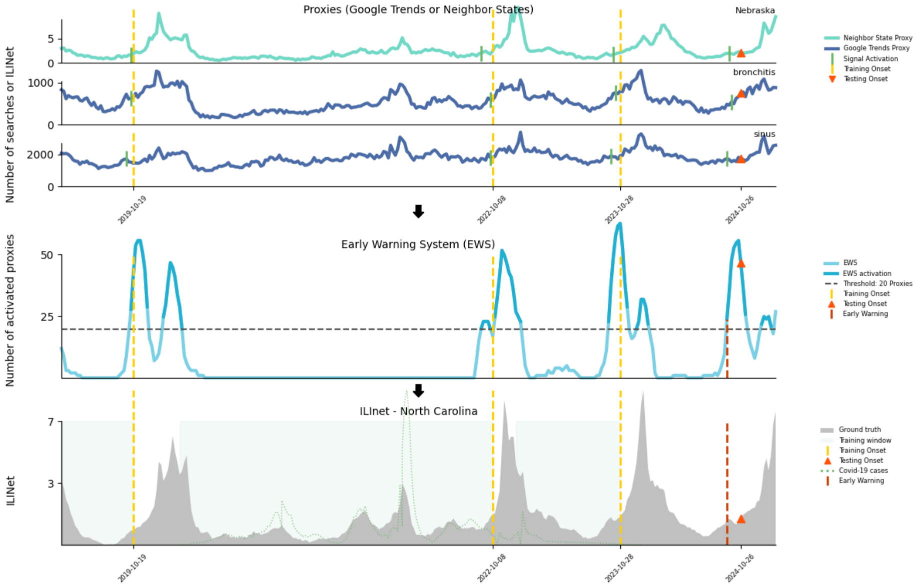


**Fig. S39**: Onset detection for North Carolina during the 2024-2025 season

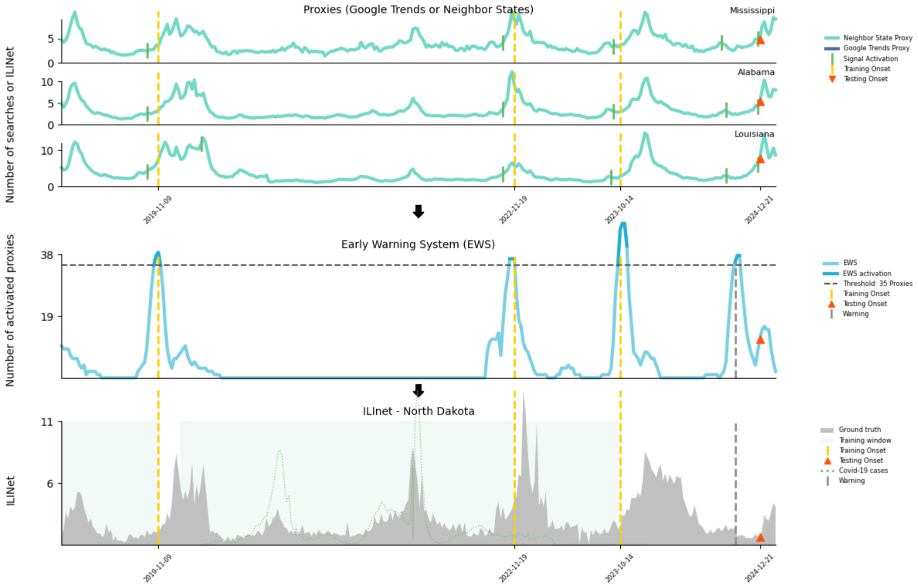


**Fig. S40**: Onset detection for North Dakota during the 2024-2025 season

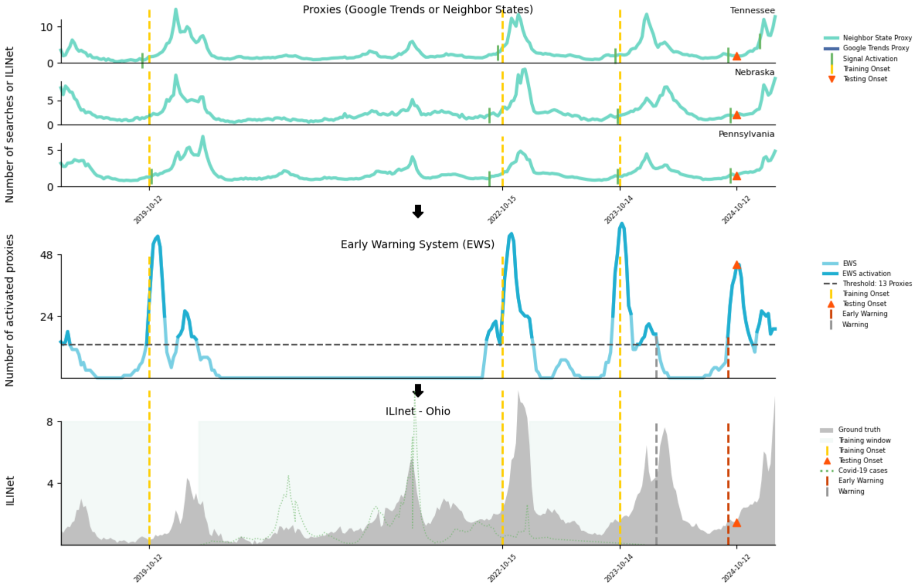


**Fig. S41**: Onset detection for Ohio during the 2024-2025 season

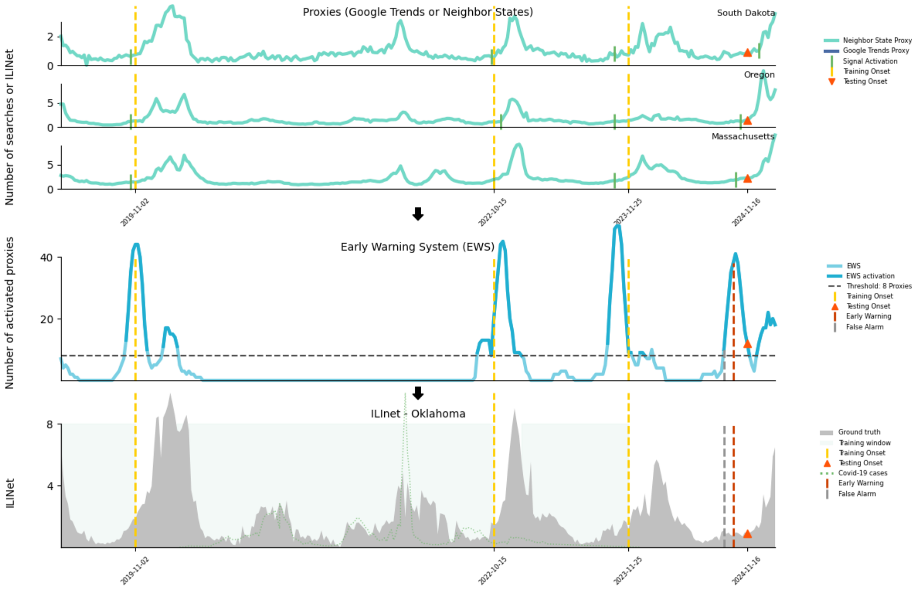


**Fig. S42**: Onset detection for Oklahoma during the 2024-2025 season

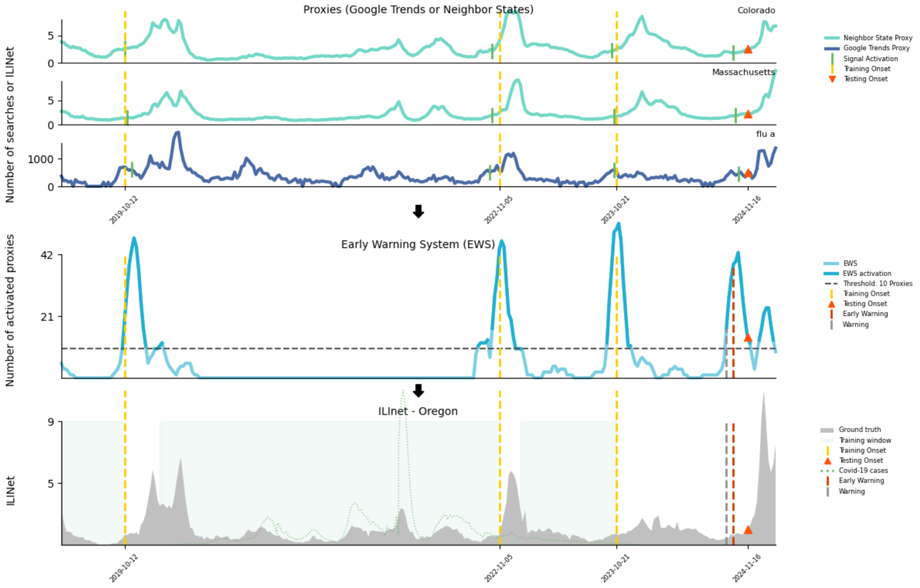


**Fig. S43**: Onset detection for Oregon during the 2024-2025 season

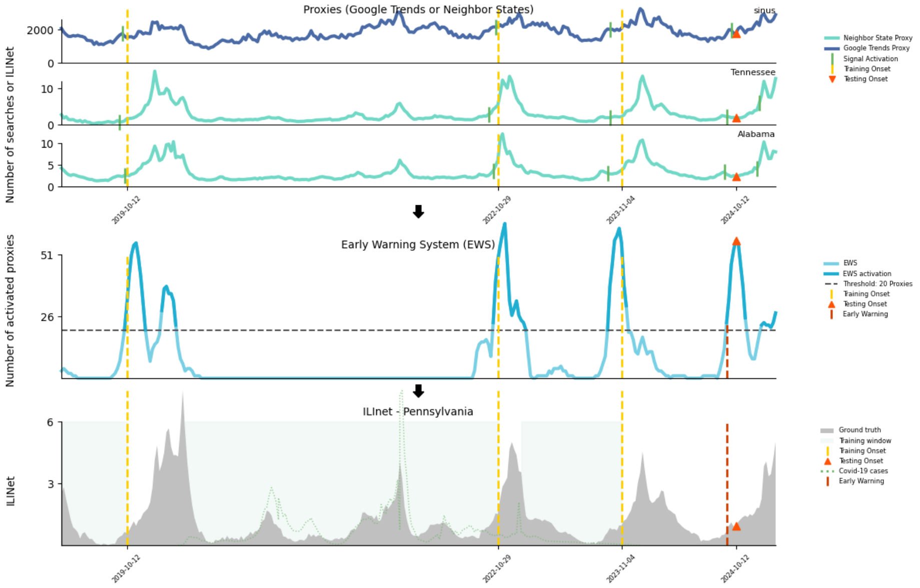


**Fig. S44**: Onset detection for Pennsylvania during the 2024-2025 season

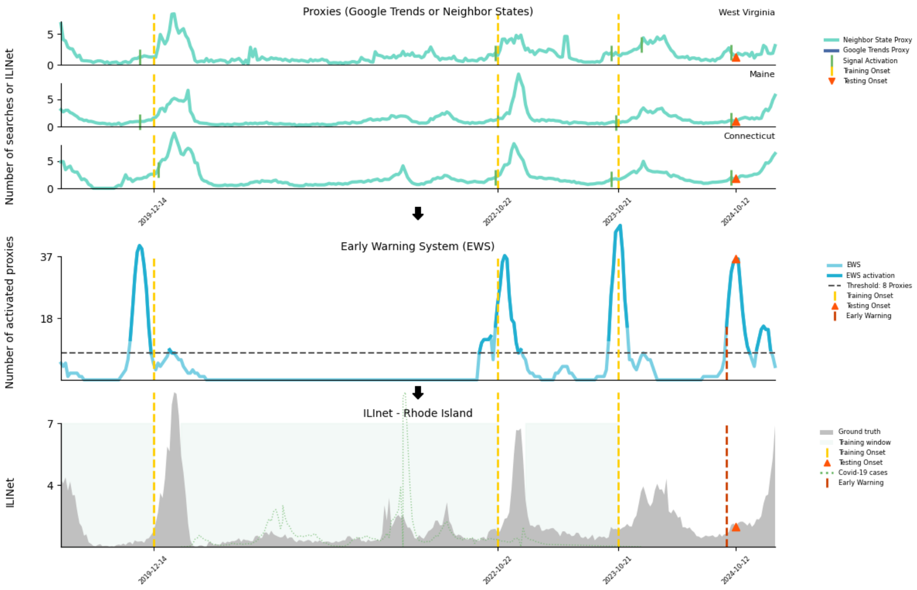


**Fig. S45**: Onset detection for Rhode Island during the 2024-2025 season

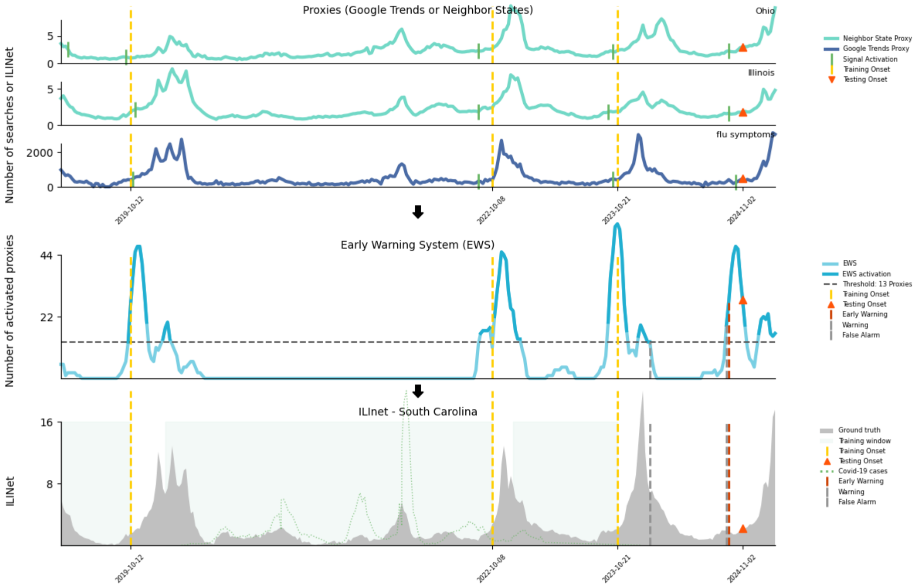


**Fig. S46**: Onset detection for South Carolina during the 2024-2025 season

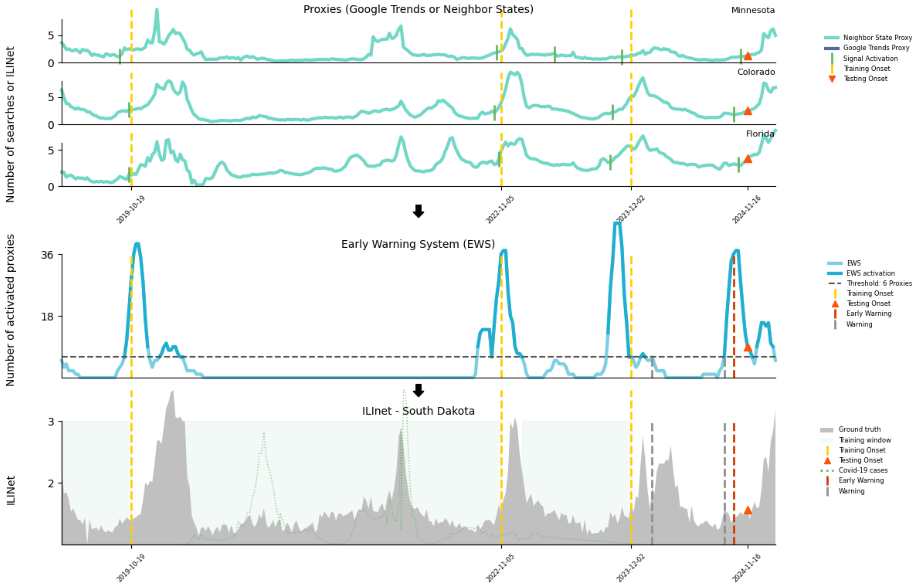


**Fig. S47**: Onset detection for South Dakota during the 2024-2025 season

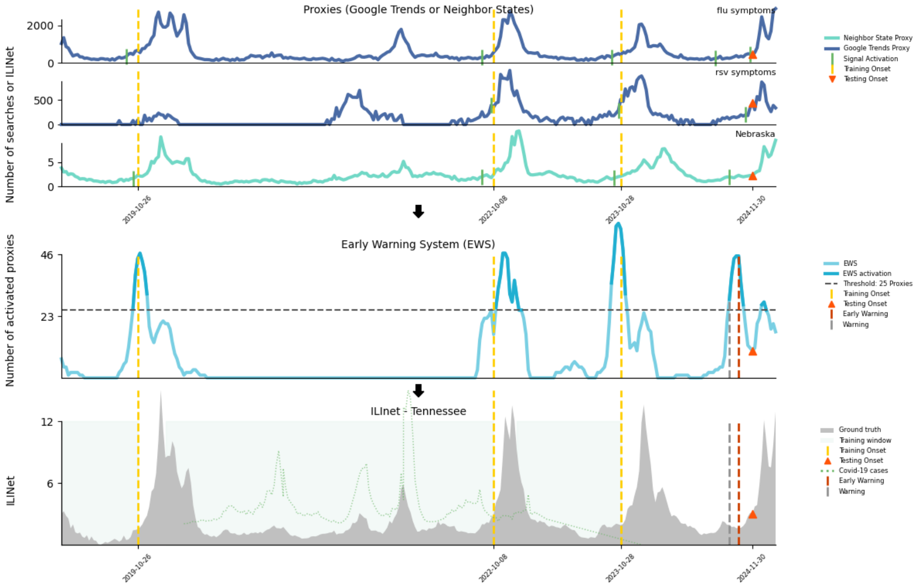


**Fig. S48**: Onset detection for Tennessee during the 2024-2025 season

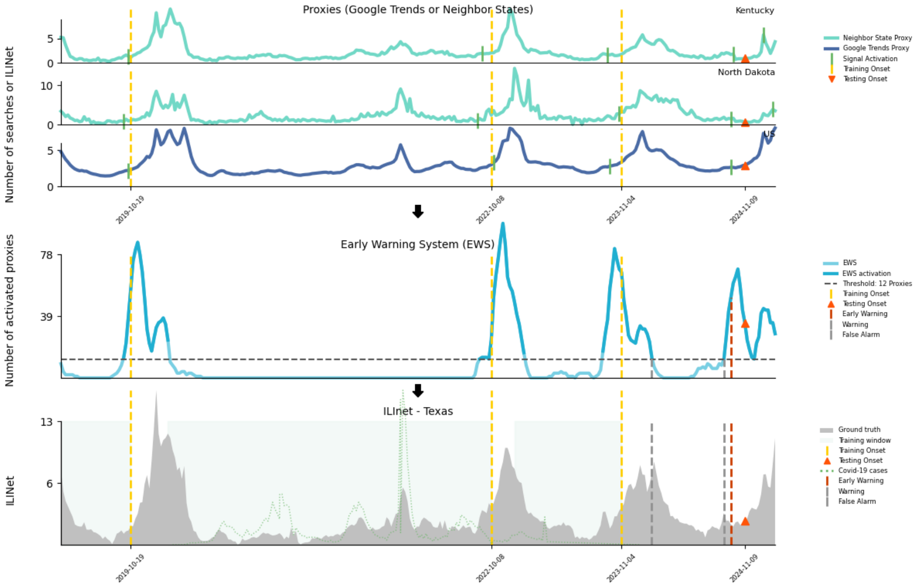


**Fig. S49**: Onset detection for Texas during the 2024-2025 season

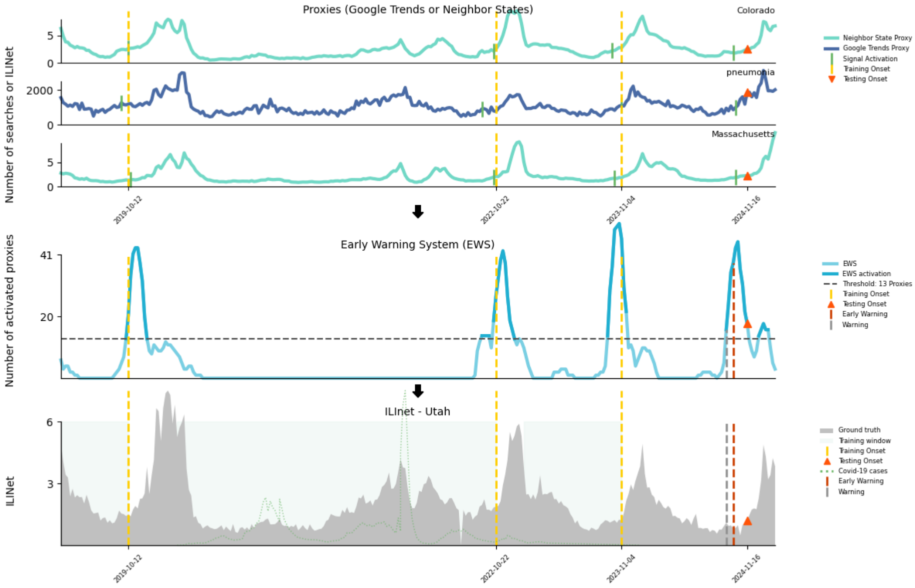


**Fig. S50**: Onset detection for Utah during the 2024-2025 season

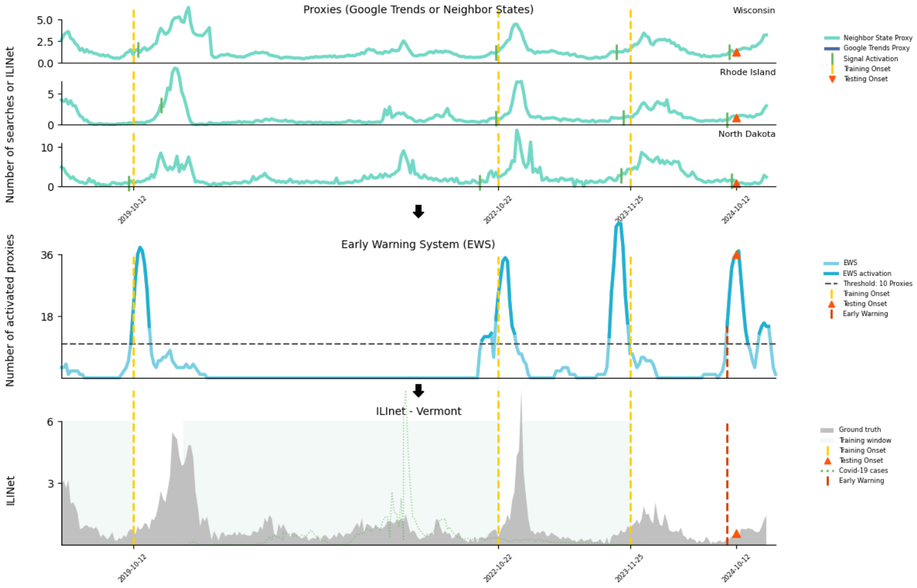


**Fig. S51**: Onset detection for Vermont during the 2024-2025 season

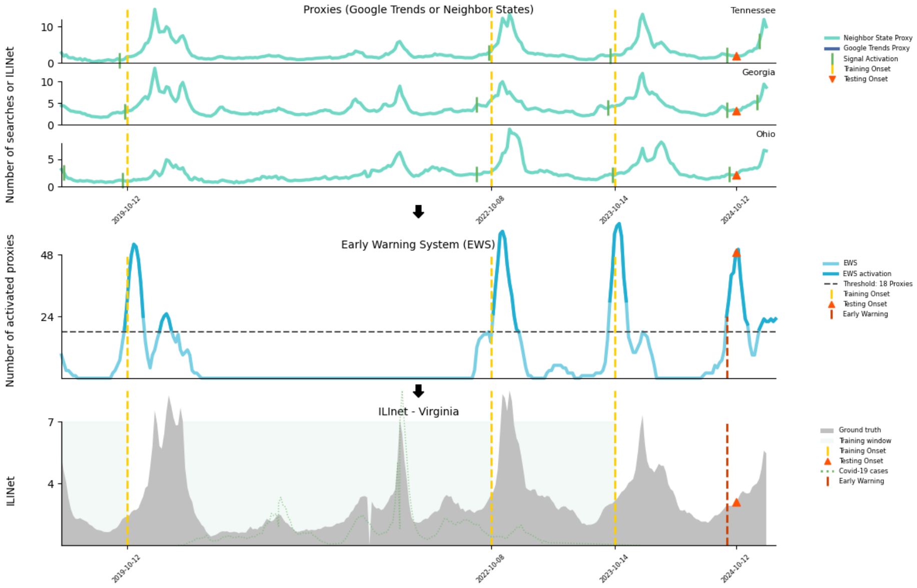


**Fig. S52**: Onset detection for Virginia during the 2024-2025 season

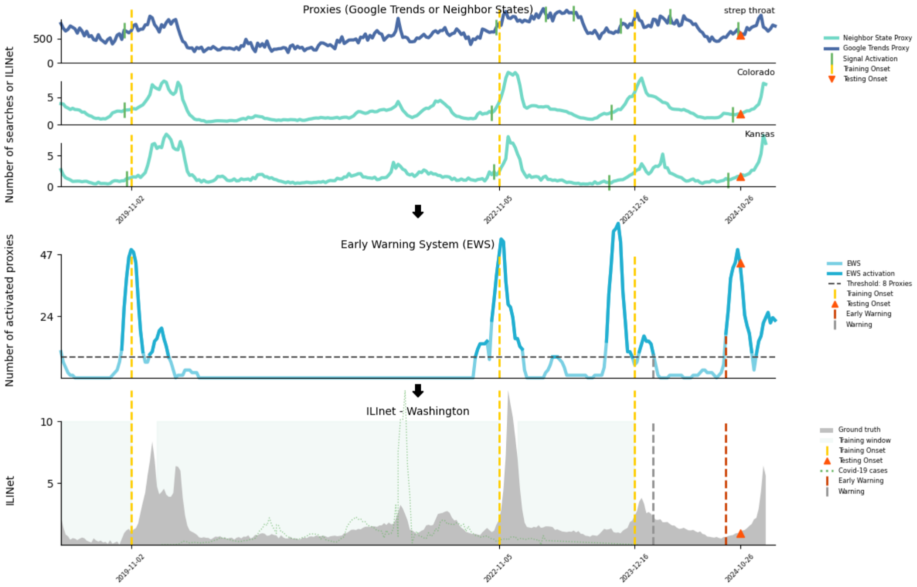


**Fig. S53**: Onset detection for Washington during the 2024-2025 season

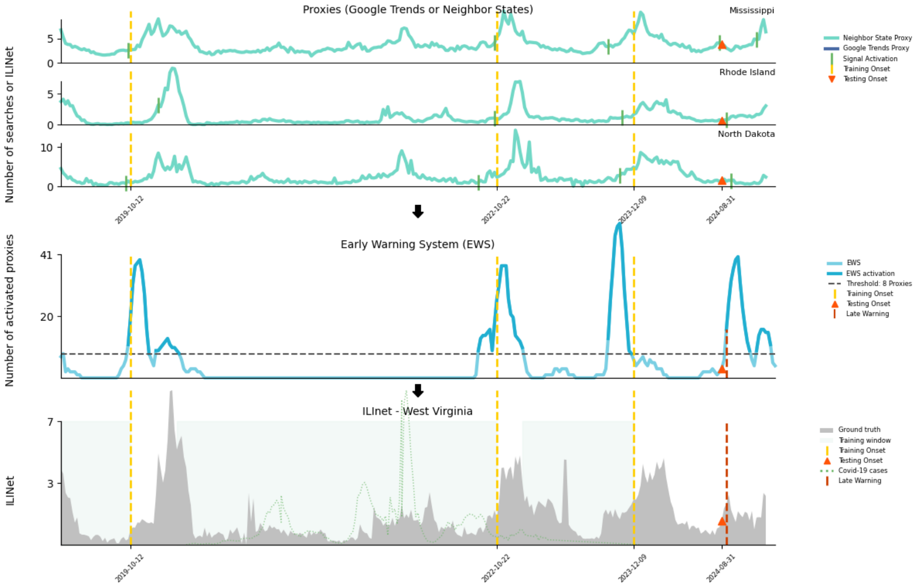


**Fig. S54**: Onset detection for West Virginia during the 2024-2025 season

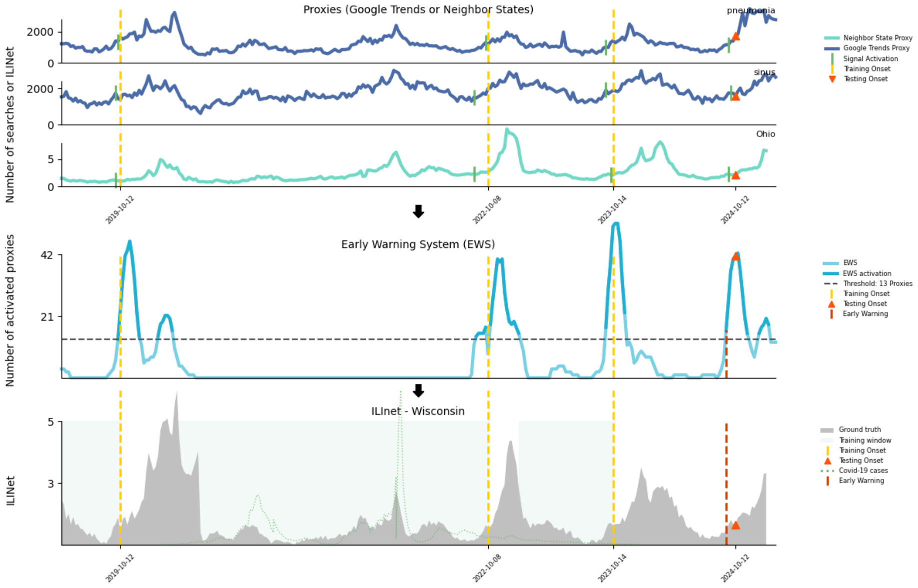


**Fig. S55**: Onset detection for Wisconsin during the 2024-2025 season

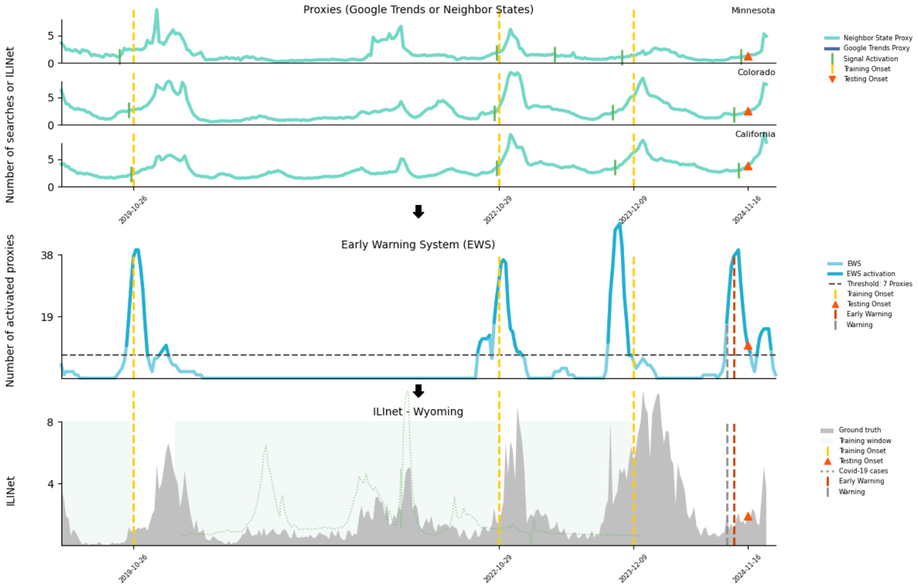


**Fig. S56**: Onset detection for Wyoming during the 2024-2025 season

### Peak Examples 2024-2025 Real-Time Predictions

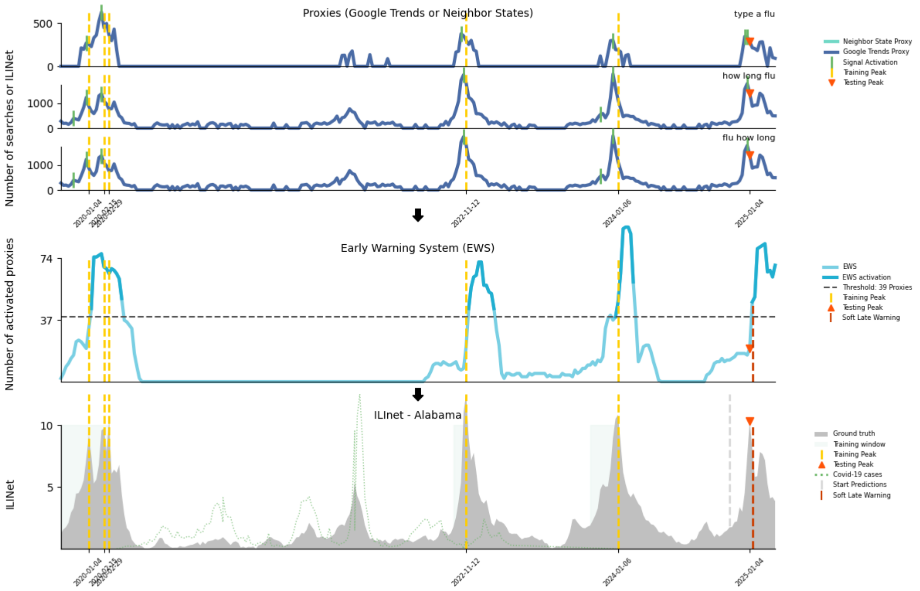


**Fig. S57**: Peak detection for Alabama during the 2024-2025 season

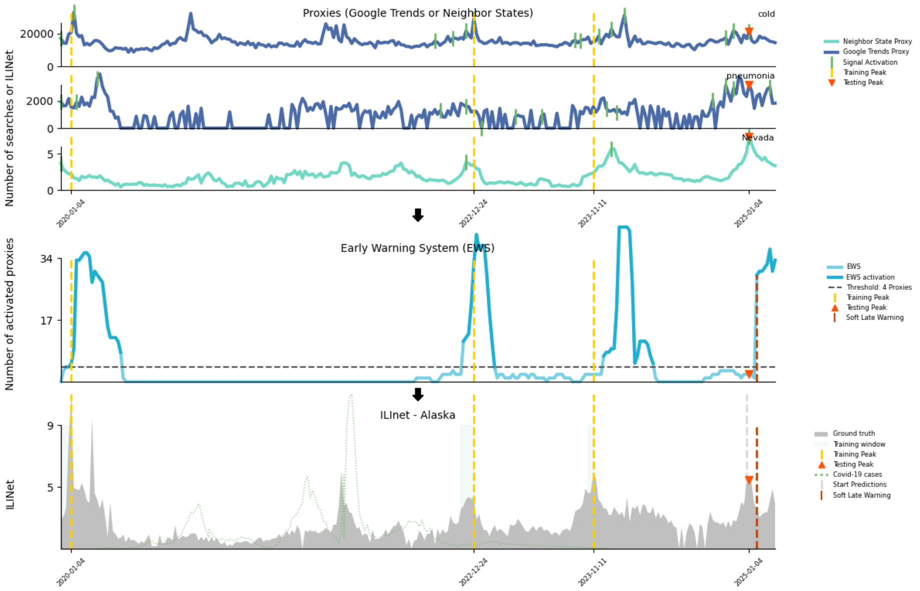


**Fig. S58**: Peak detection for Alaska during the 2024-2025 season

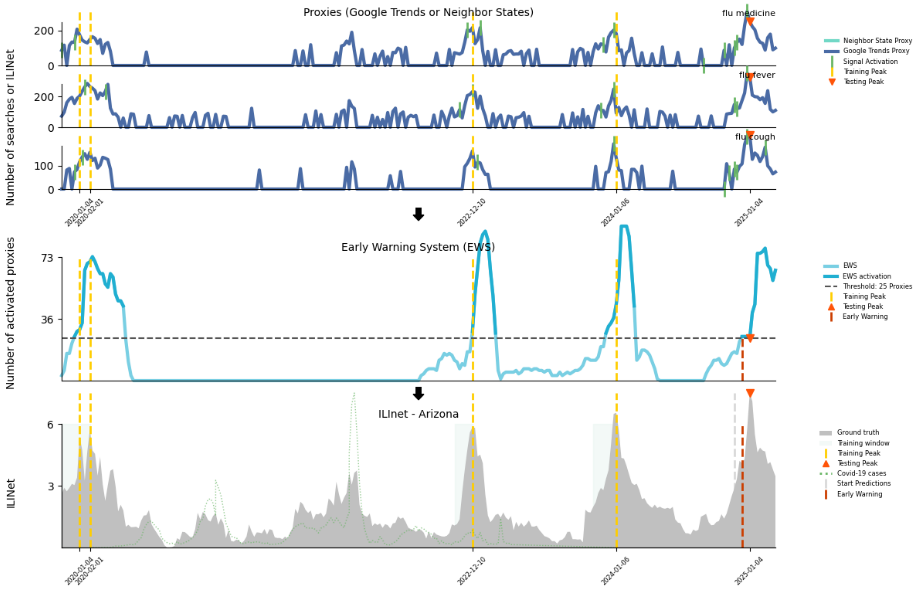


**Fig. S59**: Peak detection for Arizona during the 2024-2025 season

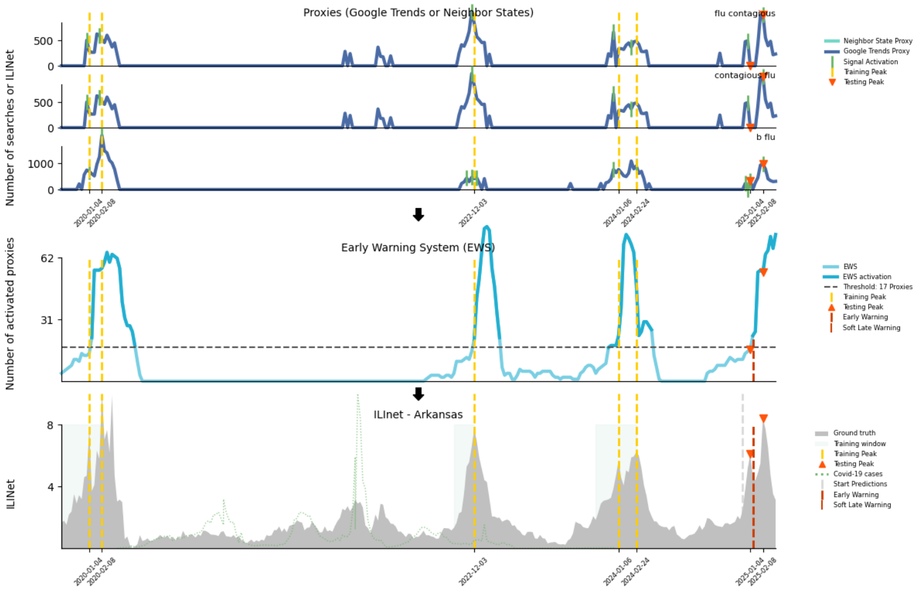


**Fig. S60**: Peak detection for Arkansas during the 2024-2025 season

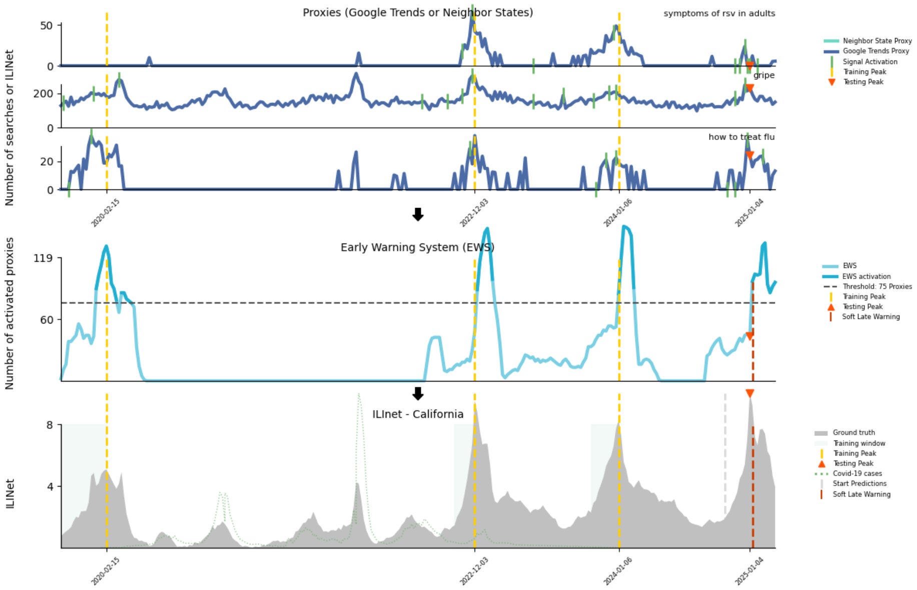


**Fig. S61**: Peak detection for California during the 2024-2025 season

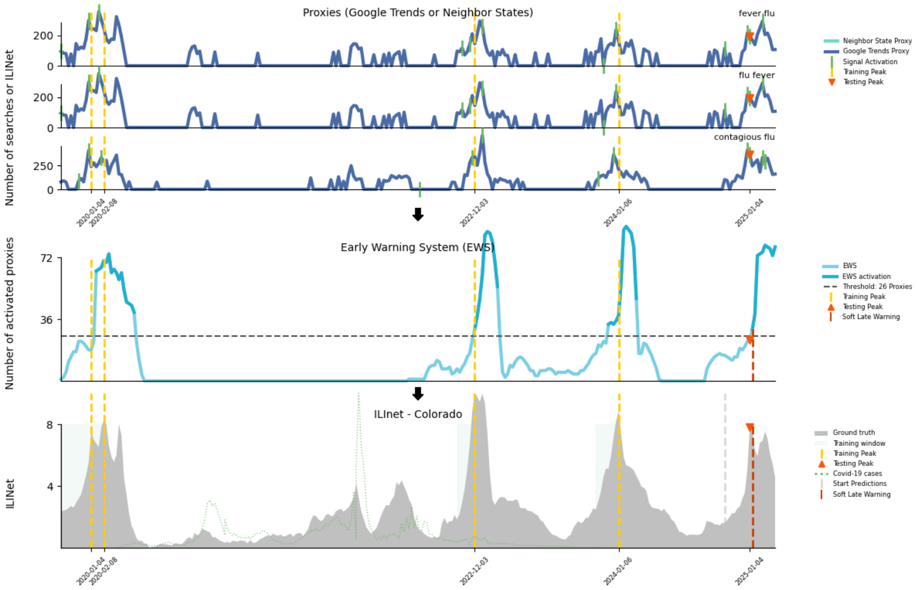


**Fig. S62**: Peak detection for Colorado during the 2024-2025 season

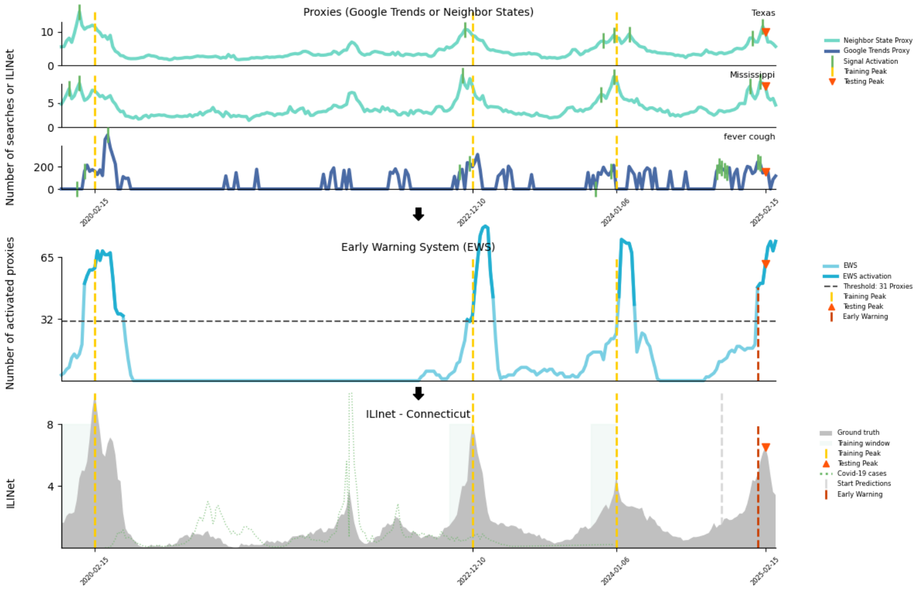


**Fig. S63**: Peak detection for Connecticut during the 2024-2025 season

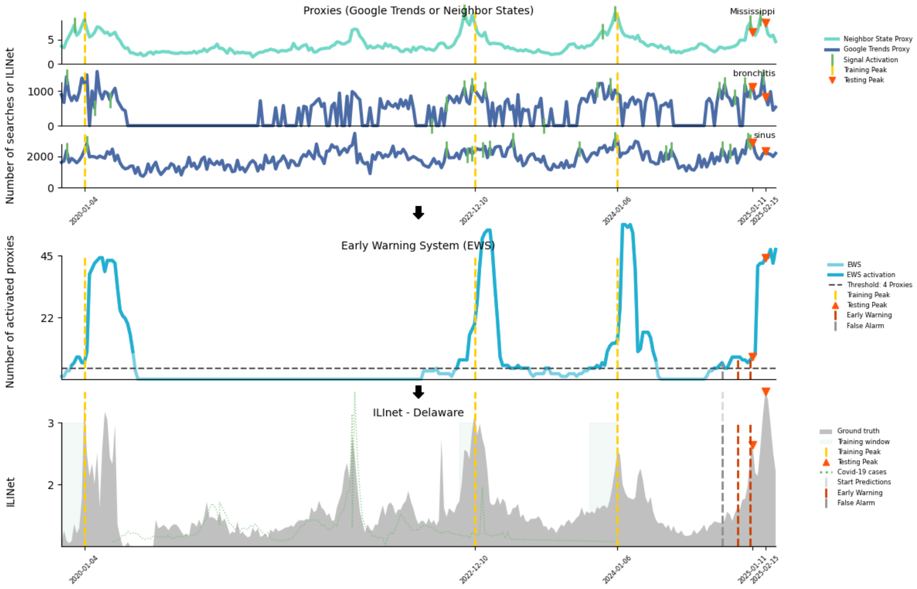


**Fig. S64**: Peak detection for Delaware during the 2024-2025 season

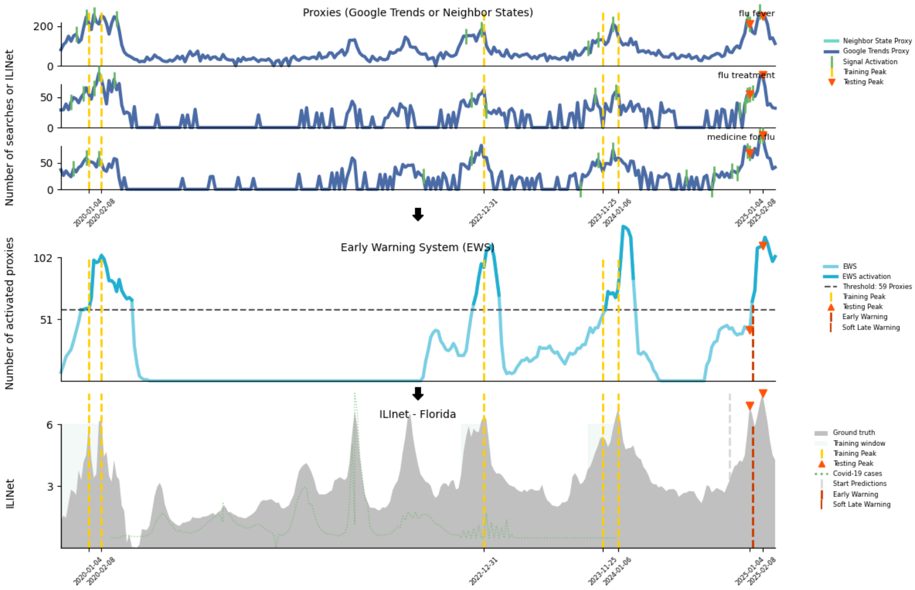


**Fig. S65**: Peak detection for Florida during the 2024-2025 season

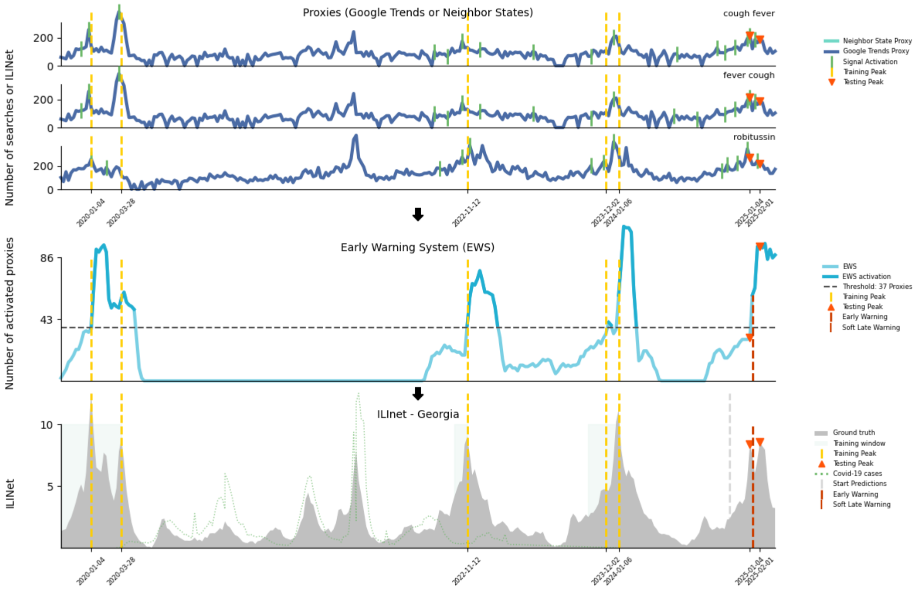


**Fig. S66**: Peak detection for Georgia during the 2024-2025 season

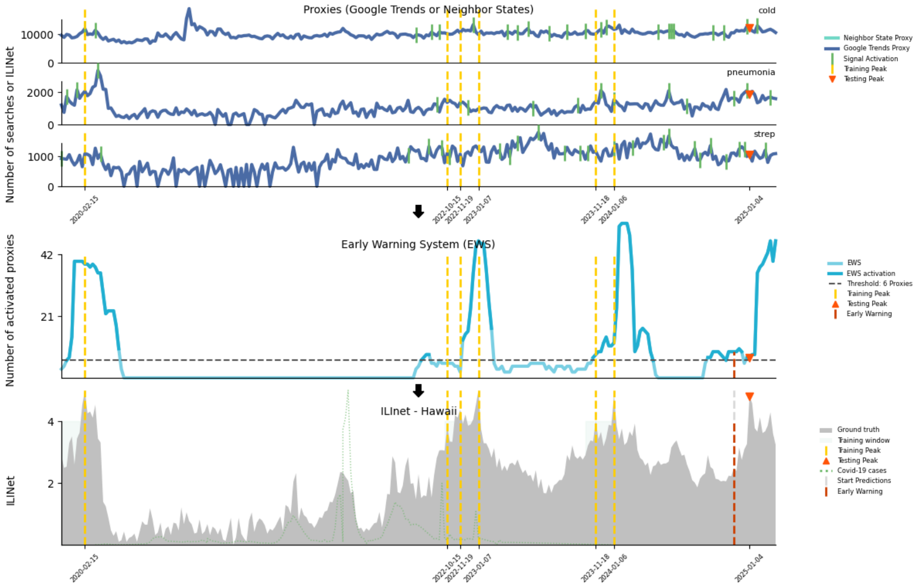


**Fig. S67**: Peak detection for Hawaii during the 2024-2025 season

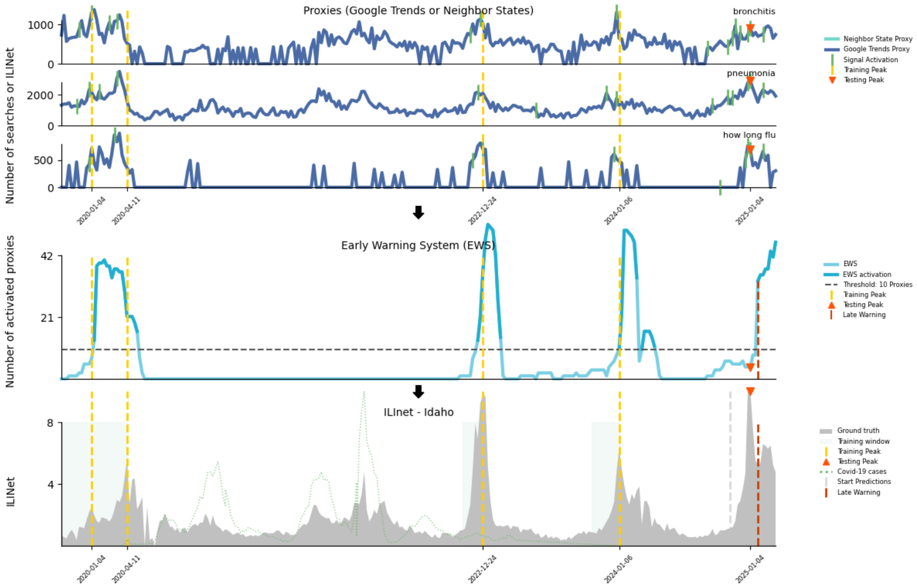


**Fig. S68**: Peak detection for Idaho during the 2024-2025 season

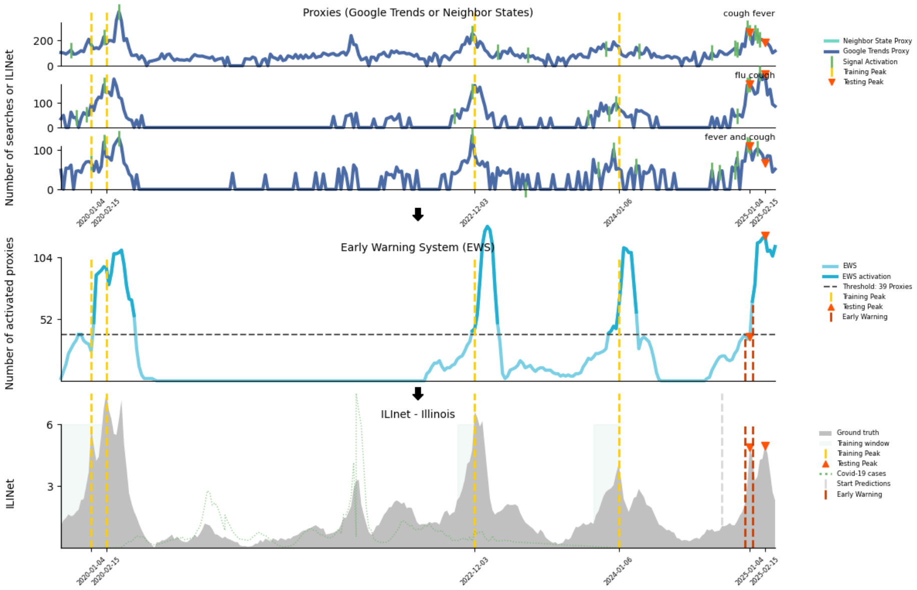


**Fig. S69**: Peak detection for Illinois during the 2024-2025 season

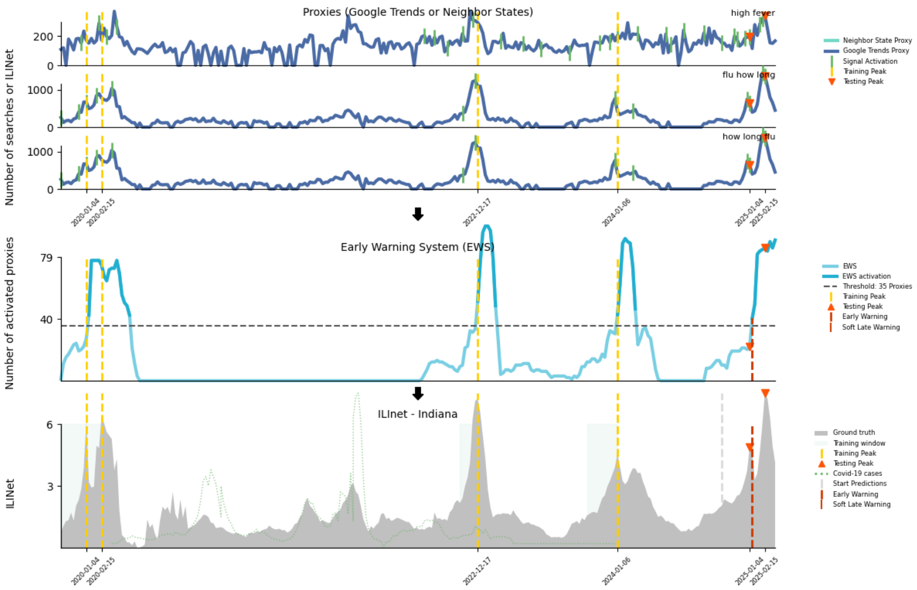


**Fig. S70**: Peak detection for Indiana during the 2024-2025 season

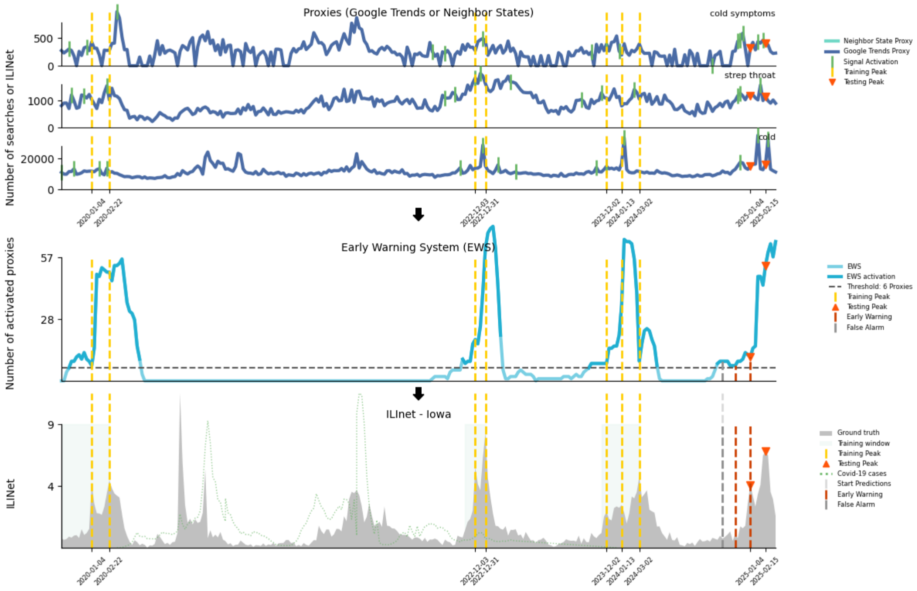


**Fig. S71**: Peak detection for Iowa during the 2024-2025 season

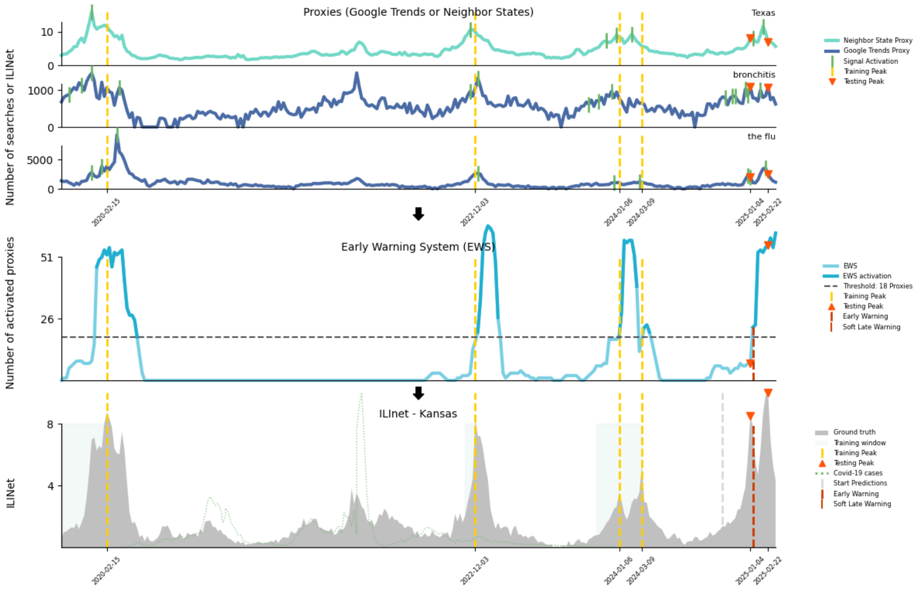


**Fig. S72**: Peak detection for Kansas during the 2024-2025 season

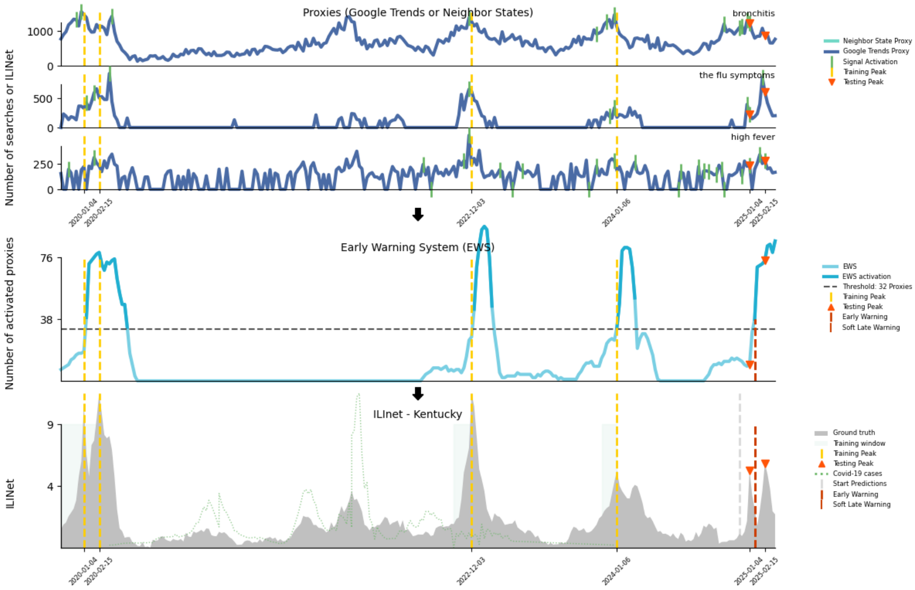


**Fig. S73**: Peak detection for Kentucky during the 2024-2025 season

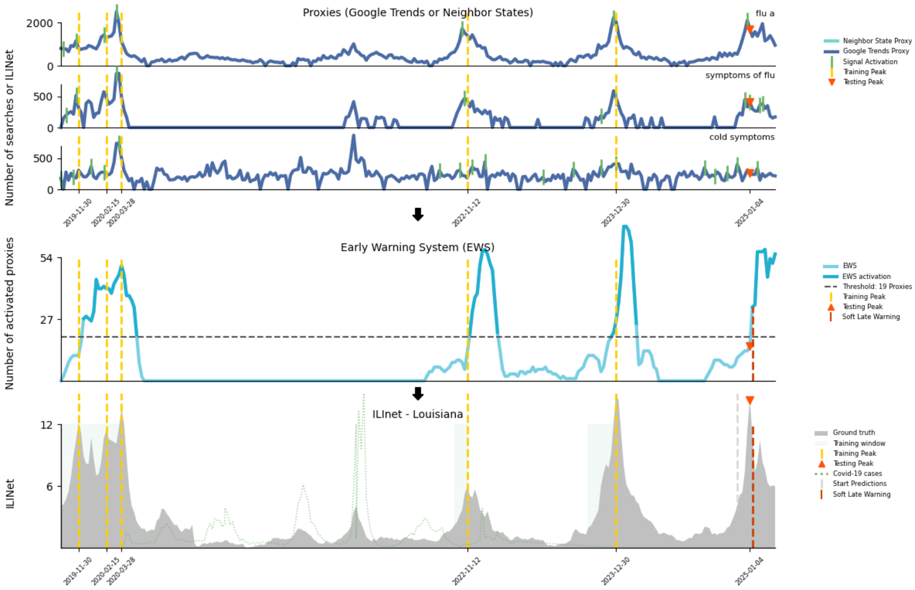


**Fig. S74**: Peak detection for Louisiana during the 2024-2025 season

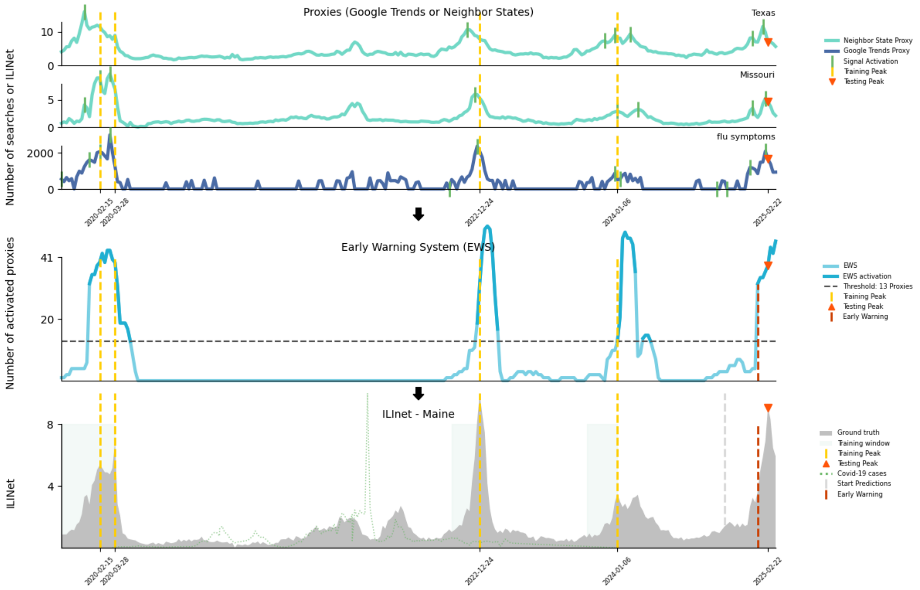


**Fig. S75**: Peak detection for Maine during the 2024-2025 season

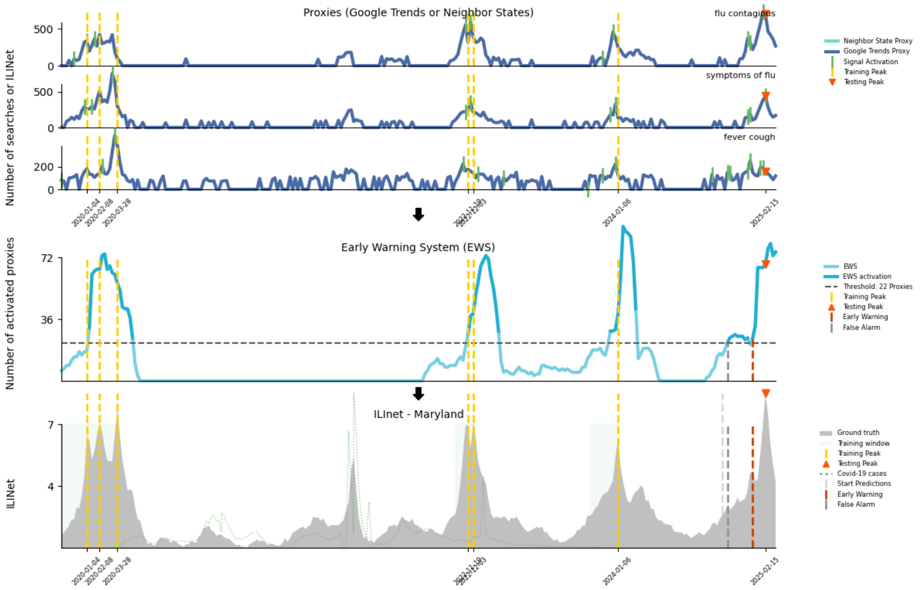


**Fig. S76**: Peak detection for Maryland during the 2024-2025 season

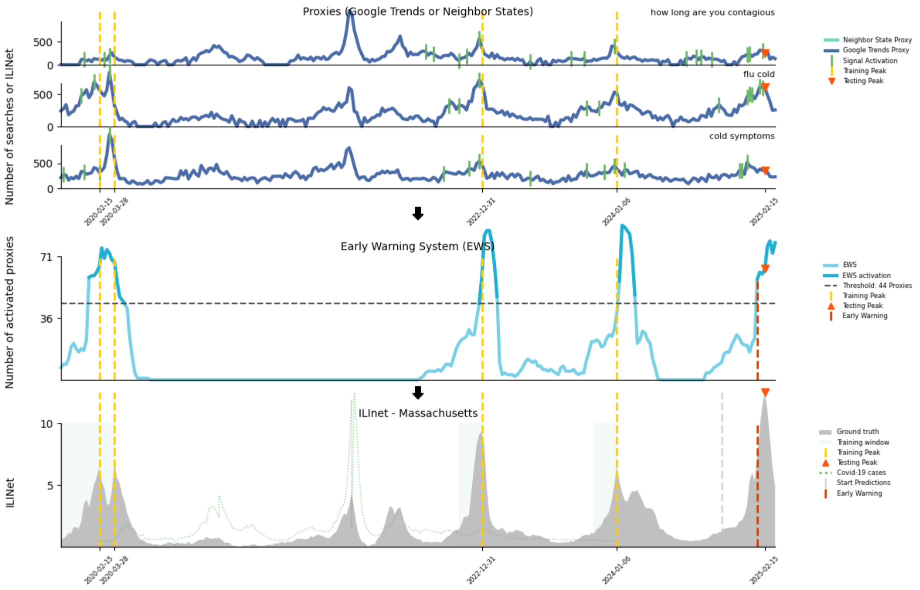


**Fig. S77**: Peak detection for Massachusetts during the 2024-2025 season

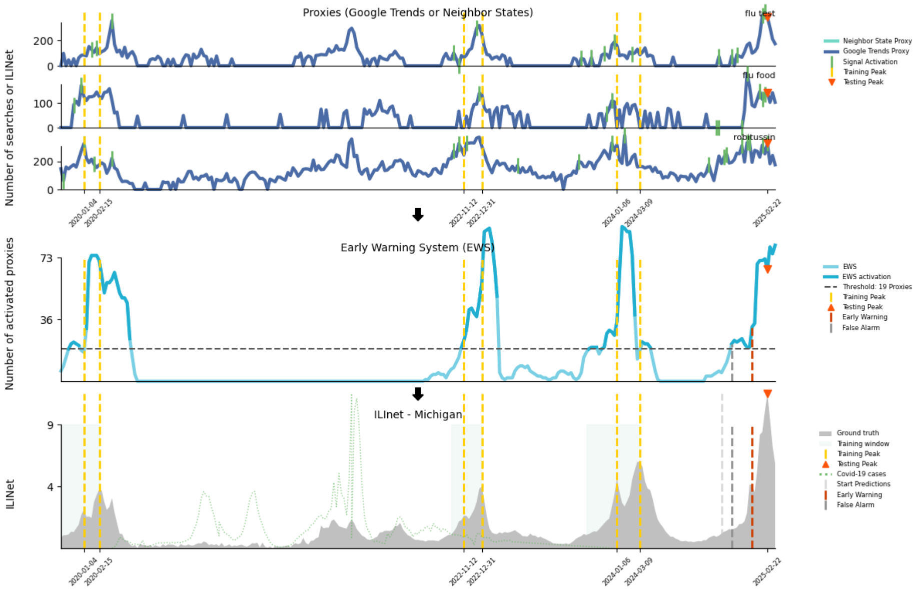


**Fig. S78**: Peak detection for Michigan during the 2024-2025 season

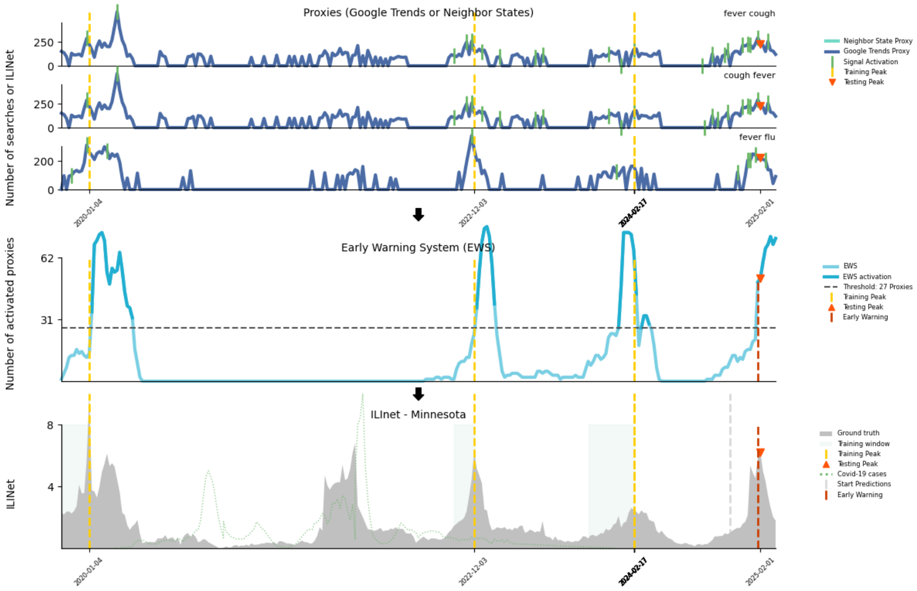


**Fig. S79**: Peak detection for Minnesota during the 2024-2025 season

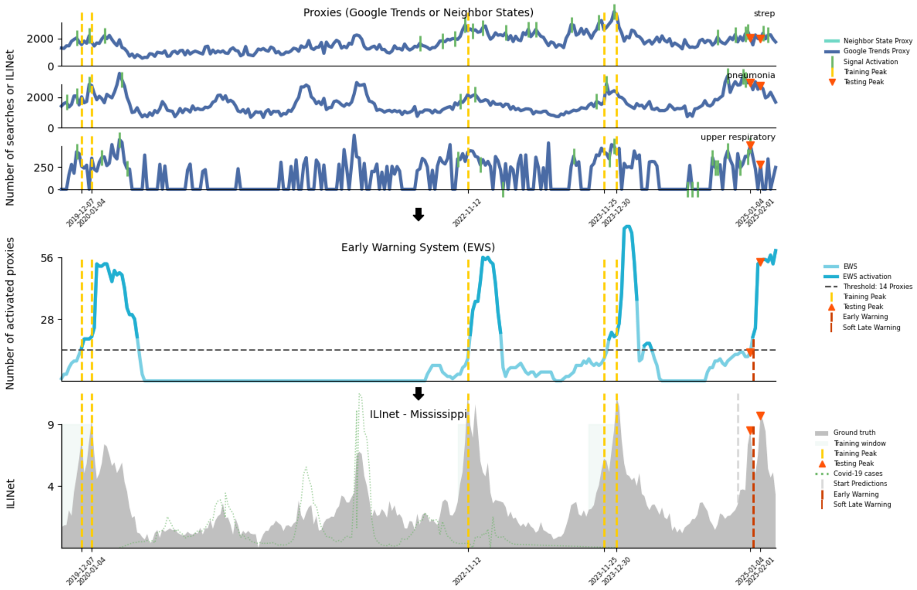


**Fig. S80**: Peak detection for Mississippi during the 2024-2025 season

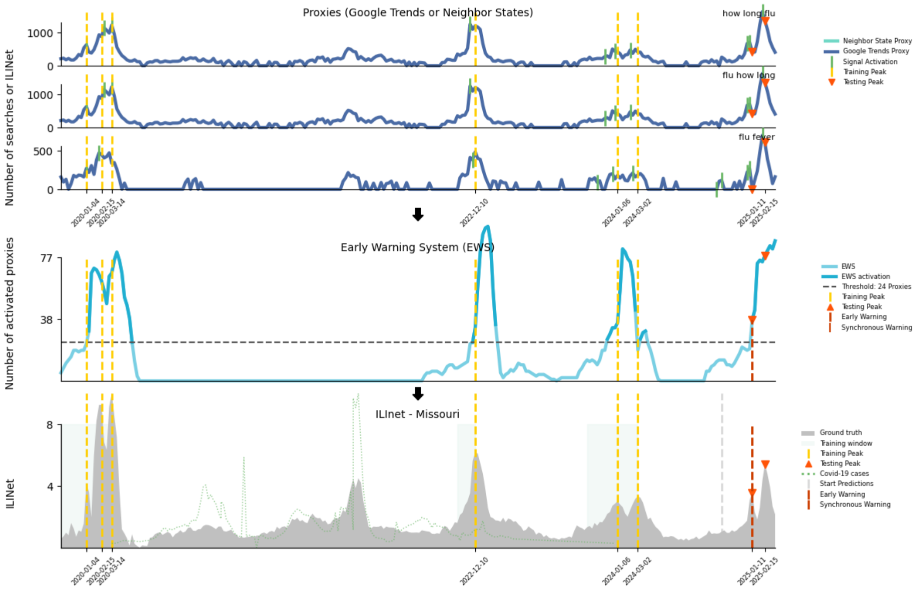


**Fig. S81**: Peak detection for Missouri during the 2024-2025 season

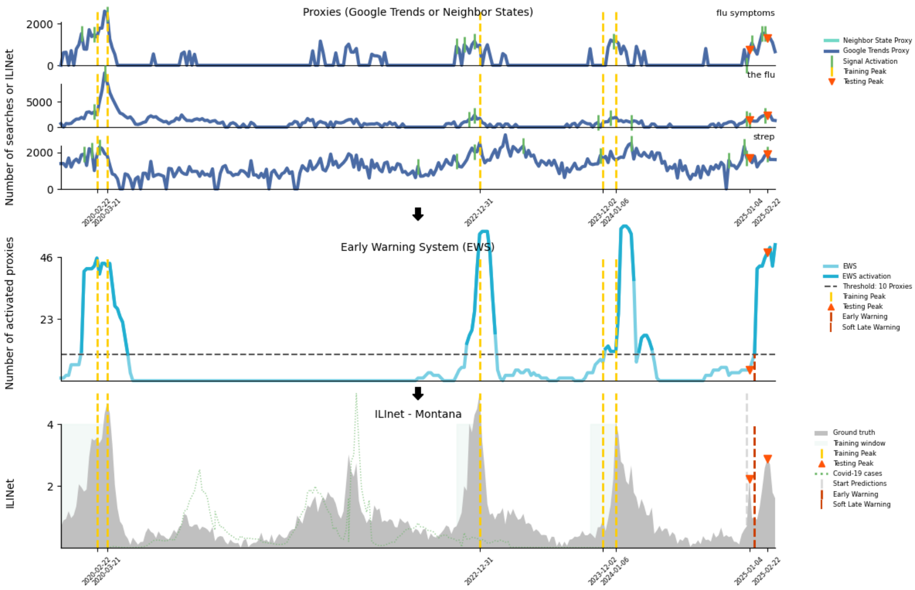


**Fig. S82**: Peak detection for Montana during the 2024-2025 season

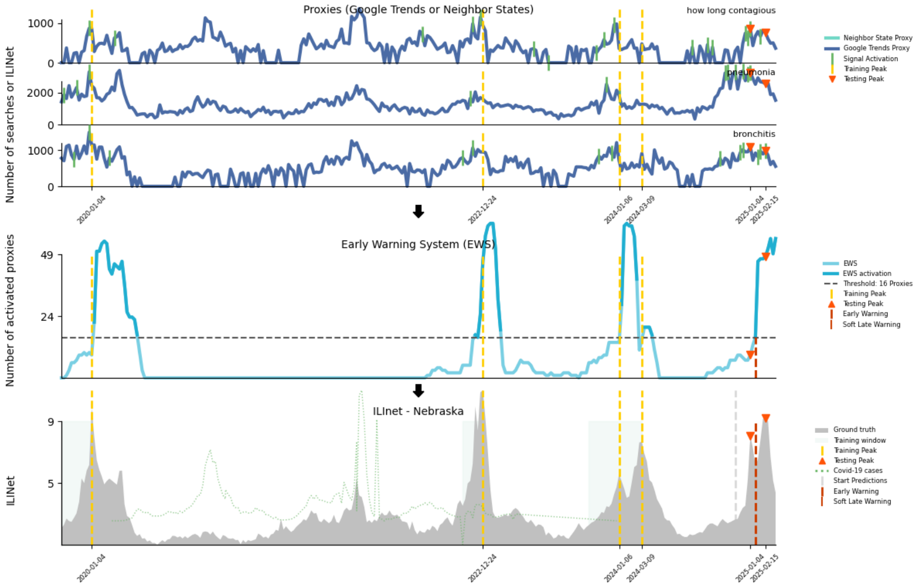


**Fig. S83**: Peak detection for Nebraska during the 2024-2025 season

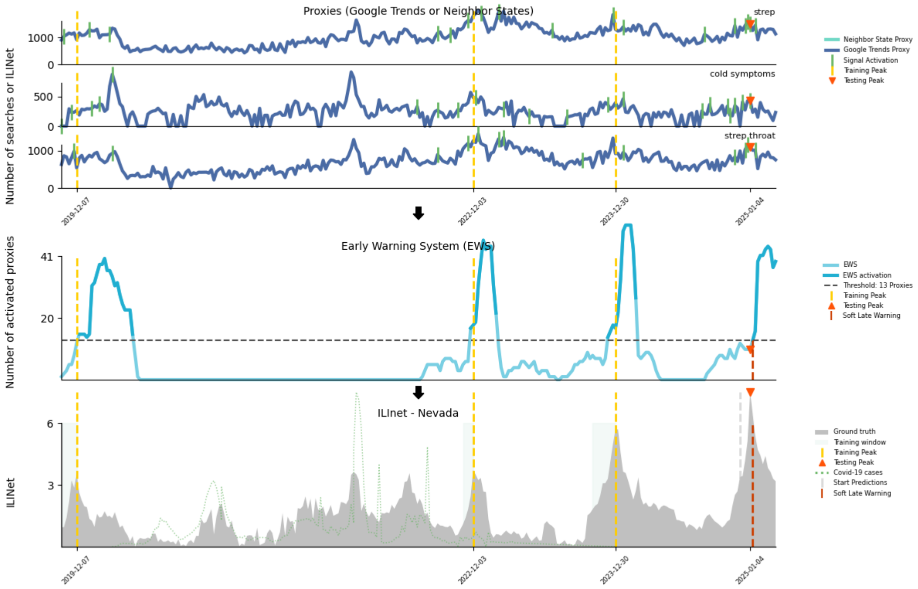


**Fig. S84**: Peak detection for Nevada during the 2024-2025 season

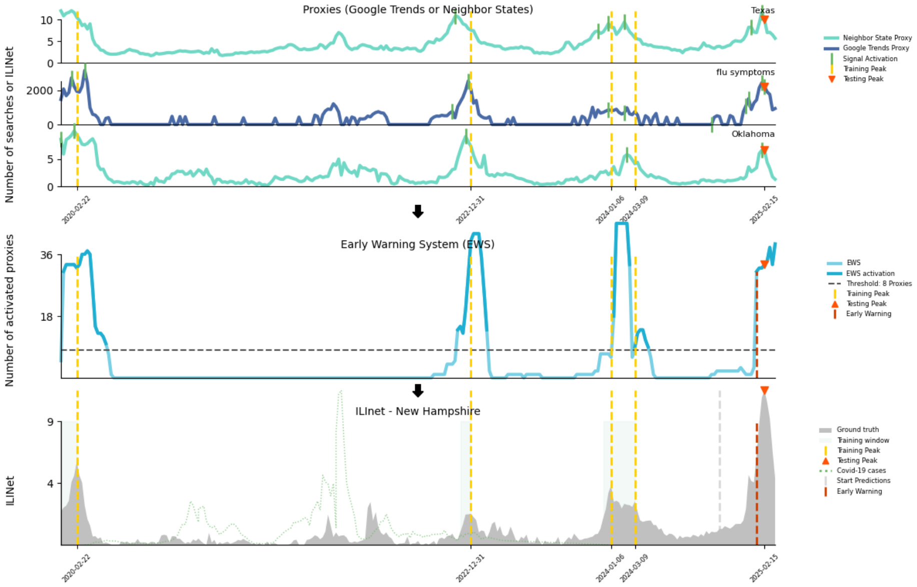


**Fig. S85**: Peak detection for New Hampshire during the 2024-2025 season

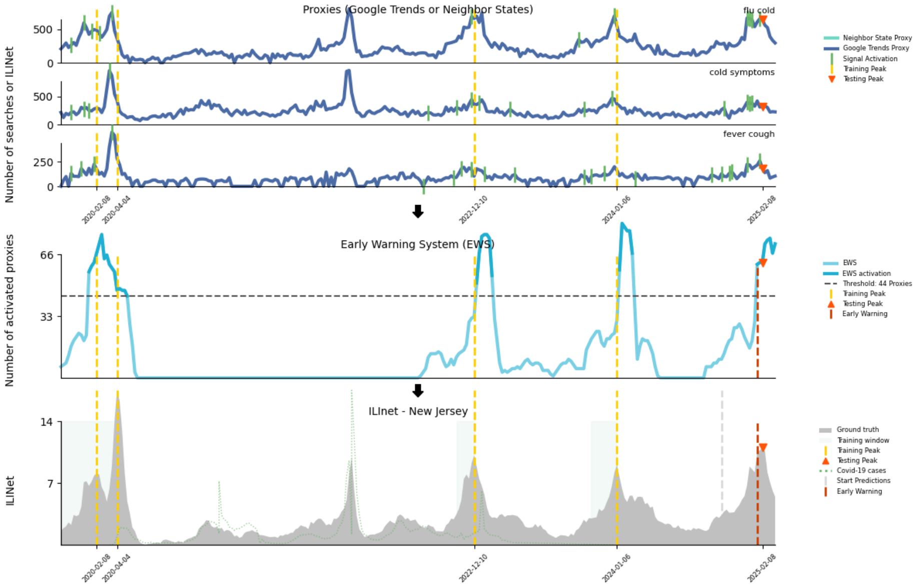


**Fig. S86**: Peak detection for New Jersey during the 2024-2025 season

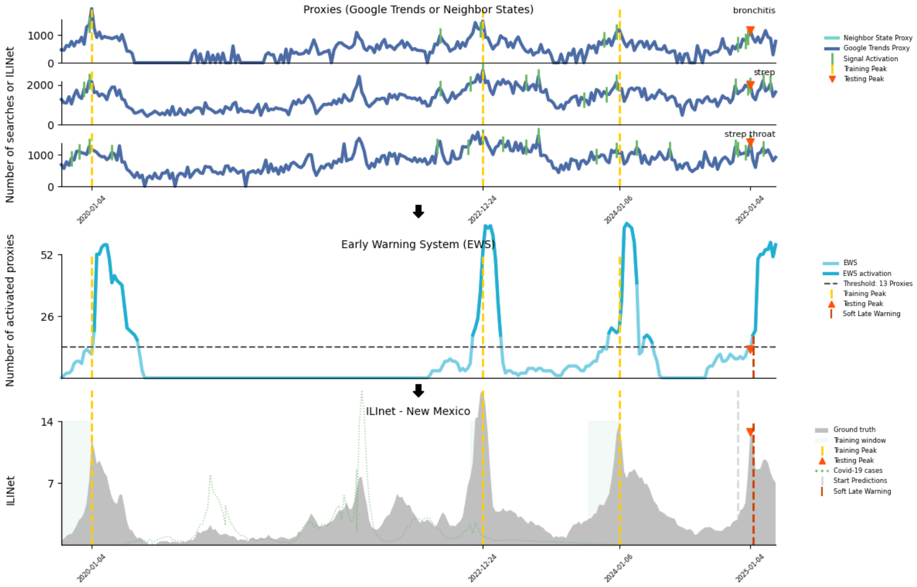


**Fig. S87**: Peak detection for New Mexico during the 2024-2025 season

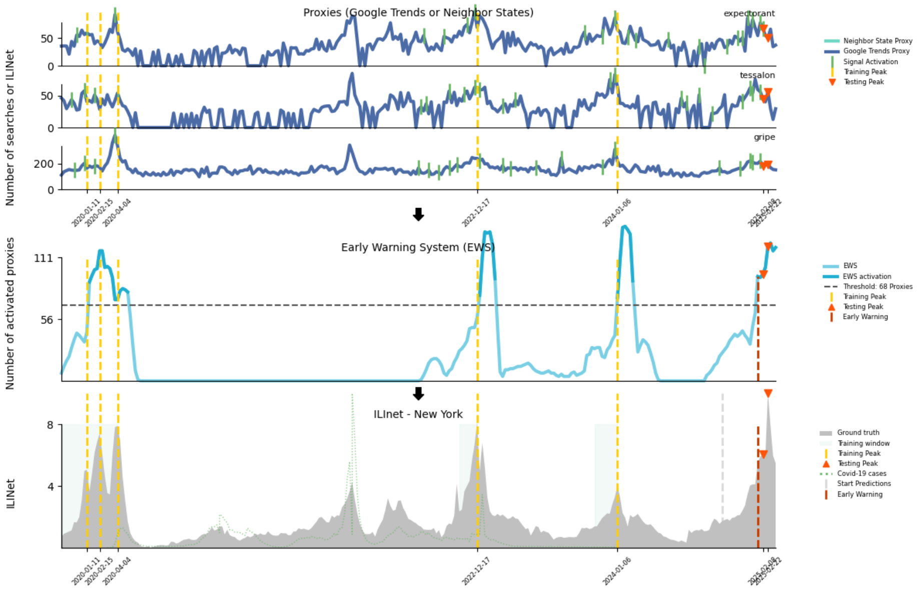


**Fig. S88**: Peak detection for New York during the 2024-2025 season

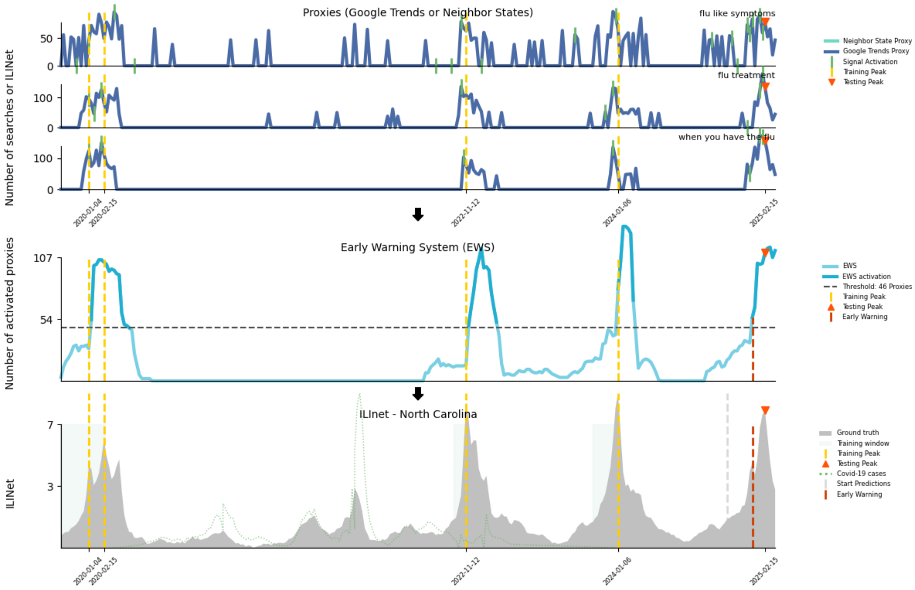


**Fig. S89**: Peak detection for North Carolina during the 2024-2025 season

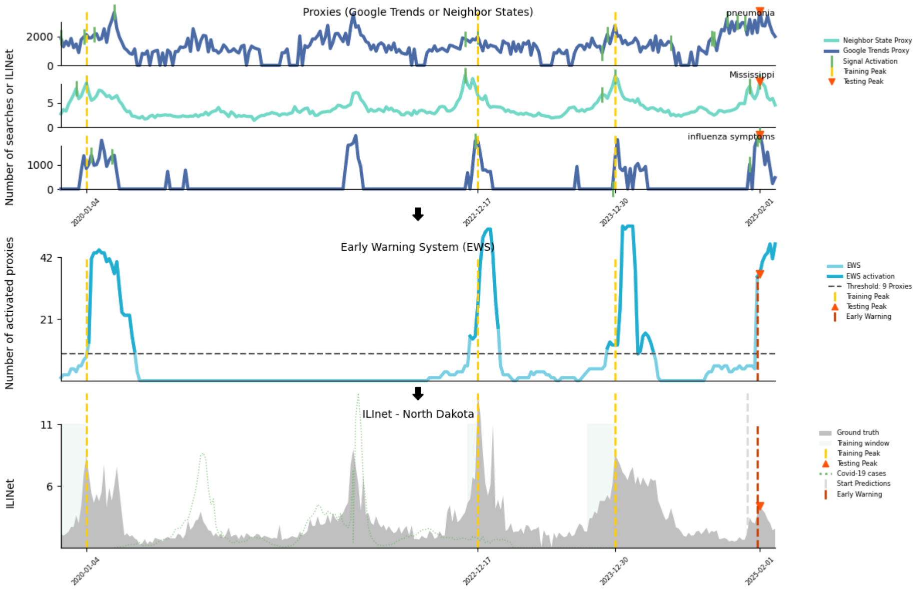


**Fig. S90**: Peak detection for North Dakota during the 2024-2025 season

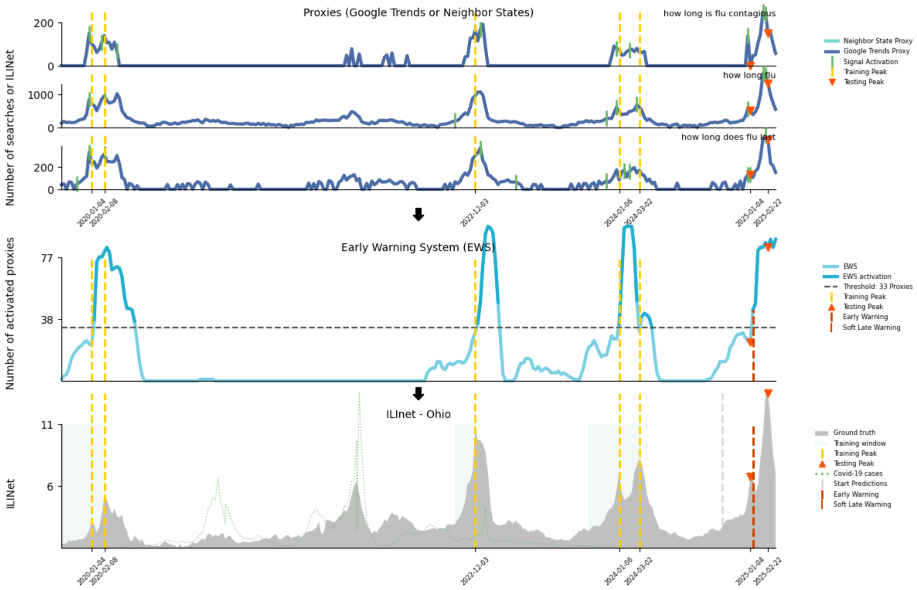


**Fig. S91**: Peak detection for Ohio during the 2024-2025 season

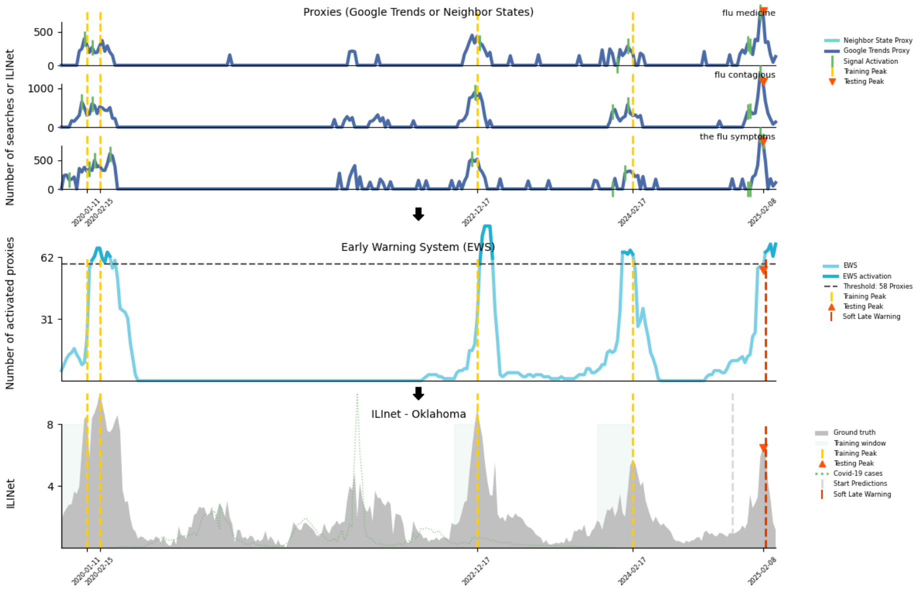


**Fig. S92**: Peak detection for Oklahoma during the 2024-2025 season

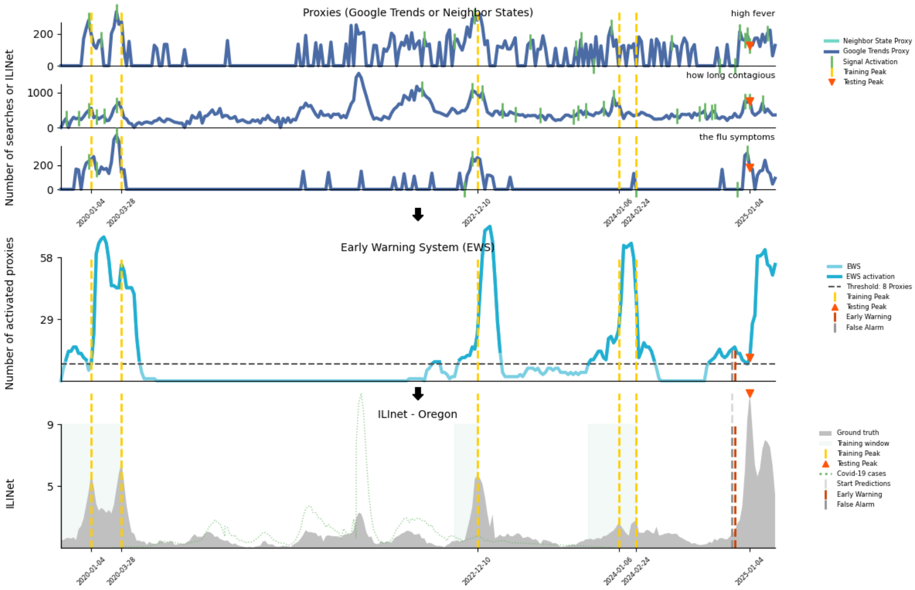


**Fig. S93**: Peak detection for Oregon during the 2024-2025 season

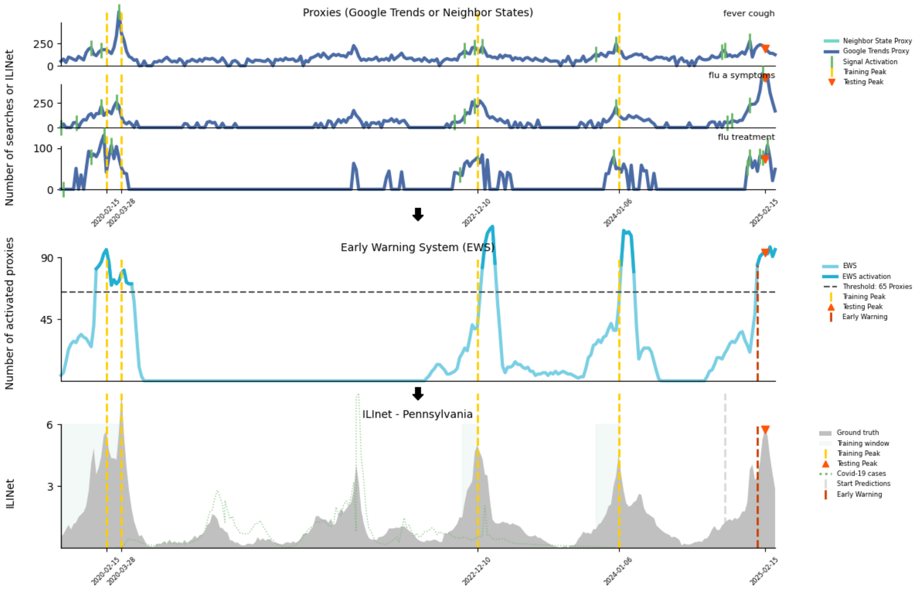


**Fig. S94**: Peak detection for Pennsylvania during the 2024-2025 season

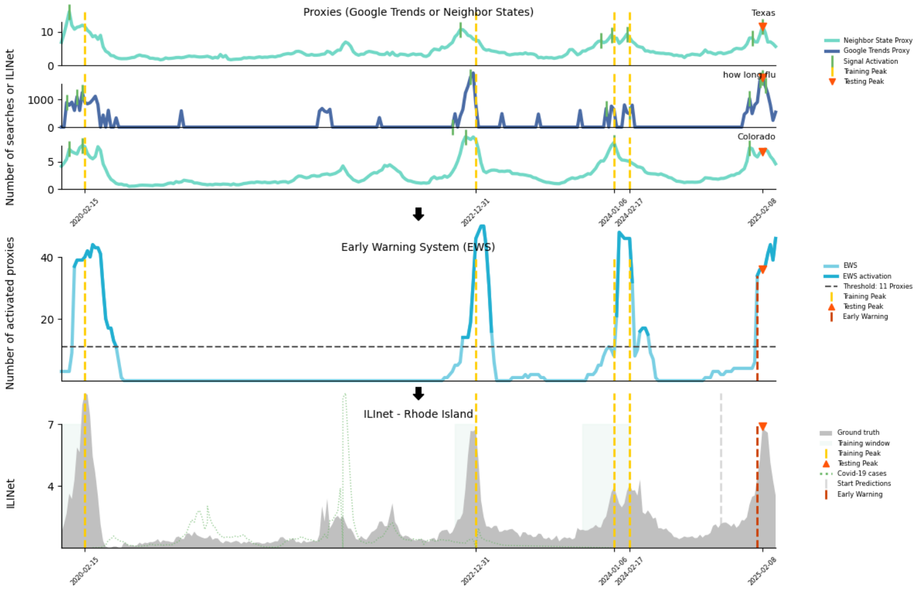


**Fig. S95**: Peak detection for Rhode Island during the 2024-2025 season

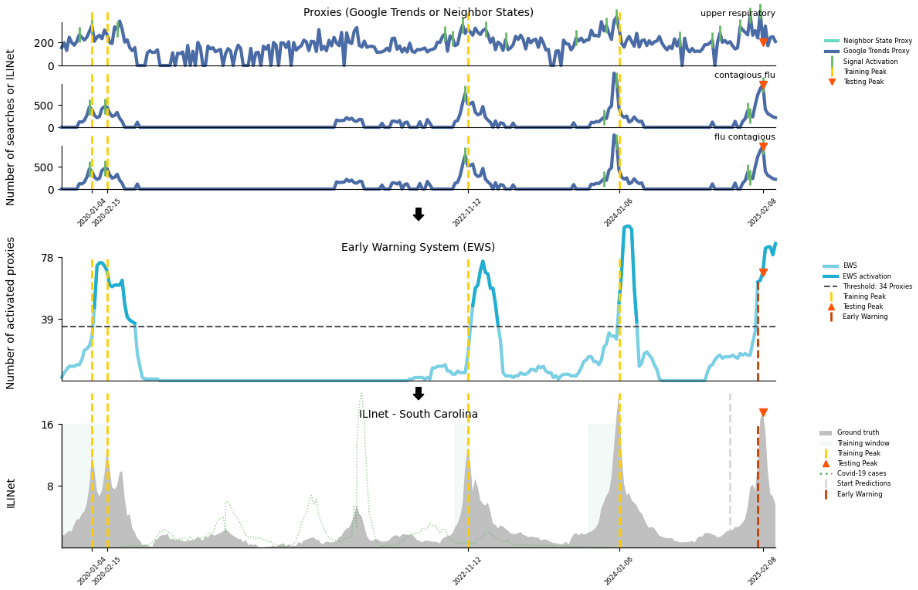


**Fig. S96**: Peak detection for South Carolina during the 2024-2025 season

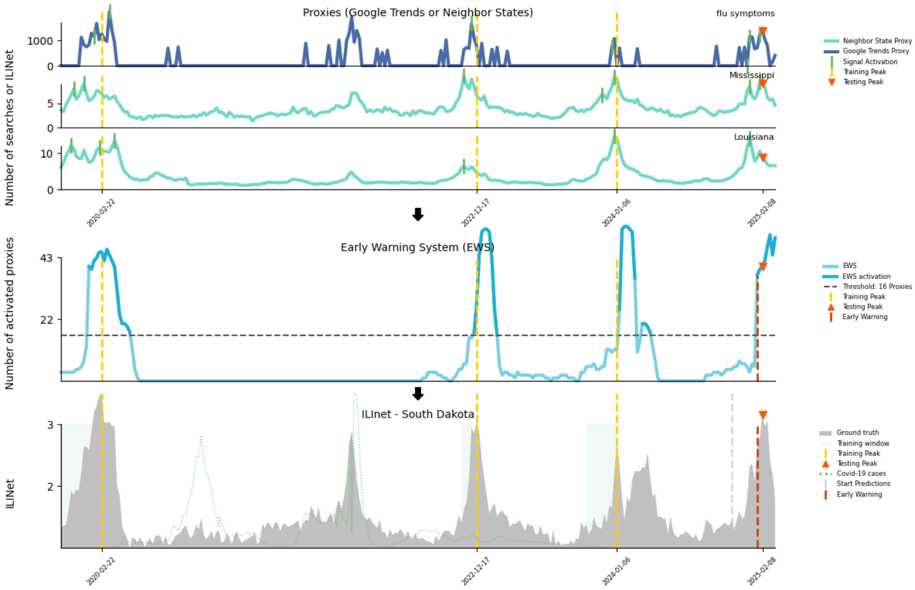


**Fig. S97**: Peak detection for South Dakota during the 2024-2025 season

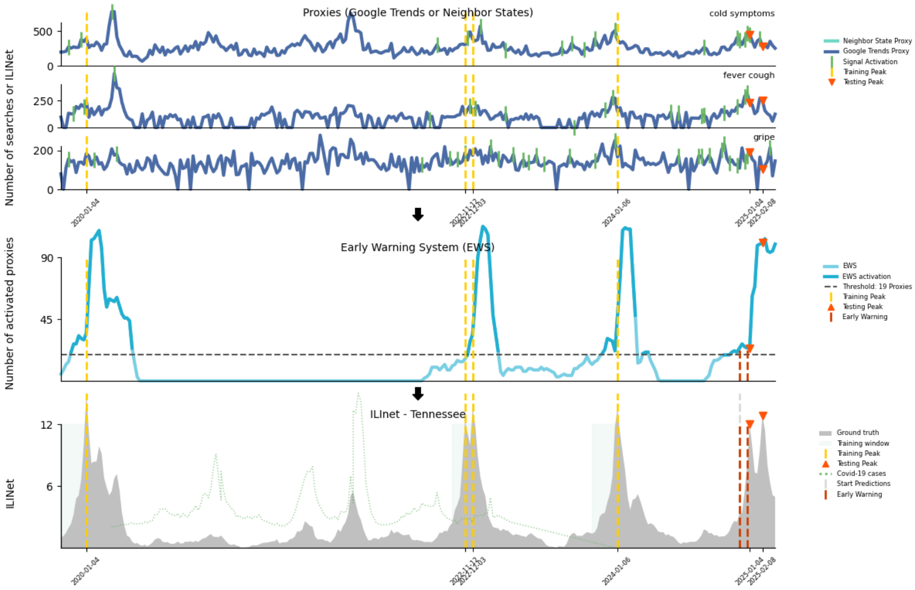


**Fig. S98**: Peak detection for Tennessee during the 2024-2025 season

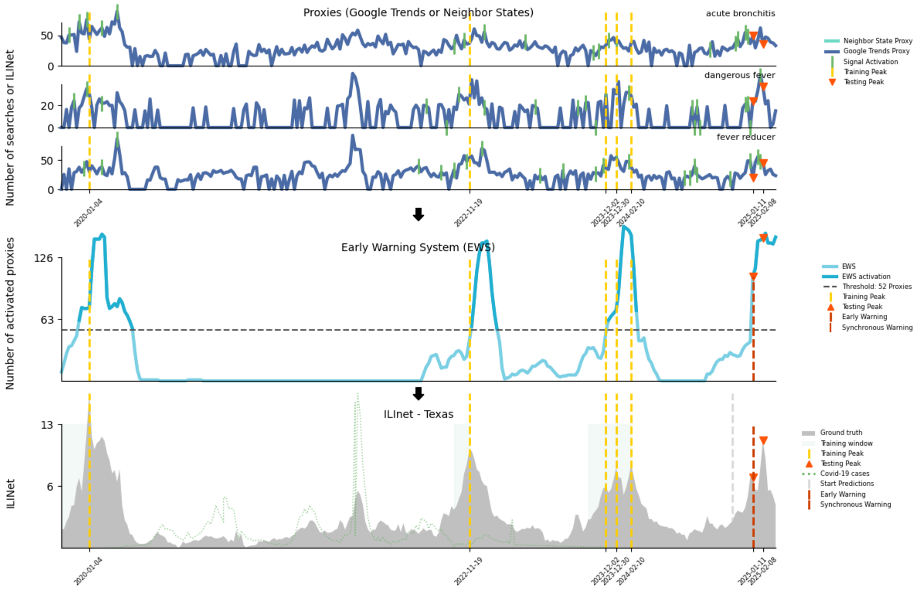


**Fig. S99**: Peak detection for Texas during the 2024-2025 season

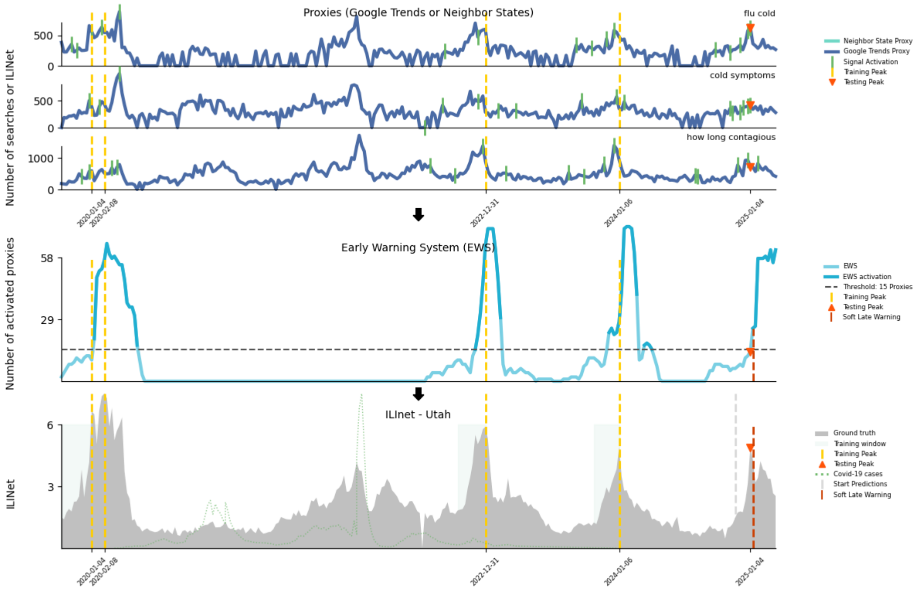


**Fig. S100**: Peak detection for Utah during the 2024-2025 season

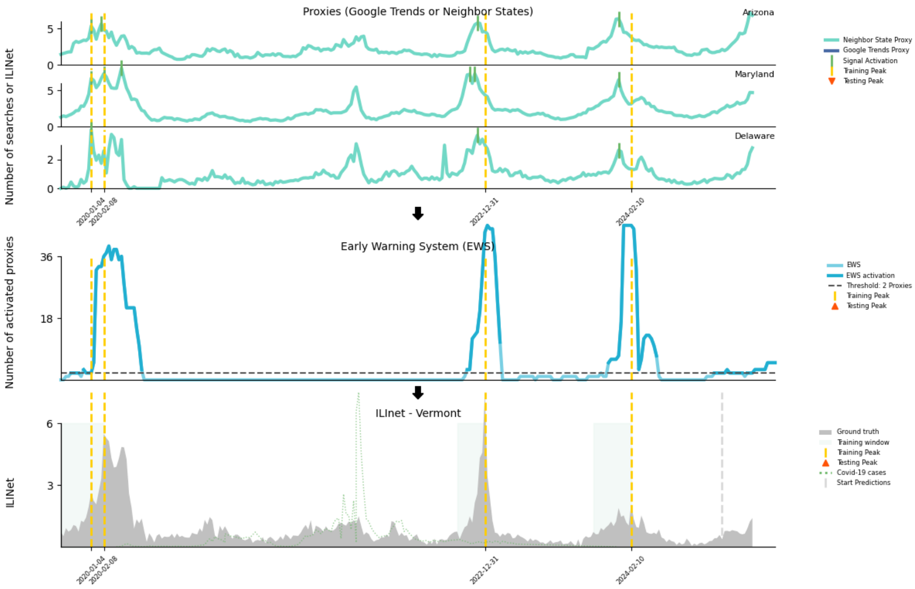


**Fig. S101**: Peak detection for Vermont during the 2024-2025 season

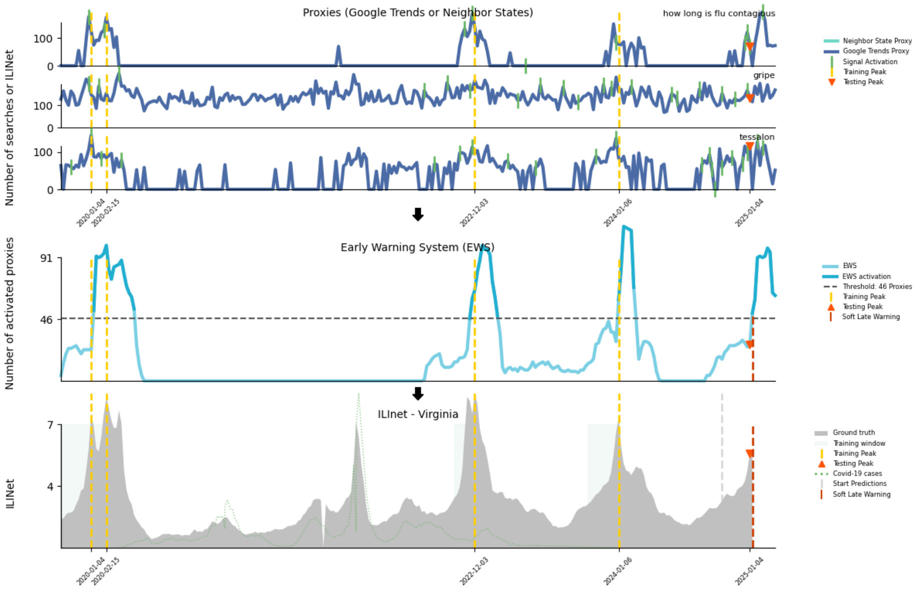


**Fig. S102**: Peak detection for Virginia during the 2024-2025 season

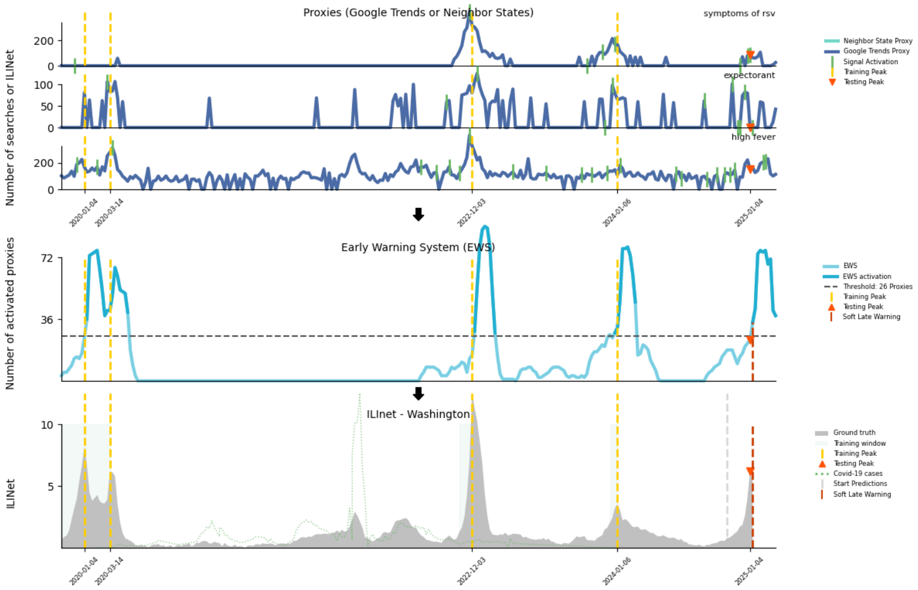


**Fig. S103**: Peak detection for Washington during the 2024-2025 season

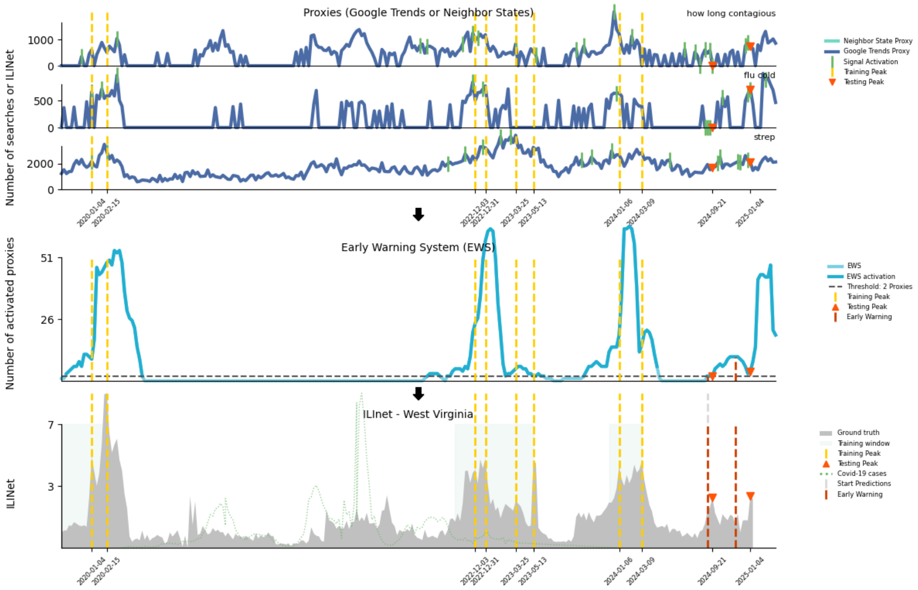


**Fig. S104**: Peak detection for West Virginia during the 2024-2025 season

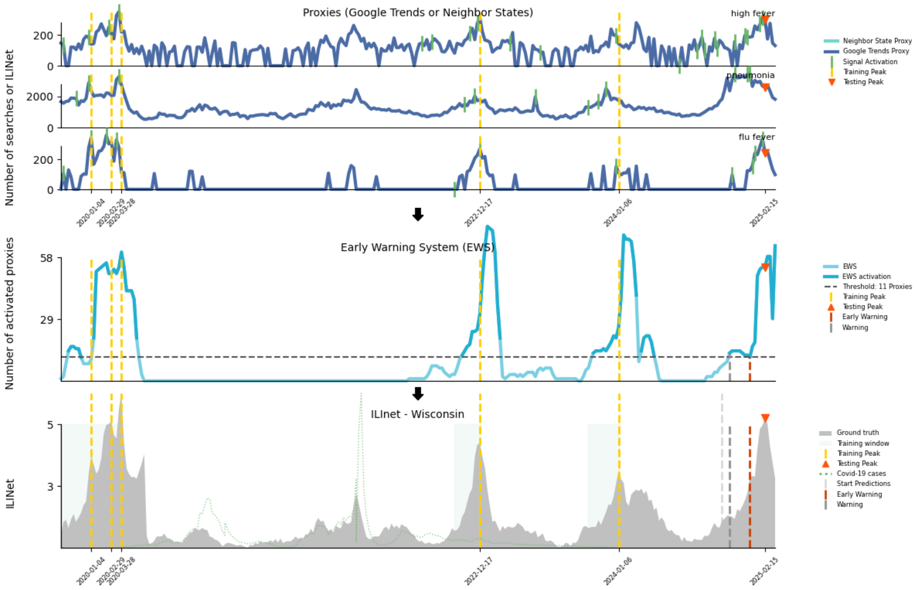


**Fig. S105**: Peak detection for Wisconsin during the 2024-2025 season

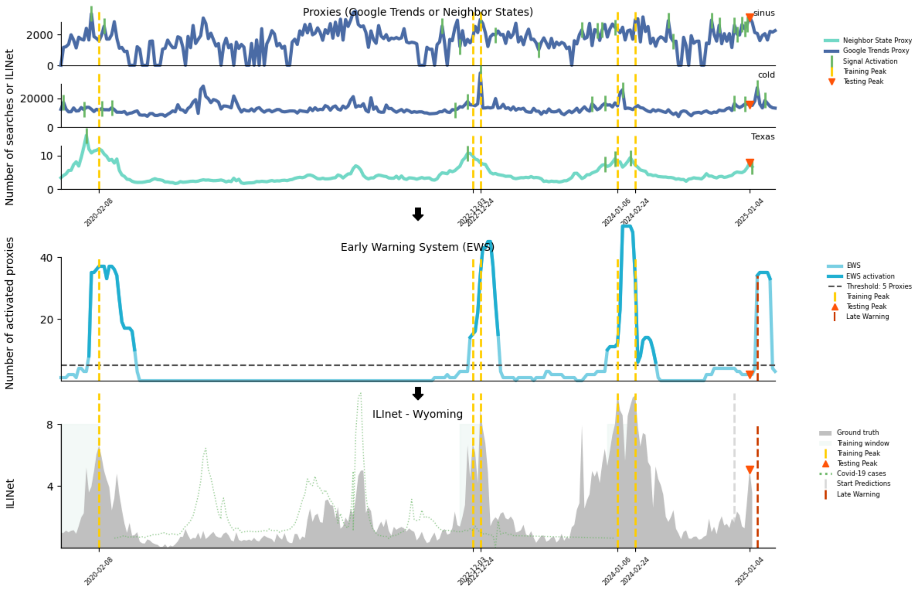


**Fig. S106**: Peak detection for Wyoming during the 2024-2025 season

#### Granger Causality Analysis

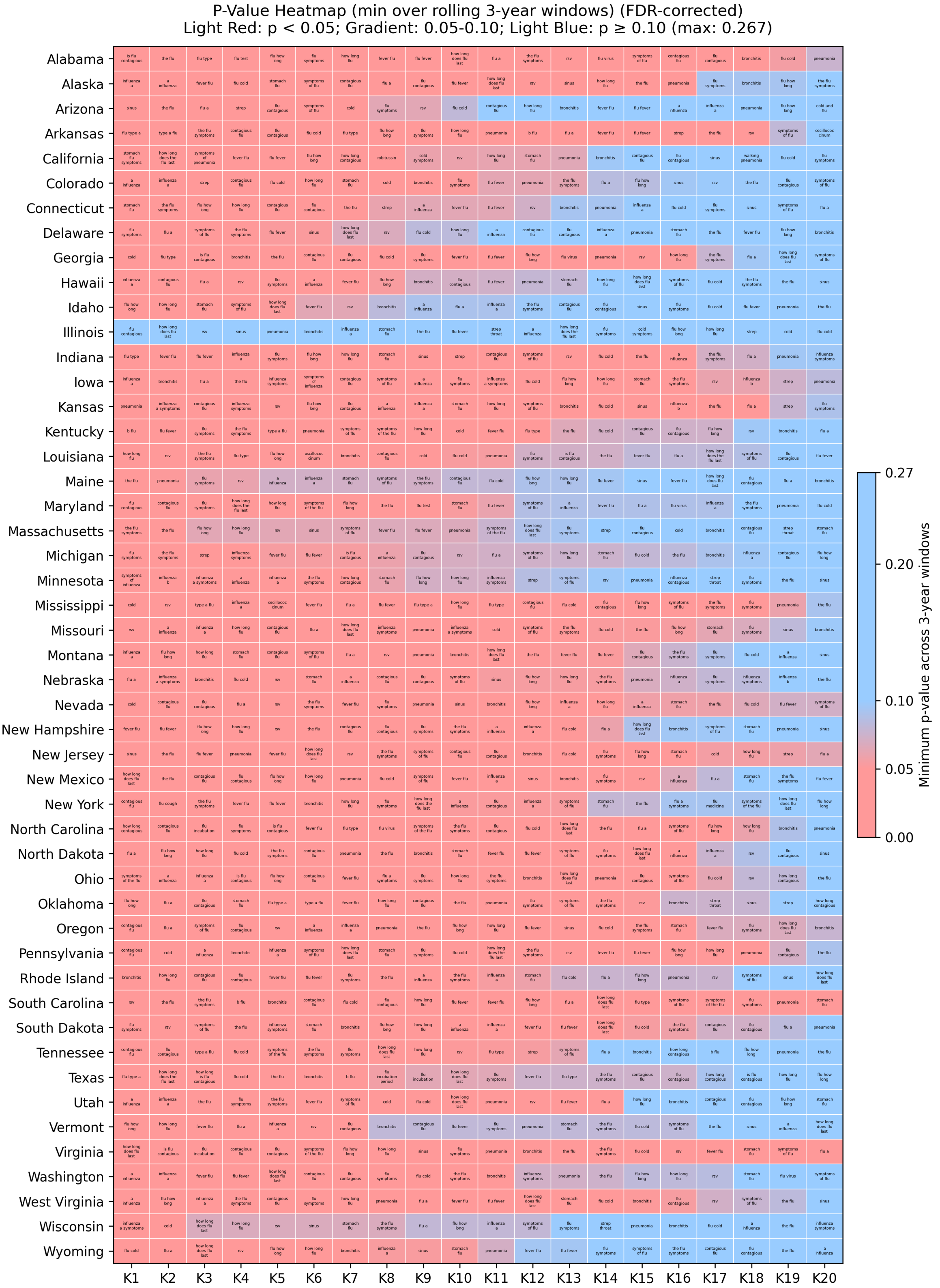


**Fig. S107**: **Granger causality $p$-value heatmap (FDR-corrected).** Heatmap showing the minimum $p$-values across rolling 3-year windows for each state (rows) and each keyword (columns), after applying false discovery rate (FDR) correction. Light red indicates statistical significance ($p < 0.05$), gradient red corresponds to $0.05 \leq p < 0.10$, and light blue indicates non-significant associations ($p \geq 0.10$).

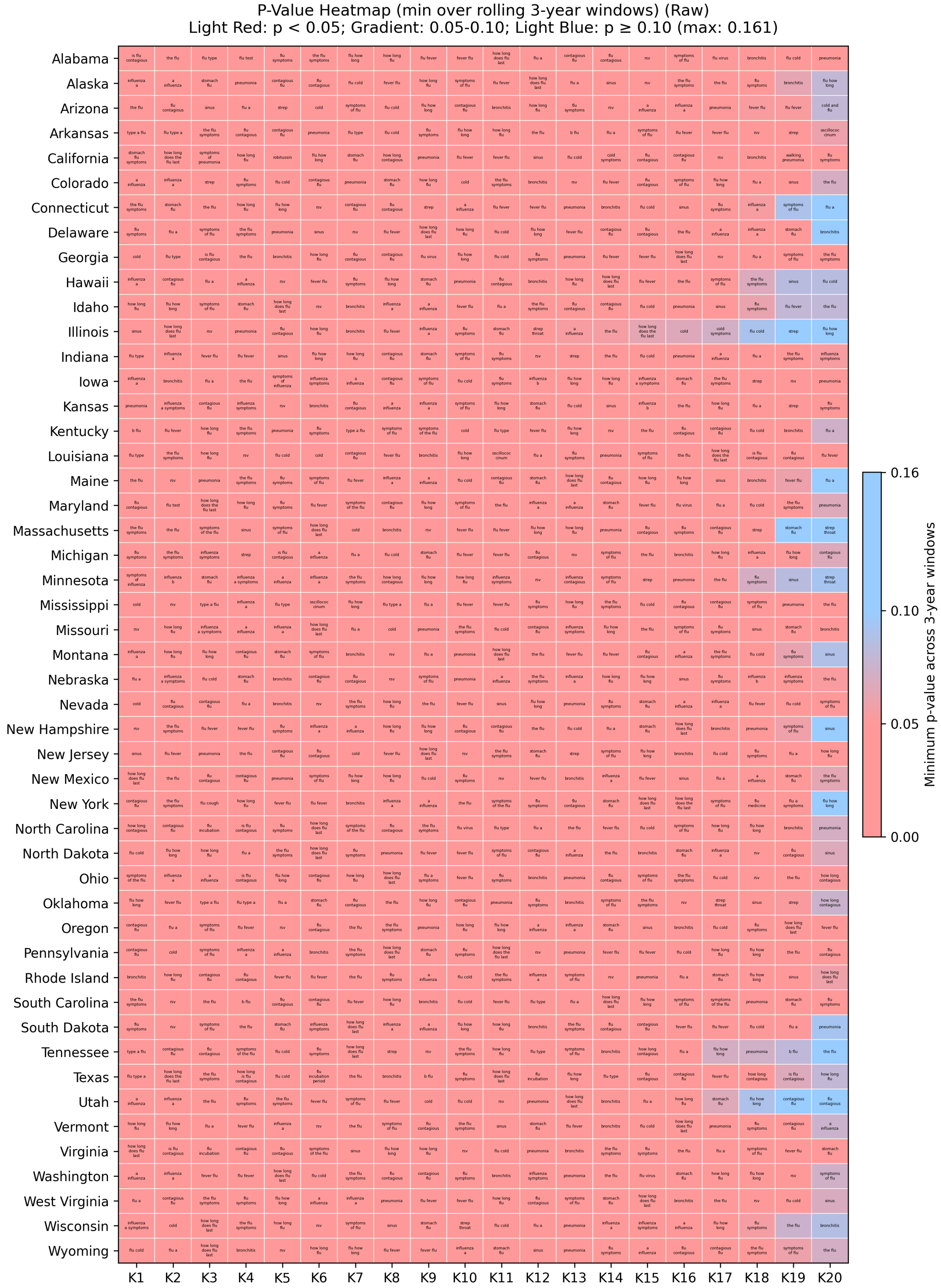


**Fig. S108**: **Granger causality $p$-value heatmap (raw, uncorrected).** Same as Figure S107, but showing raw $p$-values without FDR correction.

#### Onset Detection Methodology

**Algorithm 1** Onset detection pseudocode

```
function GET_ONSET(target_name, lambda_vector, timeseries, onset_hypervalue=3 (6-week window), gap_tolerance=4, min_week_length=2)
    Initialize empty lists: start_dates, end_dates, proxy_names, location_names
    for each column (proxy) in lambda_vector do
        Extract coefficient values and corresponding dates
        Convert coefficients into binary list: 1 if coefficient > 1 (growth), else 0
        Find consecutive windows of 1's: track start and end dates of these "onset candidate windows"
        Merge windows if they are less than gap_tolerance (4) weeks apart
        if column is the target location (process for targets, look into future) then
            for each window do
                if window ≥ min_week_length (2) weeks long then
                    Scan through window dates
                    Check if cases increase on average in next onset_hypervalue (3) weeks
                    if true then
                        if window length > 2 × onset_hypervalue (3) weeks then
                            Record start_date = current date + onset_hypervalue (3)
                            Record end_date = window_end (large window, large outbreak)
                        else
                            Record start_date = date, end_date = window_end (short window, noisy short outbreak)
                        end if
                    end if
                end if
            end for
        else column is a predictor (mimic process for targets, without look into future)
            for each window (proxy column, mimic target process without looking into future) do
                if window length ≥ 2 × onset_hypervalue (3) then
                    Record start_date = window_start + 2 × onset_hypervalue (total shift of 6 weeks)
                    Record end_date = window_end
                end if
            end for
        end if
    end for
end function
```

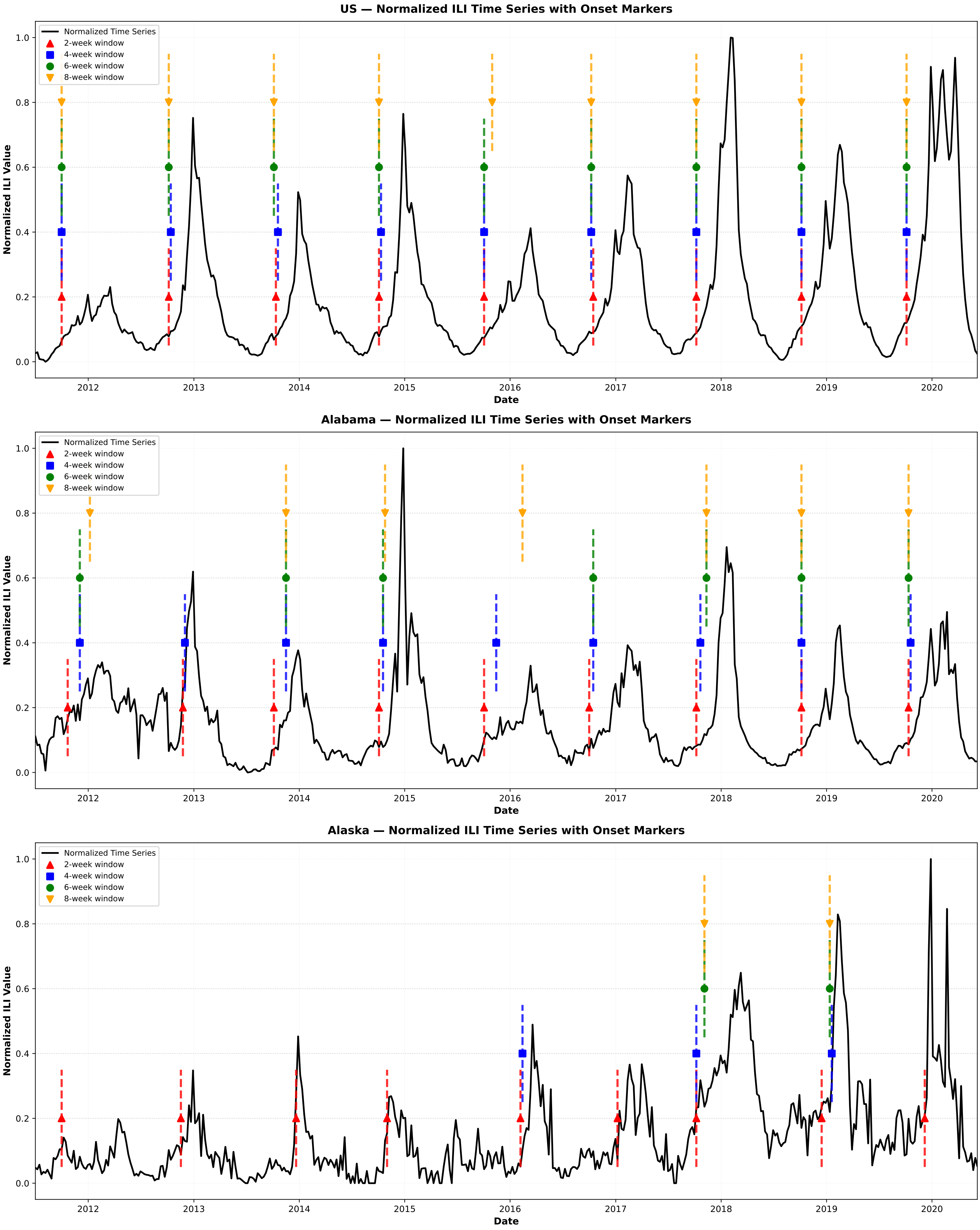

US — Normalized ILI Time Series with Onset Markers
Normalized Time Series
2-week window
4-week window
6-week window
8-week window
Normalized ILI Value
Date
Alabama — Normalized ILI Time Series with Onset Markers
Alaska — Normalized ILI Time Series with Onset Markers
0.0
0.2
0.4
0.6
0.8
1.0
2012
2013
2014
2015
2016
2017
2018
2019
2020

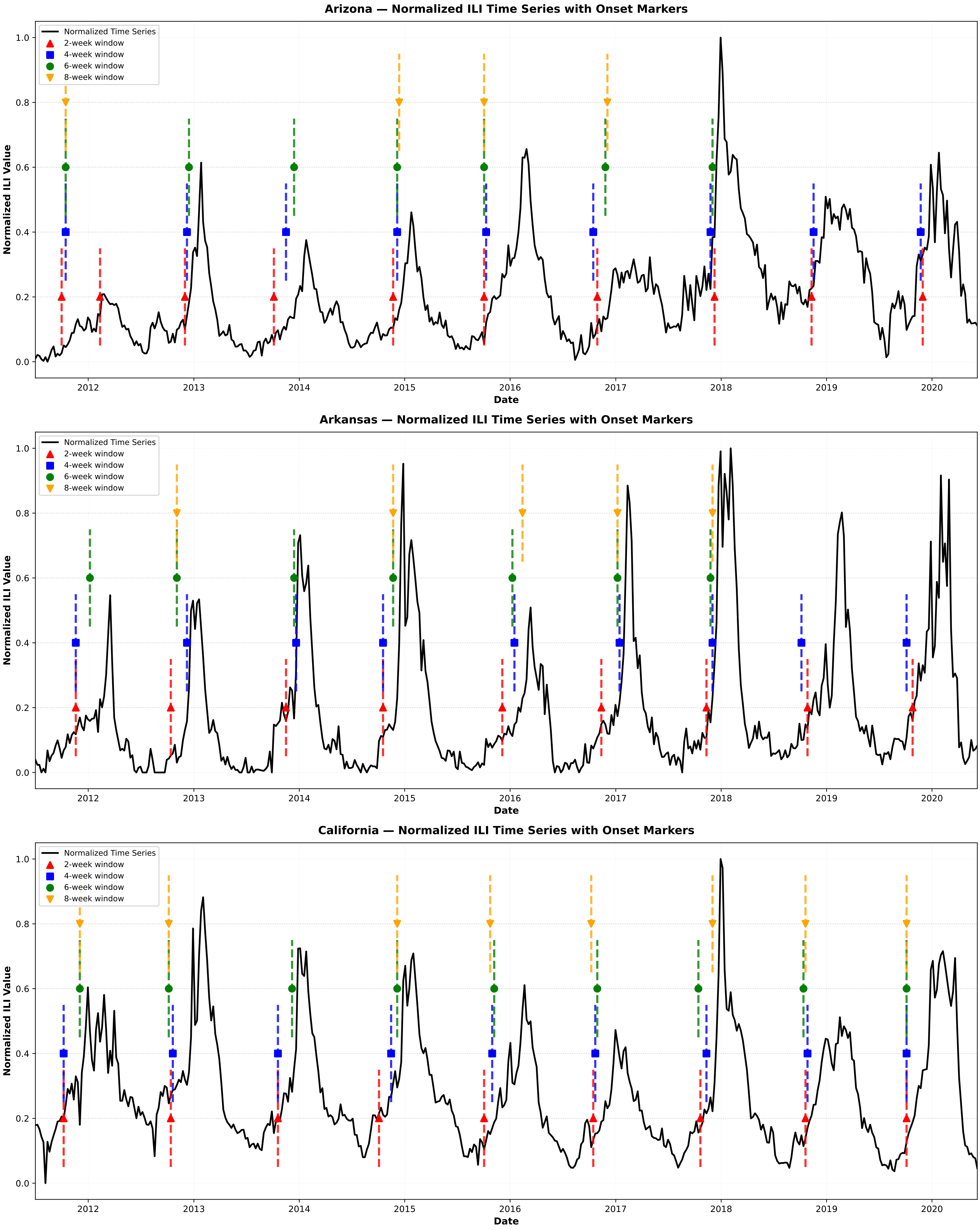

Arizona — Normalized ILI Time Series with Onset Markers
Arkansas — Normalized ILI Time Series with Onset Markers
California — Normalized ILI Time Series with Onset Markers
Normalized Time Series
2-week window
4-week window
6-week window
8-week window
Normalized ILI Value
Date
0.0
0.2
0.4
0.6
0.8
1.0
2012
2013
2014
2015
2016
2017
2018
2019
2020

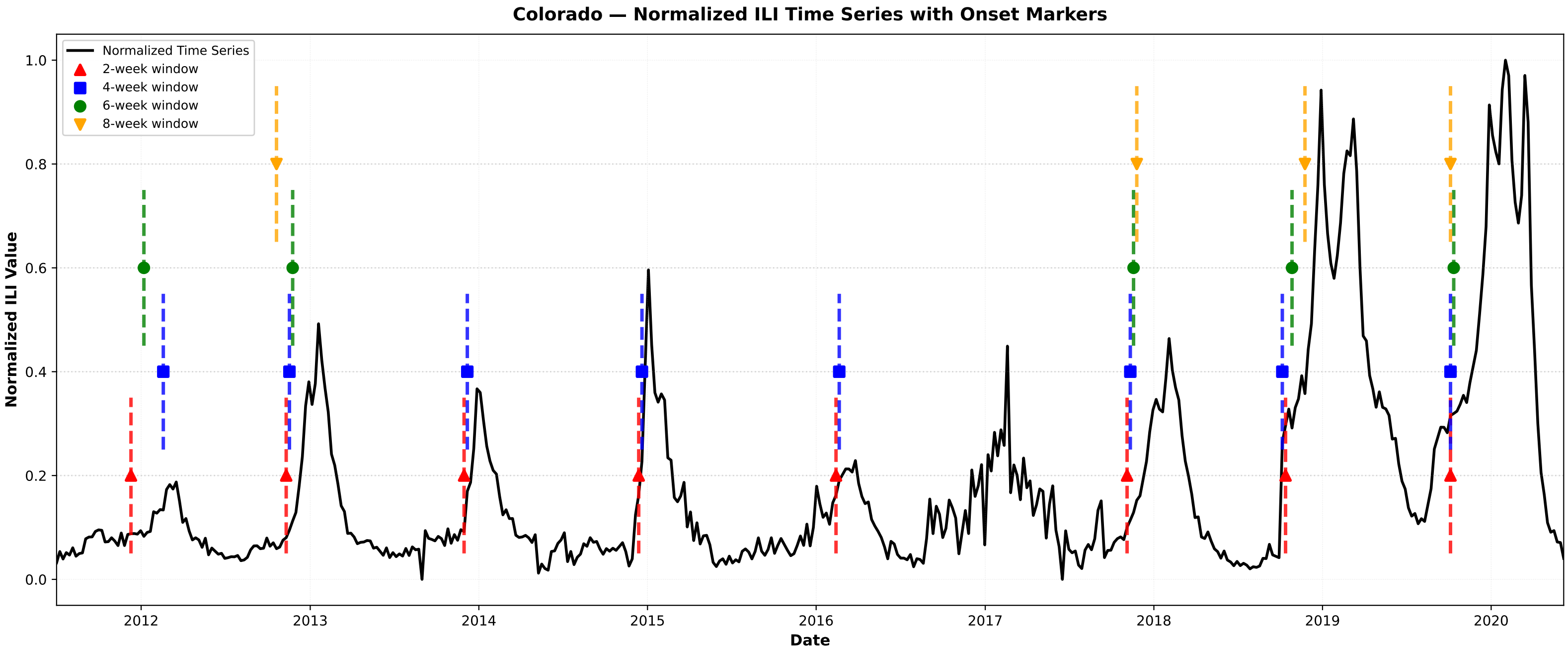

Colorado — Normalized ILI Time Series with Onset Markers
Normalized Time Series
2-week window
4-week window
6-week window
8-week window
Normalized ILI Value
Date


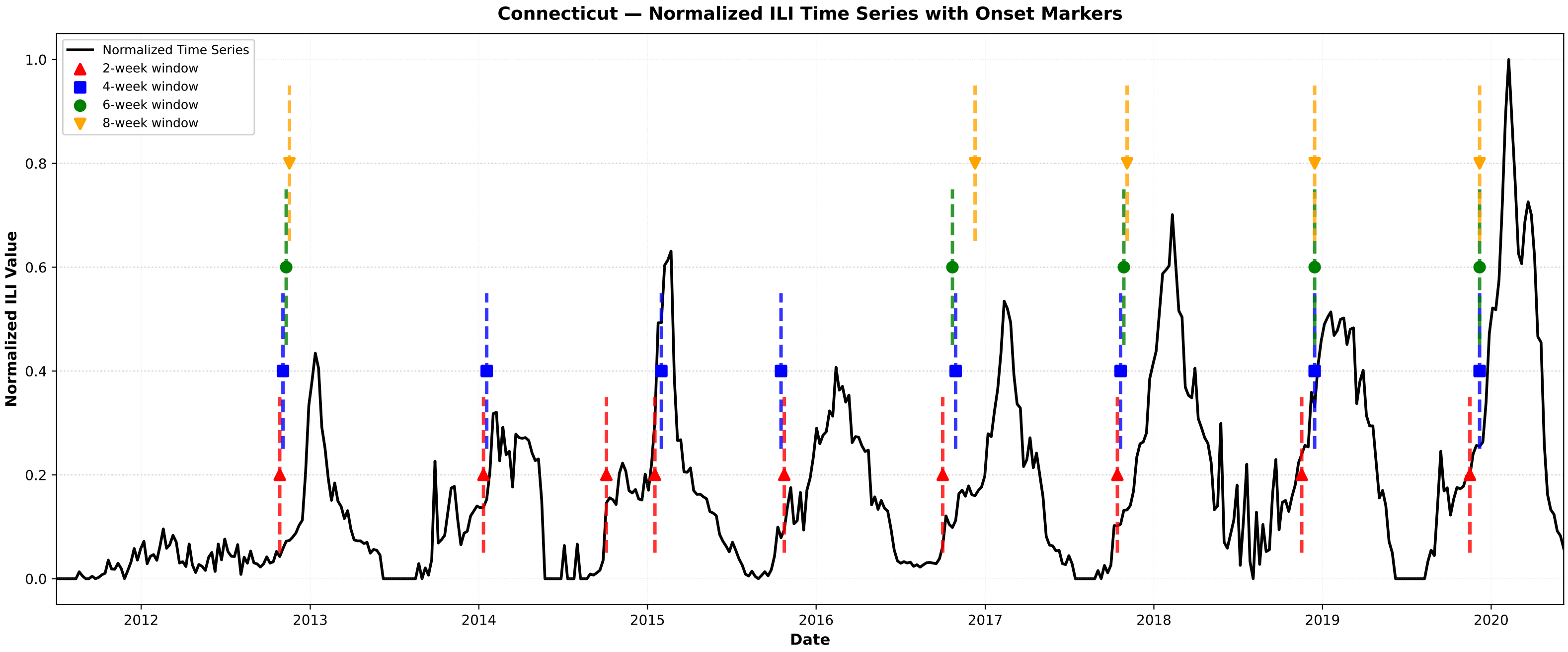

Connecticut — Normalized ILI Time Series with Onset Markers
Normalized Time Series
2-week window
4-week window
6-week window
8-week window
Normalized ILI Value
Date


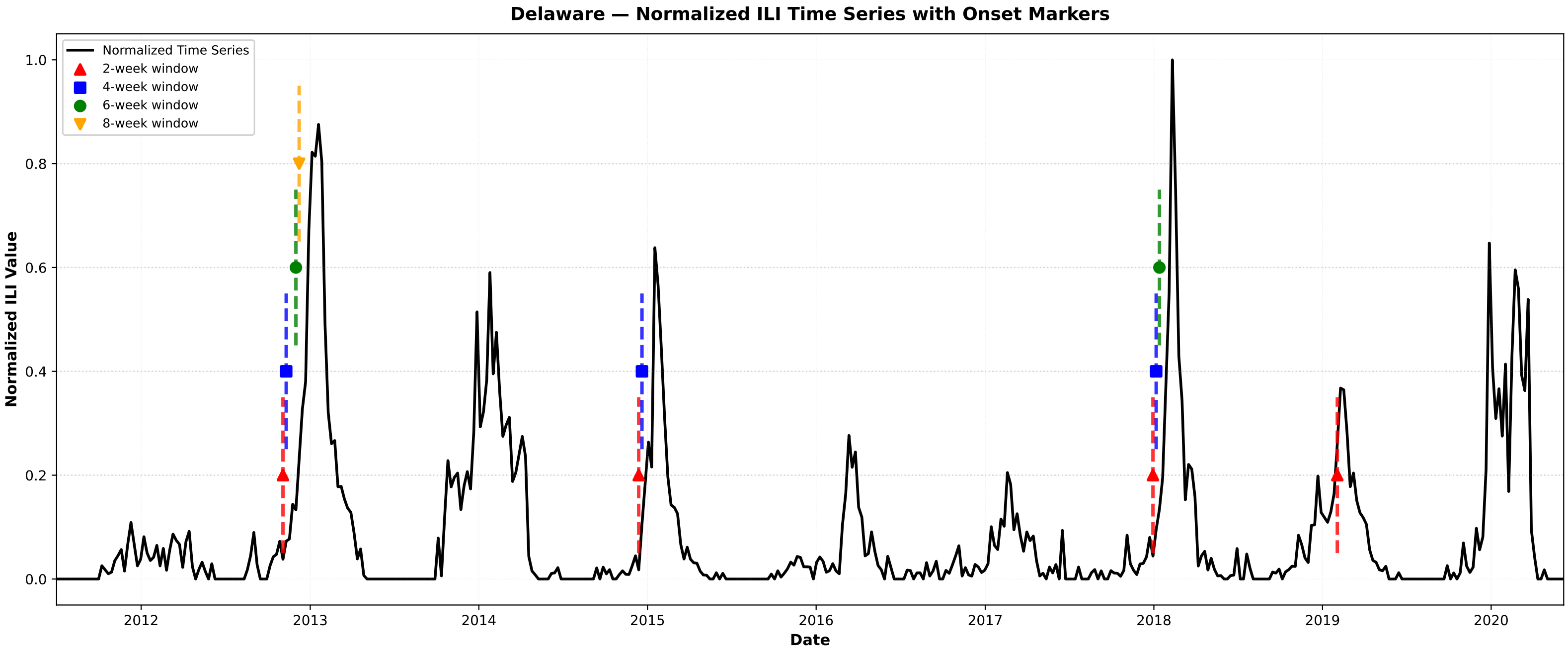

Delaware — Normalized ILI Time Series with Onset Markers
Normalized Time Series
2-week window
4-week window
6-week window
8-week window
Normalized ILI Value
Date

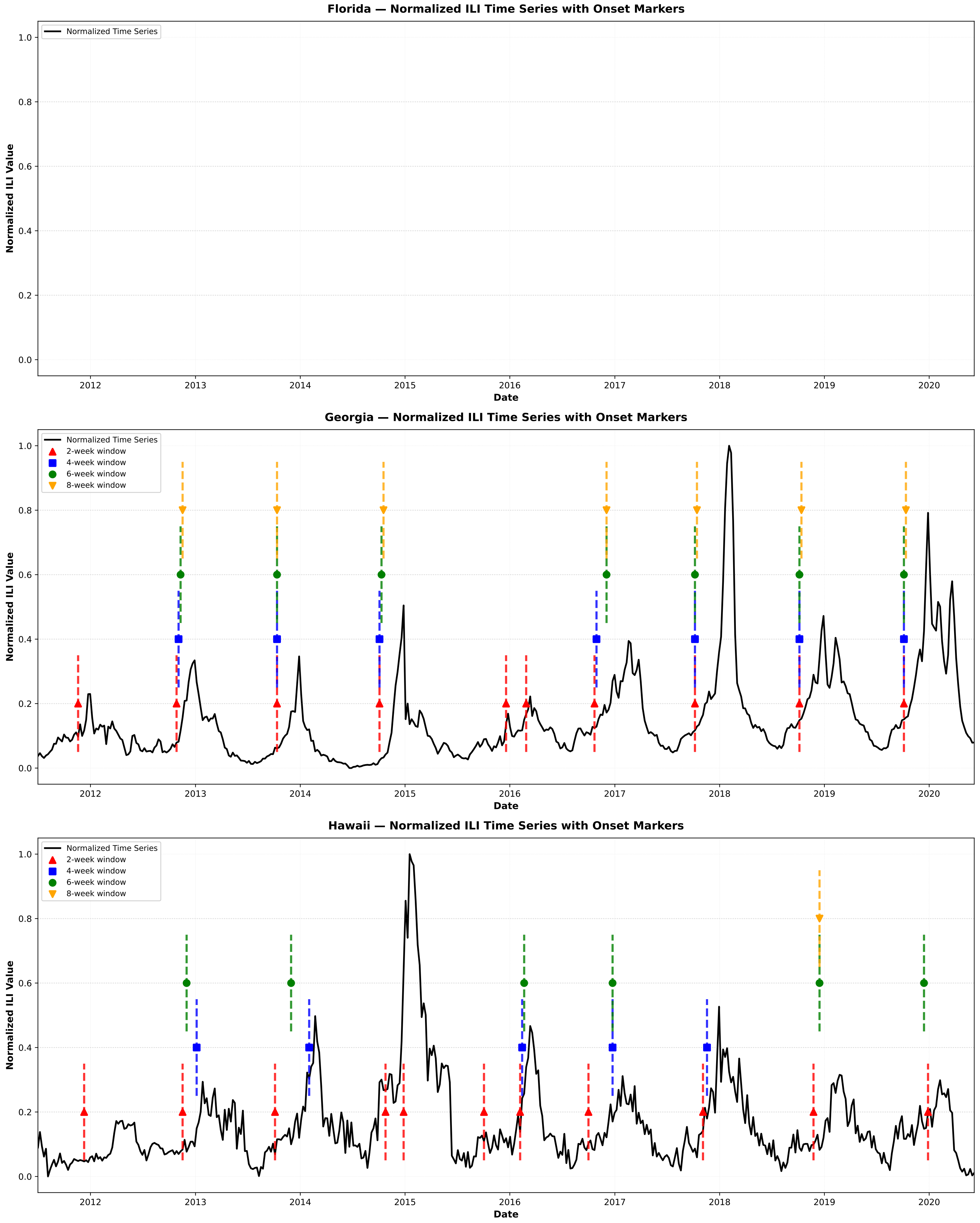

Florida — Normalized ILI Time Series with Onset Markers
Normalized Time Series
Normalized ILI Value
Date
Georgia — Normalized ILI Time Series with Onset Markers
Normalized Time Series
2-week window
4-week window
6-week window
8-week window
Normalized ILI Value
Date
Hawaii — Normalized ILI Time Series with Onset Markers
Normalized Time Series
2-week window
4-week window
6-week window
8-week window
Normalized ILI Value
Date

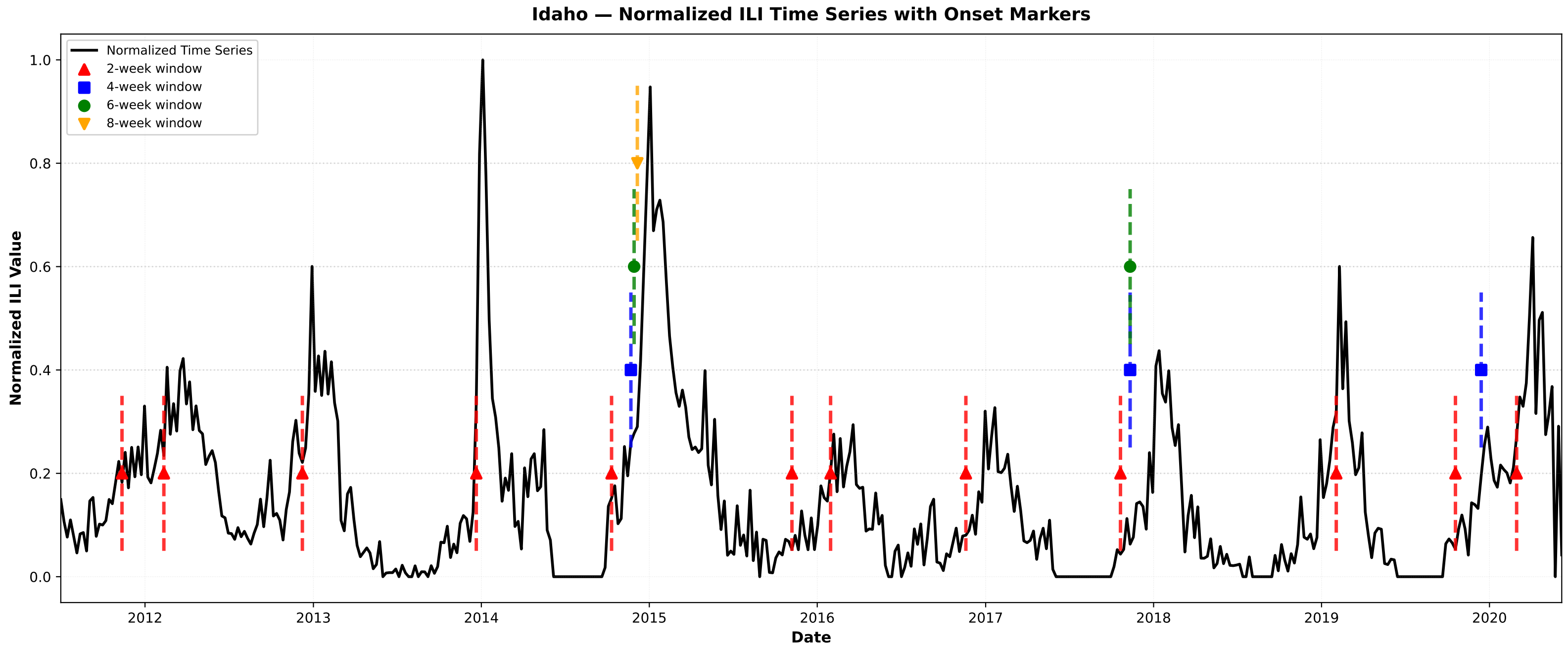
Idaho — Normalized ILI Time Series with Onset Markers
Normalized Time Series
2-week window
4-week window
6-week window
8-week window
Normalized ILI Value
0.0
0.2
0.4
0.6
0.8
1.0
2012
2013
2014
2015
2016
2017
2018
2019
2020
Date

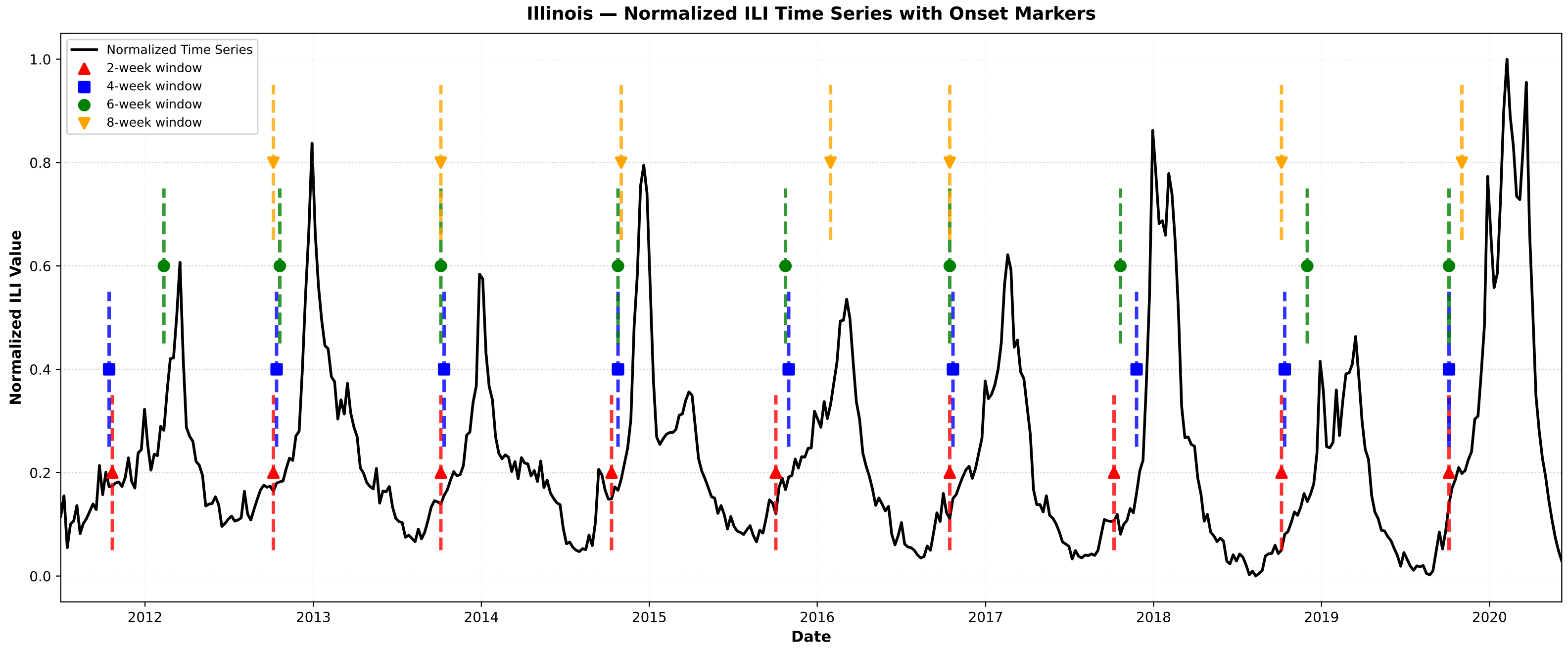
Illinois — Normalized ILI Time Series with Onset Markers
Normalized Time Series
2-week window
4-week window
6-week window
8-week window
Normalized ILI Value
0.0
0.2
0.4
0.6
0.8
1.0
2012
2013
2014
2015
2016
2017
2018
2019
2020
Date

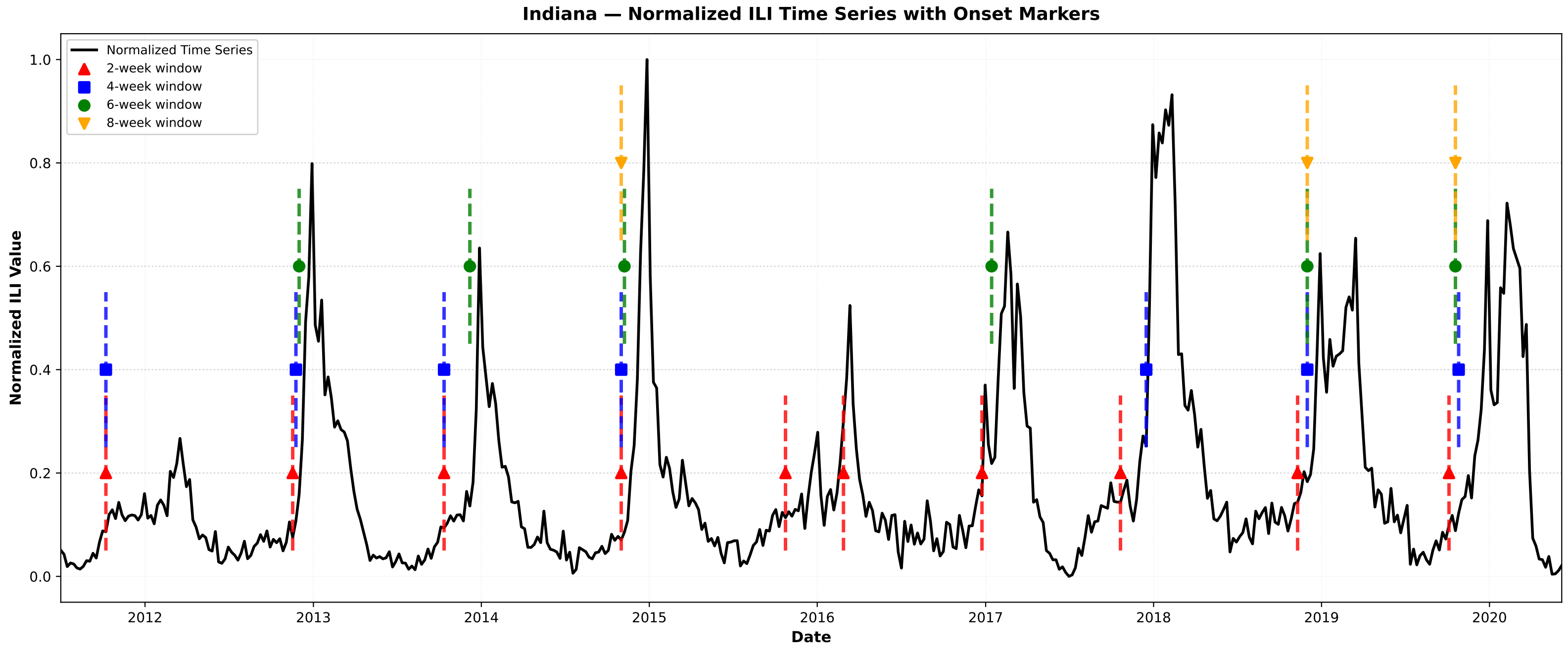
Indiana — Normalized ILI Time Series with Onset Markers
Normalized Time Series
2-week window
4-week window
6-week window
8-week window
Normalized ILI Value
0.0
0.2
0.4
0.6
0.8
1.0
2012
2013
2014
2015
2016
2017
2018
2019
2020
Date

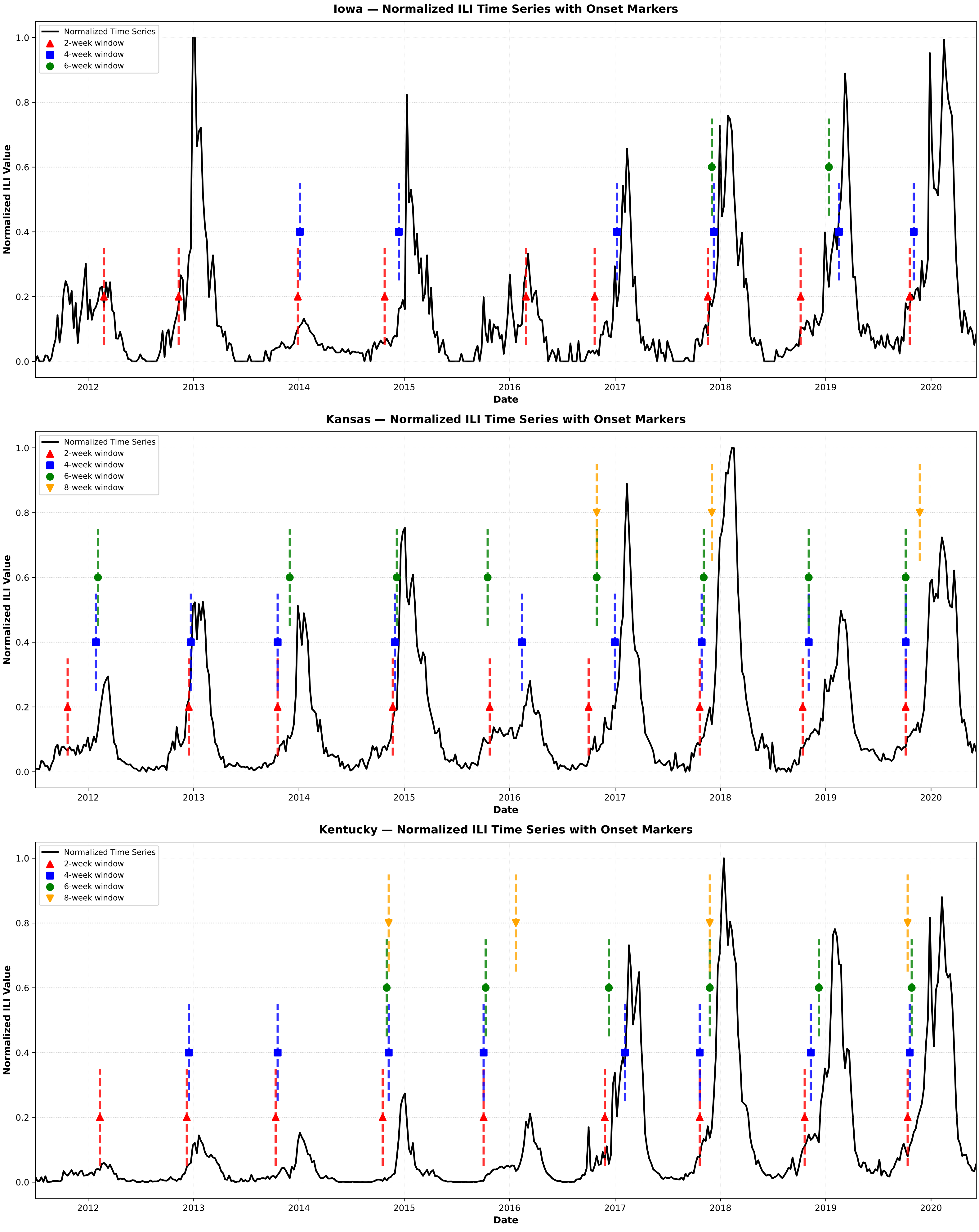

Iowa — Normalized ILI Time Series with Onset Markers
Normalized Time Series
2-week window
4-week window
6-week window
Normalized ILI Value
Date
Kansas — Normalized ILI Time Series with Onset Markers
Normalized Time Series
2-week window
4-week window
6-week window
8-week window
Normalized ILI Value
Date
Kentucky — Normalized ILI Time Series with Onset Markers
Normalized Time Series
2-week window
4-week window
6-week window
8-week window
Normalized ILI Value
Date

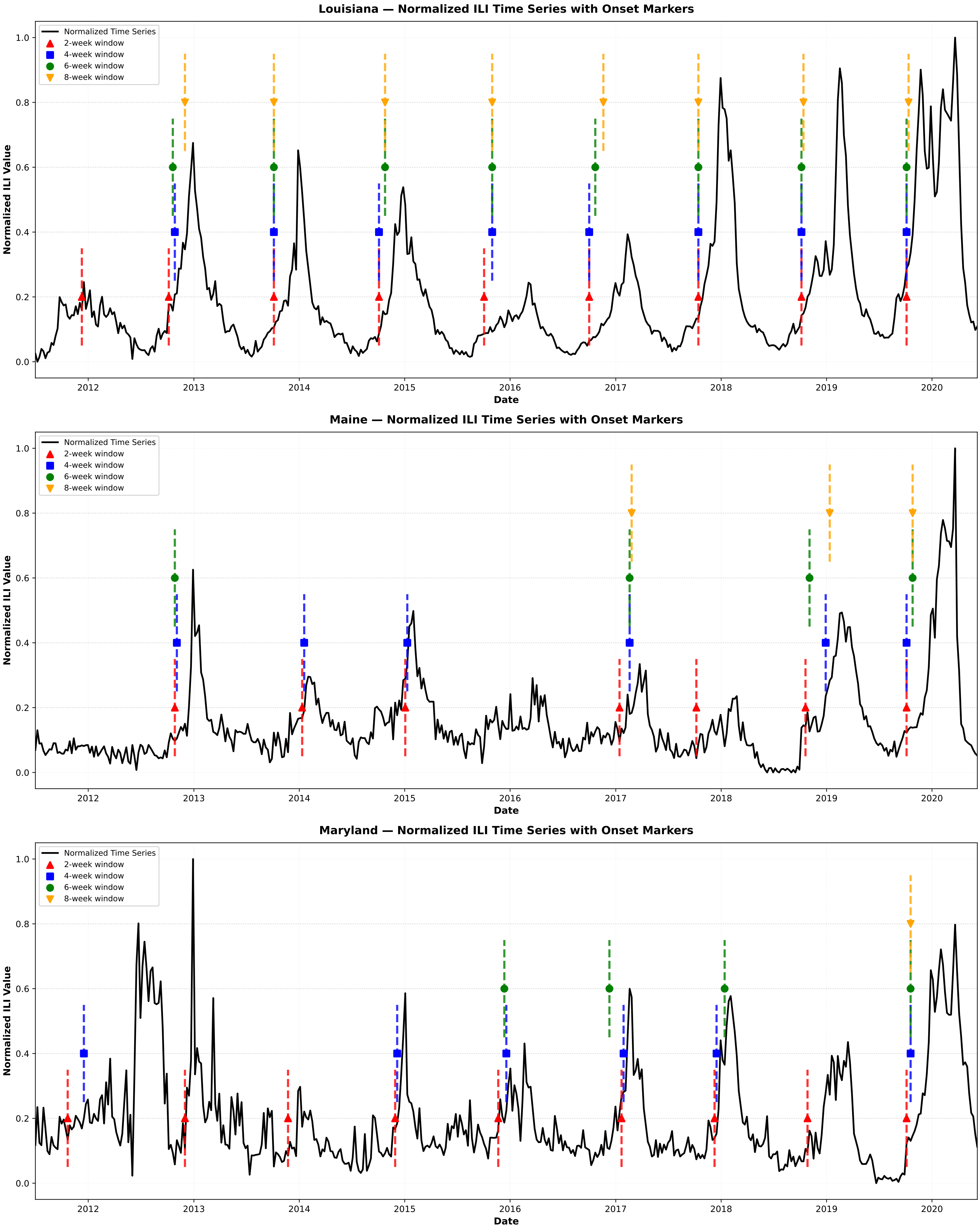

Louisiana — Normalized ILI Time Series with Onset Markers
Normalized Time Series
2-week window
4-week window
6-week window
8-week window
Normalized ILI Value
Date
Maine — Normalized ILI Time Series with Onset Markers
Normalized Time Series
2-week window
4-week window
6-week window
8-week window
Normalized ILI Value
Date
Maryland — Normalized ILI Time Series with Onset Markers
Normalized Time Series
2-week window
4-week window
6-week window
8-week window
Normalized ILI Value
Date
0.0
0.2
0.4
0.6
0.8
1.0
2012
2013
2014
2015
2016
2017
2018
2019
2020

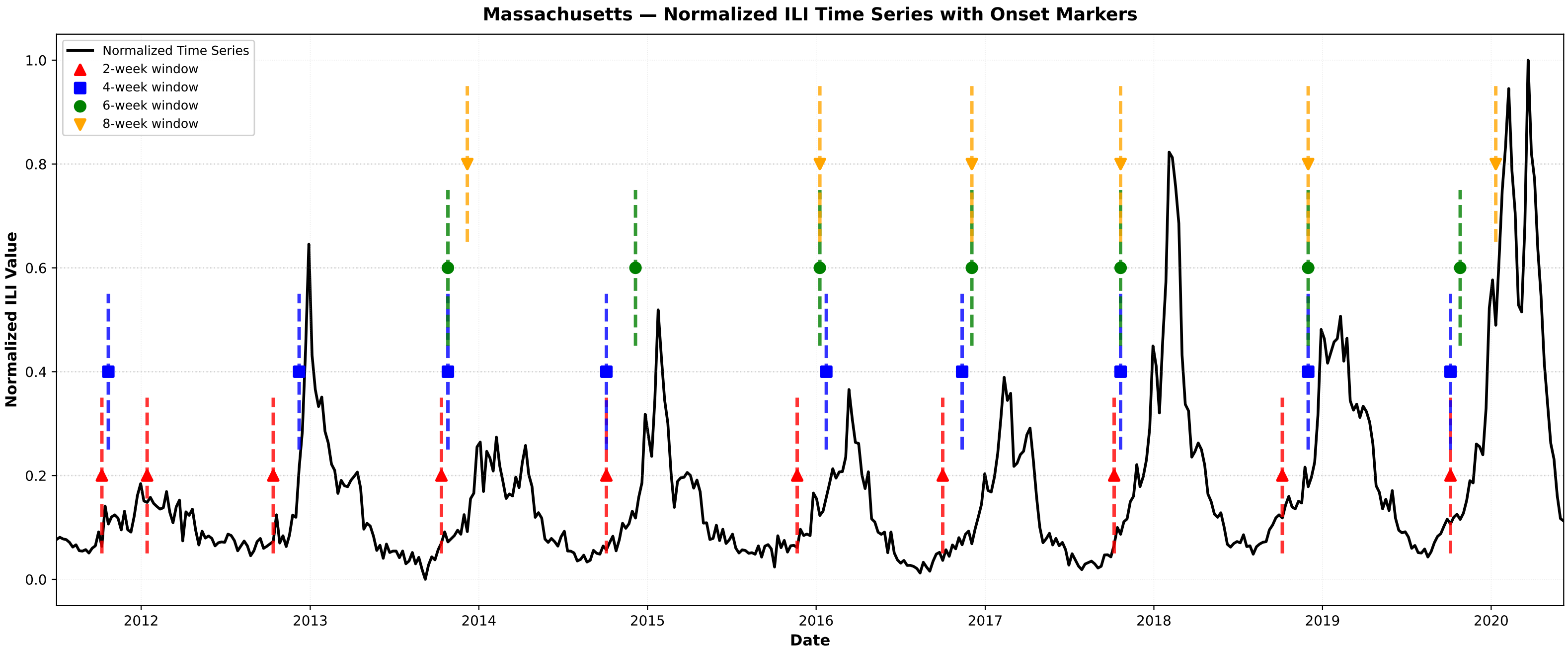
Massachusetts — Normalized ILI Time Series with Onset Markers
Normalized Time Series
2-week window
4-week window
6-week window
8-week window
Normalized ILI Value
Date
0.0
0.2
0.4
0.6
0.8
1.0
2012
2013
2014
2015
2016
2017
2018
2019
2020

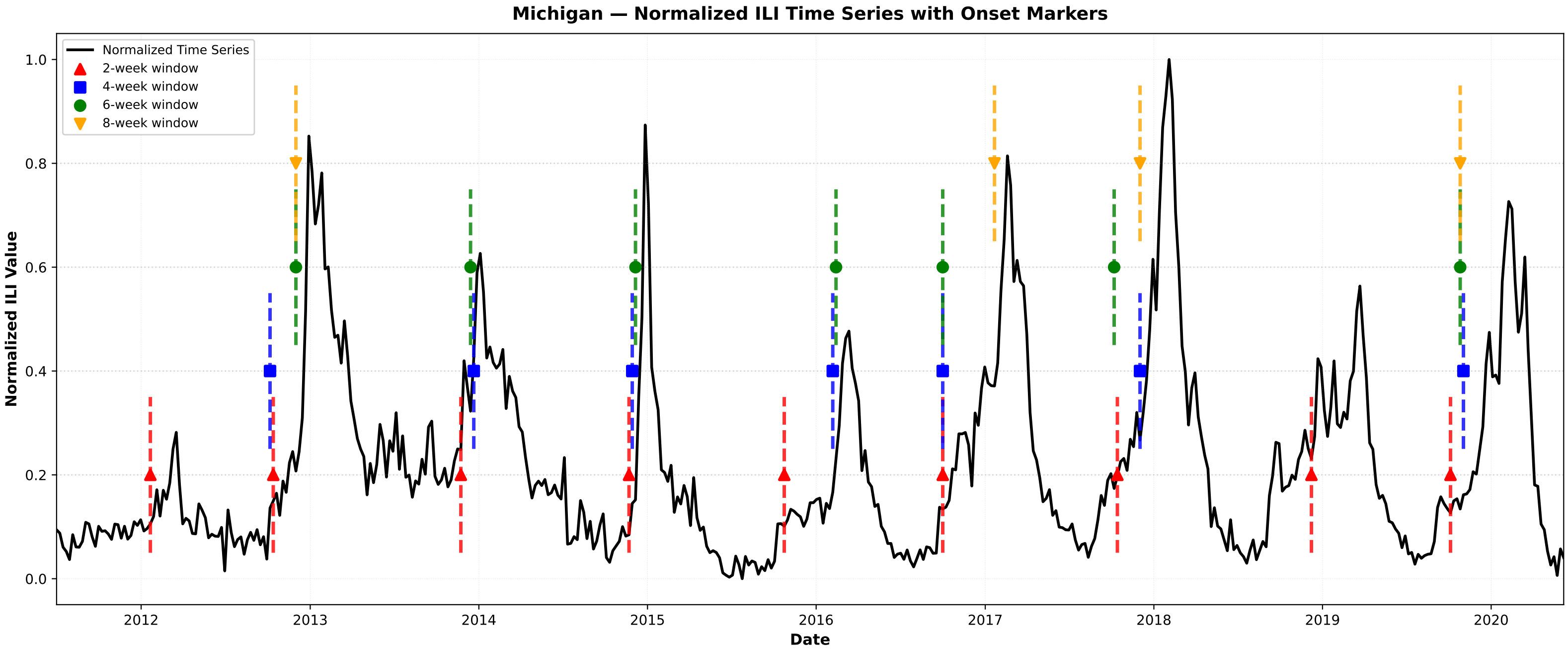
Michigan — Normalized ILI Time Series with Onset Markers
Normalized Time Series
2-week window
4-week window
6-week window
8-week window
Normalized ILI Value
Date
0.0
0.2
0.4
0.6
0.8
1.0
2012
2013
2014
2015
2016
2017
2018
2019
2020

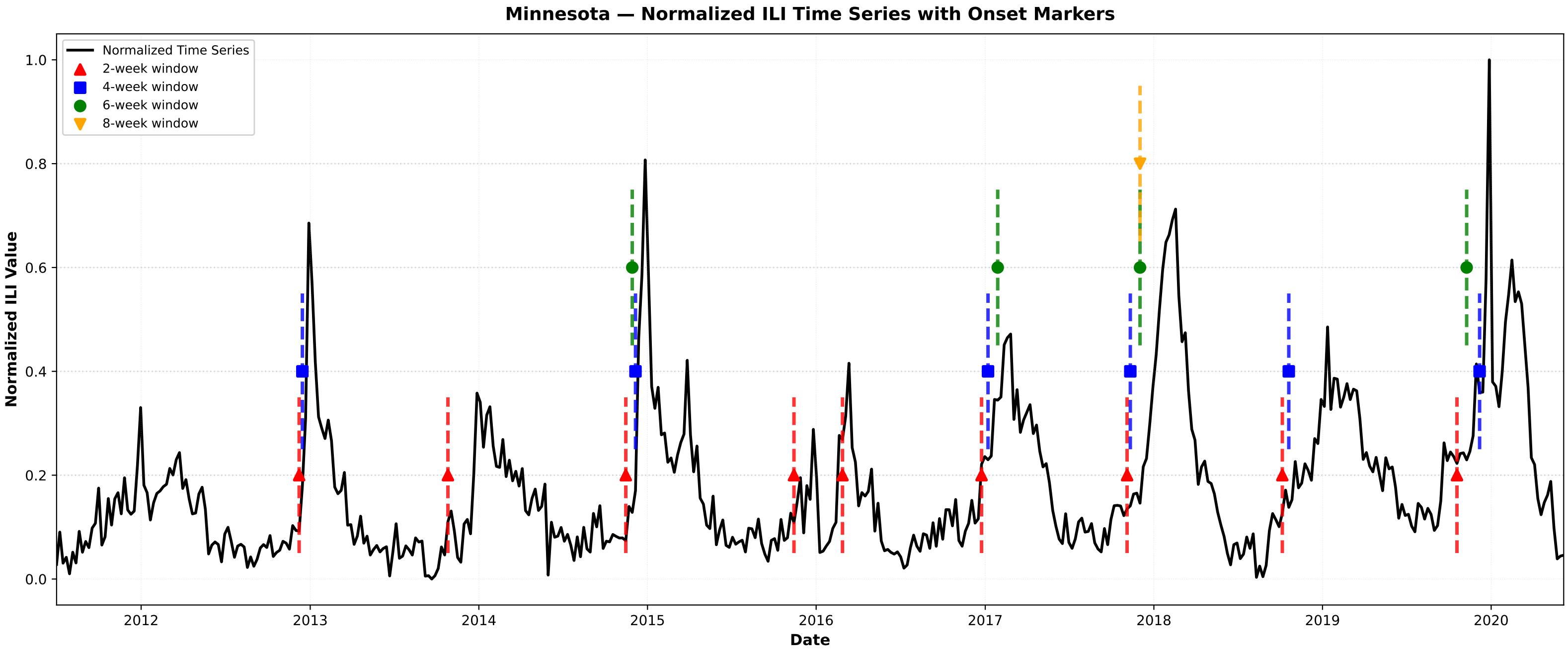
Minnesota — Normalized ILI Time Series with Onset Markers
Normalized Time Series
2-week window
4-week window
6-week window
8-week window
Normalized ILI Value
Date
0.0
0.2
0.4
0.6
0.8
1.0
2012
2013
2014
2015
2016
2017
2018
2019
2020

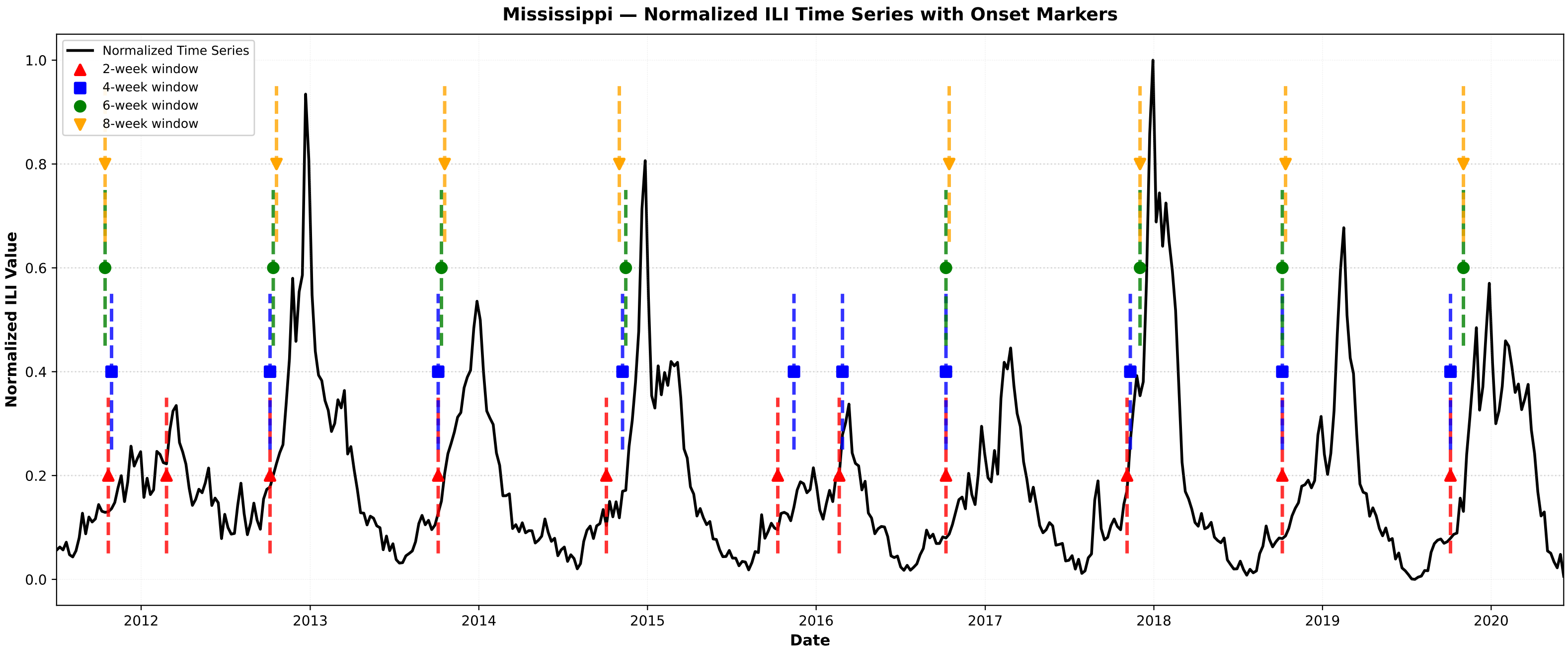

Mississippi — Normalized ILI Time Series with Onset Markers
Normalized Time Series
2-week window
4-week window
6-week window
8-week window
Normalized ILI Value
Date
0.0
0.2
0.4
0.6
0.8
1.0
2012
2013
2014
2015
2016
2017
2018
2019
2020


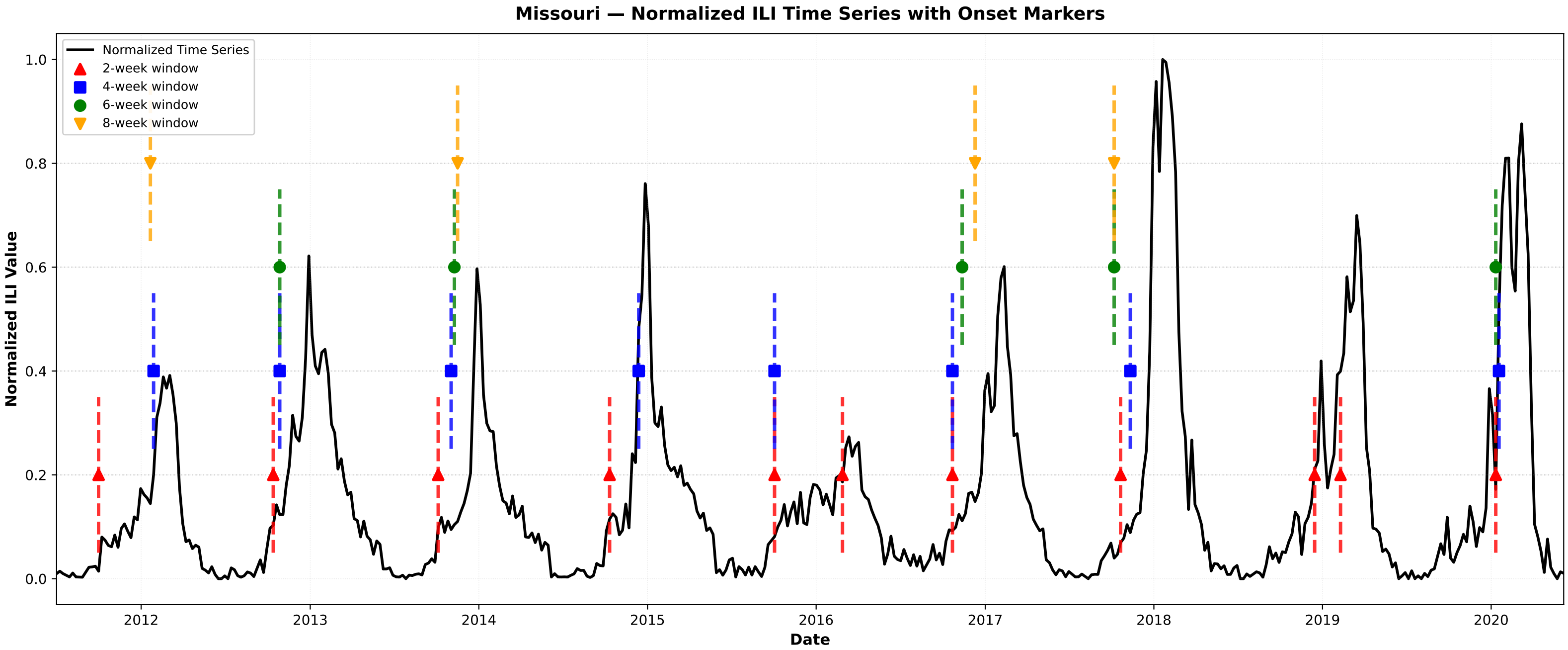

Missouri — Normalized ILI Time Series with Onset Markers
Normalized Time Series
2-week window
4-week window
6-week window
8-week window
Normalized ILI Value
Date
0.0
0.2
0.4
0.6
0.8
1.0
2012
2013
2014
2015
2016
2017
2018
2019
2020


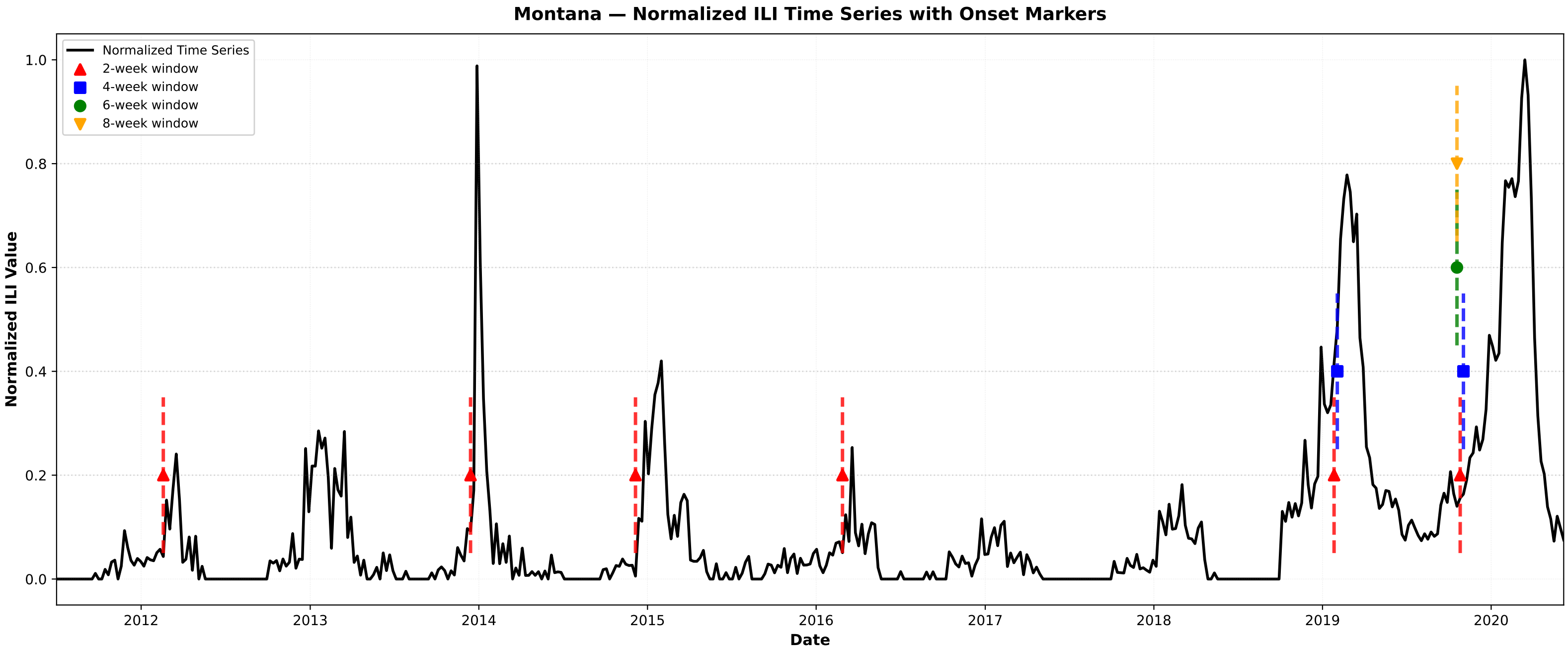

Montana — Normalized ILI Time Series with Onset Markers
Normalized Time Series
2-week window
4-week window
6-week window
8-week window
Normalized ILI Value
Date
0.0
0.2
0.4
0.6
0.8
1.0
2012
2013
2014
2015
2016
2017
2018
2019
2020

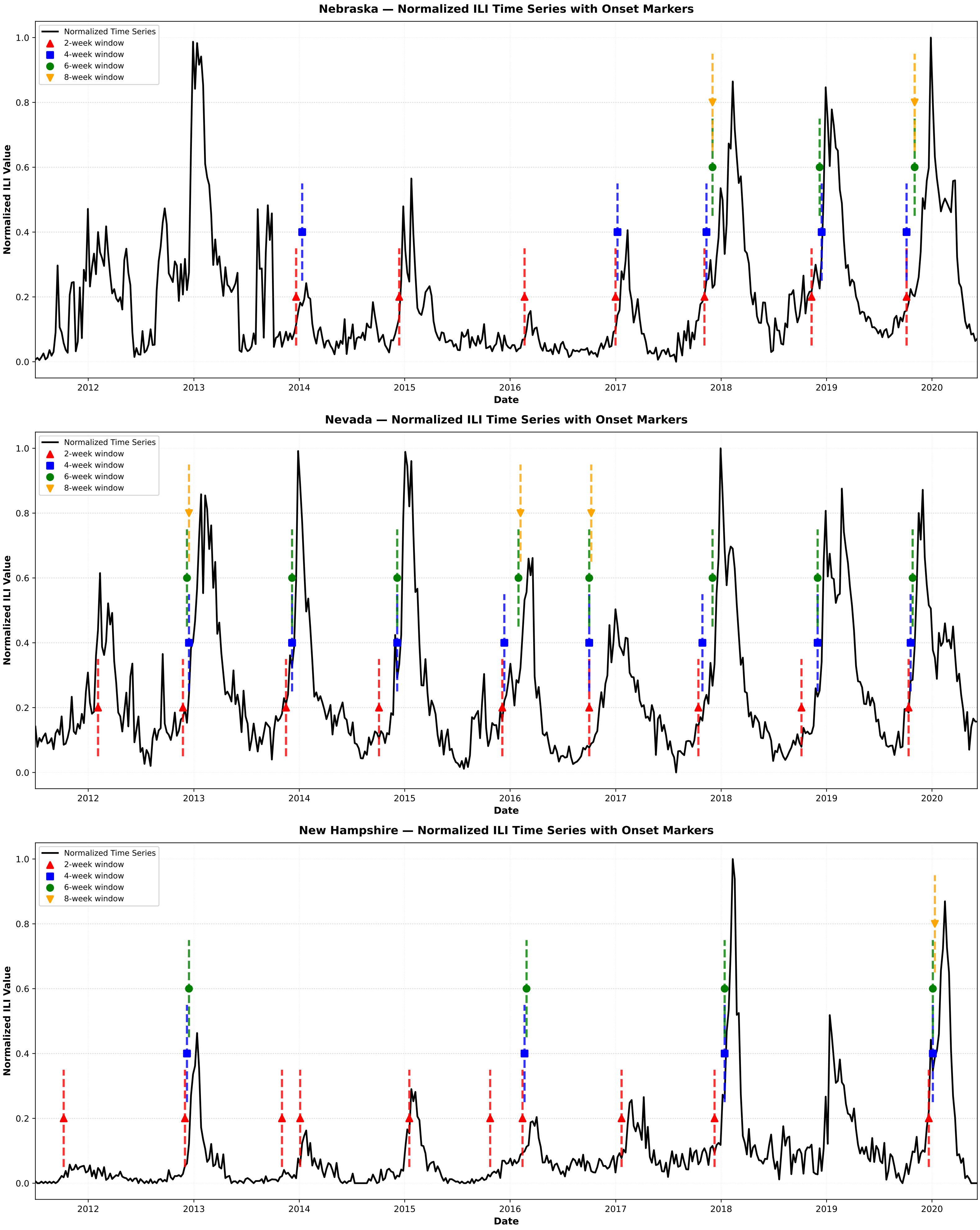

Nebraska — Normalized ILI Time Series with Onset Markers
Normalized Time Series
2-week window
4-week window
6-week window
8-week window
Normalized ILI Value
Date
Nevada — Normalized ILI Time Series with Onset Markers
New Hampshire — Normalized ILI Time Series with Onset Markers

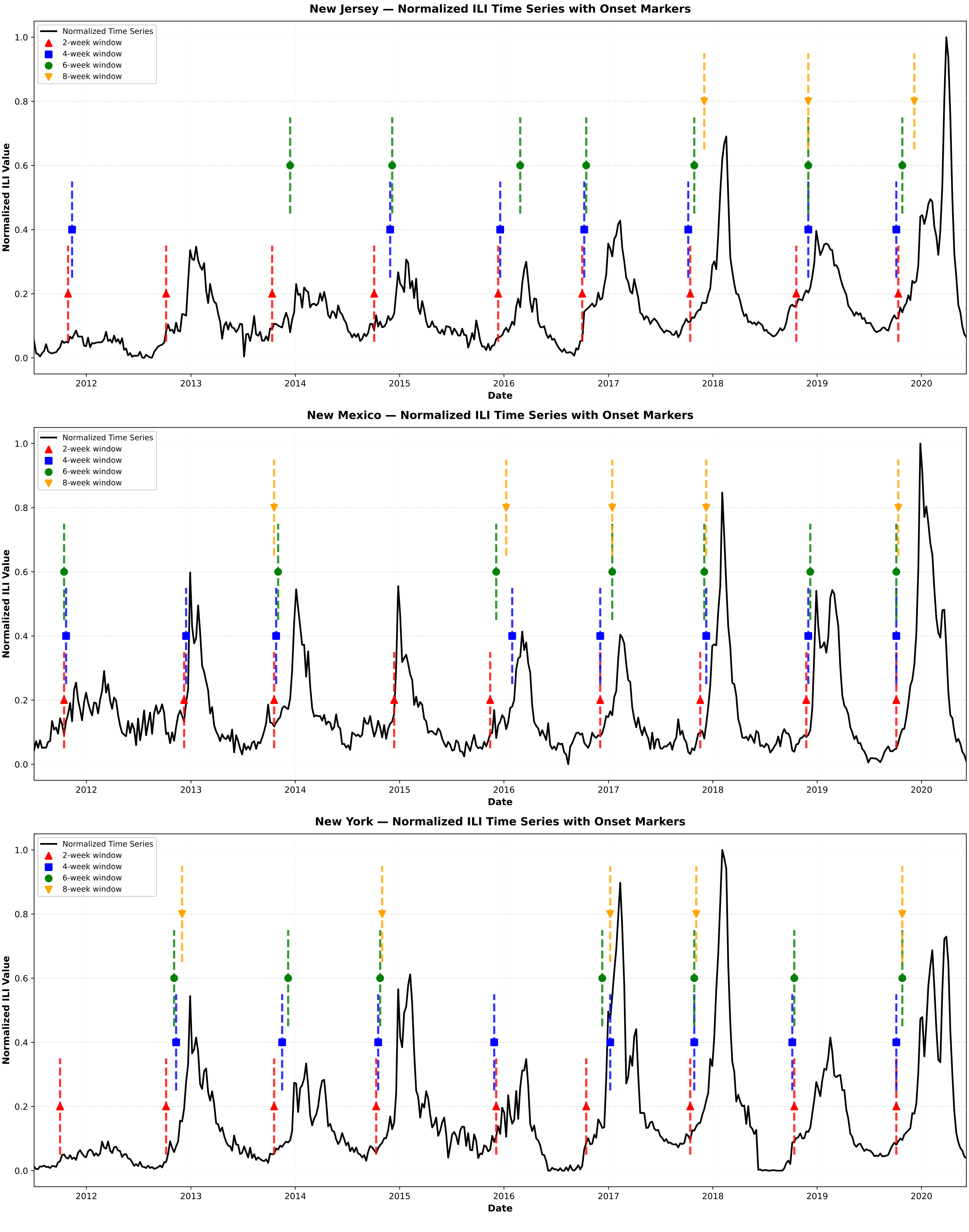

New Jersey — Normalized ILI Time Series with Onset Markers
New Mexico — Normalized ILI Time Series with Onset Markers
New York — Normalized ILI Time Series with Onset Markers
Normalized Time Series
2-week window
4-week window
6-week window
8-week window
Normalized ILI Value
Date
0.0
0.2
0.4
0.6
0.8
1.0
2012
2013
2014
2015
2016
2017
2018
2019
2020

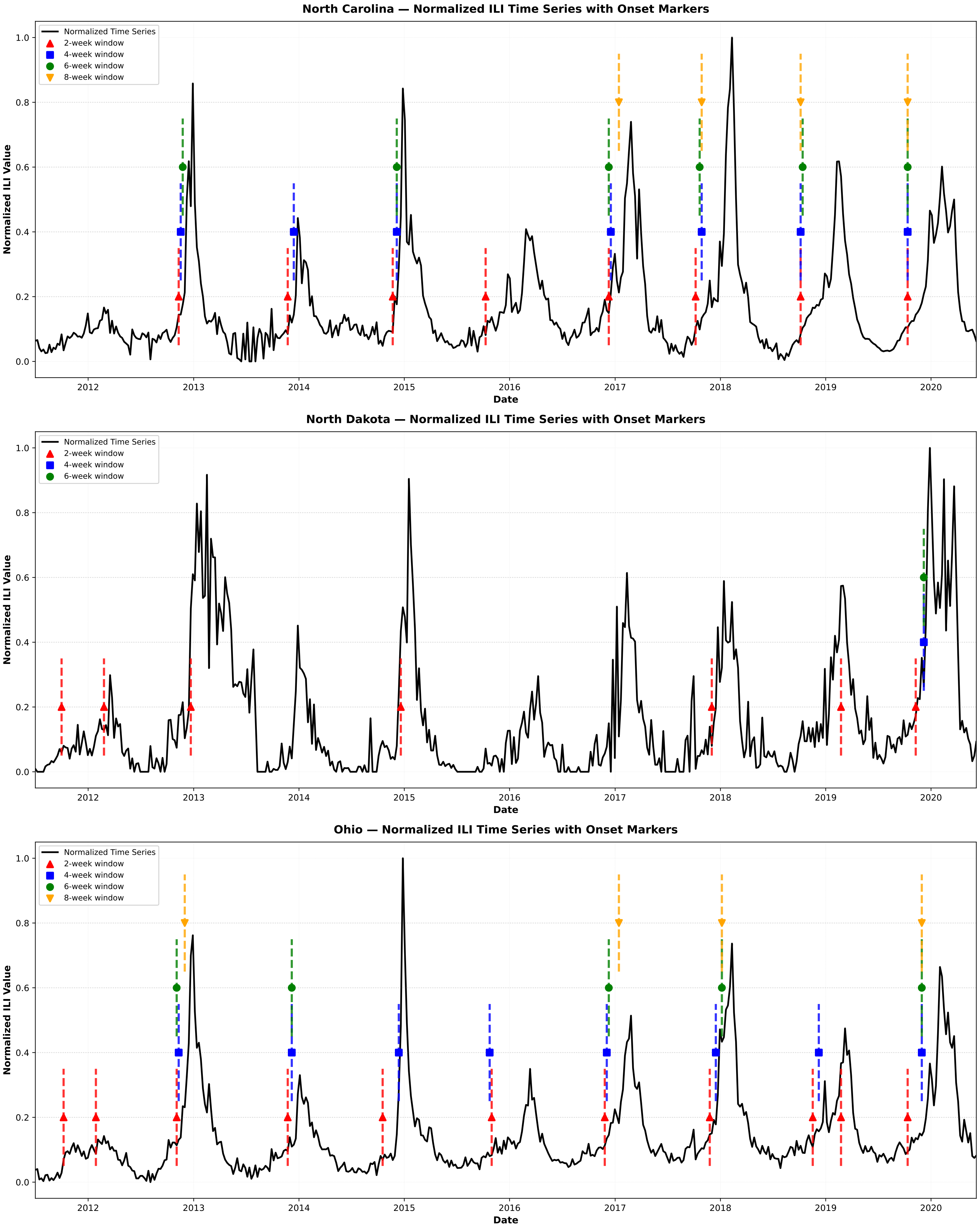
North Carolina — Normalized ILI Time Series with Onset Markers
Normalized Time Series
2-week window
4-week window
6-week window
8-week window
Normalized ILI Value
Date
North Dakota — Normalized ILI Time Series with Onset Markers
Normalized Time Series
2-week window
4-week window
6-week window
Normalized ILI Value
Date
Ohio — Normalized ILI Time Series with Onset Markers
Normalized Time Series
2-week window
4-week window
6-week window
8-week window
Normalized ILI Value
Date

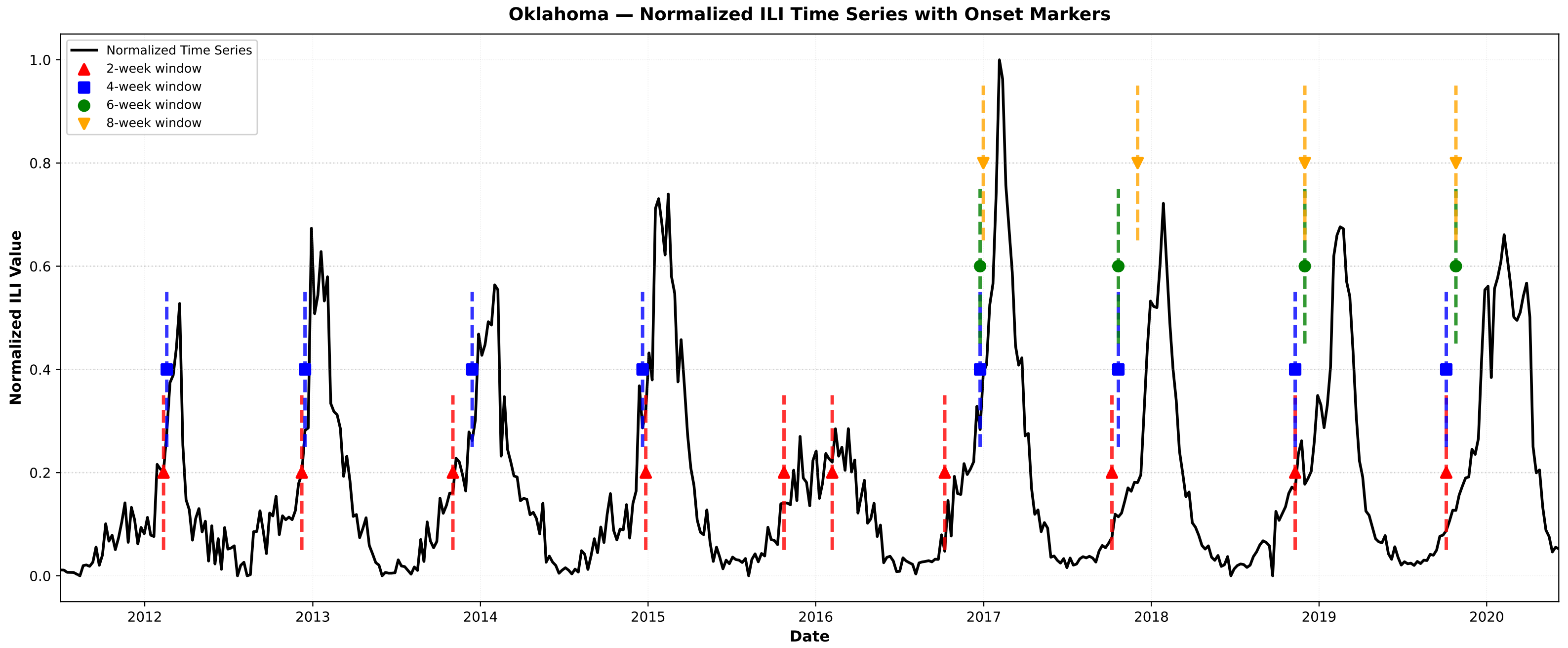
Oklahoma — Normalized ILI Time Series with Onset Markers
Normalized Time Series
2-week window
4-week window
6-week window
8-week window
Normalized ILI Value
Date
0.0
0.2
0.4
0.6
0.8
1.0
2012
2013
2014
2015
2016
2017
2018
2019
2020

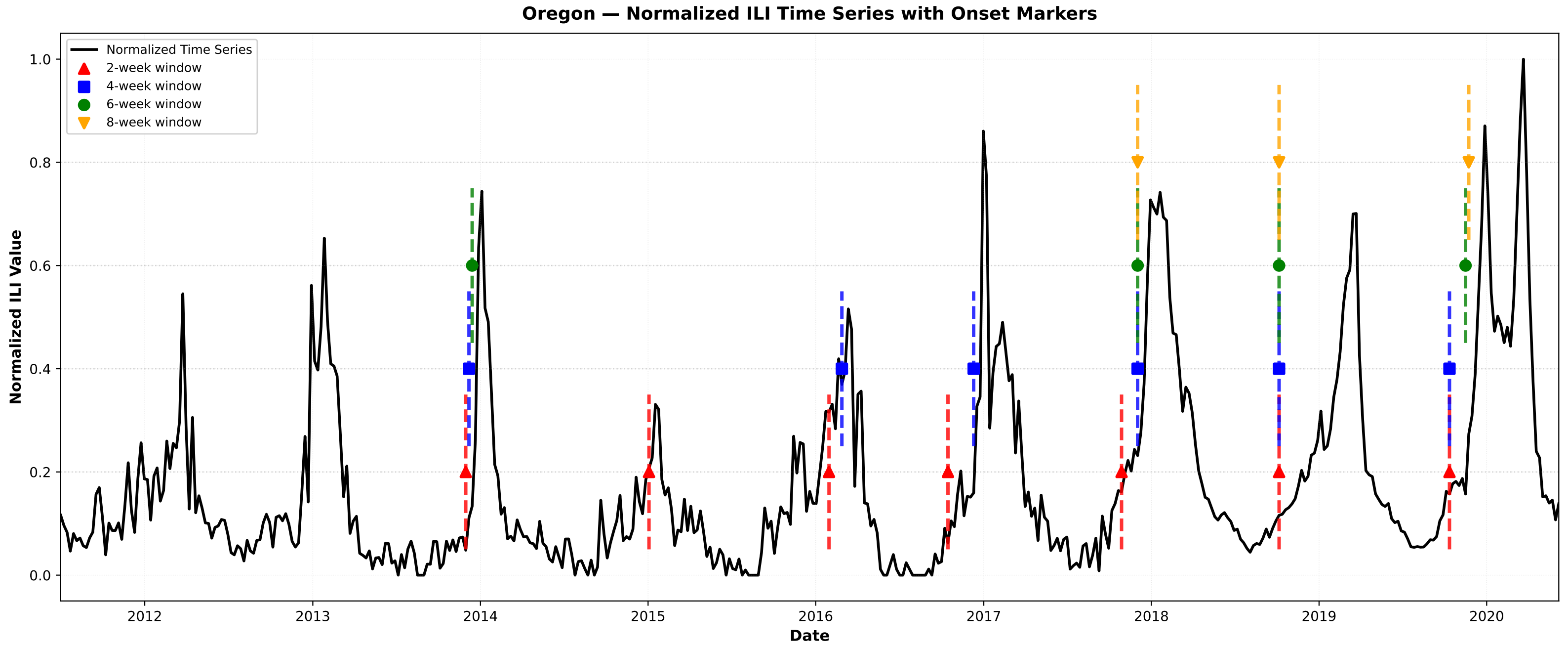
Oregon — Normalized ILI Time Series with Onset Markers
Normalized Time Series
2-week window
4-week window
6-week window
8-week window
Normalized ILI Value
Date
0.0
0.2
0.4
0.6
0.8
1.0
2012
2013
2014
2015
2016
2017
2018
2019
2020

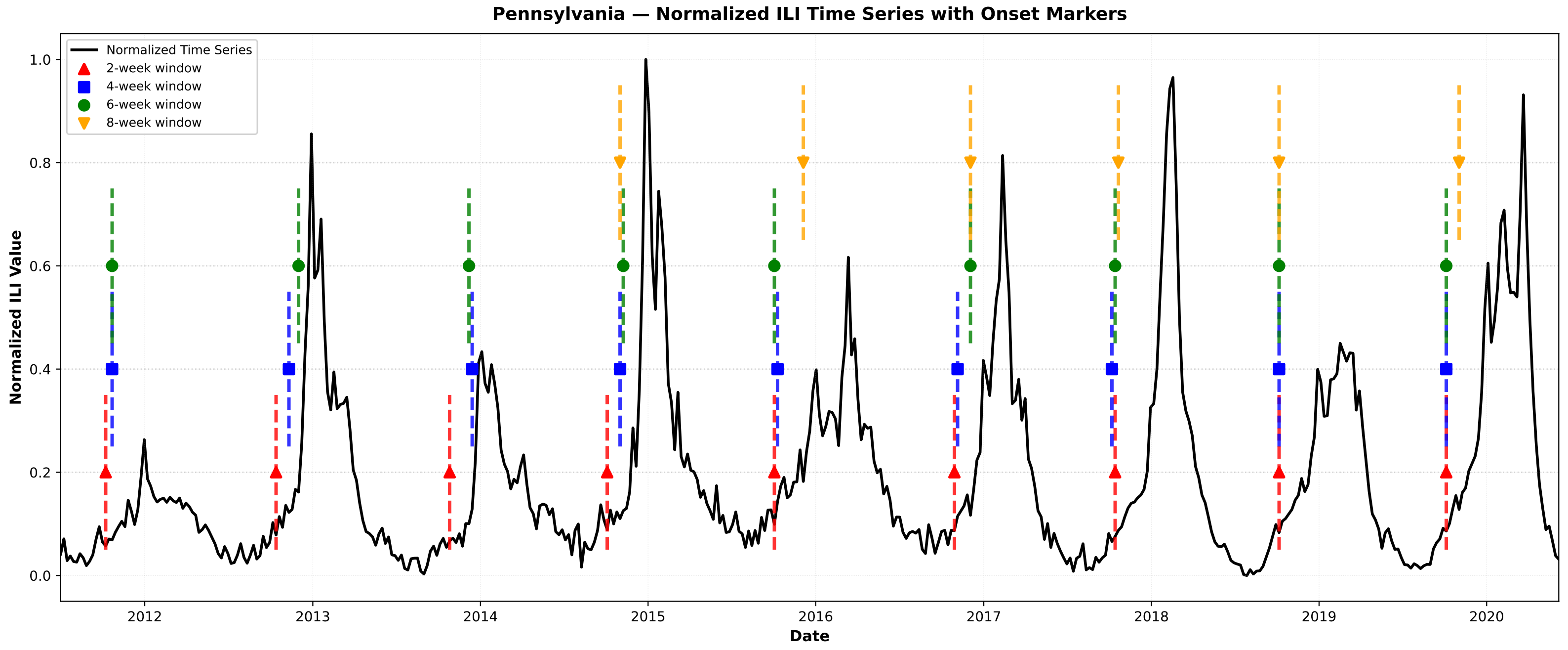
Pennsylvania — Normalized ILI Time Series with Onset Markers
Normalized Time Series
2-week window
4-week window
6-week window
8-week window
Normalized ILI Value
Date
0.0
0.2
0.4
0.6
0.8
1.0
2012
2013
2014
2015
2016
2017
2018
2019
2020

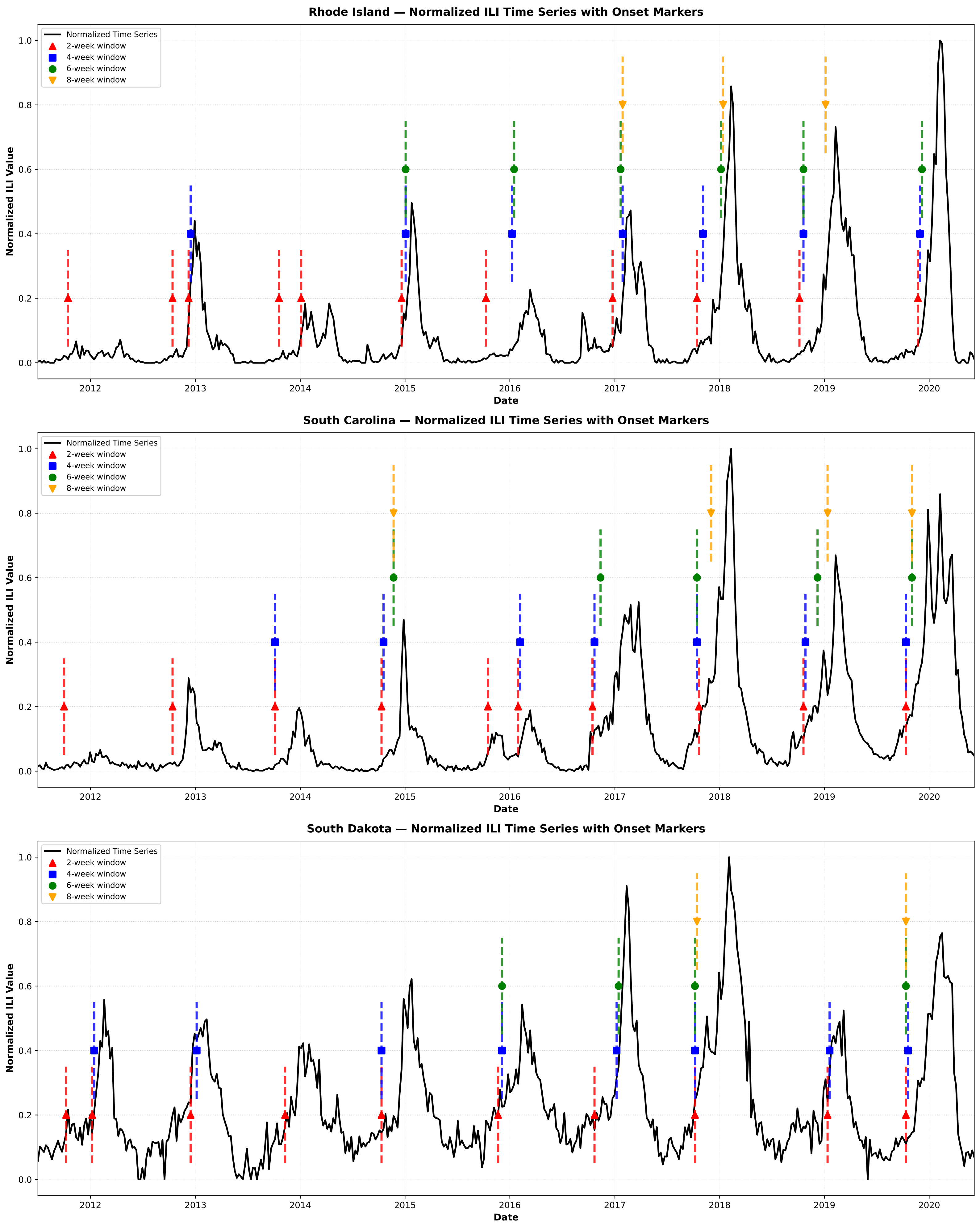

Rhode Island — Normalized ILI Time Series with Onset Markers
South Carolina — Normalized ILI Time Series with Onset Markers
South Dakota — Normalized ILI Time Series with Onset Markers
Normalized Time Series
2-week window
4-week window
6-week window
8-week window
Normalized ILI Value
Date
0.0
0.2
0.4
0.6
0.8
1.0
2012
2013
2014
2015
2016
2017
2018
2019
2020

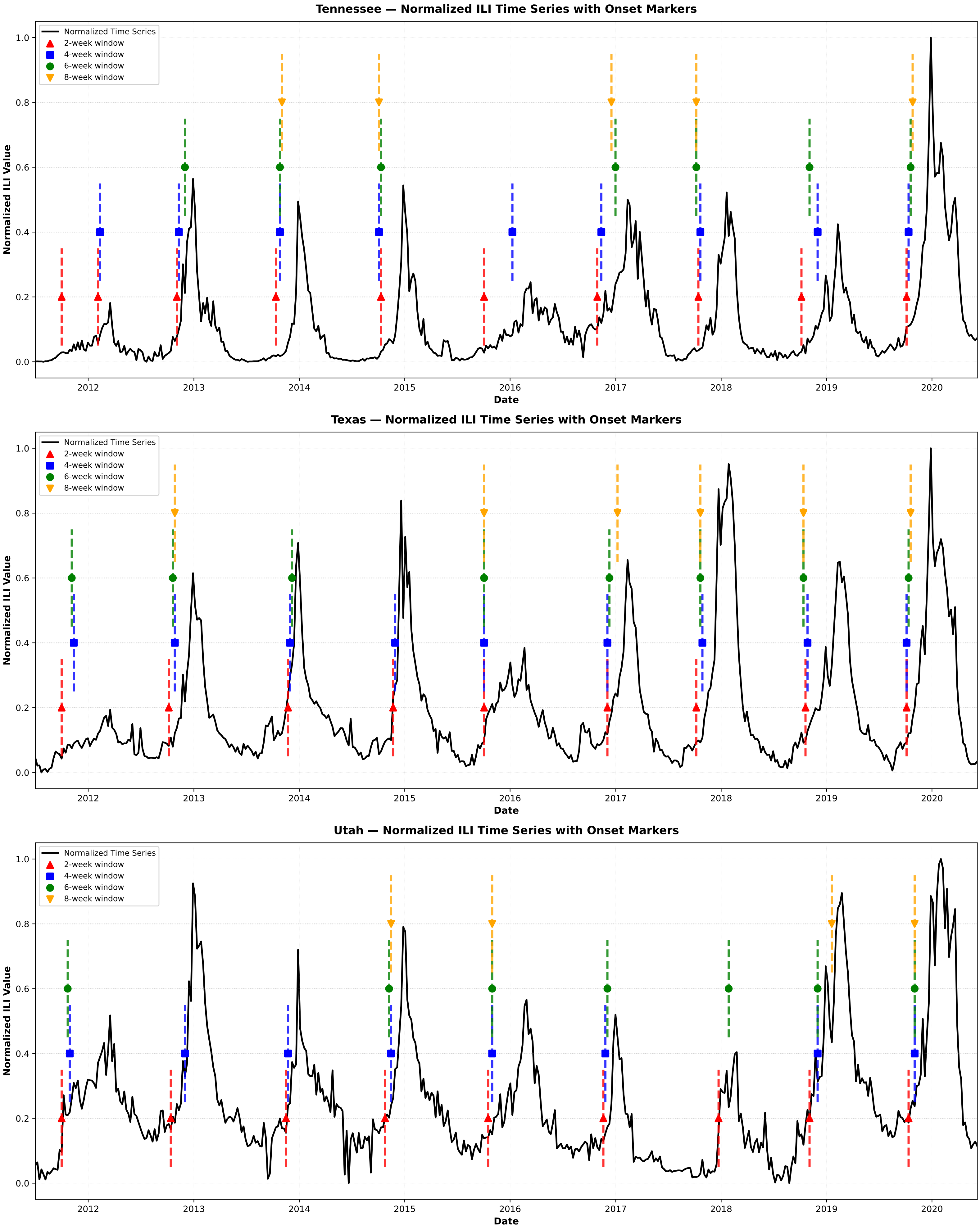

Tennessee — Normalized ILI Time Series with Onset Markers
Texas — Normalized ILI Time Series with Onset Markers
Utah — Normalized ILI Time Series with Onset Markers
Normalized Time Series
2-week window
4-week window
6-week window
8-week window
Normalized ILI Value
Date
0.0
0.2
0.4
0.6
0.8
1.0
2012
2013
2014
2015
2016
2017
2018
2019
2020

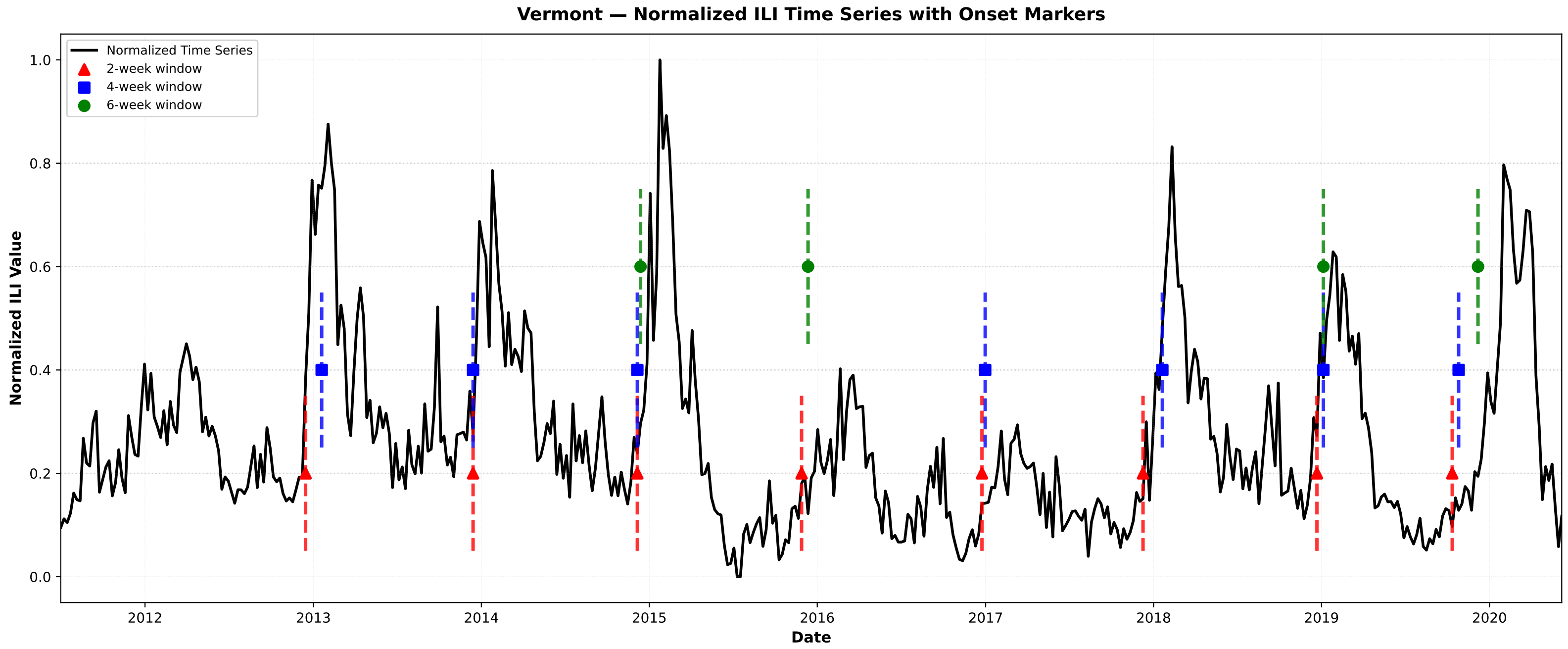

Vermont — Normalized ILI Time Series with Onset Markers
Normalized Time Series
2-week window
4-week window
6-week window
Normalized ILI Value
Date


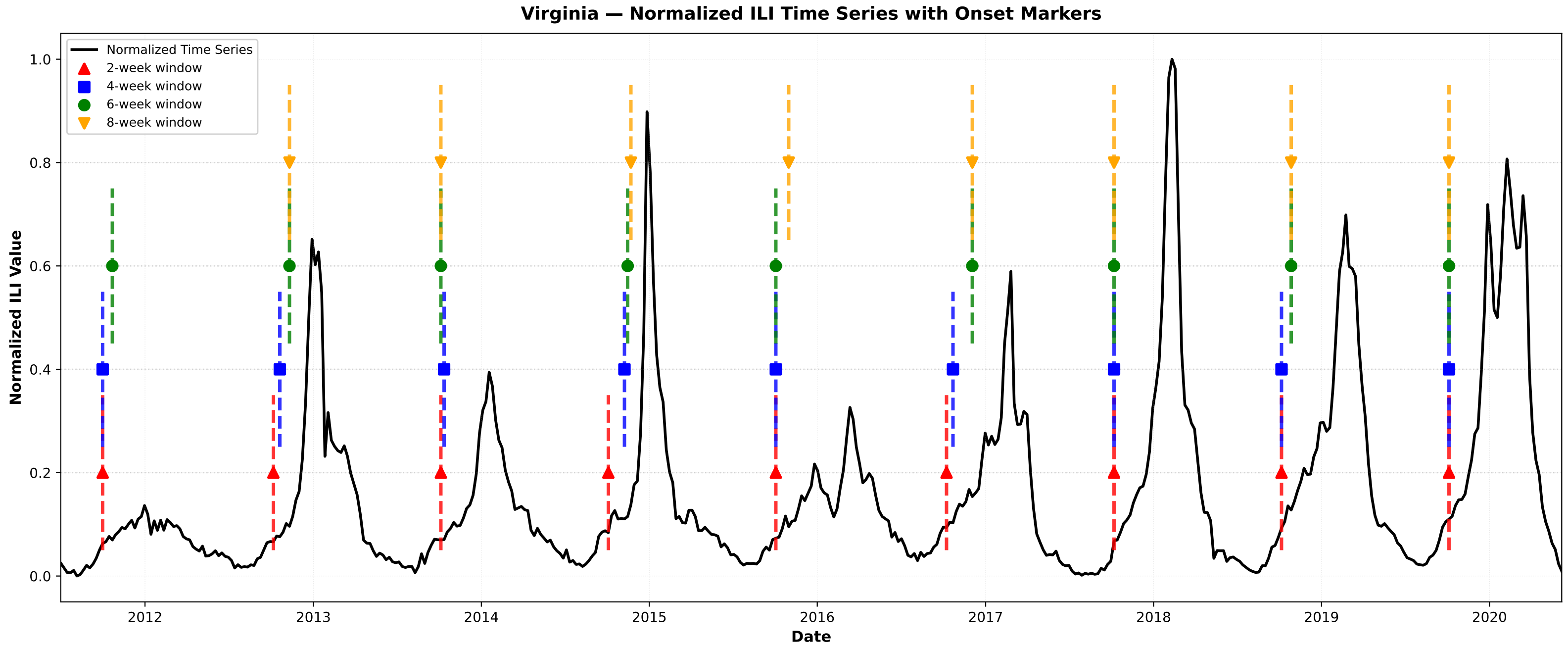

Virginia — Normalized ILI Time Series with Onset Markers
Normalized Time Series
2-week window
4-week window
6-week window
8-week window
Normalized ILI Value
Date


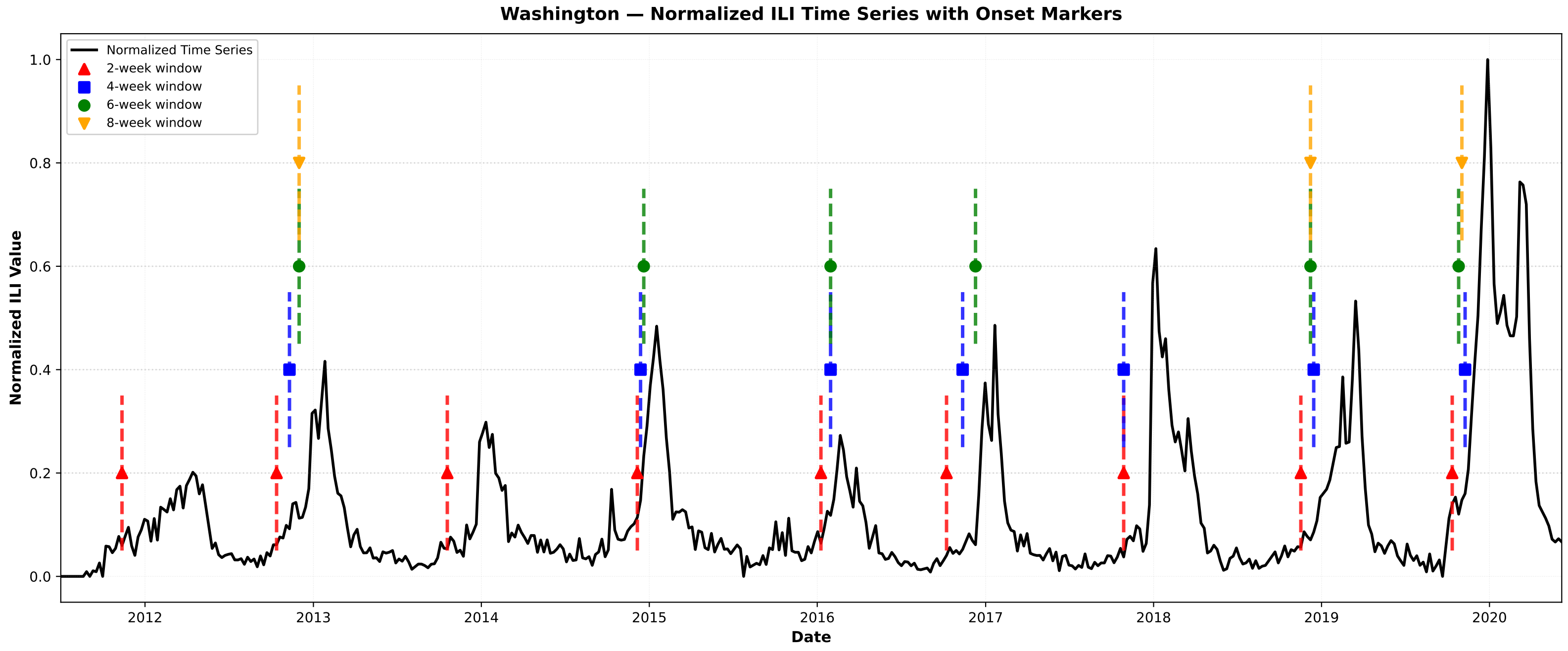

Washington — Normalized ILI Time Series with Onset Markers
Normalized Time Series
2-week window
4-week window
6-week window
8-week window
Normalized ILI Value
Date

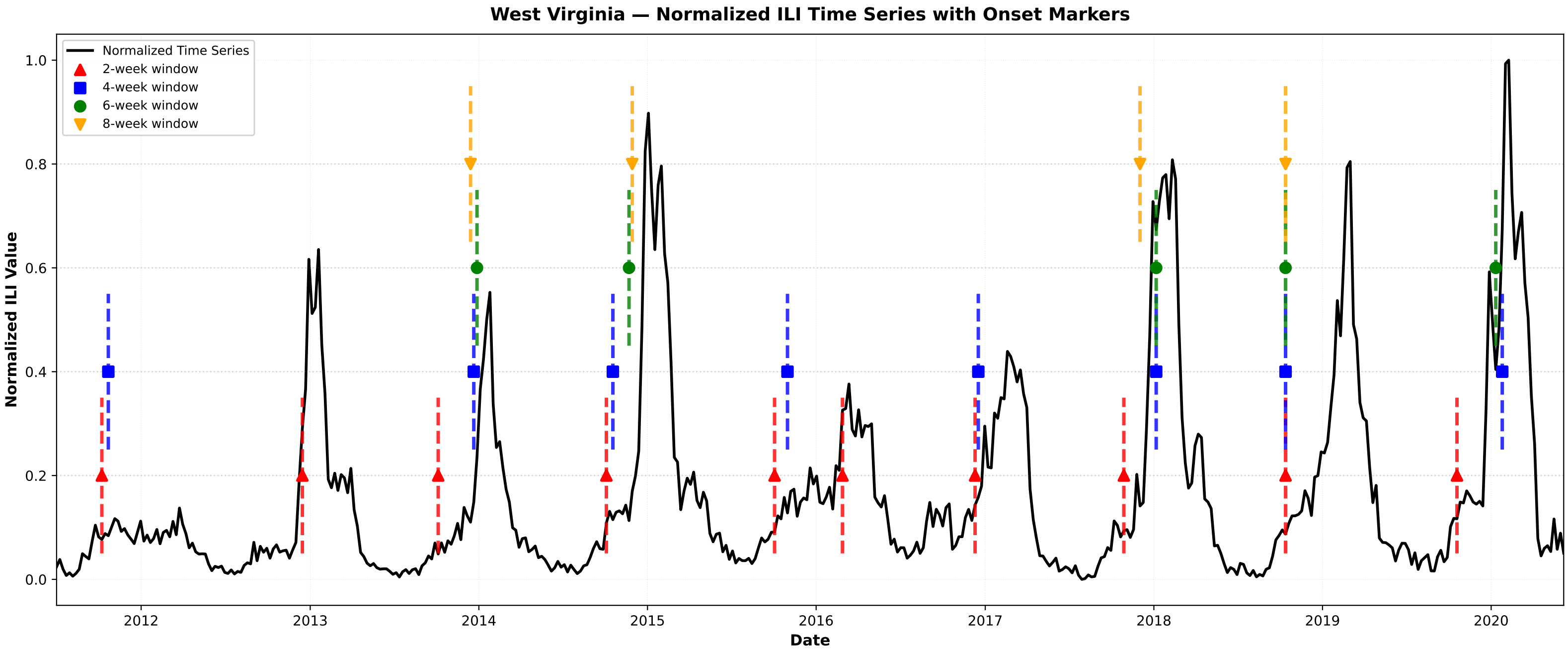
West Virginia — Normalized ILI Time Series with Onset Markers
Normalized Time Series
2-week window
4-week window
6-week window
8-week window
Normalized ILI Value
Date
0.0
0.2
0.4
0.6
0.8
1.0
2012
2013
2014
2015
2016
2017
2018
2019
2020

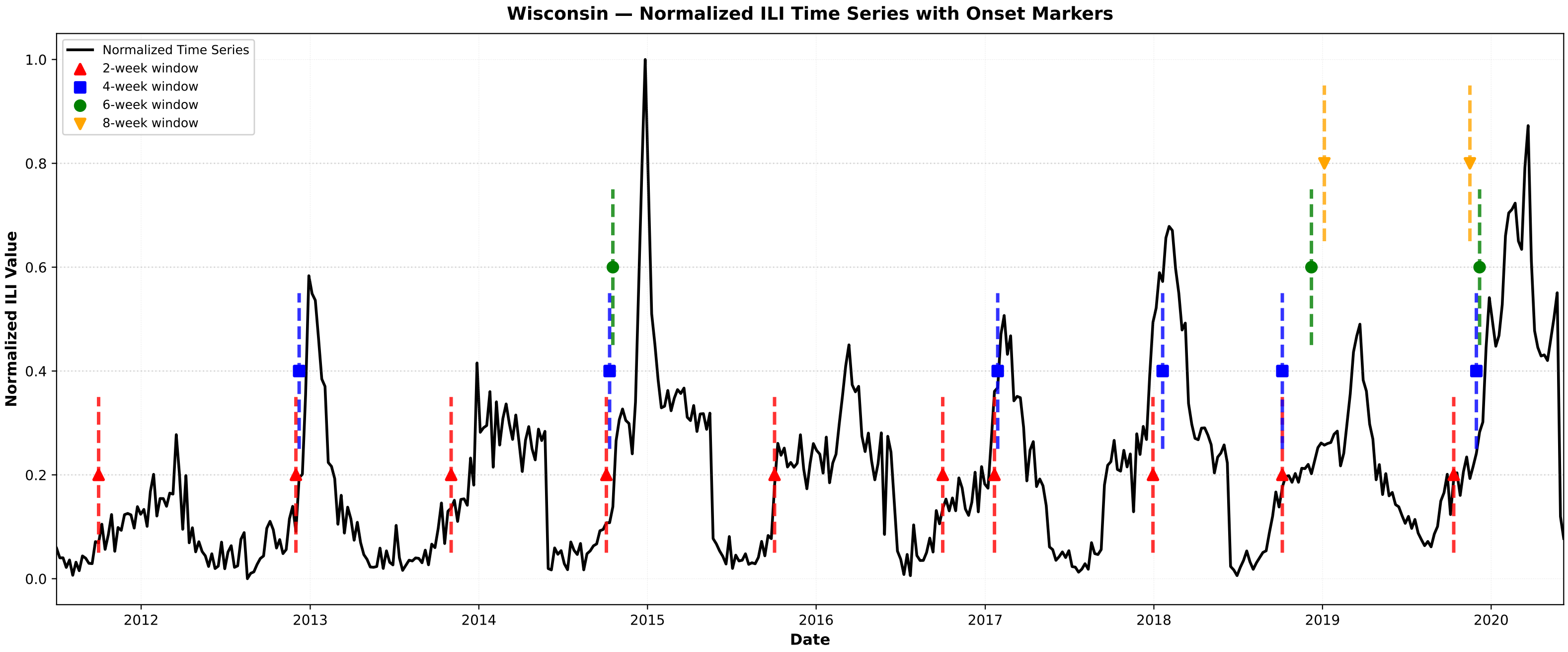
Wisconsin — Normalized ILI Time Series with Onset Markers
Normalized Time Series
2-week window
4-week window
6-week window
8-week window
Normalized ILI Value
Date
0.0
0.2
0.4
0.6
0.8
1.0
2012
2013
2014
2015
2016
2017
2018
2019
2020

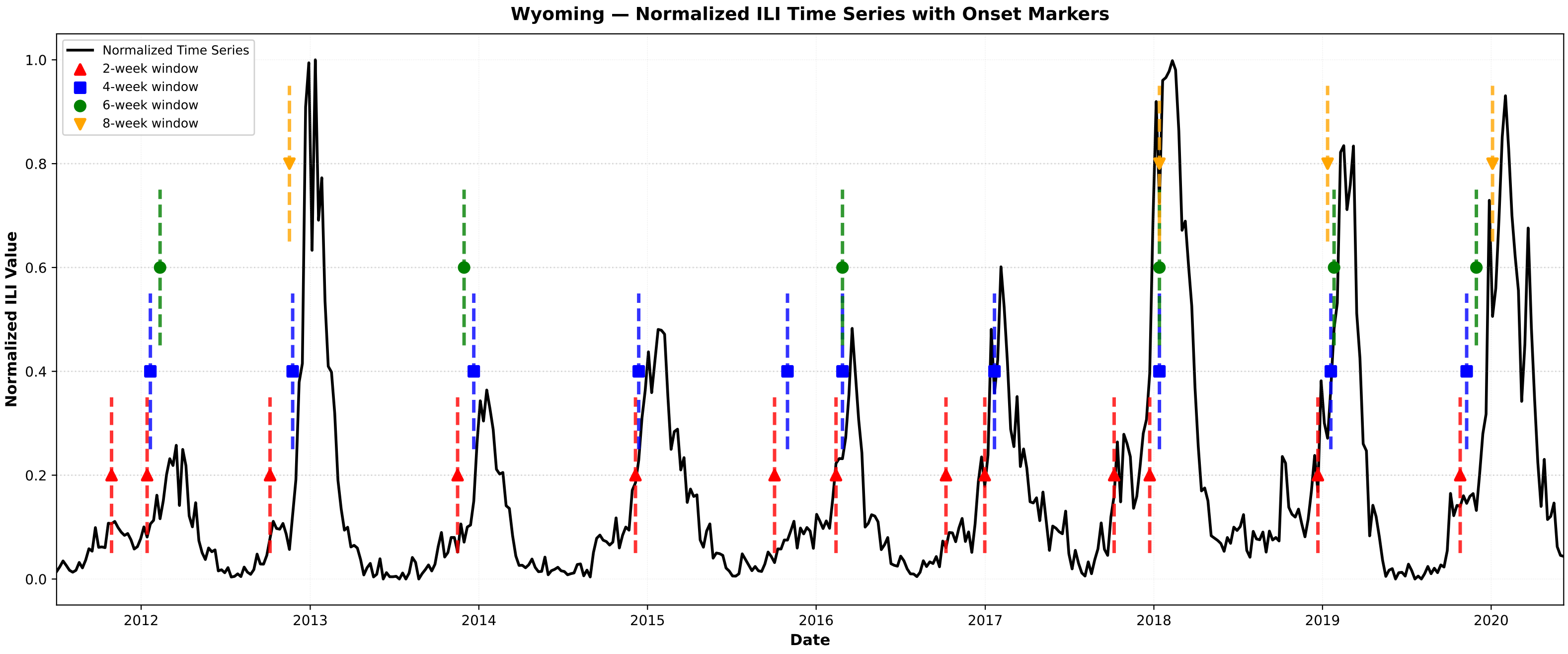
Wyoming — Normalized ILI Time Series with Onset Markers
Normalized Time Series
2-week window
4-week window
6-week window
8-week window
Normalized ILI Value
Date
0.0
0.2
0.4
0.6
0.8
1.0
2012
2013
2014
2015
2016
2017
2018
2019
2020

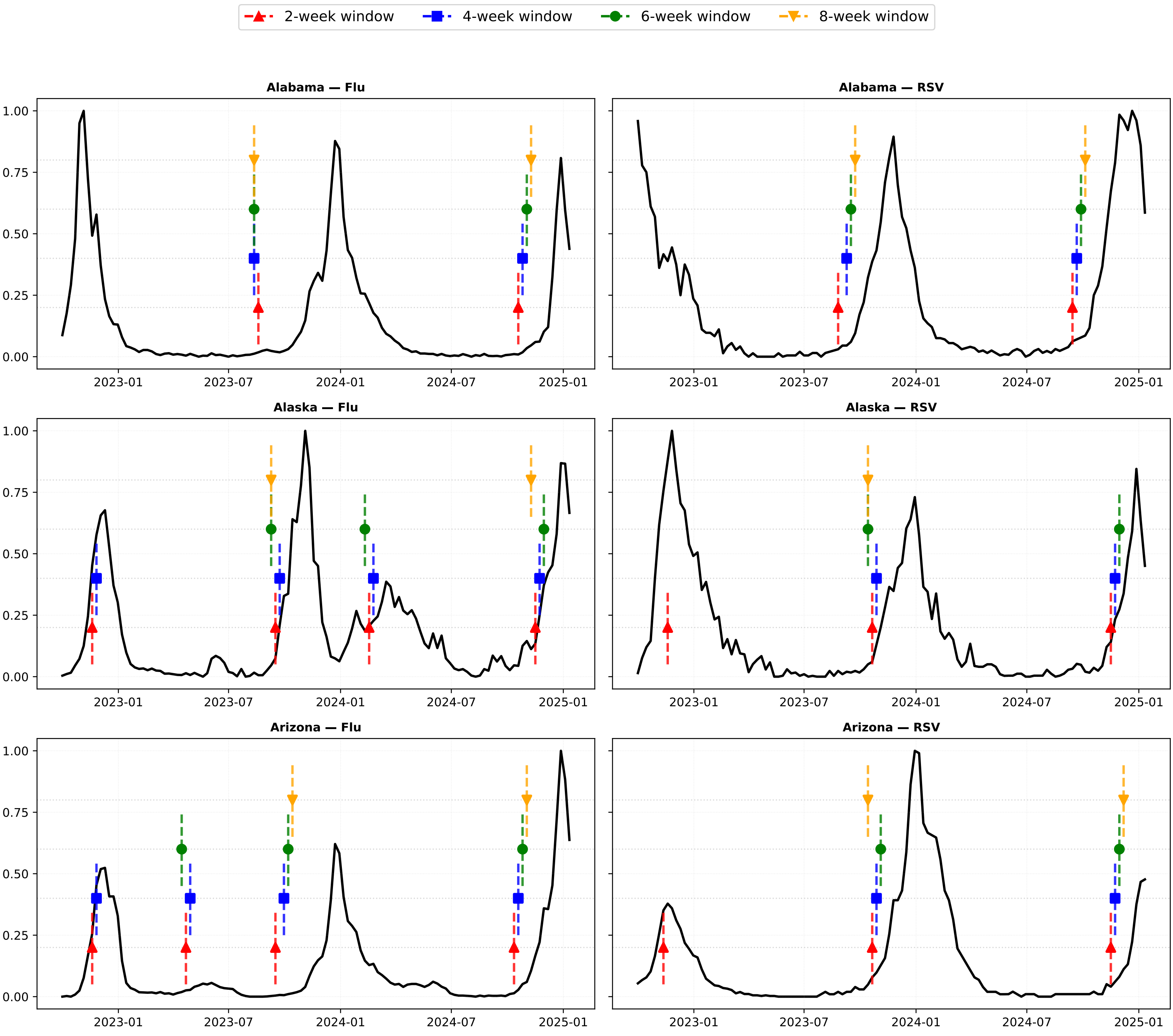

2-week window
4-week window
6-week window
8-week window
Alabama — Flu
Alabama — RSV
Alaska — Flu
Alaska — RSV
Arizona — Flu
Arizona — RSV
0.00
0.25
0.50
0.75
1.00
2023-01
2023-07
2024-01
2024-07
2025-01

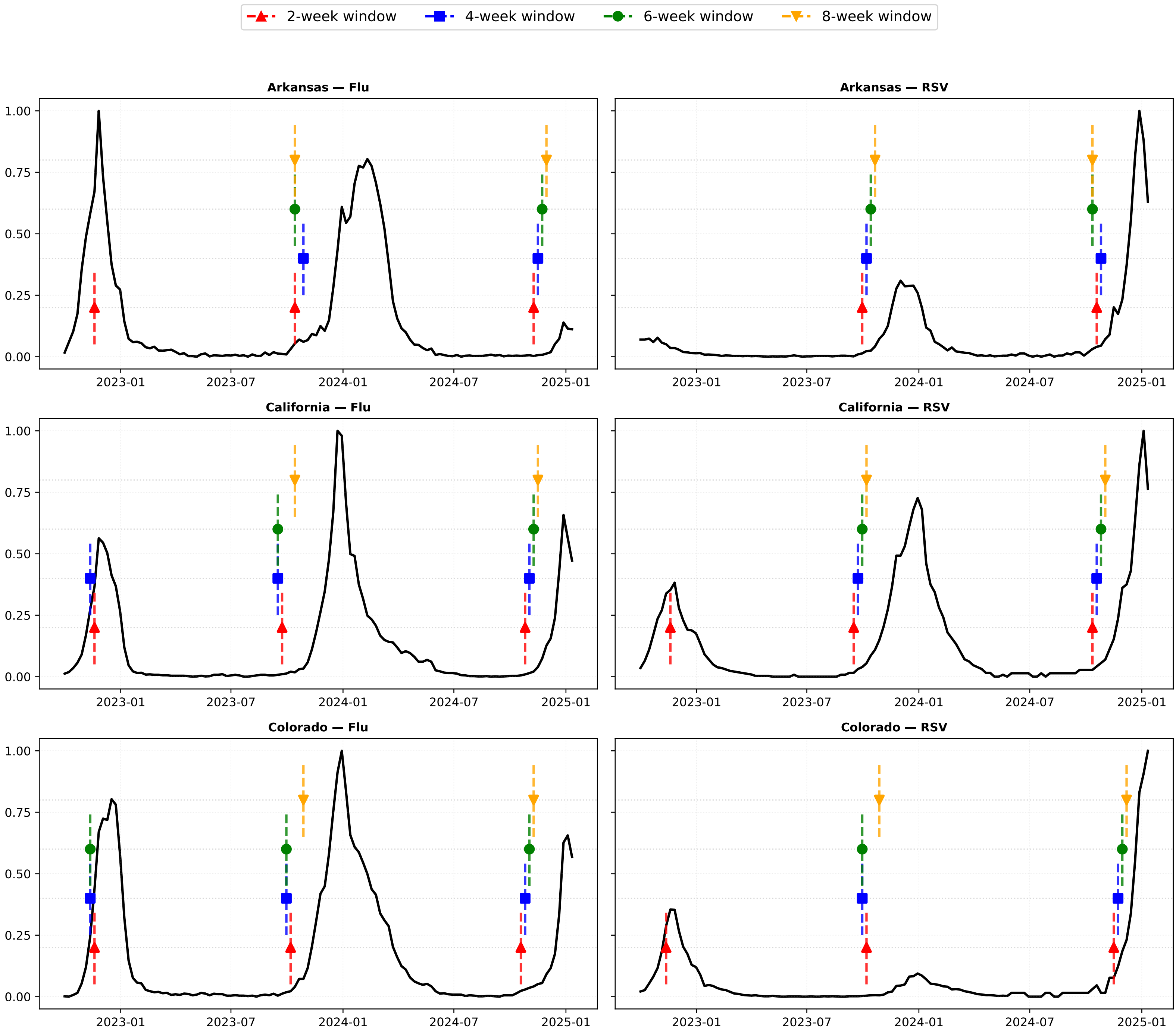

2-week window
4-week window
6-week window
8-week window
Arkansas — Flu
Arkansas — RSV
California — Flu
California — RSV
Colorado — Flu
Colorado — RSV
0.00
0.25
0.50
0.75
1.00
2023-01
2023-07
2024-01
2024-07
2025-01

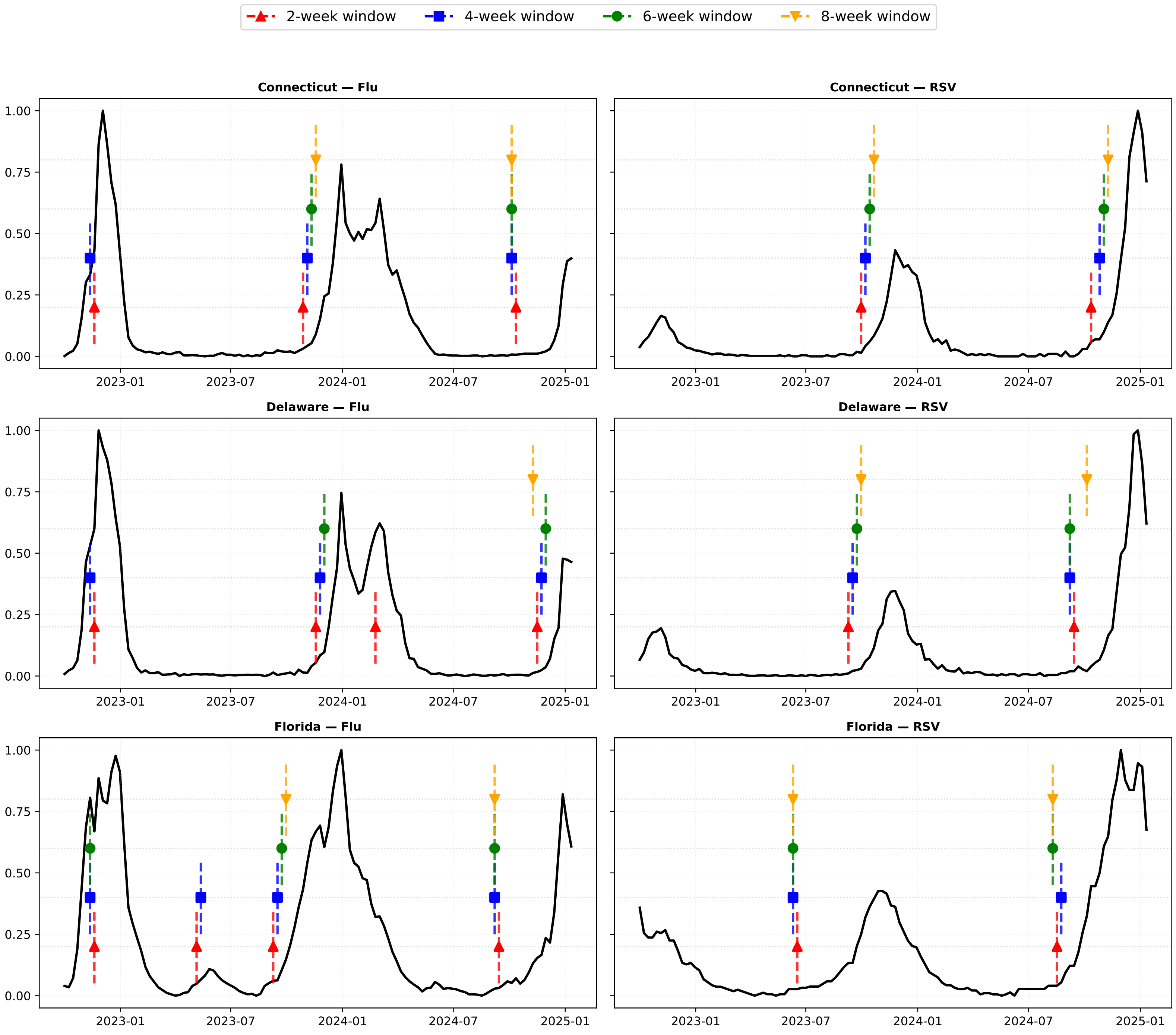

2-week window
4-week window
6-week window
8-week window
Connecticut — Flu
Connecticut — RSV
Delaware — Flu
Delaware — RSV
Florida — Flu
Florida — RSV
0.00
0.25
0.50
0.75
1.00
2023-01
2023-07
2024-01
2024-07
2025-01

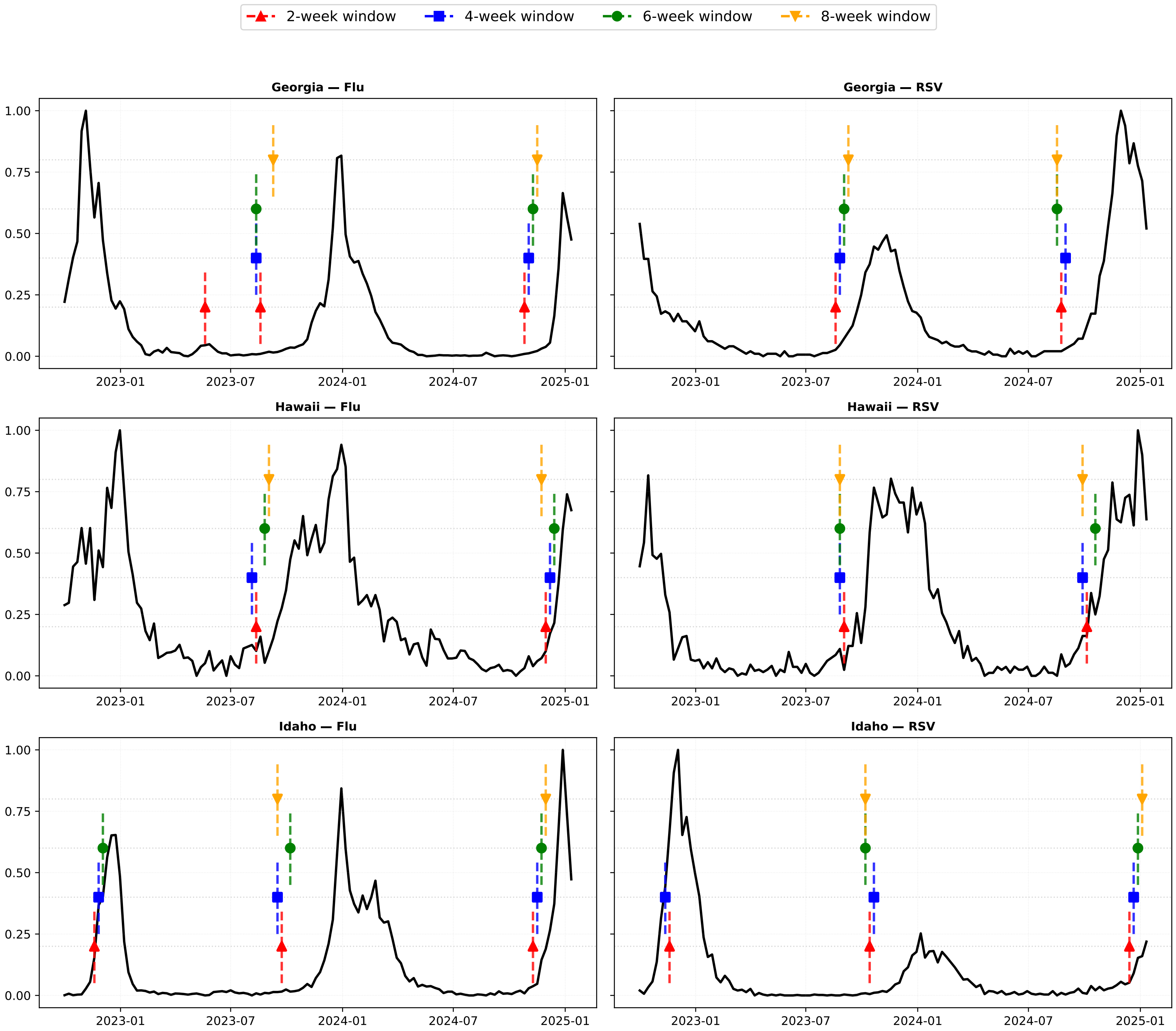

2-week window
4-week window
6-week window
8-week window
Georgia — Flu
Georgia — RSV
Hawaii — Flu
Hawaii — RSV
Idaho — Flu
Idaho — RSV
0.00
0.25
0.50
0.75
1.00
2023-01
2023-07
2024-01
2024-07
2025-01

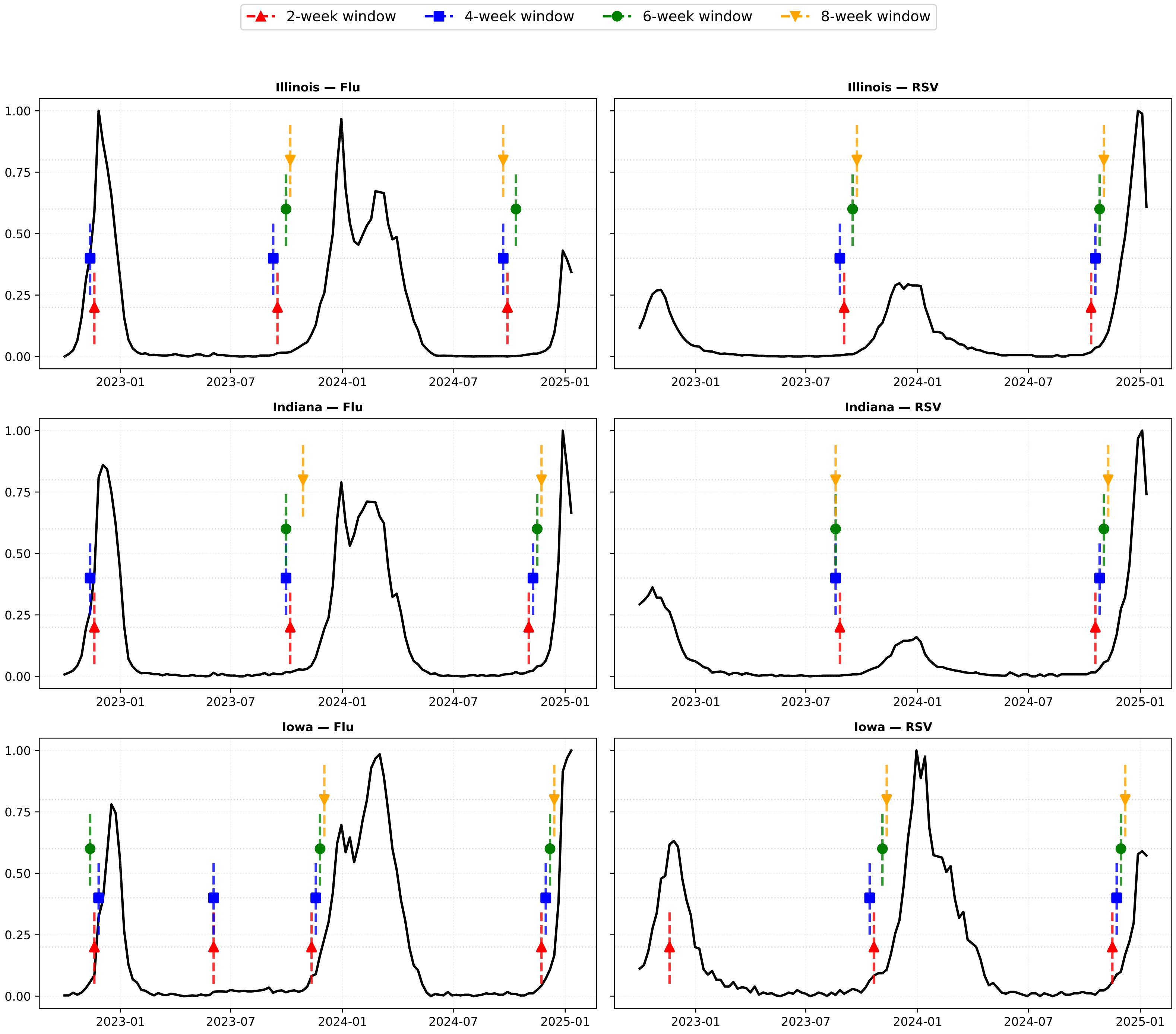
2-week window
4-week window
6-week window
8-week window
Illinois — Flu
Illinois — RSV
Indiana — Flu
Indiana — RSV
Iowa — Flu
Iowa — RSV
1.00
0.75
0.50
0.25
0.00
2023-01
2023-07
2024-01
2024-07
2025-01

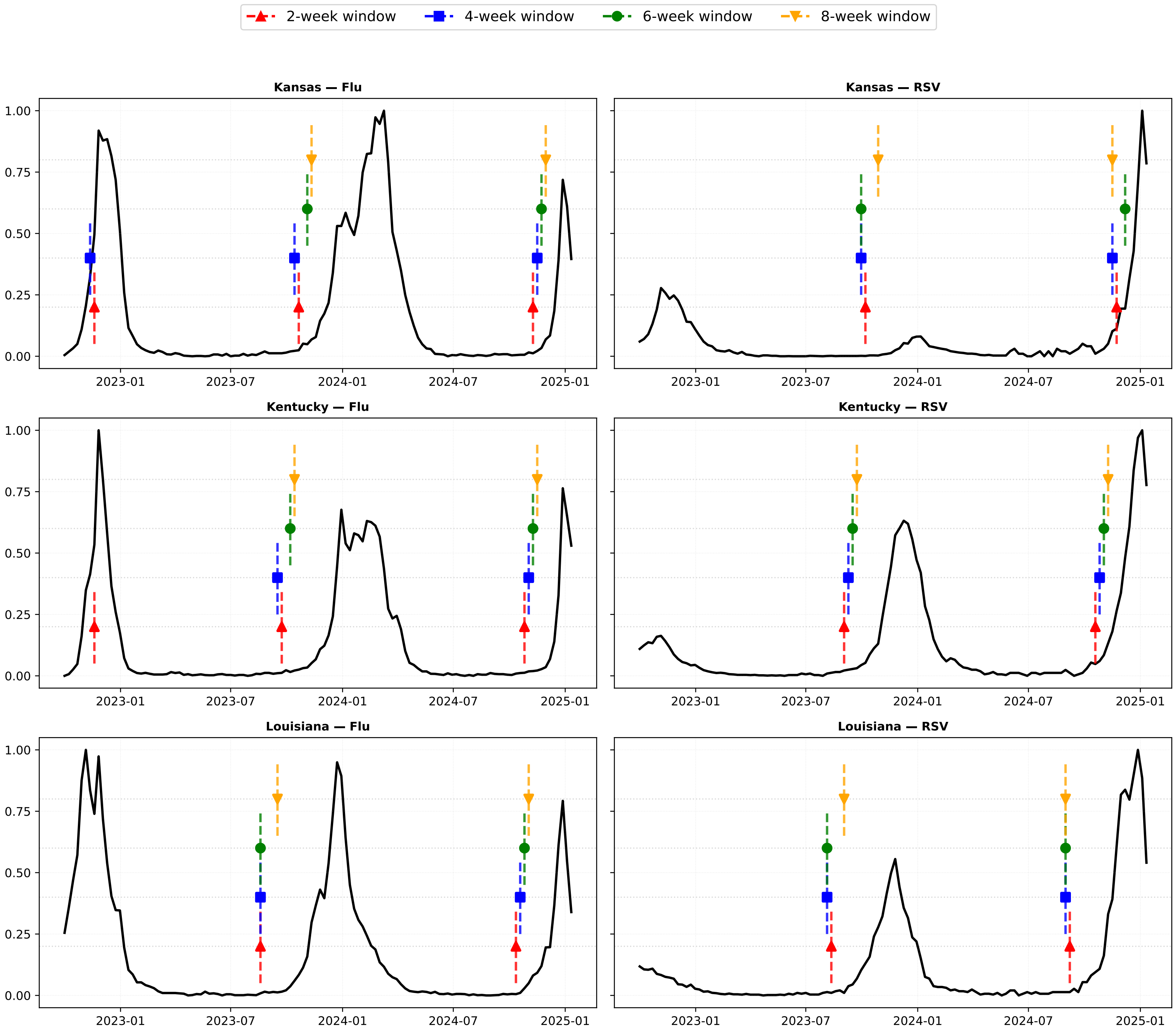

2-week window
4-week window
6-week window
8-week window
Kansas — Flu
Kansas — RSV
Kentucky — Flu
Kentucky — RSV
Louisiana — Flu
Louisiana — RSV
1.00
0.75
0.50
0.25
0.00
2023-01
2023-07
2024-01
2024-07
2025-01

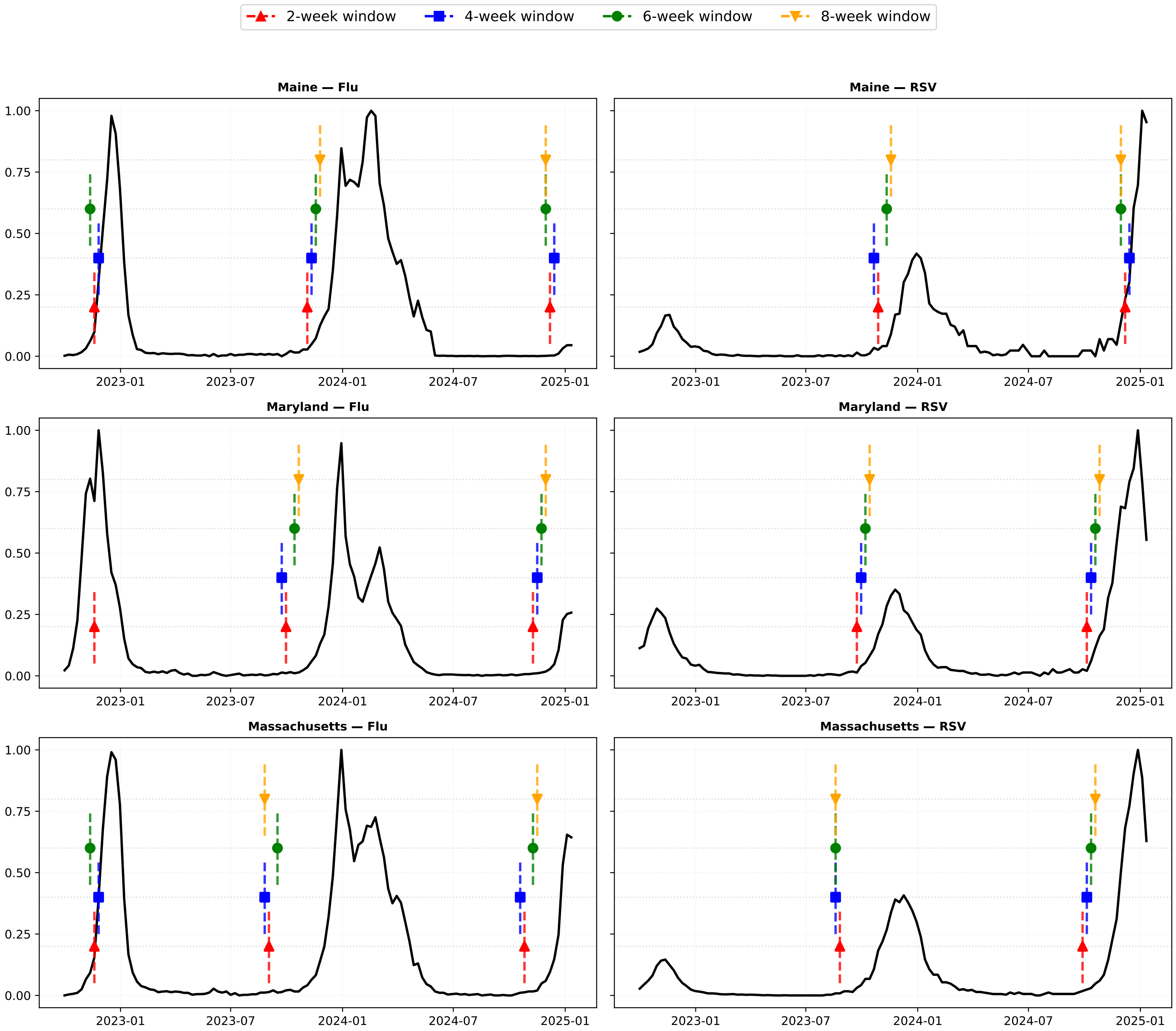

2-week window
4-week window
6-week window
8-week window
Maine — Flu
Maine — RSV
Maryland — Flu
Maryland — RSV
Massachusetts — Flu
Massachusetts — RSV
1.00
0.75
0.50
0.25
0.00
2023-01
2023-07
2024-01
2024-07
2025-01

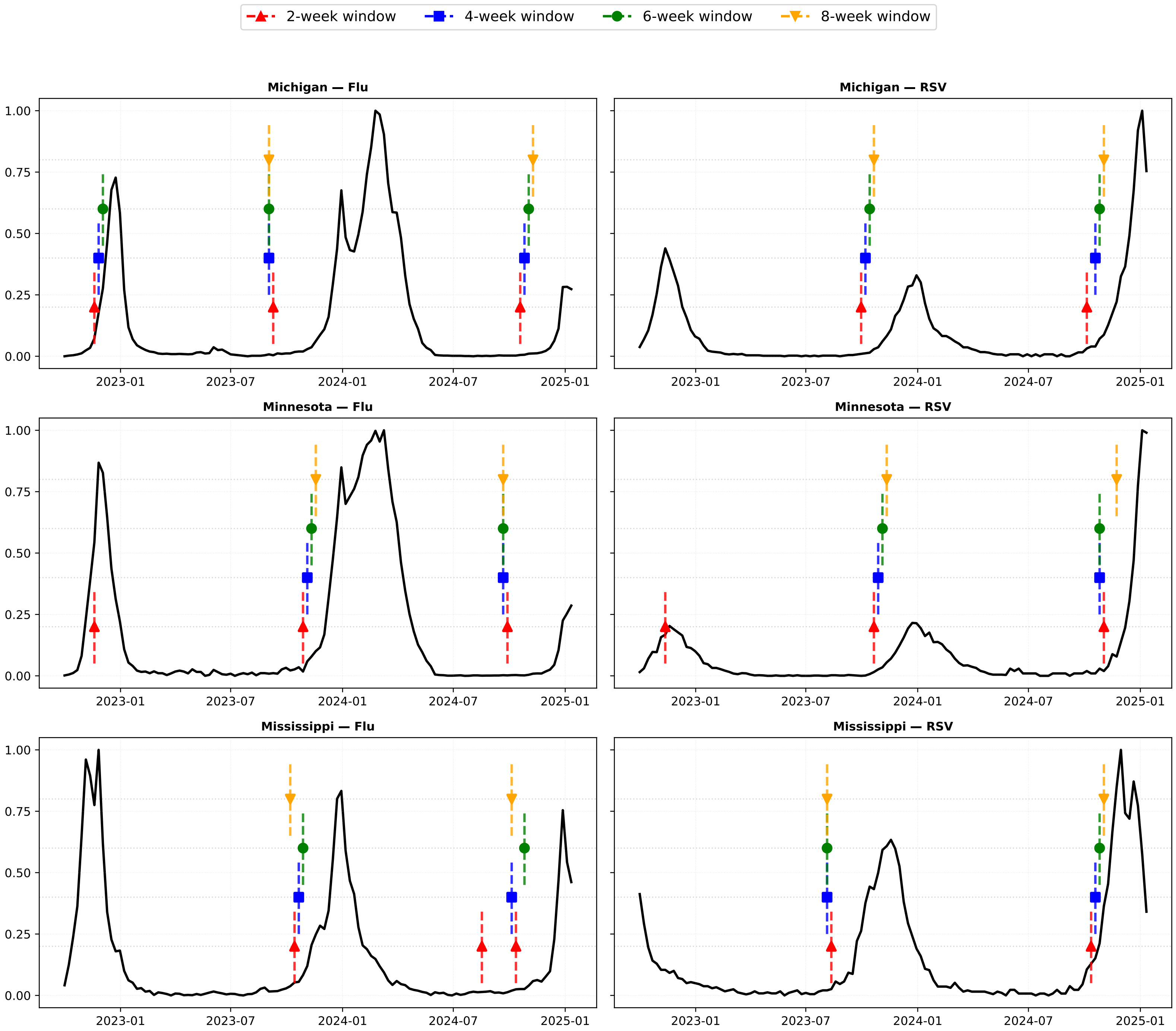

2-week window
4-week window
6-week window
8-week window
Michigan — Flu
Michigan — RSV
Minnesota — Flu
Minnesota — RSV
Mississippi — Flu
Mississippi — RSV
0.00
0.25
0.50
0.75
1.00
2023-01
2023-07
2024-01
2024-07
2025-01

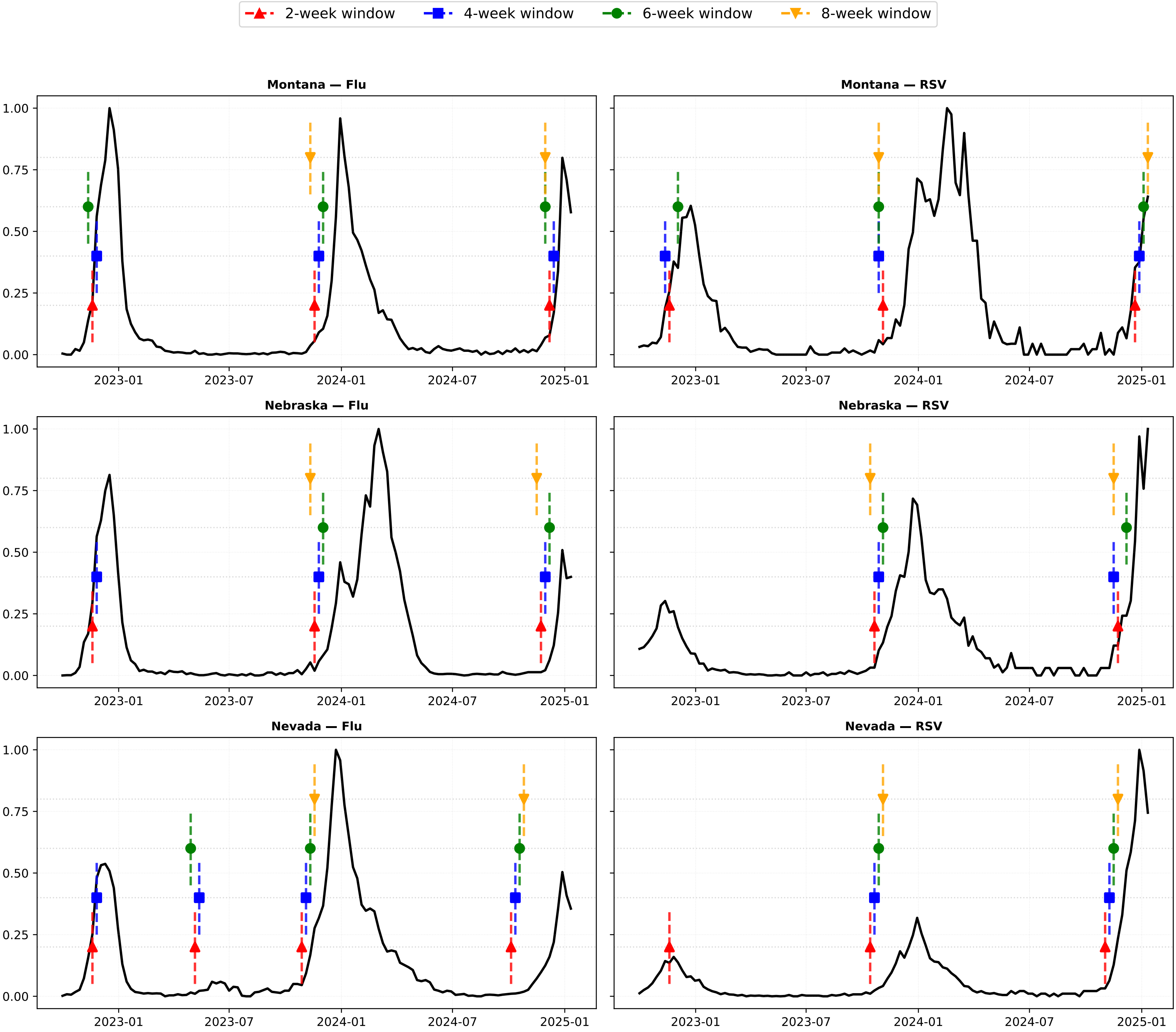
2-week window
4-week window
6-week window
8-week window
Montana — Flu
Montana — RSV
Nebraska — Flu
Nebraska — RSV
Nevada — Flu
Nevada — RSV
0.00
0.25
0.50
0.75
1.00
2023-01
2023-07
2024-01
2024-07
2025-01

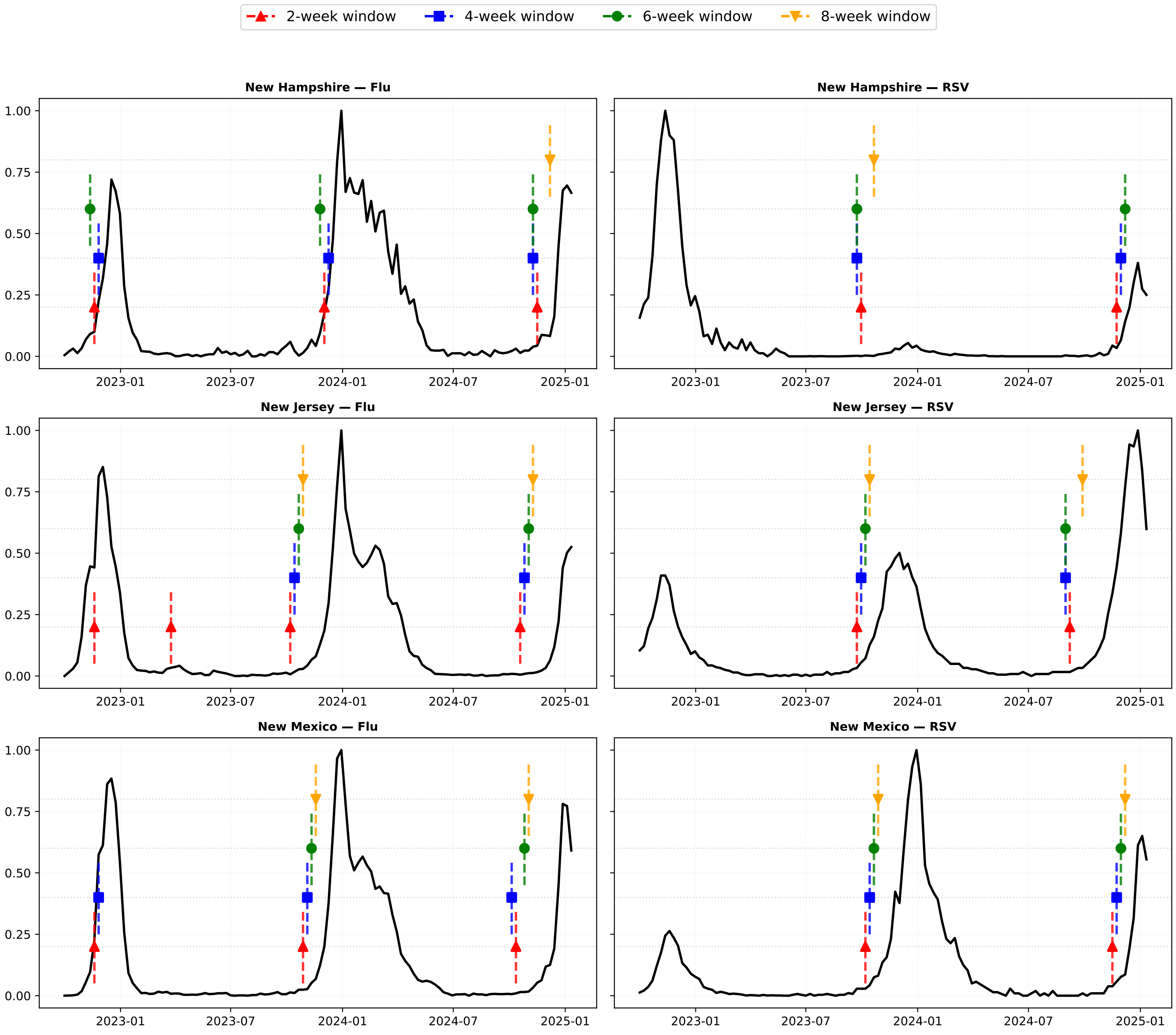
2-week window
4-week window
6-week window
8-week window
New Hampshire — Flu
New Hampshire — RSV
New Jersey — Flu
New Jersey — RSV
New Mexico — Flu
New Mexico — RSV
0.00
0.25
0.50
0.75
1.00
2023-01
2023-07
2024-01
2024-07
2025-01

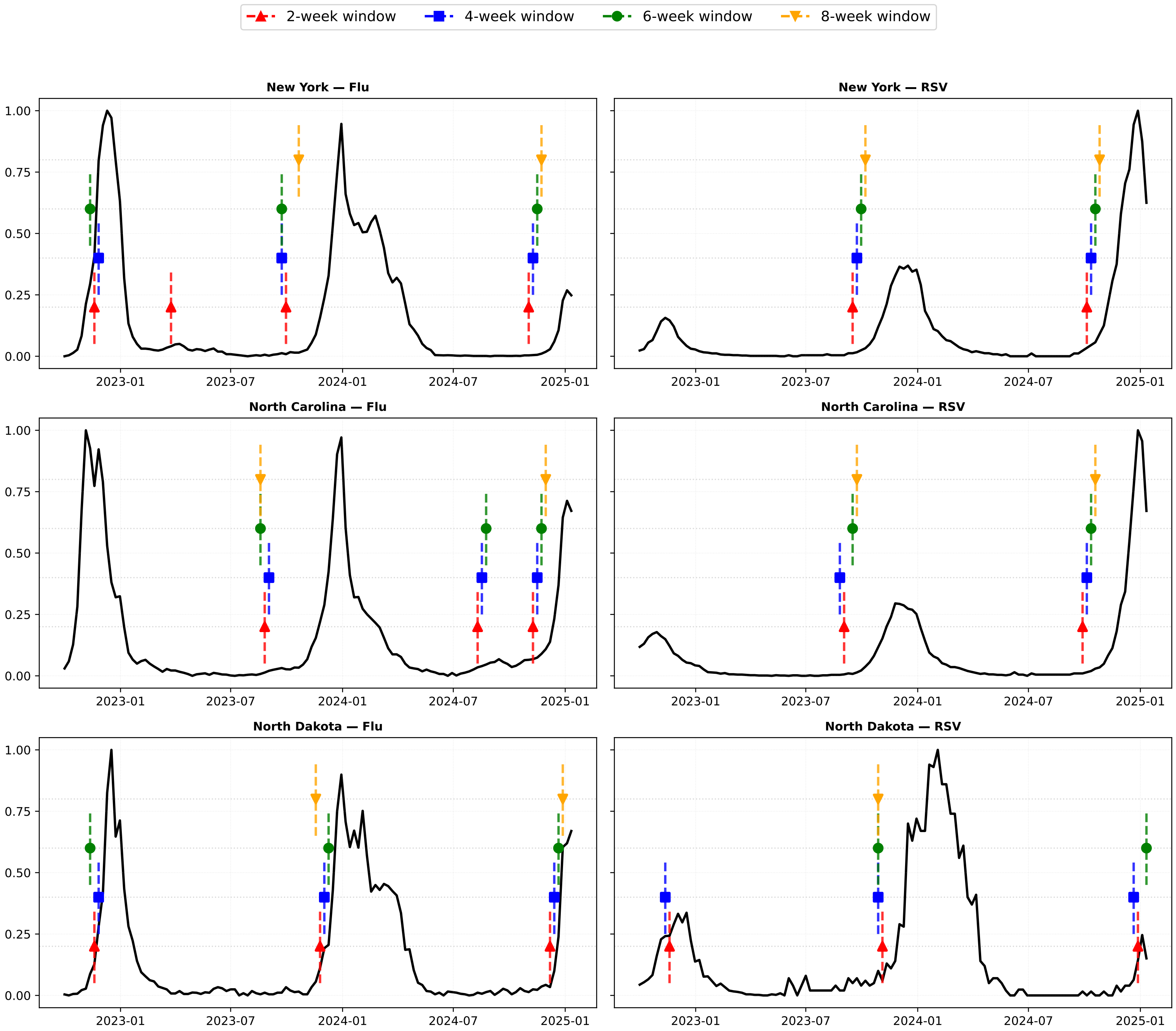

2-week window
4-week window
6-week window
8-week window
New York — Flu
New York — RSV
North Carolina — Flu
North Carolina — RSV
North Dakota — Flu
North Dakota — RSV
0.00
0.25
0.50
0.75
1.00
2023-01
2023-07
2024-01
2024-07
2025-01

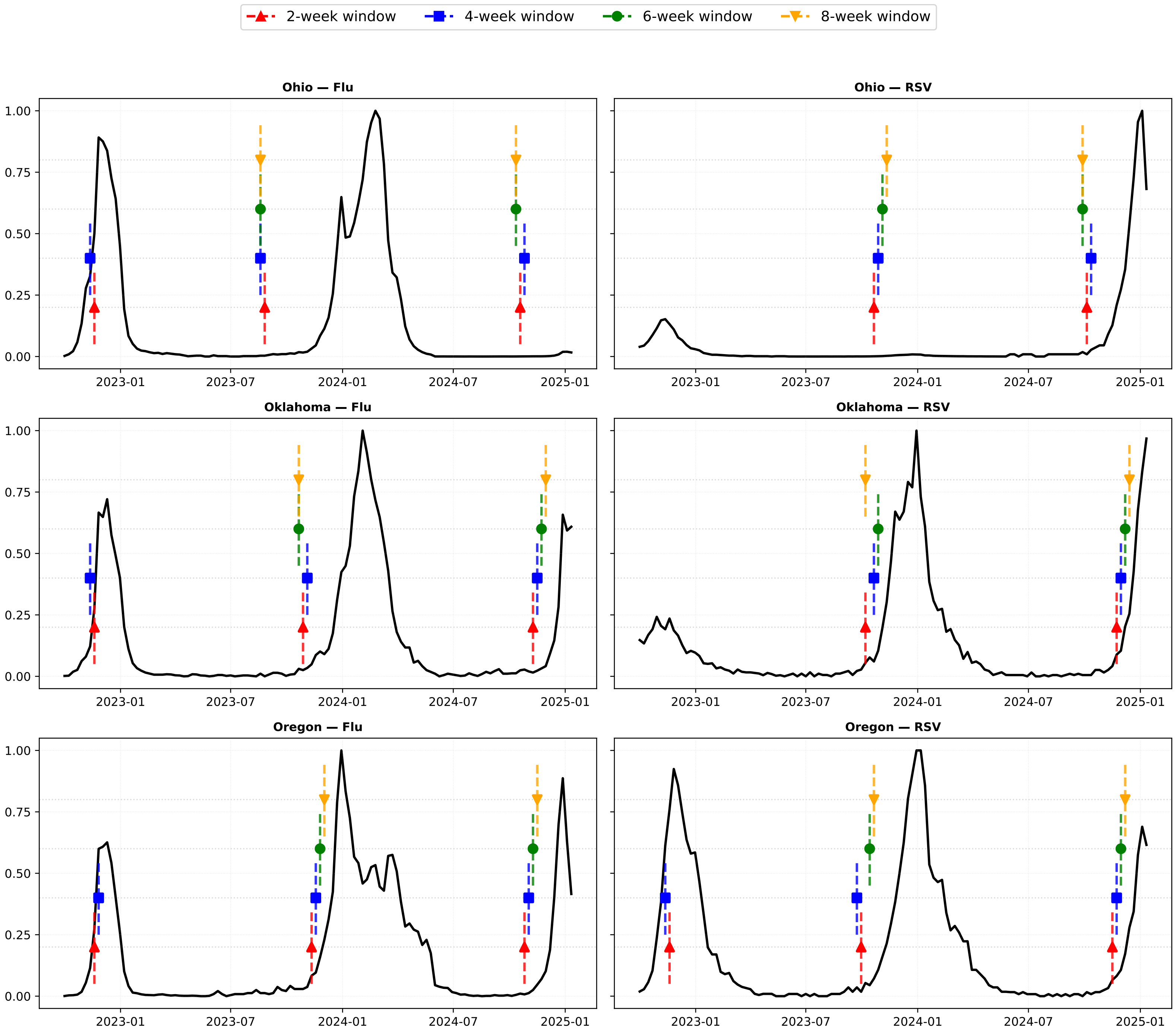

2-week window
4-week window
6-week window
8-week window
Ohio — Flu
Ohio — RSV
Oklahoma — Flu
Oklahoma — RSV
Oregon — Flu
Oregon — RSV
0.00
0.25
0.50
0.75
1.00
2023-01
2023-07
2024-01
2024-07
2025-01

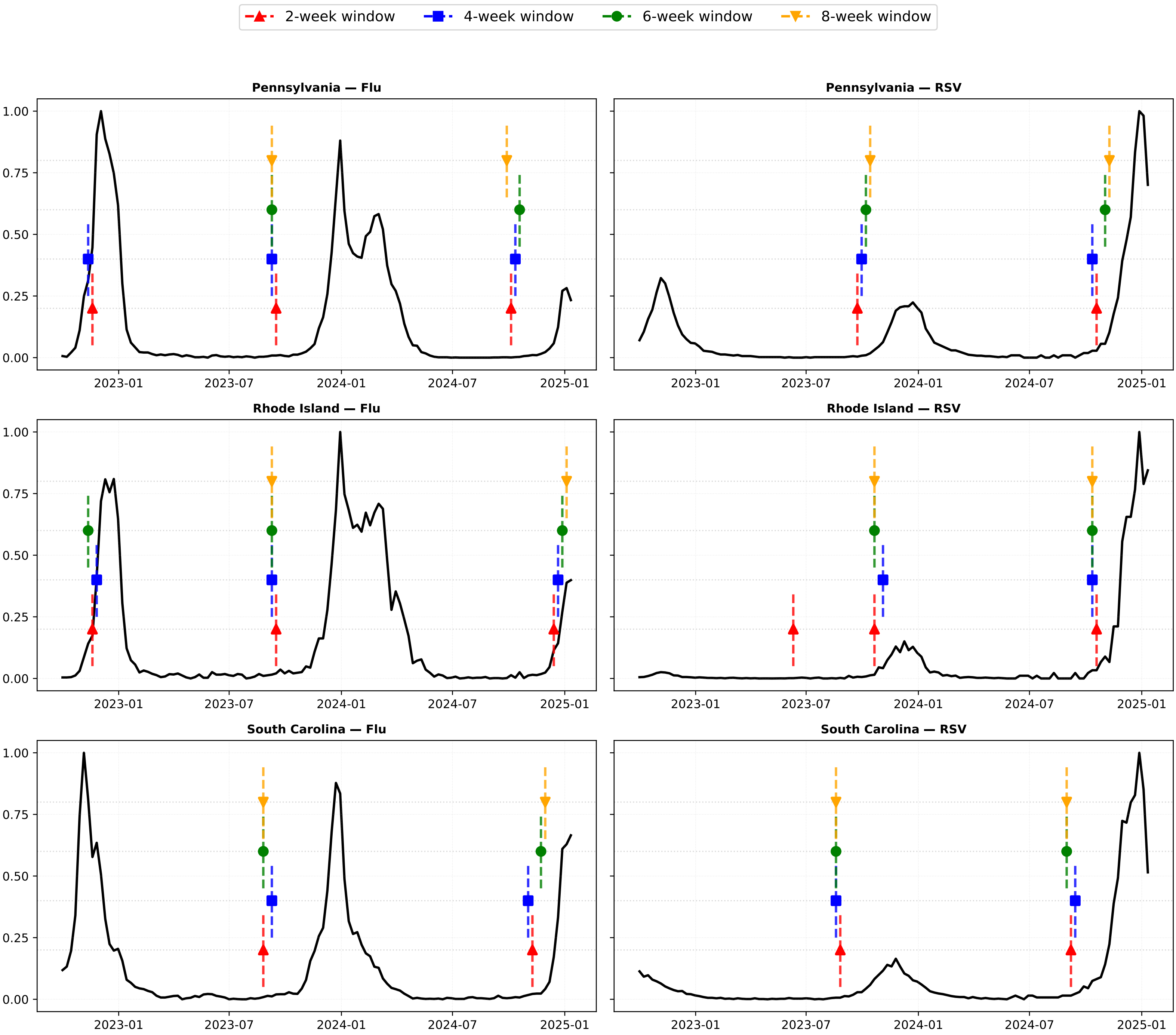

2-week window
4-week window
6-week window
8-week window
Pennsylvania — Flu
Pennsylvania — RSV
Rhode Island — Flu
Rhode Island — RSV
South Carolina — Flu
South Carolina — RSV
0.00
0.25
0.50
0.75
1.00
2023-01
2023-07
2024-01
2024-07
2025-01

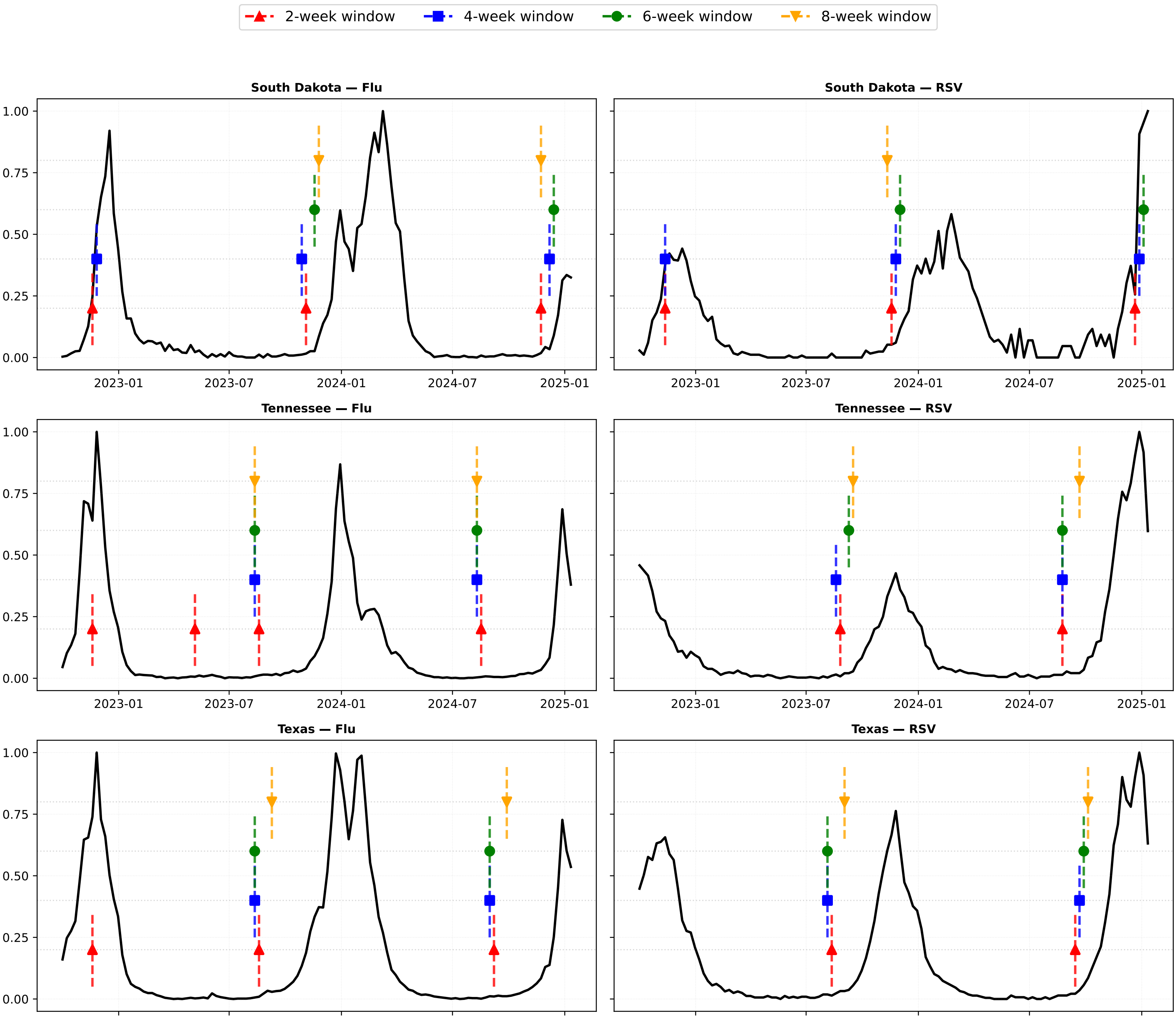

2-week window
4-week window
6-week window
8-week window
South Dakota — Flu
South Dakota — RSV
Tennessee — Flu
Tennessee — RSV
Texas — Flu
Texas — RSV
0.00
0.25
0.50
0.75
1.00
2023-01
2023-07
2024-01
2024-07
2025-01

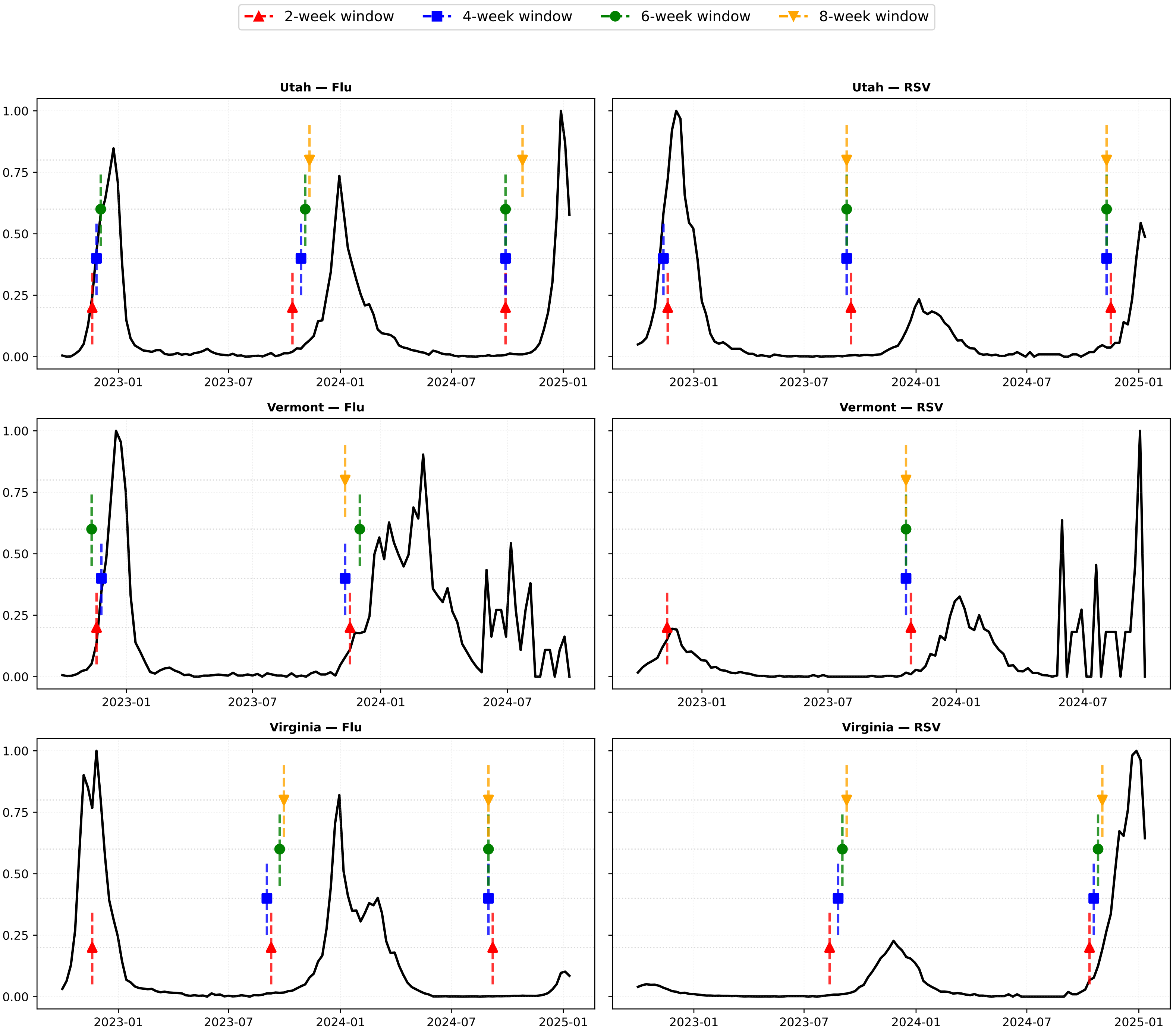
2-week window
4-week window
6-week window
8-week window
Utah — Flu
Utah — RSV
Vermont — Flu
Vermont — RSV
Virginia — Flu
Virginia — RSV
0.00
0.25
0.50
0.75
1.00
2023-01
2023-07
2024-01
2024-07
2025-01

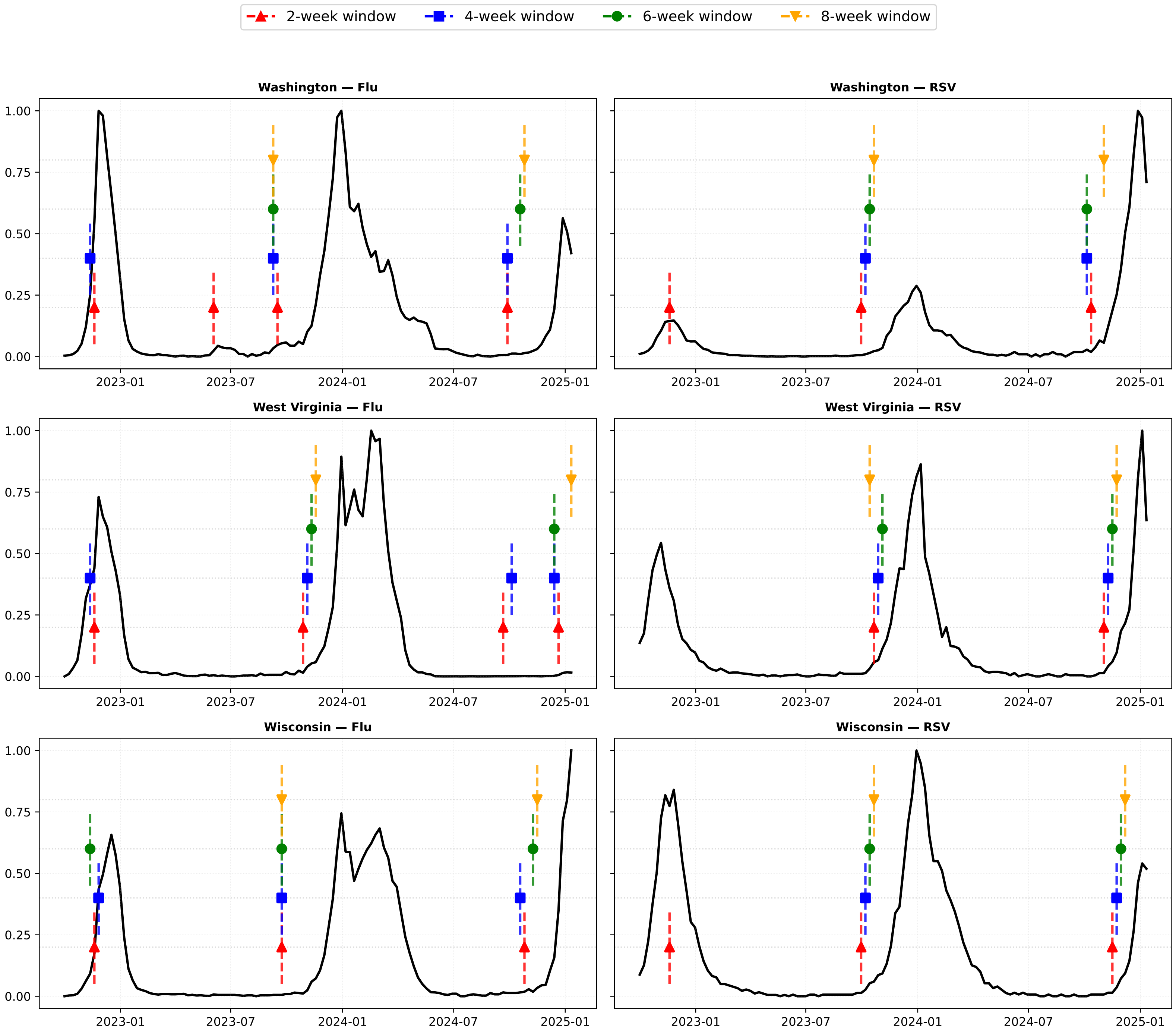

2-week window
4-week window
6-week window
8-week window
Washington — Flu
Washington — RSV
West Virginia — Flu
West Virginia — RSV
Wisconsin — Flu
Wisconsin — RSV
0.00
0.25
0.50
0.75
1.00
2023-01
2023-07
2024-01
2024-07
2025-01

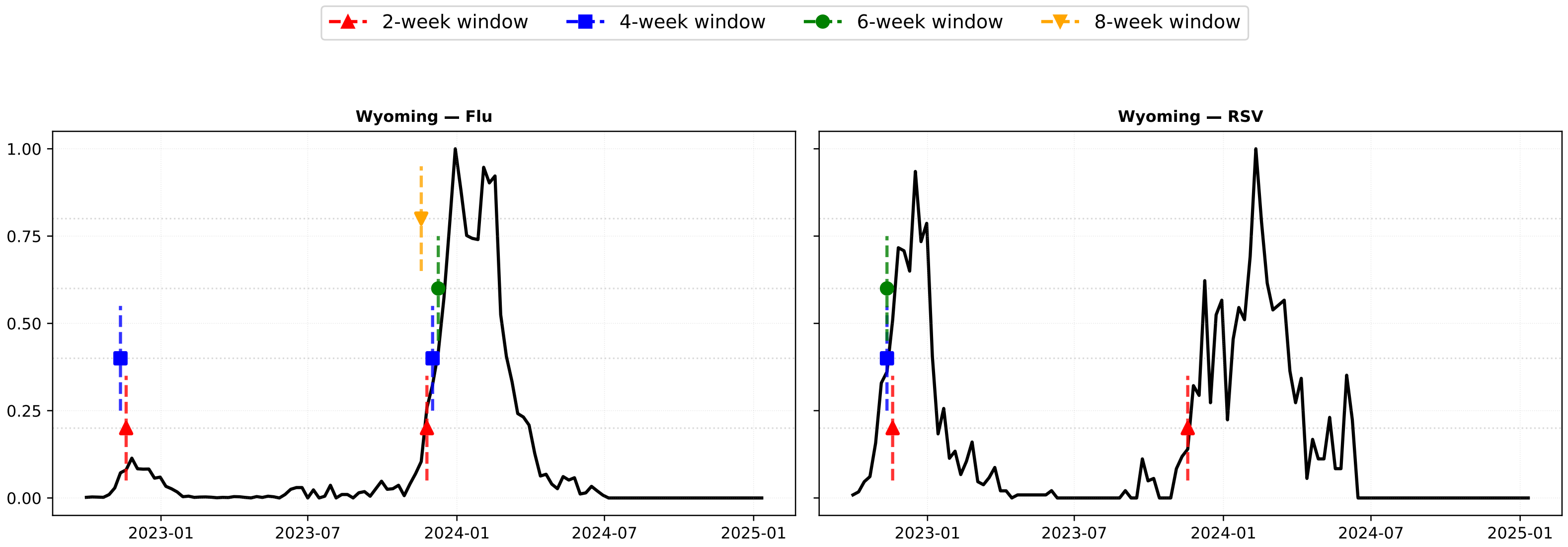

2-week window
4-week window
6-week window
8-week window
Wyoming — Flu
Wyoming — RSV
1.00
0.75
0.50
0.25
0.00
2023-01
2023-07
2024-01
2024-07
2025-01
